# Membrane theory with multimodal evidence defines blood pressure measurement

**DOI:** 10.64898/2026.10.22.26363749

**Authors:** Yingxian Sun, Haiyue Yu, Yijie Zhang, Bowei Hao, Alberto Avolio, Guozhe Sun, Lisheng Xu, Hongtai Li, Bo Bi, Xiangji Yue, Jian Li, Yang Li, Guangxiao Li, Xiaofan Guo, Tongtong Yu, Moujie Liu, Guan Wang, Xinhui Shao, Xuefeng Pang, Hongru Li, Haishan Zhang, Tiexin Zhang

## Abstract

Cuff-based blood pressure (BP) measurement is a cornerstone of health assessment, yet its mechanism remains unclear and measurement error remains substantial^1^. On the basis of membrane theory^2,3^, we propose that external pressure generated by cuff inflation follows a hyperbolic-cosine-shaped axial distribution along the artery, forming a pressure-peak zone (PPZ) beneath the cuff. By combining this external pressure with intra-arterial pressure, we constructed a mathematical model that systematically predicts site-specific vascular motion and hemodynamic changes during BP measurement. In humans, multimodal experiments integrating vascular imaging and synchronized invasive and noninvasive pressure measurements validated the model. The PPZ organizes distinct proximal and distal hemodynamics during cuff pressurization: at DBP, diastolic arterial collapse begins at the PPZ; near SBP, only weak residual blood flow traverses the PPZ and disappears once cuff pressure exceeds SBP. These transitions generated identifiable oscillogram features for DBP and SBP, precisely localized by wavelet scanning. The resulting method showed higher accuracy and lower variability than auscultation and conventional oscillometry and was the only method whose estimates did not differ significantly from invasive BP. This study redefines the hemodynamic basis of cuff-based BP measurement, and its mechanistic framework and accuracy may support clinical application and more reliable therapeutic decision-making.

**Structured Abstract:** *Introduction:* Brachial cuff-based noninvasive blood pressure (BP) measurement is among the most important measurements in medicine, yet its underlying mechanism remains unclear. Current oscillometric and auscultatory techniques rely largely on empirical criteria and show substantial errors and variability when compared with invasive BP. A core theoretical basis is therefore needed to link cuff-vessel coupling mechanics to hemodynamic changes and to support a more accurate, mechanism-consistent method for BP estimation.

*Rationale:* Traditional cuff-based BP theories assume vascular compression due to a spatially uniform external force. These theories cannot explain prior observations: tourniquet studies using cuffs similar to BP cuffs showed axial central tissue injury; intratissue pressure measurements and direct measurements of cuff contact pressure all showed curvilinear axial pressure distributions. Because the cuff is intrinsically a membrane structure, we hypothesized that sub-cuff pressure distribution should follow a membrane-theory contact model and a Laplace-type equilibrium. By combining this nonuniform external pressure with intra-arterial pressure, we constructed and validated a mathematical model that systematically predicts vascular motion, hemodynamics, and oscillogram changes during BP measurement.

*Results:* The model predicted that the cuff external pressure follows a hyperbolic-cosine-like axial distribution, forming a pressure-peak zone (PPZ) at the cuff axial center, the site of minimal curvature, with external pressure decreasing toward proximal and distal edges. Sub-cuff pressure-sensor measurements and transvenous tissue-pressure measurements in supine participants verified this pressure-distribution pattern and showed that, once cuff pressure exceeded approximately 30 mmHg, cuff pressure and tissue pressure in the PPZ were nearly identical. On this mechanistic basis, we established a physically constrained mathematical framework that reproduced vascular motion and hemodynamic changes during inflation and linked vascular dynamics to oscillogram morphology. Angiography, intravascular ultrasound (IVUS), computed tomographic angiography (CTA), and multisite invasive pressure measurements in the human brachial artery showed that the model-predicted motion, hemodynamics, multisite pressure, cross-sectional area, volume, and oscillogram changes agreed with measured results. The arterial PPZ was the primary site determining key state transitions of the sub-cuff vessel and the dominant contributor to hemodynamic and oscillogram changes. When cuff pressure rose above diastolic blood pressure (DBP) during inflation, the PPZ first collapsed during diastole, gradually creating two regions with distinct vascular motion proximal and distal to the PPZ. Near systolic blood pressure (SBP), distal pulsations became markedly delayed and attenuated into distinctive low-amplitude delayed pulsations (small waves). When cuff pressure exceeded SBP, pulsations in the PPZ and distal region disappeared, whereas proximal pulsations persisted. These hemodynamic events mapped onto characteristic oscillogram morphology transitions. Using wavelet scanning analysis, we extracted local oscillatory components as reference features for SBP and DBP. Auscultation, vascular imaging, and synchronized pressure recordings during the cardiac cycle showed that Korotkoff sounds (KS) occurred during early vascular opening and were related to expansion velocity. Our model further allowed estimation of the area-derived expansion velocity near the distal cuff edge; however, KS-generation thresholds vary across individuals. When cuff pressure is close to SBP, weak blood passage produces expansion velocities below the KS threshold, causing systematic underestimation. In contrast, near DBP, mild post-collapse re-expansion may trigger KS prematurely or with delay, leading to substantial variability. The model also showed that the conventional oscillometric amplitude-coefficient method for estimating SBP and DBP can be analytically derived from our mathematical equations. However, because these parameters are numerous and case-specific, they cannot be practically determined; therefore, the maximum amplitude algorithm (MAA) must rely on empirical ratios relative to the maximum oscillogram amplitude. Consequently, MAA exhibits not only systematic bias but also substantial variability. On the basis of this mechanism, we developed the novel Morphology-Transition Oscillometry (MTO) method for noninvasive BP measurement. For SBP, MTO showed consistently much higher accuracy and lower dispersion in 175 participants (0.39 ± 6.41 mmHg) than conventional auscultation (KS) (−6.58 ± 10.30 mmHg) and oscillometry based on MAA (-6.79 ± 10.58 mmHg), with no significant difference from invasive pressure. For DBP, the bias was 0.14 ± 6.25 mmHg for MTO, −1.15 ± 7.24 mmHg for KS, and 4.85 ± 9.14 mmHg for MAA.

**Conclusion:** This membrane-theory-based framework establishes a reliable new theory of cuff-based BP measurement, accurately predicting hemodynamic transitions, vascular motion, and oscillogram morphology, as validated by multimodal in vivo measurements. The MTO method derived from this theory measured arterial BP more accurately than auscultation and traditional oscillometry.

## Introduction

Blood pressure (BP) is a core physiological variable for health assessment and therapeutic decision-making. Although brachial cuff-based BP measurement has been used for more than a century and remains the most widely used noninvasive BP (NIBP) method ^1,4^, its hemodynamics, vascular motion, and external-internal pressure coupling still lack a unified theoretical explanation and systematic validation. With increasing use of intensive BP lowering ^5–8^ and AI-enabled wearable devices ^9^, an accurate, low-variability, interpretable NIBP method grounded in physical mechanisms is increasingly needed.

Auscultation has long been regarded as the standard NIBP method. Korotkoff sounds (KS) appearance is generally considered to indicate intermittent arterial opening and blood passage through the compressed segment, whereas KS disappearance indicates restoration of arterial patency throughout the cardiac cycle ^4^. Yet auscultatory and oscillometric methods remain essentially empirical and lack mechanistic anchors to hemodynamic events ^10,11^. Since the 1970s, cuff compression has often been simplified as a single, spatially uniform external pressure in models of oscillation amplitude and auscultation, implicitly assuming an axially uniform arterial response ^12–23^. This conflicts with evidence that tourniquet cuffs comparable to BP cuffs cause tissue injury within a narrow mid-cuff region ^24^, intratissue pressure is centrally elevated and decreases curvilinearly toward the edges ^25^, and cuff contact is similarly enhanced centrally ^26,27^. This incomplete mechanistic understanding may contribute to substantial measurement error.

Synchronized invasive studies have shown substantial systolic blood pressure (SBP) underestimation by KS-based endpoints (mean bias, −20 to −14 mmHg; standard deviation (SD), 9.5 to 11.6 mmHg; root mean square error (RMSE) up to 23.2 mmHg) ^28^. Oscillometric devices likewise rely on empirical fixed amplitude ratios ^19,29^ and underestimate SBP (mean bias, −19 to −17 mmHg; SD, 11.2 to 13.2 mmHg; RMSE up to 23.0 mmHg) ^28^, with extreme errors > 50 mmHg ^1^ and SDs up to 19.25 mmHg ^30^, indicating substantial individual error dispersion. Thus, current NIBP methods are limited by both substantial error dispersion relative to invasive pressure and the absence of a mechanistic framework linking cuff signals to hemodynamic events.

We therefore hypothesized that the cuff behaves as a membrane structure whose deformation and tension distribution after inflation follow membrane theory and Laplace-type mechanical equilibrium ^2,3,31^, while tissue stiffness transmits the resulting axial pressure distribution to the vascular wall ^32,33^. We combined intra-arterial BP with the nonuniform external-pressure field predicted by membrane theory to construct a coupled cuff–artery model of vascular motion, hemodynamics, and oscillogram morphology. Synchronized multimodal measurements of vascular imaging, intravascular pressure, and cuff-interface loading validated the model and spatially linked oscillogram signals to axial vascular events. Wavelet transform was used to localize SBP-and diastolic blood pressure (DBP)-related oscillogram features and establish a novel BP measurement method. We compared this method with conventional NIBP approaches to assess performance and identify their limitations and error mechanisms. This study establishes a new theoretical framework and an accurate, interpretable mechanistic basis for NIBP measurement, with potential to improve measurement reliability while supporting future clinical translation and wearable-device development.

## Results

### 1. A coupled cuff–artery model based on membrane theory reveals a nonuniform axial pressure field and pressure-peak zone

On the basis of membrane theory and a Laplace-type mechanical equilibrium relation describing the relationship between transmural pressure (P) and circumferential tension (T) in a thin-walled tube of radius r (T = P·r), we modeled the cuff-tissue interface as a thin-membrane system driven by cuff pressure and constrained by soft-tissue support. Under the small-slope approximation, the governing equation is:

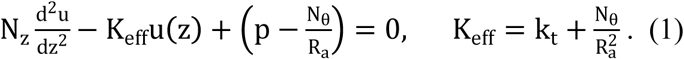

Here, u(z) is normal indentation displacement along the axial coordinate (z); N_θ_ and N_z_ are circumferential and axial effective membrane-force resultants, respectively; R_a_ is effective upper-arm radius; k_t_ is soft tissue support stiffness; p is the driving pressure acting on the cuff membrane before subtraction of the local tissue-contact reaction (Supplementary Fig. 3). We defined normalized contact pressure as 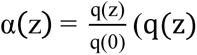 (q(z) and q(0) are defined in Supplementary Information, Section S2.1) and introduced the dimensionless coordinate 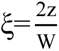, yielding the closed-form expression:

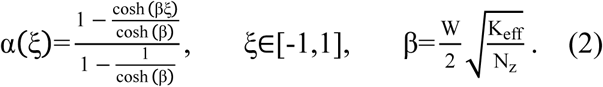

Here, W is effective cuff width, and β is a dimensionless structural parameter that collectively represents tissue support, membrane-force coupling, and edge-constraint effects. The resulting cuff–skin contact-pressure profile forms a hyperbolic-cosine-like arc, with a central pressure-peak zone (PPZ) where peak pressure equals cuff pressure and decreases toward both edges (Fig. 1a). Full assumptions, boundary conditions, and derivation are provided in Supplementary Information, Section S2.1.

**Fig. 1.**
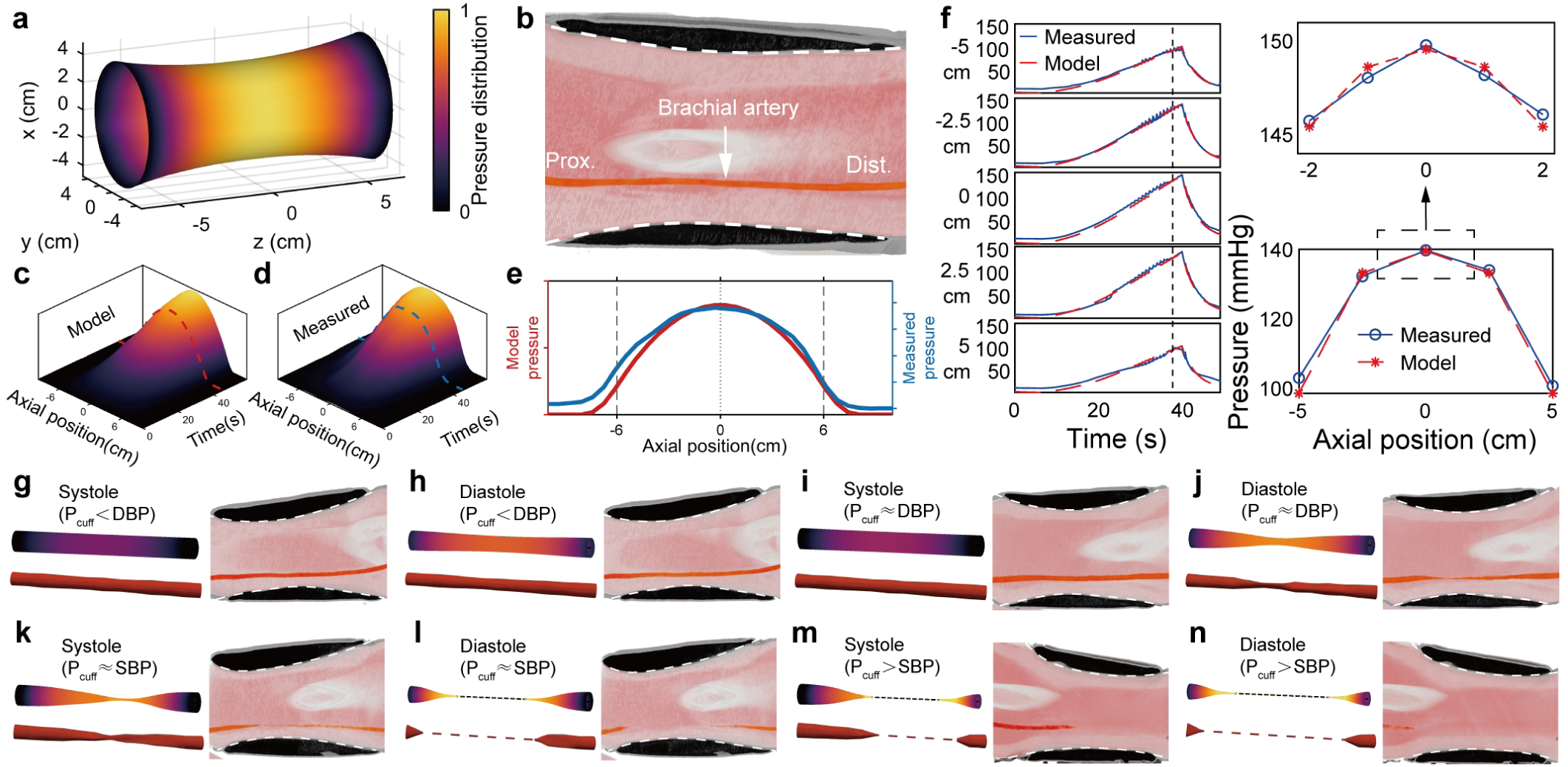
Theoretical prediction and multimodal validation of the axial cuff-pressure distribution. **a**, Cuff inner-edge geometry derived from the membrane-structure assumption and model-predicted sub-cuff pressure distribution; z axis denotes axial position. The cuff midpoint was defined as axial position 0, with the proximal direction assigned negative values and the distal direction assigned positive values. The heat map denotes the predicted normalized contact-pressure distribution. **b**, Three-dimensional photon-counting computed tomography (CT) reconstruction showing the spatial relationship between the inflated cuff and upper-arm tissues. The white dashed line indicates the curved cuff–skin contact interface. **c-e**, Model-predicted (**c**) and flexible pressure sensor-measured (**d**) axial cuff–skin contact-pressure distributions during cuff inflation. The z-axis height and heat-map color encode contact pressure; the x and y axes denote axial position and time, respectively. Dashed lines indicate the representative time point used for the axial pressure-profile comparison in (**e**). **e**, Compares the model-predicted and measured axial pressure profiles at that time point. **f**, Comparison between multisite surrogate deep-tissue pressure measurements and model-predicted pressures during the complete inflation measurement (left), and a snapshot comparison of the axial pressure distribution at a representative time point (right). The upper plot focuses on the central pressure-peak zone (PPZ), whereas the lower plot shows the pressure distribution across most of the cuff-covered segment. **g-n**, Morphological changes of the sub-cuff brachial artery during systole and diastole at different cuff pressures. Each set includes model-predicted arterial motion (left upper), three-dimensional reconstruction of multisite intravascular ultrasound (IVUS) contours (left lower), and vascular morphology observed by CT angiography (right).

Three-dimensional (3D) photon-counting computed tomography (CT) reconstruction supported this predicted arc shape (white dashed line, Fig. 1b). The conical upper-arm geometry could shift the PPZ slightly from the cuff midpoint, usually proximally, but subsequent vascular and hemodynamic transitions remained centered on this region.

In 50 participants, flexible pressure-sensing pads placed beneath the cuff showed a curvilinear contact-pressure distribution closely matching model predictions (Fig. 1, c to e).To determine whether this axial arc-shaped compression was transmitted into deeper tissue, we measured sub-cuff brachial venous pressure with pressure guidewires in 5 participants. Once cuff pressure exceeded approximately 30 mmHg, the axial pressure profile showed a curvilinear pattern, with PPZ pressure closely matching cuff pressure (Fig. 1f), in close agreement with model predictions.

We next coupled this nonuniform external-pressure field with intra-arterial pressure, defining transmural pressure as:

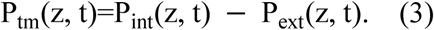

Here, P_tm_(z, t) is the transmural-pressure field, P_ext_(z, t) is the nonuniformly transmitted external cuff-pressure field, and P_int_(z, t) is the intravascular pressure field. The model expressed the target cross-sectional area (CSA) A_eq_(z, t) as a smooth combination of open and closed arterial states:

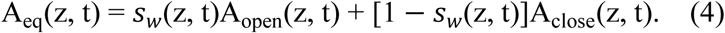

Here, A_open_ denotes the area governed by positive transmural pressure, A_close_ denotes the collapsed-segment area governed by negative transmural pressure, and (z, t) is the smooth switching coefficient between the open and closed segments, determined by transmural pressure and ranging from 0 to 1.

Model predictions and multimodal observations across different cuff pressures are shown in Fig. 1, g to n. For cuff pressure ≤ DBP (Phase Ⅰ), arterial pulsation differed little from the pre-measurement state (Fig. 1, g and h). Once cuff pressure > DBP (Phase II), the artery continued to expand during systole, whereas during diastole it underwent abrupt collapse at the PPZ (Fig. 1i, j). Progressive changes then divided the proximal and distal sides of the PPZ into two hemodynamically distinct regions, with the artery remaining open during systole but occluded during diastole. As cuff pressure approached SBP, vascular motion distal to the PPZ became delayed relative to that in the proximal segment (Fig. 1, k to l, Supplementary Video 1). When cuff pressure exceeded SBP (Phase Ⅲ), the PPZ became occluded, pulsation disappeared in the PPZ and distal segment, while proximal systolic flow persisted (Fig. 1m, n). The fully occluded segment subsequently extended toward both sides as cuff pressure increased. Complete model equations, parameter definitions, default values, and dynamic processes are provided in Supplementary Information, Section S2.2, Supplementary Tables 2 and 3, and Supplementary Video 1. To clearly characterize the sequence of arterial compression, collapse, and occlusion under increasing external pressure, the main results focus primarily on cuff inflation; corresponding deflation findings are provided in the Supplementary Information.

For validation, intravascular ultrasound (IVUS) imaging was performed at 1-cm axial intervals beneath the cuff. Cross-sectional images from 13 positions were time-registered and spatially reconstructed to obtain a 3D representation of brachial arterial motion throughout cuff pressurization. The model prediction agreed well with the IVUS-based dynamic imaging and CT imaging at different preset measurement-stage pressures (Fig. 1, g to n, Supplementary Videos 2–6). The CTA measurement protocol is described in Supplementary Information, Section S1.1, S1.7 and Supplementary Fig. 1.

### 2. The pressure-peak zone organizes sub-cuff vascular motion and hemodynamic separation

Figure 2, a to f, shows brachial artery motion measured by angiography and multisite IVUS during cuff inflation (Supplementary Video 7), consistent with the model predictions in Fig. 1, g to n. IVUS shows CSA changes at five representative axial sites (Supplementary Information, Section S1.6).

**Fig. 2.**
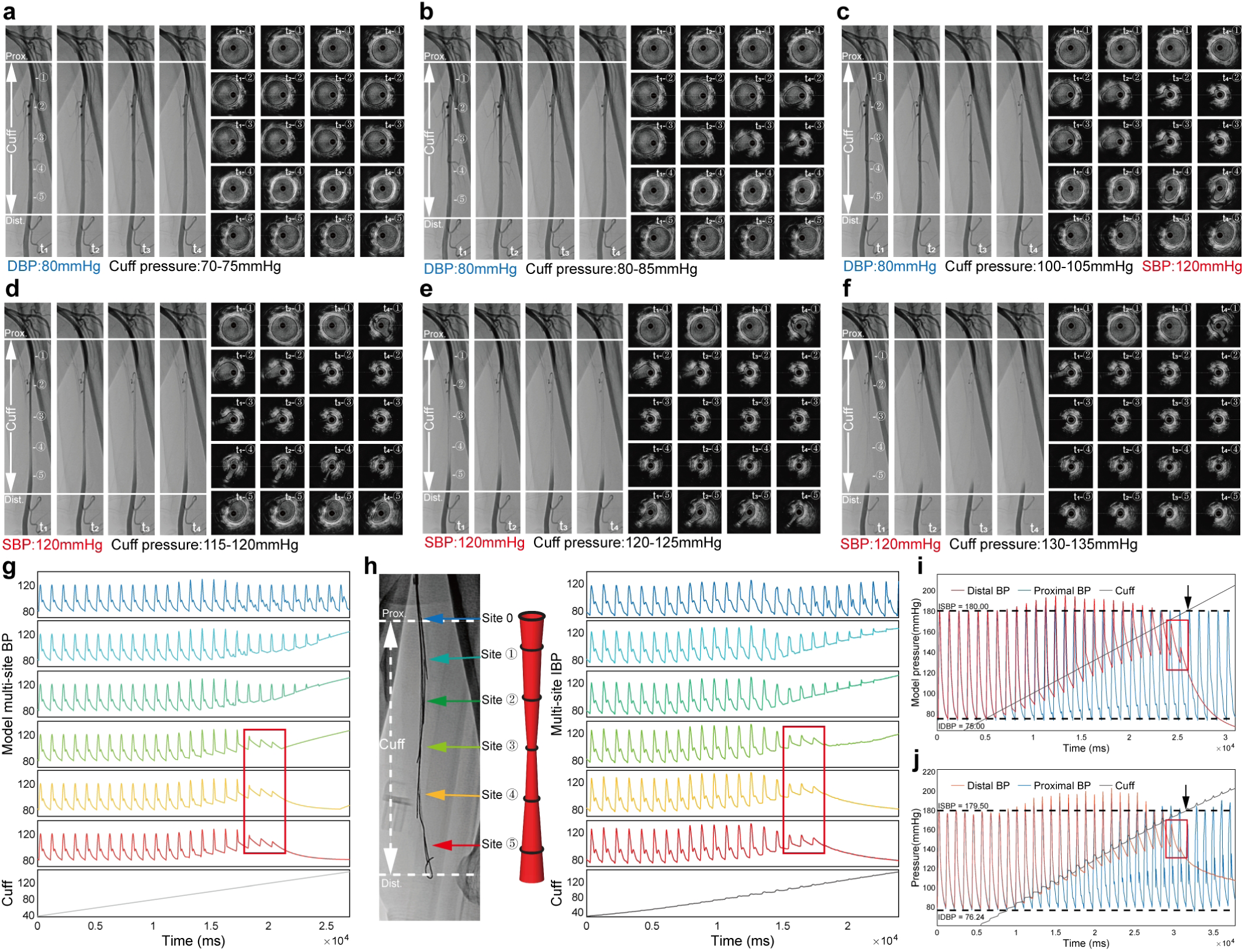
Experimental validation of global arterial motion and model-predicted hemodynamics. **a-f**, Angiographic and multisite IVUS observations during cuff inflation at cuff-pressure ranges of 70–75 mmHg, 80–85 mmHg (approximately diastolic blood pressure, DBP), 100–105 mmHg, 115–120 mmHg, 120–125 mmHg (approximately systolic blood pressure, SBP), and 130–135 mmHg, respectively. In each panel, t1 to t4 indicate four sequential time points within the diastolic interval of a single cardiac cycle. Site ① to ⑤ indicate five IVUS measurement sites arranged along the arterial axis; their axial positions are marked on the angiographic image at time point t1. **g** and **h**, Model-predicted and experimentally measured intra-arterial pressure changes at multiple sub-cuff brachial artery sites. In (**h**), the left image shows the axial position of the measured intravascular site, while the middle section depicts a schematic diagram of the brachial artery. Red boxes mark the appearance of mid-to-distal small waves, defined here as residual low-amplitude pressure pulsations, when cuff pressure approaches SBP. **i** and **j**, Model-predicted and experimentally measured relationships between proximal and distal sub-cuff pressure waveforms. Dashed lines indicate the baseline mean invasive blood pressure (IBP) before cuff inflation. Red boxes mark small waves in the distal pressure waveform when cuff pressure approaches SBP; black arrows indicate the cuff pressure at disappearance of the last small wave.

When cuff pressure was below DBP, Sites ① to ⑤ showed only slight systolic and diastolic motion that increased gradually with cuff pressure (Fig. 2a). At the beginning of Phase Ⅱ, just above DBP, end-diastolic narrowing first appeared within the PPZ (Fig. 2b, angiography at t4), with IVUS showing collapse at Site ③ (Fig. 2b, IVUS at Site ③ at t4; corresponding to Fig. 1i, j). During Phase Ⅱ, the PPZ and adjacent arterial segment gradually became occluded during diastole and reopened during systole (Fig. 2c, angiography at t1–t4). Diastolic occlusion and systolic reopening centered on Site ③ and extended axially toward Sites ② and ④ (Fig. 2c, IVUS ② to ④).

In the cardiac cycle immediately before cuff pressure reached SBP, corresponding to the end of Phase Ⅱ, systolic blood flow still traversed the PPZ, whereas during diastole the segment closed rapidly, gradually forming an occlusion zone centered on the PPZ (Fig. 2d, angiography at t2–t4, IVUS ② to ④, corresponding to Fig. 1k, l). When cuff pressure rose just above SBP (onset of Phase Ⅲ), a conical column of proximal systolic blood flow extended past Site ② but did not reach Site ③. Sites ③ and ④ remained continuously occluded throughout the observation cycle, forming a stable occlusion point (Fig. 2e, angiography at t1–t4, IVUS ③ to ④).

Meanwhile, the distal brachial artery remained open but lost volumetric pulsation and was gradually compressed closed (Fig. 2e, IVUS ⑤). With further cuff pressurization, the occluded segment continued to extend proximally and distally, while the systolic blood column entered progressively less of the sub-cuff artery (Fig. 2f, angiography at t1). Sites ③ to ⑤ remained completely occluded (Fig. 2f, IVUS ③ to ⑤; corresponding to Fig. 1m, n). These findings matched the model-predicted Phase Ⅰ-Ⅲ changes and were consistently observed at the available measurement sites across all 22 angiography and 28 IVUS participants.

Figure 2, g and h, shows model-predicted and measured multisite hemodynamic changes during inflation, respectively. The measurement sites included the proximal cuff edge (Site 0) and the five sites described above (Sites ① to ⑤). During Phase Ⅱ at Site 0, SBP and DBP rose modestly for a short period and then declined, with maximal increases from baseline of 12.35 ± 7.87 mmHg for SBP and 3.65 ± 3.96 mmHg for DBP (n = 37; N = 124 measurements). At Site ③, SBP showed a similar pattern, whereas DBP increased with cuff pressure because of vascular occlusion. The PPZ thereby separated proximal and distal sub-cuff brachial artery hemodynamics. As cuff pressure approached SBP, restricted blood flow propagation through the PPZ and distal region produced delayed, low-amplitude residual pressure pulsations (small waves; measured mean number of small waves, 4.81 ± 2.58; n = 37; N = 124 measurements) (red box, Fig. 2, g and h). When cuff pressure reached SBP, the artery became completely occluded, and small waves in the middle-to-distal segments disappeared simultaneously. Model predictions at all six sites agreed closely with the measured results and indicated that the complete occlusion point was generally located at the cuff center. The same patterns were observed in all 8 participants undergoing multisite arterial pressure measurements (Supplementary Information, Section S2.3 and Supplementary Figs. 4-6).

Figure 2i, j, shows sub-cuff proximal and distal pressure waveforms obtained from model calculations and measurements, respectively. For each measurement, invasive arterial pressure averaged over approximately 30 s before cuff inflation was used as the reference BP, and the cuff pressure at the end of the final small wave in the middle-to-distal artery was closest to this reference SBP (black arrows, Fig. 2i, j). This represented the only cuff-measurable SBP-related point and could therefore serve as a signature for measurable SBP. In contrast, DBP was associated with the first rapid arterial collapse.

### 3. Axial arterial volume changes generate oscillogram morphology

Figure 3, a to e, shows close agreement between model-predicted and measured CSA changes at five axial arterial sites, consistent with the staged vascular and hemodynamic changes shown in Figs. 1 and 2.

**Fig. 3.**
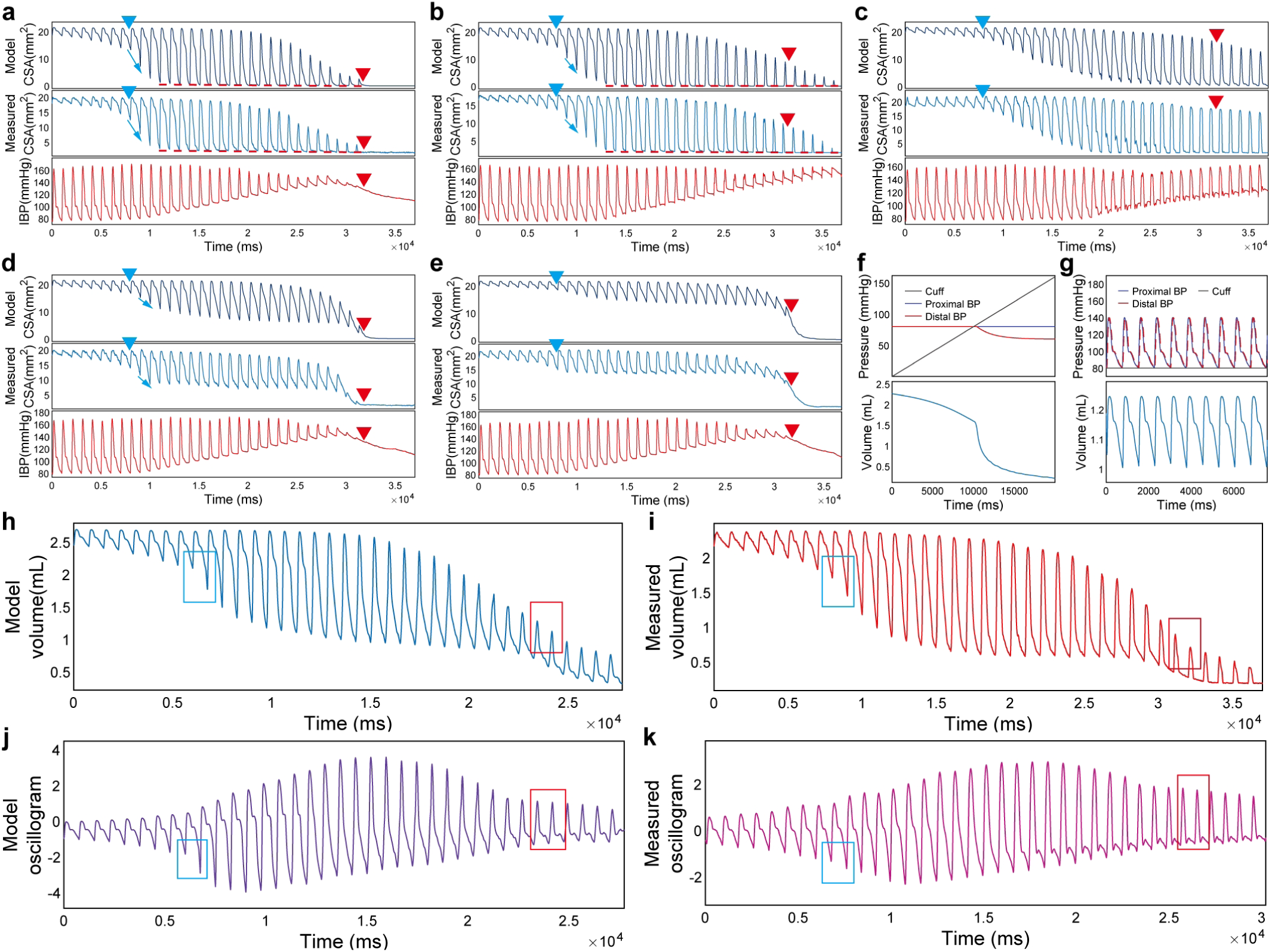
Axial vascular wall motion, segmental volume changes, and their impact on oscillometric waveform morphology. **a-e**, Axial measurement sites corresponding to Site ③, Site ②, Site ①, Site ④, and Site ⑤, respectively. Within each panel, the upper, middle, and lower subpanels show model-predicted vascular cross-sectional area (CSA), IVUS-measured CSA, and the corresponding IBP, respectively. The model was driven by the subject’s baseline arterial pressure waveform to predict site-specific vascular wall motion during cuff inflation. Blue and red triangles indicate phase boundaries, red dashed lines denote diastolic occlusion, and blue arrows mark the vascular collapse process. **f** and **g**, Model-predicted proximal pressure, distal pressure, cuff pressure, and sub-cuff vascular segment volume under constant arterial pressure with progressively increasing cuff pressure **(f)** and under constant cuff pressure with pulsatile proximal pressure input **(g)**. **h** and **i**, Model-predicted and IVUS-reconstructed sub-cuff brachial arterial segment volume during cuff inflation, respectively. The measured volume was reconstructed by axial interpolation of CSA values from the five IVUS measurement sites. Blue boxes indicate the volume change associated with abrupt PPZ collapse near DBP, whereas red boxes indicate the volume change associated with disappearance of distal arterial pulsation near SBP. **j** and **k**, Model-predicted and experimentally measured oscillometric waveforms, respectively. Blue and red boxes indicate DBP-and SBP-related oscillometric morphological transitions, respectively; these transitions are consistent with the corresponding segmental volume waveform changes shown in **(h)** and **(i)**.

Figure 3a corresponds to Site ③ in Fig. 2, representing the PPZ. Diastolic CSA decreased mildly and progressively during Phase Ⅰ (blue triangles in Fig. 3, a to e mark the end of Phase Ⅰ). At the beginning of Phase Ⅱ, maximal systolic CSA remained largely preserved, whereas diastolic CSA fell abruptly because of arterial collapse (blue arrow, Fig. 3a) and reached 0 after several cardiac cycles (red dashed line, Fig. 3a), when CSA variation peaked. The PPZ then separated the proximal and distal sub-cuff arterial segments into distinct hemodynamic systems. During mid-Phase Ⅱ, maximal systolic CSA and the systolic–diastolic CSA difference progressively declined. Once cuff pressure exceeded DBP, the diastolic pressure level continued to rise with cuff pressure, whereas SBP rose slightly for a period and then declined; as cuff pressure approached SBP, the waveform gradually evolved into increasingly delayed, lower-amplitude small waves. At the beginning of Phase Ⅲ (red triangles in Fig. 3, a to e mark the Phase Ⅱ–Ⅲ boundary), the PPZ became completely occluded, CSA and its pulsatile variation fell to 0, and the small waves disappeared.

Figure 3b corresponds to Site ②. Phase I changes were similar to those at Site ③. In Phase Ⅱ, initial CSA collapse occurred simultaneously with Site ③, but pronounced CSA collapse and DBP elevation developed later (blue arrow and red dashed line, Fig. 3b), whereas SBP increased slightly. After cuff pressure exceeded SBP (Phase Ⅲ), pressure and CSA pulsations persisted for several cardiac cycles at Site ② but disappeared immediately at Site ③ (red triangle, Fig. 3b).

Figure 3c corresponds to Site ①. Diastolic CSA declined gradually during Phase Ⅱ and reached 0 later than at Site ②, after which systolic CSA declined. Area variation remained relatively large in Phase Ⅲ. Diastolic pressure rose in late Phase Ⅱ but less than at Sites ② and ③, whereas the SBP increase was similar. Large area and pressure waveform pulsations persisted in Phase Ⅲ. Overall, Sites ① and ② represented the proximal sub-cuff region, where pulsations persisted after cuff pressure exceeded SBP because proximal systolic flow continued to enter beneath the cuff but could not pass through the occluded PPZ.

Figure 3d corresponds to Site ④. At the beginning of Phase Ⅱ, diastolic collapse began simultaneously with that at Site ③ but became pronounced later (blue triangle and arrow, Fig. 3d), while CSA variation gradually increased. After diastolic PPZ occlusion, incomplete emptying caused diastolic CSA to plateau above 0. Near the end of Phase Ⅱ, minimum diastolic and maximal systolic CSA declined synchronously, with a greater decline in maximal systolic CSA, leaving residual CSA pulsations corresponding to the pressure small waves; both disappeared simultaneously with those at Site ③. In Phase Ⅲ, pulsatile CSA variation was lost and CSA fell to 0 (red triangle, Fig. 3d). Pressure changes resembled those at Site ③, including DBP elevation, slight SBP elevation, small-wave formation and eventual disappearance.

Figure 3e corresponds to Site ⑤. CSA changes similarly showed a plateau and disappearance of residual pulsations but DBP-related collapse was less pronounced, minimum diastolic CSA declined more slowly to a higher plateau, and CSA variation was smaller. Pressure changes also resembled those at Site ④, including DBP elevation, small-wave formation and synchronous disappearance. Overall, Sites ④ and ⑤ represented the distal sub-cuff region, where all pulsatile volume and pressure changes disappeared after cuff pressure exceeded SBP, leaving only progressive compression and emptying as cuff pressure increased. The same pattern was observed in the other 4 participants undergoing multisite IVUS measurements.

Figure 3f shows the model-inferred relationship among cuff pressure, intra-arterial pressure, and lumen volume under constant intra-arterial pressure. With smoothly changing cuff pressure, the volume curve showed only one transition near zero transmural pressure. In contrast, when cuff pressure was held constant and intra-arterial pressure varied over the cardiac cycle (Fig. 3g), multiple matched inflection points appeared in the pressure and volume curves. These findings indicate that volume-curve inflections were determined mainly by BP pulsation rather than smoothly changing cuff pressure. Although waveform scale differed across cuff-pressure stages, the timing of these features remained consistent.

Figure 3h shows model-calculated arterial volume change during cuff inflation using measured proximal pressure as input. Figure 3i shows the continuous volume waveform reconstructed by axial linear interpolation from measured CSA at the five sites in Fig. 3, a to e (procedure described in Supplementary Information, Section S1.3.3 and Supplementary Fig. 7). Model prediction agreed closely with the reconstructed waveform in overall morphology and the site-specific CSA changes explained its major features. At the beginning of Phase Ⅱ, arterial collapse at and around the PPZ produced an abrupt diastolic trough in the total arterial volume waveform once cuff pressure exceeded DBP (blue box, Fig. 3, h and i). From Phase Ⅱ to early Phase Ⅲ, complete PPZ occlusion had separated the proximal and distal sub-cuff arterial segments into distinct hemodynamic systems: pulsations at Sites ③ to ⑤ became progressively delayed and attenuated and then disappeared, whereas proximal pressure and volume pulsations persisted at Sites ① and ②. Loss of the distal volume contribution caused the delayed component on descending limb to disappear and the overall volume waveform to approach the morphology of the proximal volume waveform (red box, Fig. 3, h and i). At both DBP and SBP, these features reflected the summed vascular motion beneath the cuff at the same time point. Volume changes in the PPZ and adjacent proximal and distal segments are shown in Supplementary Fig. 8.

Figure 3, j and k compare the oscillogram calculated by the model with that derived from measured cuff pressure, showing close agreement in key features, including the DBP-and SBP- related morphological changes. These results indicate that model-predicted arterial volume change accurately reflects the actual mechanism of oscillogram formation and evolution during BP measurement (Supplementary Fig. 9).

### 4. Oscillogram morphology transitions identify DBP- and SBP-related vascular events

Based on the relationship among intra-arterial pressure, vascular-wall motion, and oscillogram morphology, we identified DBP-and SBP-related volume and hemodynamic changes and mathematically localized their oscillogram features. Local segment-replacement experiments further tested the causal contribution of dominant arterial segments to these transitions.

Figure 4a shows the dynamic relationships among CSA change, hemodynamics, and the oscillogram. When cuff pressure rose just above DBP, model-predicted (Fig. 3a) and IVUS- measured CSA showed marked diastolic collapse (blue arrow, Fig. 4, a and b), coinciding with an abrupt increase in negative oscillogram amplitude (orange arrow, Fig. 4, a and b). This collapse defined the DBP-related oscillogram transition. When cuff pressure was slightly below SBP, model-predicted and measured pressure waveforms showed increased delay and reduced amplitude of the PPZ-to-distal pressure component (Fig. 2, g and h), corresponding to waveform morphology changes in the oscillograms (blue box, Fig. 4a). Once cuff pressure exceeded SBP, the model and multimodal imaging showed complete PPZ occlusion. Loss of distal arterial pulsation eliminated the PPZ-to-distal contribution to oscillogram formation (Supplementary Fig. 8), producing a further main-peak transition (red box, Fig. 4a), typically including disappearance of a subtle descending-limb notch, main-peak narrowing, and reduced amplitude. This transition localized small-wave disappearance and was consistent with model predictions (representative waveforms are shown in Supplementary Figs. 12-19). Phase-specific oscillogram, pressure, and IVUS features are provided in Section S2.4 and Supplementary Fig. 10, with synchronized angiographic evidence in Supplementary Videos 9 and 10. Because these DBP-and SBP-related oscillogram features were transient local curvature changes, the Mexican-hat (Ricker) wavelet was selected to localize these morphology transitions across time and scale.

**Fig. 4.**
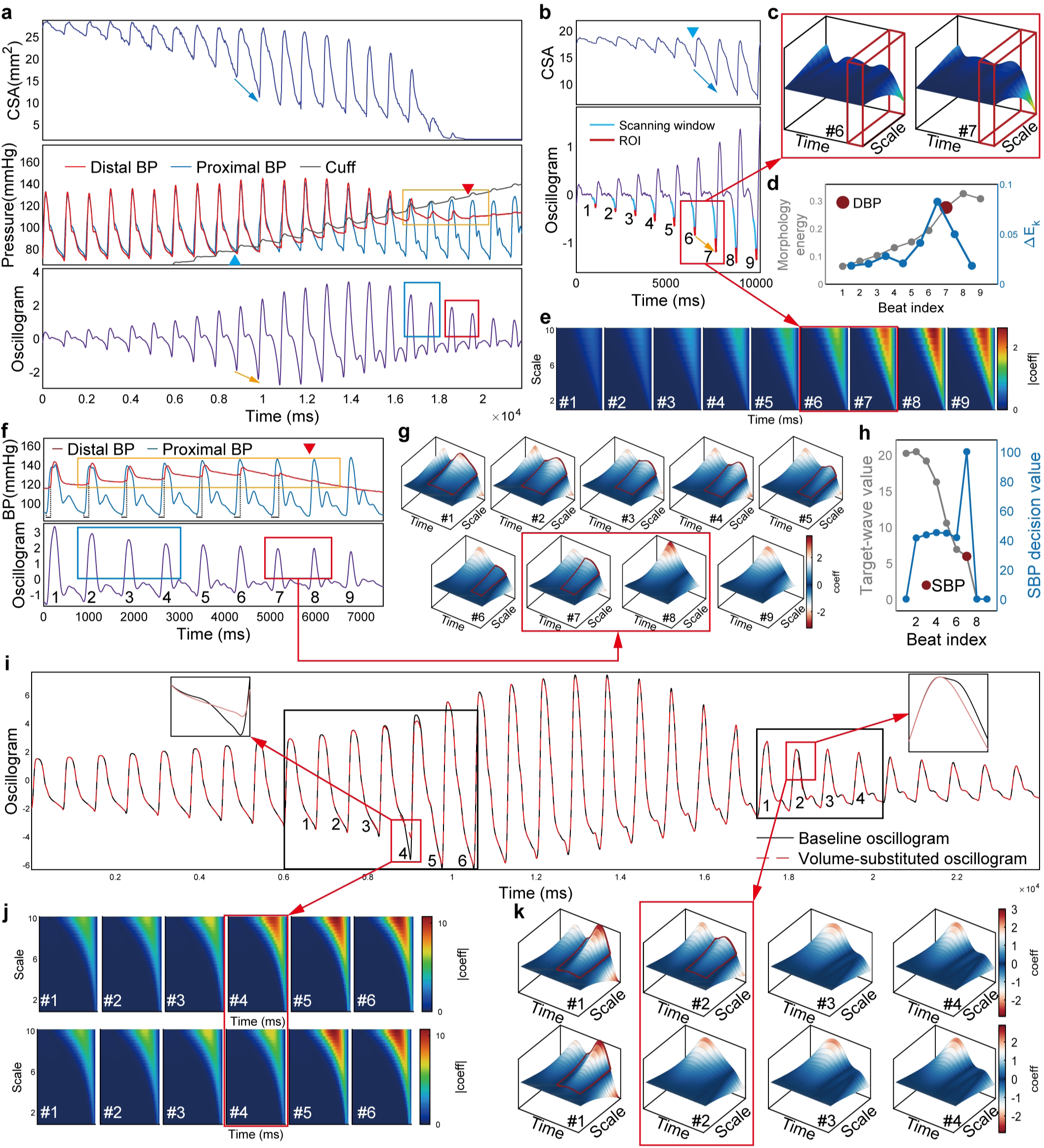
Extraction and validation of oscillometric morphological features. **a**, Synchronously recorded Site ④ CSA, proximal and distal invasive arterial pressures, cuff pressure, and oscillogram during cuff inflation. Blue arrows mark abrupt collapse near DBP, with orange arrows indicating the corresponding oscillometric change. Orange boxes highlight progressive small-wave attenuation/disappearance near SBP, while blue and red boxes indicate peak-side oscillometric shifts and oscillograms before and after small-wave disappearance, respectively. **b**, Representative CSA and oscillometric segment for DBP analysis; blue lines indicate the scanning window, red lines indicate the target ROI, and the blue triangle indicates the invasive DBP point. **c**, Wavelet-transform analysis of the two adjacent ROIs around the DBP point, shown as 3D time–scale–coefficient surfaces. **d**, Beat-to-beat morphology energy and interbeat change ΔEk for DBP transition detection (gray, morphology energy; blue, ΔEk; red point, DBP localization). **e**, Beat-to-beat time–scale coefficient map for DBP identification. **f**, Measured proximal/distal invasive pressure waveforms and oscillogram during small-wave appearance/disappearance; orange, blue, and red boxes are defined as in (**a**), and the red triangle marks the invasive SBP point. **g**, Beat-to-beat 3D time–scale–coefficient surfaces for SBP identification. **h**, Beat-to-beat small-wave tracking and SBP localization. **i**, Dominant-segment volume substitution comparing baseline and volume-substituted oscillograms. **j** and **k**, DBP-related coefficient maps and SBP-related 3D surfaces before and after volume substitution.

DBP identification required beat-by-beat assessment of oscillogram morphology within the region of interest (ROI) to localize the initial marked collapse of the PPZ. Figure 4b shows the temporal correspondence between diastolic brachial CSA collapse and the oscillogram and the wavelet scanning window. Across the two cardiac cycles immediately before and after the DBP identification point, the terminal portion of the wavelet-scanning result, defined as the ROI, showed marked enhancement of a small-scale local negative oscillatory component (red box, Fig. 4c), which was tracked beat by beat (Fig. 4, d and e). For the two beats showing the largest change (red boxes, Fig. 4, c and e), cuff pressure at the onset of the later beat (red dot, Fig. 4d) corresponded to the vascular-collapse transition in Fig. 4b.

SBP identification required localization of the pulsatile component distal to the PPZ. As this component became progressively delayed relative to proximal flow (orange box, Fig. 4, a and f), it separated from the main oscillogram peak, altered peak morphology, and eventually appeared as a subtle descending-limb notch (blue box, Fig. 4, a and f); its disappearance indicated complete arterial occlusion (red box, Fig. 4, a and f). Wavelet transform was therefore used to track this delayed component in the latter part of the oscillogram (Fig. 4g). Before cuff pressure exceeded SBP, residual distal systolic pulsation generated a small-scale local oscillatory component after the main peak (red boxes, Fig. 4g, #1 to #7). Once cuff pressure exceeded SBP, PPZ occlusion abolished distal pulsation, and this component disappeared between #7 and #8 (red box, Fig. 4g). In some measurements, the small-wave-related component disappeared directly; in others, a distinguishable reflected-wave component emerged after its disappearance (Supplementary Information, Section S2.4). The abrupt changes in local wavelet energy and temporal delay marked disappearance of residual trans-cuff flow (red dot, Fig. 4h). The preceding beat was the final beat with residual blood flow, and cuff pressure at its end was taken as the SBP reading. Thus, despite the small distal volume pulsation near SBP, loss of this delayed component produced a marked beat-to-beat oscillogram transition.

Figure 4, i to k, provides a causal mathematical test of these features through local segment-replacement experiments within a hypothetico-deductive framework. While preserving cuff pressure and volume contributions from other axial segments, we locally replaced the dominant segmental volume component. At the beat where cuff pressure equaled DBP, the PPZ volume-pulsation segment identified by the model—undergoing rapid negative-transmural-pressure collapse and occupying approximately 20% of the sub-cuff length—was replaced by the corresponding non-collapsed segment from the preceding beat while preserving endpoint continuity. This altered the negative diastolic oscillogram component and shifted the maximal DBP-related wavelet transition to the next beat (Fig. 4j). For the beat immediately before cuff pressure exceeded SBP, removing the volume-pulsation component distal to the PPZ, which occupied approximately 50% of the sub-cuff length, changed the oscillogram morphology to resemble the subsequent beat and shifted the SBP identification point one beat earlier (Fig. 4k). These experiments demonstrate the decisive role of the dominant segment in DBP-and SBP- related oscillogram morphology transitions (Fig. 4i; Supplementary Information, Section S2.2.13).

Throughout the cuff-based BP measurement, multisite IVUS data, intra-arterial pressure, and oscillogram changes were consistent with model predictions. We therefore established the novel Morphology-Transition Oscillometry (MTO), which identifies DBP and SBP from local oscillogram morphology transitions using wavelet analysis. Mean arterial pressure (MAP)-related feature identification is described in Supplementary Information, Section S1.5.6 and Supplementary Fig. 11.

### 5. Mathematical modeling explains errors in auscultatory and amplitude-ratio BP measurement

#### Korotkoff sounds

According to Baranger’s research, KS occur during the earliest phase of arterial wall expansion and are closely related to wall expansion velocity ^34^. Because IVUS could not reliably determine the local wall segment beneath the stethoscope, we used CSA changes as an approximate substitute for local wall motion and analyzed the maximum CSA expansion rate at Site ⑥, closest to the stethoscope, in relation to KS generation.

In single-cycle analysis, model-predicted arterial expansion closely matched IVUS measurements at Site ⑥. Arterial expansion lasted an average of 134.89 ms, whereas the early rapid expansion period averaged 40.76 ms and coincided with detectable KS (Fig. 5a) (n = 25 participants; 46 measurements). Frame-by-frame IVUS images illustrated the corresponding arterial opening process (Fig. 5b). Across the complete cuff measurement, model-predicted and measured CSA expansion rates showed similar temporal patterns and were compared with KS (Fig. 5c). At the systolic and diastolic boundaries, detectable KS appeared only when the CSA expansion rate exceeded a certain level, whereas small arterial expansions produced no detectable KS. These observations suggested an expansion-rate threshold for KS generation, differing between Korotkoff phase I (K1)-and phase V (K5)-related phases.

**Fig. 5.**
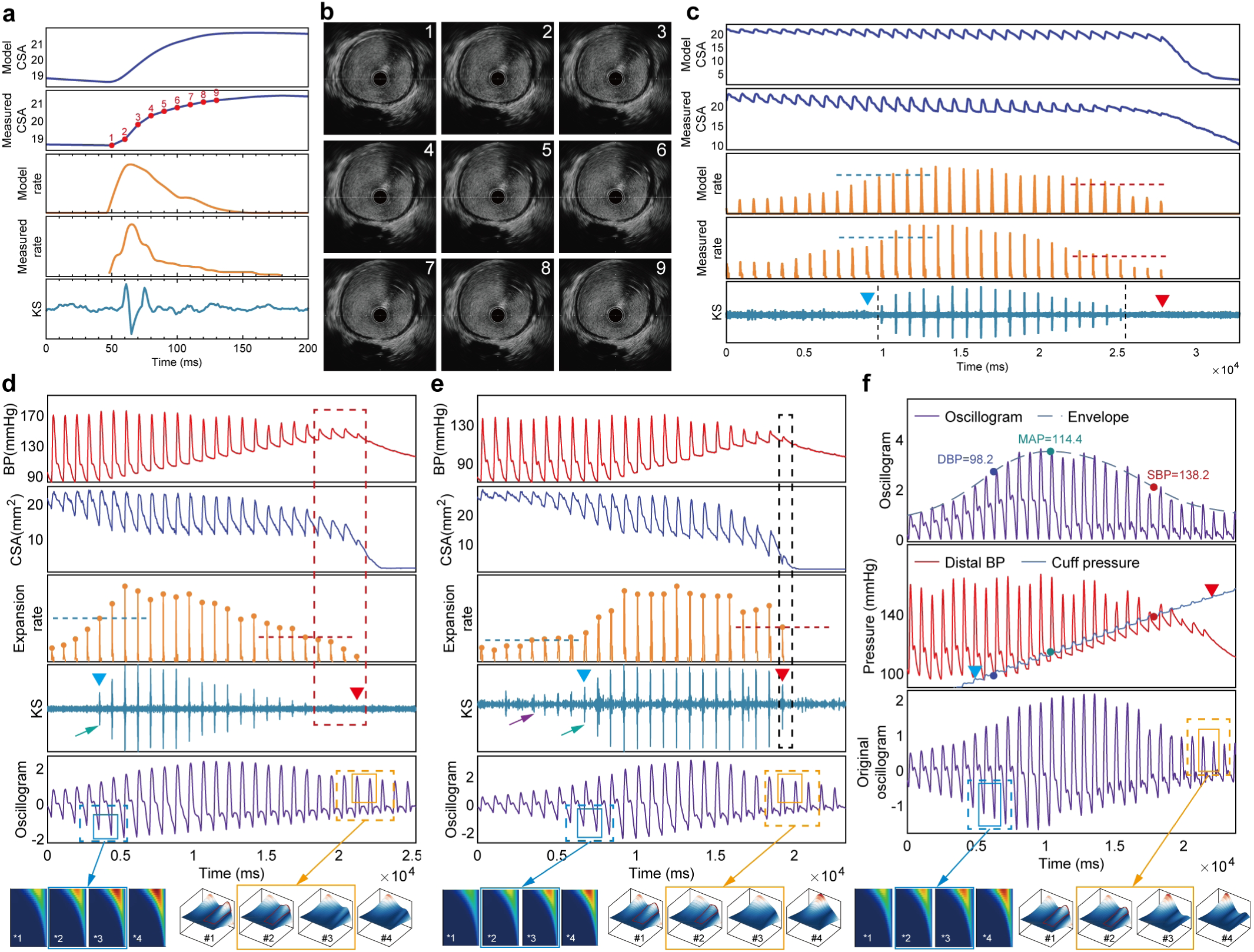
Mechanistic basis of errors in conventional cuff-based BP measurement. **a**, Single- beat rising-phase model-predicted and IVUS-measured CSA, CSA expansion rate, and synchronous Korotkoff sound (KS) at Site ⑥. **b**, Nine sequential IVUS frames; red dots in **(a)** indicate the corresponding sampling times. **c**, Complete cuff-inflation recording at Site ⑥ showing model-predicted and IVUS-measured CSA, CSA expansion rate, and KS. Blue and red dashed lines indicate expansion-rate thresholds for detectable KS, black dashed lines mark detected KS intervals, and blue and red triangles denote invasive DBP and SBP, respectively; these labels apply throughout Fig. 5. **d** and **e**, Examples of concordance and discordance between KS occurrence and trans-occlusion flow passage. From top to bottom: intra-arterial pressure at Site ⑤, co-localized IVUS CSA, CSA expansion rate, KS signal, and oscillogram. Distal pressure small waves and pulsatile IVUS CSA changes indicate trans-occlusion flow passage. Red boxes mark flow passage without detectable KS; black boxes mark KS–flow concordance near SBP. In **(d)**, the green arrow indicates KS onset coinciding with DBP; in **(e)**, purple and green arrows indicate KS onset and DBP, respectively. Lower rows show schematic wavelet-transform scanning, with blue and yellow boxes indicating DBP-and SBP-related feature changes. **f**, Examples of maximum-amplitude algorithm (MAA) readout and oscillometric waveform analysis from the same cuff-measurement run. From top to bottom: the detrended oscillogram and its envelope, with SBP and DBP determined by fixed envelope ratios and mean arterial pressure (MAP) assigned to the envelope peak; synchronously recorded distal intra-arterial pressure and cuff pressure; the raw oscillogram from the same cuff-measurement run; and the schematic wavelet-transform scanning of the oscillogram.

To characterize the relationship between KS and BP, multimodal measurements were performed at Site ⑤, where IVUS, invasive pressure, KS, and oscillogram were synchronously recorded, allowing comparison of CSA expansion rate with KS occurrence. In Fig. 5d, K5 coincided with DBP, whereas K1 did not reliably correspond to the SBP-related hemodynamic transition. Using K1 expansion rate as the SBP-related KS threshold showed that beat-by-beat peak expansion rate did not always reach detectable levels, particularly during the rounded, low-amplitude residual small waves of Phase Ⅱ. In contrast, MTO identified the prominent local morphological changes near DBP and SBP. The time–scale coefficient map in Fig. 5d identified the distal small-wave component and localized its disappearance. Figure 5e showed another pattern: K1 was relatively consistent with the SBP-related transition, but persistent KS near DBP prevented K5 from accurately identifying DBP, whereas MTO localized the DBP-related transition from the largest local oscillogram change. Across measurements, K5 occurred before, after, or within the same cardiac beat as invasive DBP, producing both over- and underestimation and substantial error dispersion. Near SBP, some terminal small-wave-related distal pulsations were too weak to generate detectable KS, making KS disappearance difficult to determine and causing K1-based SBP underestimation and systematic bias. Detailed statistics and individualized threshold analyses are provided in the Supplementary Information, Section S2.5, Supplementary Fig. 20 and Supplementary Video 8.

### Oscillogram amplitude ratios

Our theoretical derivation showed that maximum oscillogram amplitude, SBP, DBP, and their amplitude-ratio coefficients could be analytically derived from the mathematical model (Supplementary Information, Section S2.6). However, these coefficients are not fixed; they are determined by six coupled mechanistic domains: cuff loading state, vascular compliance, collapse sensitivity, residual transmission capacity, distal emptying and refilling state, and proximal–distal pressure propagation synchrony (Supplementary Table 4).

Consequently, individualized SBP and DBP amplitude ratios cannot be accurately represented by universal fixed coefficients. In practice, conventional amplitude-coefficient methods approximate SBP and DBP using preset empirical ratios on either side of the maximum amplitude. For maximum amplitude algorithm (MAA) comparison, fixed coefficients of 0.593 for SBP and 0.717 for DBP were used ^19^, within previously reported ranges of 0.45 to 0.73 for SBP and 0.69 to 0.83 for DBP. Fixed empirical coefficients cannot reliably correspond to true SBP and DBP across individuals and compressed-vessel states or explain their hemodynamic origin. By contrast, MTO localized the cardiac cycles containing the characteristic morphology transitions (Fig. 5f). MAA showed larger errors and greater error variability, with a representative SBP-overestimation case in Supplementary Fig. 24. Because traditional fixed ratios are often calibrated against auscultatory boundaries, which are themselves shifted by acoustic detection thresholds, fixed-ratio oscillometry may inherit this reference-standard shift as systematic error.

MTO localization more closely corresponded to the characteristic hemodynamic transitions; analyses of the same measurements shown in Fig. 5, d to f, are provided in Supplementary Figs. 21-23.

### 6. Paired statistical comparison of three methods against synchronized invasive BP

BP was measured in 175 participants and 630 measurements, with invasive SBP ranging from 98.08 to 203.22 mmHg, DBP from 46.72 to 121.53 mmHg, and MAP from 72.32 to 155.33 mmHg.

For SBP, MTO showed the smallest error and dispersion and did not differ significantly from invasive SBP (Bias = 0.39 ± 6.41 mmHg, mean absolute error (MAE) = 4.79 mmHg, P = 0.100). By contrast, auscultation and MAA systematically underestimated SBP (auscultation, −6.58 ±

10.30 mmHg, MAE = 9.67 mmHg, P < 0.001; MAA, −6.79 ± 10.58 mmHg, MAE = 10.25 mmHg, P < 0.001) (Fig. 6a). Violin plots showed fewer large errors and tighter distributions around zero with MTO whereas MAA showed extreme errors approaching 40 mmHg (Fig. 6b). For DBP, MTO again showed the smallest error and variability without a significant difference from invasive DBP (Bias = 0.14 ± 6.25 mmHg, MAE = 5.05 mmHg, P = 0.388). Auscultation showed a small bias but greater variability (Bias = −1.15 ± 7.24 mmHg, MAE = 5.63 mmHg, P < 0.001). MAA showed a larger positive bias (Bias = 4.85 ± 9.14 mmHg, MAE = 7.95 mmHg, P < 0.001) and occasional extreme errors > 30 mmHg (Fig. 6, a and c). For MAP, MTO showed no significant difference from waveform-integration MAP and the lowest variability (Bias = −0.37 ± 5.92 mmHg, MAE = 4.59 mmHg, P = 0.281). Cuff pressure at maximum oscillation amplitude showed greater variability and extreme individual errors exceeding 50 mmHg (MAA, 2.38 ± 11.56 mmHg, MAE = 8.78 mmHg, P < 0.001), while formula-derived auscultatory MAP also showed greater error dispersion (Bias = −6.77 ± 7.42 mmHg, MAE = 8.08 mmHg, P < 0.001) (Fig. 6, a and d). Bland–Altman and linear regression analyses are shown in Supplementary Fig. 25-26. The derivative-of-envelope method showed larger deviations from invasive BP, especially for SBP (Supplementary Information, Section S2.7).

**Fig. 6.**
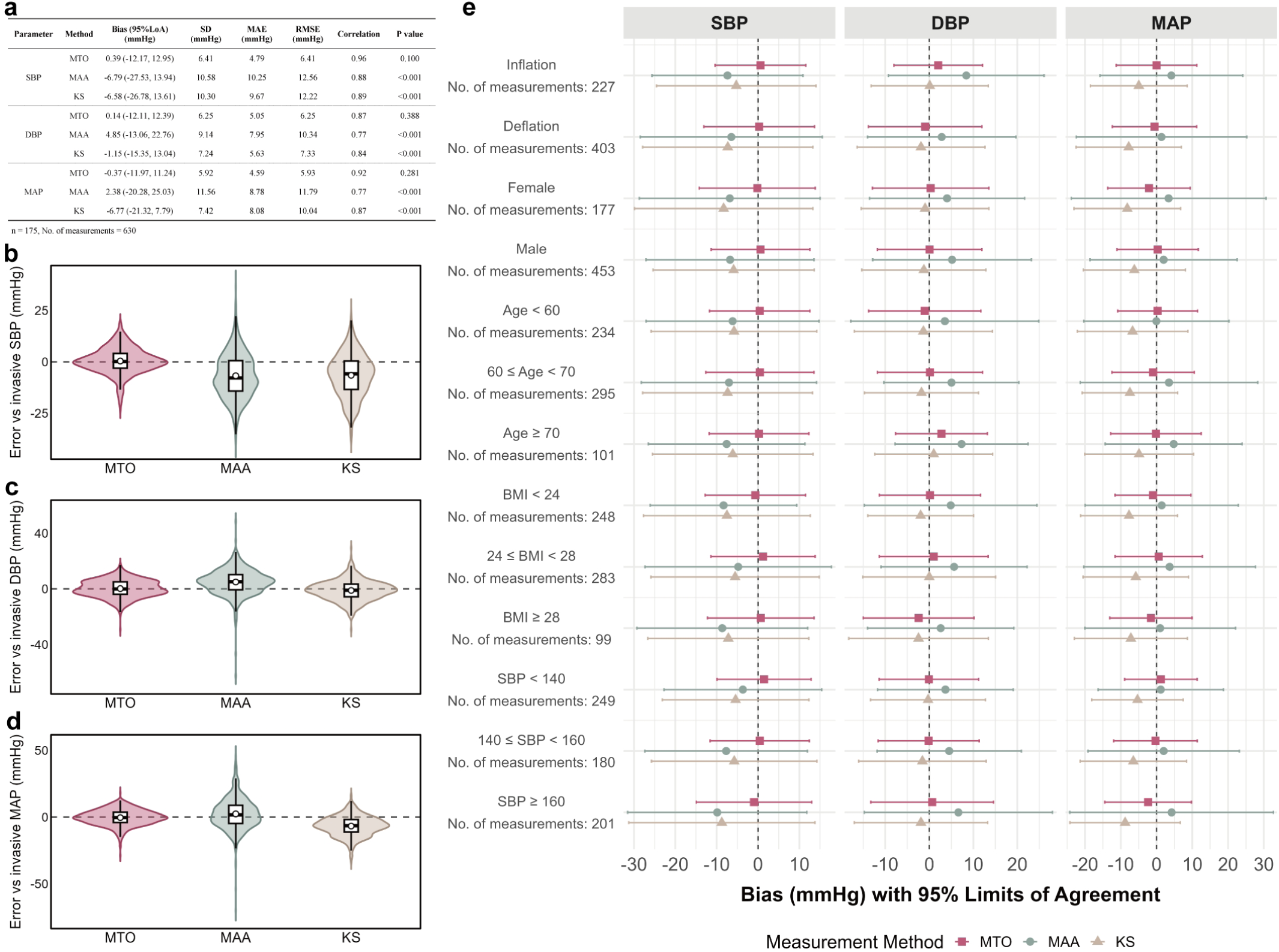
Accuracy assessment and subgroup agreement analysis relative to IBP. **a**, Performance comparison of KS auscultation, MAA, and Morphology-Transition Oscillometry (MTO) for SBP, DBP, and MAP measurement using IBP as the reference standard; participant numbers and measurement counts are shown in the table. Metrics include bias, 95% limits of agreement (95% LoA), standard deviation (SD), mean absolute error (MAE), root mean square error (RMSE), correlation coefficient, and P values from two-sided Wilcoxon signed-rank tests comparing noninvasive and invasive BP. **b** to **d**, Violin plots of SBP, DBP, and MAP errors (noninvasive minus invasive) for MTO, MAA, and auscultation. Violin width indicates density; open circles indicate means, center lines medians, boxes interquartile ranges, whiskers 1.5× interquartile ranges, and gray dashed lines zero error. **e**, Forest plots for subgroup agreement analysis showing the bias and 95% LoA of auscultation, MAA, and MTO relative to IBP across measurement phase, sex, age, body mass index (BMI) and BP level subgroup, with the number of measurements labeled for each subgroup.

Across predefined subgroups by inflation or deflation mode, sex, age, body mass index (BMI), and BP level, MTO maintained small bias, lower dispersion, and narrower 95% limits of agreement (95% LoA) (Fig. 6e). Repeated-measures sensitivity analyses confirmed these findings, with MTO retaining minimal bias and narrower 95% LoA for SBP, DBP, and MAP than auscultation and MAA. MTO also showed the lowest pooled within-participant SD of method-specific errors for SBP, DBP, and MAP (3.93, 3.78, and 3.96 mmHg, respectively), supporting closer agreement with invasive BP and greater stability across repeated measurements within the same participant. Additional statistical results are provided in Supplementary Information, Section S2.7, Supplementary Fig. 27 and Supplementary Tables 5–10.

## Discussion

This study began with the fundamental question of how the cuff acts as an external compression source. Treating the cuff as a membrane governed by membrane theory and a Laplace-type normal force balance, we derived a hyperbolic-cosine-like axial pressure distribution transmitted through tissue to the brachial artery wall. By coupling the external cuff pressure field with intra-arterial pressure, we formulated a mathematical cuff–artery coupling model and derived a set of governing equations that systematically predict vascular motion and hemodynamic changes in the sub-cuff brachial artery, as well as the resulting evolution of oscillogram morphology during BP measurement, and validated these predictions using in vivo multimodal measurements in supine participants. The model identified the PPZ near the cuff center and corresponding to the minimum-curvature region of the sub-cuff brachial artery, as the driving region of vascular and hemodynamic changes. When cuff pressure first exceeded DBP, diastolic arterial collapse first occurred at the PPZ, separating the proximal and distal sub-cuff arterial segments into distinct hemodynamic systems. With further pressurization, the diastolic occlusion zone extended from the PPZ toward both sides, producing diastolic occlusion and systolic re-expansion within this region. When cuff pressure approached SBP, limited blood flow traversed the PPZ for several cardiac cycles, increasing proximal–distal delay and producing delayed small waves. When cuff pressure exceeded SBP, the PPZ became completely occluded and distal vascular pulsation disappeared (Figs. 2 and 3; Supplementary Videos 1–7). These mechanical events explain characteristic oscillogram morphology transitions. Within this mechanistic framework, wavelet transform localized DBP-related local diastolic collapse and SBP-related complete occlusion, providing localization features for BP estimation.

Beyond direct experimental validation, we used a physics-based hypothetico-deductive perturbation approach to selectively remove model components from the dominant vascular segments responsible for DBP- or SBP-related oscillogram features; the wavelet-identified DBP and SBP positions shifted accordingly. Our mechanistic framework systematically explains vascular and hemodynamic changes during BP measurement, establishes a theoretical basis for BP measurement, and identifies a new BP measurement method anchored to hemodynamic changes. In 175 participants, MTO showed greater accuracy and precision than clinically used auscultation and fixed-amplitude-ratio oscillometry and no significant difference from invasive pressure (Fig. 6). To our knowledge, this is the first study to derive a series of cuff-artery coupling equations that establish this complete mechanistic chain linking cuff external-pressure distribution, vascular transmural pressure, hemodynamic response, and oscillogram morphology transitions. It further applies these hemodynamic transitions as the localization logic for BP measurement, with consistent measurement advantages across the whole cohort and major subgroups (Fig. 6, Supplementary Figs. 25 and 26, and Supplementary Tables 5–10).

Cuff-based BP measurement was introduced by Riva-Rocci in the late 19th century using the palpable pulse method ^35^, followed by the Korotkoff auscultation. Both approaches assume that the first perceptible signal during cuff deflation—a tactile pulse or Korotkoff sound (K1)—marks initial blood passage through the compressed brachial artery and SBP. Auscultation further provides a diastolic endpoint, with disappearance of Korotkoff sounds (K5) taken to mark DBP ^36^. It is true that our measurements related SBP to the final blood passage during cuff inflation. However, this event was marked not by an audible or palpable signal, but by the distal sub-cuff pressure small wave, whose final occurrence is relatively weak and may escape auscultatory or palpatory detection. The mechanism of KS generation has been debated for over a century, with proposed explanations including water-hammer phenomena, the preanacrotic wave, cavitation bubbles, flow-instability-induced wall vibration, pulse-wave distortion, and dynamic arterial wall instability ^37–45^. Baranger et al. used ultrafast ultrasound to show that KS arise from rapid motion of the compressed arterial wall transmitting shear vibrations to the stethoscope ^34^. Consistently, our model calculations and experimental observations both showed that detectable KS require sufficient arterial expansion rate and therefore depend on individualized thresholds. Near SBP, terminal small waves are flat and produce weak, slow expansion that often remains below this threshold, explaining systematic SBP underestimation and error dispersion with auscultation, and the greater underestimation by less-sensitive palpation. MTO has the further advantage that the cuff records pulsations from the sub-cuff arterial segment on the distal side of the PPZ over approximately half of the cuff length, whereas palpation and auscultation detect only weaker signals after propagation beyond the cuff. MTO instead captures the temporal separation of small-wave-related components near SBP, allowing clear localization of small-wave disappearance. At DBP, K5 is likewise constrained by individualized acoustic thresholds and may deviate from the first collapse-related event, whereas MTO directly localizes this initial oscillogram transition. Notably, our auscultatory analysis used visually displayed and frequency-filtered signals; true human-ear auscultation may produce larger errors. Beyond the traditional vascular opening–closing interpretation, the model links cuff BP measurement to local transmural pressure, vascular-volume change, and hemodynamic evolution during compression. If individual KS-generation thresholds can be defined, our mathematical model may further predict KS appearance from simulated distal arterial area dynamics.

In 1904, Nobel laureate Joseph Erlanger combined the Riva-Rocci cuff with a recording apparatus to capture cardiac-cycle-related oscillations and attributed the oscillation amplitude to arterial volume change under compression ^46^. In 1969, Posey et al. first established that maximum cuff oscillation approximates MAP ^16,29^. Building on this observation, Geddes et al. first proposed an empirical amplitude-ratio method for estimating SBP and DBP relative to the maximum cuff-oscillation amplitude. Drzewiecki et al. reported and validated specific coefficients of 0.593 for SBP and 0.717 for DBP, and this empirical ratio framework was subsequently translated into fixed, device-specific coefficients ^19,47^. Conventional models derived oscillation amplitude from arterial volume changes under effectively uniform external cuff pressure ^16^. Later models retained this simplification, representing cuff–artery interaction with a single external-pressure variable to reproduce the oscillogram envelope and optimize MAA-based BP estimation ^12–15^. However, this approach overlooked experimentally observed nonuniform axial compression and could not fully simulate true sub-cuff hemodynamics; consequently, current MAA methods still rely on empirical ratios ^24–27^. Subsequent studies introduced patient-specific model fitting, analytical reformulation, and optimized arterial-compliance functions, but remained centered on oscillogram envelopes, multi-parameter estimation, and a single external-pressure representation that did not resolve axial sub-cuff pressure heterogeneity ^12–15^. Theoretically, MAP, SBP, and DBP can be analytically linked, and their corresponding ratio coefficients can be mathematically derived through our model.

However, this derivation depends on six categories of highly individualized and variable parameters that cannot be directly measured (Supplementary Table 4). MAA therefore relies on fixed empirical coefficients that cannot universally accommodate different populations or measurement conditions, contributing to substantial error dispersion ^10,48^. Celler et al. similarly reported SBP underestimation of 18.2 ± 13.2 mmHg with conventional oscillometry versus invasive pressure ^49^. Auscultation-derived coefficients may additionally propagate auscultatory bias into oscillometry. In contrast, MTO is mechanism-based. For SBP, absolute errors greater than 10 mmHg accounted for only 11.75% of MTO measurements, compared with 46.51% of MAA measurements. Violin plots showed broader MAA error distributions than those of auscultation and especially MTO (Fig. 6b-d), with maximum MAA SBP errors approaching 40 mmHg. These approaches also extract only amplitude-envelope information from the oscillogram. Our study mathematically defined and experimentally validated the curvilinear distribution of cuff external pressure, resolving the long-standing discrepancy between legacy model calculations and physical evidence. Built upon first-principles mechanics, our framework defines pressure relationships among the cuff, tissue, and brachial artery, predicts vascular motion, hemodynamic transitions, and oscillogram morphology, and applies these insights to BP localization.

MTO was the only method whose BP estimates did not differ significantly from invasive reference values, indicating higher accuracy and precision. With the increasing use of wearable health monitoring and AI-enabled physiological assessment, BP measurement increasingly requires non-auscultatory, rapid, and reliable methods. BP can be inferred from beat-by-beat oscillogram morphology when cuff pressure corresponds to SBP or DBP. This beat-specific localization also suggests that MTO could be extended to short-term dynamic BP monitoring, irregular and unstable hemodynamic states such as arrhythmia. MTO therefore has the potential to replace auscultation, which remains a required noninvasive reference under regulatory requirements and professional standards such as the ISO 81060 series ^48,50^, and may serve as a candidate reference method for NIBP measurement. More broadly, this modeling concept may support analysis of other dynamic vascular states, including pressure-waveform-based assessment of vascular stenosis, coronary pressure-waveform morphology, and hemodynamic modeling in other vascular systems. By transforming cuff BP measurement from an empirical approximation into a mechanism-based mathematical framework, this study exemplifies how practical measurement methods can be translated into mathematical–physical mechanisms for physiological application.

### Limitations

This study has three main limitations. First, participants with invasive BP references were recruited from catheterization-laboratory populations, limiting population representativeness and requiring further validation in broader community-based and primary-care populations. Second, for ethical reasons, patients with extreme BP values had to receive treatment first to achieve a relatively stable state before study measurements, which limited the number of cases with extreme BP values and may restrict the evaluation of MTO performance at the outer ranges of BP. Third, all measurements were performed supine because of catheterization-laboratory constraints; therefore, synchronized invasive reference data in the standard seated BP measurement posture were unavailable.

## Conclusion

By integrating membrane theory with Laplace-law-based mathematical and physical principles, this study establishes a unified framework for cuff-based BP measurement that resolves prior mechanistic inconsistencies, predicts the dynamic transformation from intra-arterial pressure to vascular volume change and oscillogram morphology, and experimentally validates this framework through multimodal measurements. On this basis, we developed a BP measurement method consistent with hemodynamic events, which employs wavelet-transform-based morphology tracking to identify the vascular transitions corresponding to SBP and DBP, achieving higher accuracy than conventional methods.

## Methods

### Study design and participants

This prospective human mechanistic study of cuff-based BP measurement comprised three coordinated cohorts. Cohort 1 consisted of patients undergoing cardiac catheterization and provided invasive arterial pressure recordings, brachial venous pressure measurements, vascular imaging, and synchronized cuff, oscillometric, and KS signals. Cohort 2 comprised healthy participants studied with noninvasive cuff-skin pressure mapping, and Cohort 3 comprised healthy volunteers undergoing upper-limb computed tomographic angiography (CTA) during controlled cuff compression (Supplementary Fig. 1).

For Cohort 1, 182 consecutive patients undergoing cardiac interventional procedures at the First Affiliated Hospital of China Medical University between July 2024 and July 2026 were prospectively screened. Eligible participants were older than 18 years and younger than 80 years and had complete baseline demographic and clinical information. Exclusion criteria were acute myocardial infarction requiring primary percutaneous coronary intervention, hemodynamic instability or cardiogenic shock, congestive heart failure, unrevascularized triple-vessel coronary artery disease, left-main coronary artery disease, severe brachial artery stenosis or occlusion caused by peripheral vascular disease, dialysis dependence, or refusal to participate. Four screened patients were excluded because of significant arrhythmia, poor KS detectability, inability to tolerate the procedure, or incorrect arterial access. Cohort 1 therefore included 178 patients: 175 contributed invasive arterial pressure recordings and 3 underwent venous pressure measurements only. A subset of 37 participants with 124 synchronized measurements was used to characterize arterial pressure changes during cuff BP measurement. No patient underwent cardiac catheterization solely for this study.

Sample size for the invasive-reference cohort was determined from pilot data using paired two one-sided equivalence tests with prespecified equivalence bounds of −5 to 5 mmHg, a one-sided significance level of 0.05, and 95% power. The calculated requirements were 64 participants for SBP and 120 for DBP; allowing for up to 20% nonevaluable enrollments yielded a target of at least 150. The final invasive-reference cohort comprised 175 participants and 630 paired measurements.

Cohort 2 included 50 healthy participants. Cohort 3 included 22 healthy volunteers aged 18–60 years with BMI 18–26 kg/m² and no contraindication to CT examination. Baseline characteristics are provided in Supplementary Table 1.

### Ethics and informed consent

All study procedures were approved by the Ethics Committee of the First Affiliated Hospital of China Medical University, Shenyang, China (approval no. [2026] 2024-609-5), and were conducted in accordance with the Declaration of Helsinki. Written informed consent was obtained from participants in the cardiac-catheterization and healthy-volunteer CTA cohorts before enrollment. The noninvasive pressure-mapping study was approved under the same institutional protocol. No procedure performed solely for research altered the standard clinical care delivered to patients in the catheterization cohort.

### Integrated instrumentation and synchronized acquisition

The experimental platform incorporated a HIERS multichannel physiological recorder (Shanghai Hongtong Industrial), a custom cuff-pressure control system, COMET II pressure guidewires with an FFR Link interface (Boston Scientific), electronic auscultation, digital subtraction angiography, and a 60-MHz IVUS system (VH-60; Shenzhen Insight Lifetech). Devices were synchronized through a common trigger pulse. Commercial upper-arm cuffs (OMRON HEM-

CR24 Type B or GS CUFF2, size M) were positioned with the lower cuff edge 1–2 cm above the antecubital fossa and the artery marker aligned with the brachial artery. Cuff pressure was controlled by a low-vibration closed-loop pneumatic system. Full hardware specifications are provided in Supplementary Information, Section S1.2.

### Cuff-pressure measurement protocols

Cuff measurements in the catheterization cohort were performed supine. During inflation-mode measurements, the cuff was inflated at 3–5 mmHg/s from baseline to 30 mmHg above invasive SBP and then rapidly released. During deflation-mode measurements, the cuff was rapidly inflated to 30 mmHg above invasive SBP and then deflated at 3–5 mmHg/s. Only the slow approximately linear measurement phase was used for BP determination and waveform analysis. Two inflation-mode and two deflation-mode measurements were obtained in the primary synchronized protocol, with at least 1 min of rest after each run.

Five complementary catheterization protocols were used. The primary synchronized protocol included 92 patients and recorded invasive arterial pressure at the distal-internal sub-cuff site (Site ⑤), with an optional proximal edge reference (Site 0), together with cuff pressure, oscillograms, and KS. Site-specific IVUS measurements were obtained in 28 patients. Additional protocols covered multisite arterial and venous pressure mapping. A fluid-filled catheter protocol at Site ⑤ was used in 75 patients, each undergoing three repeated deflation measurements.

Detailed configurations are provided in Supplementary Information, Section S1.6.

### Invasive arterial-pressure measurement and reference definition

Pressure guidewires were flushed, zeroed, and equalized against the same fluid-filled catheter reference before advancement to the target site. When multiple guidewires were used, all sensors were compared at the same vascular position before recording; if concordance was not achieved, zeroing and equalization were repeated. Fluid-filled catheter systems were prefilled and deaerated, the transducer was positioned at right-atrial level, and the system was zeroed to atmospheric pressure. Dynamic response was assessed in vivo by fast-flush testing; the measured natural frequency was 14.29 Hz and the damping coefficient was 0.474 ^51^.

For each cuff measurement, sub-cuff intra-brachial arterial pressure served as the run-specific invasive reference. The reference window was selected from a continuous, analyzable, artifact-free arterial pressure segment of approximately 30 s within 1 min before cuff inflation, whenever available, with preference given to the segment closest to cuff inflation. Selection was based on signal quality and temporal proximity and was independent of the result of any noninvasive BP method. Invasive SBP and DBP were defined as the means of beat-wise systolic maxima and diastolic minima, respectively. Beat-wise MAP was calculated as the time integral of the pressure waveform over each cardiac cycle divided by cycle duration, and run-specific MAP was the mean of these values across the reference window.

### Multisite arterial-pressure measurements

Multisite arterial pressure was recorded in 8 patients using five or six pressure guidewires at predefined axial locations. Whole-cuff configurations sampled the proximal edge and internal sub-cuff sites; local configurations sampled the PPZ. Signals were acquired simultaneously during two inflation and two deflation runs at the predefined positions. Detailed sensor configurations and additional proximal arterial recordings are provided in Supplementary Information, Section S1.6.3.

### Multisite brachial venous-pressure measurements

Brachial venous pressure was measured in five participants, including two who also underwent arterial pressure mapping and three with venous recordings only. Pressure guidewires were positioned at predefined sub-cuff venous sites during cuff inflation and deflation. Whole-cuff and central PPZ configurations were used, as detailed in Supplementary Information, Section S1.6.4. Available venous-pressure channels were recorded synchronously within each run.

Venous pressure was used as a surrogate of cuff-transmitted deep-tissue and perivascular pressure rather than as an arterial BP reference ^52,53^. Arterial and venous measurements, when both obtained, were performed sequentially rather than simultaneously.

### Brachial angiography

Brachial angiography was performed in 22 patients. The examined arm was positioned to allow stable visualization of the cuff-covered segment while minimizing motion and direct external compression. The catheter tip was positioned at the proximal end of the cuff-covered segment rather than within the compressed segment, and selective contrast injection was used to visualize arterial morphology during cuff pressurization. Depending on the combined procedure, radial or femoral access was used. Because of technical limitations, angiography and IVUS were acquired in separate cuff-measurement runs when both were required.

### IVUS acquisition and CSA analysis

IVUS-synchronized intravascular pressure measurements were performed in 28 patients with a 60-MHz IVUS catheter operating at 100 frames/s. During each BP measurement run, the catheter was maintained at a fixed vascular position, and continuous imaging was acquired throughout the cuff-pressure change. After measurements at one site were completed, the catheter and co-localized pressure guidewire were repositioned to the next predefined site.

For continuous CSA measurement, 30 sampling points were selected within each analyzed cardiac cycle at approximately equal temporal intervals, with both maximal and minimal CSA included. The luminal boundary was delineated along the inner intimal contour. At least two trained clinicians independently analyzed the same segment, with additional specialist review; measurers and reviewers were blinded to site identity and acquisition order. Quantitative stage-specific CSA analyses used 46 measurements from 25 participants.

### Multisite IVUS reconstruction

Whole-segment luminal volume was reconstructed from CSA waveforms obtained at five axial IVUS sites during separate matched cuff measurements. Cuff pressure relative to invasive DBP and SBP was used as the common reference, and corresponding cardiac cycles were normalized in duration. Axial delays were corrected using electrocardiogram (ECG)-derived pulse-arrival timing, with Site ① used as the temporal reference. After temporal alignment, CSA waveforms were linearly interpolated along the vessel axis and integrated to obtain total sub-cuff luminal volume.

Four participants additionally underwent cuff-inflation measurements with IVUS acquisition at 13 evenly distributed sub-cuff locations. Frame-wise vascular contours were manually delineated, and an AI-assisted segmentation workflow generated candidate masks for contour regularization and quality control. Frame indices were combined with pressure and ECG timing for inter-site alignment, and spatially ordered contours were interpolated using a signed-distance-function method to generate time-resolved 3D representations. Full procedures are provided in Supplementary Information, Sections S1.3.3 and S1.3.4.

### Surface pressure mapping

Cuff-skin contact pressure was measured in 50 healthy participants using a flexible pressure-sensing pad containing 1,024 sensing points arranged in a 32 × 32 matrix over a 200 × 200 mm active area, sampled at 100 Hz. The pad was positioned between the cuff and upper-arm skin during controlled cuff inflation. Frame-wise sensor arrays were reconstructed into two-dimensional pressure maps, and axial pressure profiles were obtained by averaging across the transverse dimension. A three-point moving-average smoothing step was applied to the axial profile only for visualization; unsmoothed profiles were retained for quality control and model comparison.

### CTA acquisition and reconstruction

Upper-limb CTA was performed in 22 healthy volunteers at predefined cuff-pressure levels spanning below DBP to above SBP, with retrospective ECG gating. Brachial cuff BP was measured three times before scanning, and the mean DBP and SBP values were used as references for cuff-pressure settings. Cuff pressure was maintained at the assigned level during each acquisition. The complete pressure protocol and numbers of acquisitions at each level are provided in Supplementary Information, Section S1.7.2 and Supplementary Fig. 1. Imaging used a photon-counting CT system (NeuViz P10; Neusoft Medical Systems) at 120 kVp, rotation time 0.27 s, pitch 0.15, and collimation 584 × 0.274 mm. Images were reconstructed at 33 phases of the R–R interval with 0.274-mm slice thickness and 0.137-mm increment. Cuff and brachial artery segmentation used a semi-automated AI-assisted workflow followed by manual review before 3D rendering.

### Korotkoff-sound measurement

KS were recorded synchronously with cuff and invasive pressure signals using an electronic auscultation module. The chest piece was placed over the point of strongest brachial pulsation distal to the cuff and fixed without adding direct compression to the artery. KS signals were represented in low-(20–50 Hz), medium- (50–150 Hz), and high-frequency (150–900 Hz) bands; when separation was inadequate, the bands were adjusted to maintain a three-band representation ^54–56^. Visual assessment was primary and auditory review supportive. Two senior clinicians independently identified systolic and diastolic boundaries while blinded to other experimental data except measurement direction; disagreements or visual SBP estimates lower than auditory estimates were reviewed by a third physician. Auscultatory MAP was estimated as DBP + (SBP − DBP)/3 using SBP and DBP from the same cuff run.

### Conventional oscillometric methods

For the fixed-ratio MAA, the oscillogram envelope was constructed after waveform extraction. MAP was assigned to the cuff pressure at the envelope maximum, and fixed amplitude coefficients of 0.593 for SBP and 0.717 for DBP were applied ^19^. A derivative-based oscillometric method was also evaluated as a supplementary empirical comparator and assigned BP landmarks according to maximal first-derivative changes in the corresponding oscillometric feature trajectory.

### Oscillometric signal processing

Cuff-pressure signals were processed with a third-order Butterworth band-pass filter with a passband of 1–15 Hz to extract the oscillometric waveform ^57^. The extracted waveforms were used for wavelet analysis, envelope analysis, and model-related comparisons. No additional filtering or smoothing was applied to the original invasive arterial pressure waveforms used for physiological interpretation. Signal processing and model calculations were implemented in MATLAB R2023a (MathWorks).

### Morphology-Transition Oscillometry

MTO was implemented as a semi-automated method that identified DBP, MAP, and SBP from beat-to-beat changes in oscillogram morphology. For SBP and DBP localization, oscillograms were segmented by cardiac cycle and analyzed with a Mexican-hat (Ricker) continuous wavelet transform using dyadic scale sampling ^58,59^. The method localized pressure-dependent morphology transitions corresponding to vascular events identified by the multimodal experiments rather than fixed oscillation-amplitude thresholds.

For DBP localization, analysis was restricted to the terminal downstroke immediately preceding the beat valley. A 120-ms terminal window and a 40-ms valley-proximal ROI were used in the primary implementation. Wavelet coefficients were summarized into a local morphology-saliency measure, and the largest absolute change between adjacent beats defined the DBP-related transition. During cuff inflation, DBP was assigned to cuff pressure at the onset of the later beat in the transition pair.

For SBP localization, the primary above-baseline component of each beat was analyzed. Positive wavelet-coefficient hotspots on the right side of this component were tracked across consecutive beats according to temporal and scale continuity, local morphology similarity, and relative amplitude. During cuff inflation, the SBP-associated beat was the final beat before the tracked small-wave-related morphology entered its terminal stable low-value state. Cuff pressure at the end of that beat was assigned as SBP. MAP was localized from crossover of an early dicrotic-like and a later reflection-like feature on the descending limb using derivative-defined feature prominence. The mean cuff pressure of the crossover beat was assigned as MAP. For each measurement, algorithm-generated candidate SBP and DBP transitions were verified by at least two investigators familiar with the characteristic MTO features and blinded to invasive reference BP. Mathematical definitions are provided in Supplementary Information, Section S1.5.

Complete implementation code and numerical settings will be provided with the submission.

### Cuff membrane model

The cuff-arm system was modeled as an axisymmetric thin membrane supported by deformable soft tissue. The analytical formulation assumed quasi-static inflation, continuous contact within the effective loaded width, an equivalent cylindrical arm geometry, linear local tissue support of Winkler type, approximately uniform effective membrane-force resultants, and small-slope deformation ^2,3,31,60–63^. The governing equation and normalized axial pressure distribution are given in equations (1) and (2), respectively. Zero displacement was prescribed at the effective cuff edges, with symmetry at the cuff center. Contact pressure was normalized by its central value. The dimensionless structural parameter β combines tissue support, membrane-force coupling, cuff width, and edge constraints. The analytical model characterizes the relative axial pressure profile rather than its absolute magnitude. Full derivation and limitations are provided in Supplementary Information Section S2.1.

### Cuff-brachial artery model

The membrane-derived normalized pressure profile was multiplied by cuff pressure to define the local external load in a reduced-order cuff-brachial artery model. Local intraluminal pressure, Pint(z,t), entered the transmural-pressure definition and open–closed area relation given in equations (3) and (4). Proximal arterial pressure was supplied as a measured invasive waveform, a standardized arterial-pressure template, or a constant-pressure control. Pressure propagation was represented by site-to-site delay, with additional delay developing as local compression increased. Positive-transmural-pressure area followed a Langewouters-type pressure-area relation ^64–66^. Negative transmural pressure was represented by a hybrid collapse model incorporating geometric collapse, closure memory, residual patency, and a functional conductance threshold ^65–67^. When proximal-distal transmission became restricted, distal pressure evolved toward an effective reservoir according to a lumped resistance-compliance representation ^12,15,68^.

Under severe compression, mechanism-inspired pressure-field-remodeling terms represented distal pressure enhancement, a proximal mirror-like response, and localized DBP- and SBP-related pressure structures ^69–71^. These terms entered the intraluminal pressure field before calculation of transmural pressure and vascular area and were not applied as oscillogram post-processing. Local arterial areas were integrated along the cuff-covered segment to obtain total arterial volume, which was converted to recovered cuff pressure using an air-chamber volume-pressure readout. Representative simulations used a 120-mm modeled segment discretized into 30 axial sites, β = 3.5, a baseline pulse-wave velocity of 6 m/s, and a numerical sampling frequency of 1,000 Hz. Except for explicit inputs and protocol conditions, representative parameters were kept fixed across simulations. Measured multisite IVUS CSA sequences were also passed through the same interpolation, volume-integration, and cuff-readout framework, and local volume perturbations tested PPZ and distal contributions to DBP- and SBP-related oscillogram transitions. Complete equations, parameter definitions, and numerical settings are provided in Supplementary Information, Section S2.2 and Supplementary Tables 2 and 3.

### Statistical analysis

The principal invasive-reference analyses used 630 paired measurements from 175 participants. Mechanistic pressure analyses used 124 synchronized measurements from 37 participants, and stage-specific IVUS analyses used 46 measurements from 25 participants. The three venous-only participants were included exclusively in the venous pressure substudy. Measurement error was defined as the noninvasive estimate minus the invasive reference.

Primary agreement analyses were performed at the measurement level and evaluated separately for SBP, DBP, and MAP using bias, SD of the differences, 95% LoA (bias ± 1.96 × SD), MAE, RMSE, and Pearson correlation coefficients. Two-sided one-sample Wilcoxon signed-rank tests were used to test whether measurement errors differed from zero. Subgroup analyses were performed at the measurement level according to measurement phase, sex, age (<60, 60–69, or ≥70 years), BMI (<24, 24–27.9, or ≥28 kg/m²), and concurrent invasive SBP (<140, 140–159, or ≥160 mmHg). Mann-Whitney U tests were used for two-level comparisons and Kruskal-Wallis tests for three-level comparisons, followed where appropriate by Dunn post hoc tests. Subgroup P values were adjusted using the Benjamini-Hochberg false discovery rate procedure.

Because participants contributed repeated cuff-measurement runs, repeated-measures analyses were performed as sensitivity analyses. Measurement error was modeled using a linear mixed-effects model with participant-specific random intercepts. The model intercept represented repeated-measures bias, and between-participant and within-participant variance components were used to calculate repeated-measures LoA. The 95% confidence intervals for the LoA were estimated using 5,000 participant-level cluster-bootstrap resamples. A separate participant-level sensitivity analysis averaged paired measurements within each participant. Within-participant repeatability was evaluated from method-specific BP errors. Within-participant variances were pooled with degrees-of-freedom weighting. Comparator-minus-MTO differences in pooled within-participant error SD were estimated using 5,000 paired participant-level cluster-bootstrap resamples. Absolute-error proportions >10, >15, and >20 mmHg were reported descriptively.

Statistical analyses were conducted with R 4.5.1, IBM SPSS Statistics 26, and MATLAB R2023a. All tests of measurement errors were two-sided; P < 0.05, or false-discovery-rate-adjusted P < 0.05 where applicable, was considered statistically significant. Full statistical procedures are provided in Supplementary Information, Section S1.9.

## Data Availability

All data produced in the present study are available upon reasonable request to the authors.

## Use of large language models

ChatGPT (OpenAI) was used for language polishing and MATLAB code debugging. All outputs were reviewed and revised by the authors.

## Data availability

Deidentified source data underlying the figures, tables, and statistical analyses will be made publicly available through Dryad upon publication.

## Code availability

Custom code used for MTO, mathematical modeling, signal processing, reconstruction, and statistical analysis will be made publicly available with the associated research record.

## Acknowledgements

We thank APT Medical Inc. (Shenzhen Huitai Medical Equipment Co., Ltd.) for technical assistance in establishing the multimodal physiological monitoring and data-acquisition platform. We thank Neusoft Medical Systems Co., Ltd. for providing the photon-counting CT system and technical support for subsequent image reconstruction and rendering. We also thank Boston Scientific Corporation for support with experimental equipment used in this study.

## Author contributions

Y.S., L.X. and G.S. conceived the study. Y.S., H.Y., Y.Z., B.H., A.A., L.X., B.B., X.Y., X.S., G.L., H.R.L. and T.Z. developed the methodology. Y.S., H.Y., Y.Z., B.H., G.S., L.X., H.T.L., B.B., J.L., Y.L., X.G., T.Y., M.L., G.W., X.P. and H.Z. carried out the investigation. H.Y., Y.Z., B.H. and G.W. prepared the visualizations. Y.S. and G.S. acquired funding and administered the project. Y.S. supervised the study. Y.S., H.Y., Y.Z., B.H. and A.A. wrote the original draft. Y.S., H.Y., Y.Z., B.H., A.A., G.S. and L.X. reviewed and edited the manuscript.

Yingxian Sun, Haiyue Yu, Yijie Zhang, Bowei Hao, Alberto Avolio, Guozhe Sun and Lisheng Xu contributed equally to this work.

## Additional information

Supplementary Information is available for this paper.

Correspondence and requests for materials should be addressed to Yingxian Sun.

