## Supplementary Information for "Membrane theory with multimodal evidence defines blood pressure measurement"

### Supplementary Methods

#### *Section S1.1 Study design, cohorts, and ethics*

##### S1.1.1 Overall study design

This study was organized around three coordinated cohorts that addressed complementary questions. The catheterization cohort provided invasive arterial pressure recordings, brachial venous pressure measurements, vascular imaging, and synchronized multimodal recordings required for mechanistic inference during cuff-based blood pressure (BP) measurement. A noninvasive healthy-participant cohort was used to measure cuff-skin contact pressure directly. A separate healthy-volunteer computed tomographic angiography (CTA) cohort was used to define cuff-tissue-artery geometry under controlled cuff pressure levels. The overall design therefore linked invasive physiology, external pressure mapping, and CTA-based three-dimensional (3D) structural imaging within a single mechanistic framework (Supplementary Fig. 1).

##### S1.1.2 Study cohorts

***Cohort 1***: Cardiac catheterization patients. A total of 182 consecutive patients undergoing cardiac interventional therapy at our institution between July 2024 and July 2026 were prospectively screened. Procedures included coronary angiography (CAG), percutaneous coronary intervention (PCI), and electrophysiological procedures. The inclusion criteria were as follows: (1) age > 18 years and < 80 years and (2) availability of complete baseline demographic and clinical data. The exclusion criteria were as follows: (1) acute myocardial infarction requiring primary PCI, (2) hemodynamic instability or cardiogenic shock, (3) congestive heart failure, (4) unrevascularized triple-vessel coronary artery disease, (5) left main coronary artery disease, (6) severe brachial artery stenosis or occlusion caused by peripheral vascular disease, (7) dialysis dependence, and (8) refusal to participate. Among the initially screened patients, 4 were excluded because of significant arrhythmia, poor Korotkoff sound (KS) detectability, inability to tolerate the procedure, or incorrect arterial access. After exclusion, the catheterization-laboratory cohort included 178 patients. Of these, 175 patients contributed invasive arterial pressure recordings and were used for invasive-reference BP analyses, including agreement analyses, error-distribution analyses, repeated-measures sensitivity analyses, and repeatability analyses. Within Cohort 1, a subset of 37 participants with 124 synchronized measurements was used to characterize arterial pressure changes during cuff-based BP measurement. Three additional patients underwent brachial venous pressure measurements only and were included exclusively in the venous pressure sub-study; these 3 patients did not contribute invasive arterial pressure recordings and were not included in invasive-reference BP analyses. Invasive arterial pressure measurements, when performed, were obtained as part of routine clinical care, with additional non-invasive cuff-based recordings acquired synchronously. No patient underwent cardiac catheterization solely for participation in this study. The invasive-reference cohort therefore comprised all consecutively recruited participants with valid arterial recordings during the study period.

The sample size was determined on the basis of the pilot study using paired two one-sided equivalence tests. The equivalence bounds were prespecified as −5 to 5 mmHg, with a one-sided significance level of 0.05 and 95% power. In the pilot data, the expected mean differences between the oscillometric method and the invasive reference measurements were −1.93 mmHg for systolic blood pressure (SBP) and 2.62 mmHg for diastolic blood pressure (DBP), with corresponding standard deviations of the paired differences of 7.379 and 7.853 mmHg, respectively. The calculated sample sizes were 64 participants for SBP and 120 participants for DBP. Because both SBP and DBP were evaluated, the larger estimate of 120 participants was adopted as the minimum number of evaluable participants. To allow for up to 20% of enrolled participants being nonevaluable because of incomplete paired measurements or failure to meet the prespecified signal-quality criteria, the recruitment target was increased to at least 150 participants. The final cohort included 175 participants contributing 630 valid paired measurements, thereby exceeding the required sample size.

***Cohort 2***: Healthy participants for pressure-mapping measurements. A separate cohort of 50 healthy participants was recruited for noninvasive surface pressure-mapping measurements during controlled cuff inflation. Cuff-transmitted surface pressure was recorded using a flexible external pressure-sensing pad positioned between the upper-arm cuff and the skin. The protocol was designed to characterize the spatiotemporal distribution of cuff-induced surface pressure and involved no invasive procedures, vascular access, radiation exposure, or administration of contrast agents.

***Cohort 3***: Healthy volunteers for upper-limb CTA under cuff compression. Under the overall study framework, an additional cohort of 22 healthy volunteers was recruited for supplemental CTA-based imaging studies. This cohort was designed specifically for upper-limb CTA under cuff compression and did not overlap with Cohort 2. No invasive procedures were performed in this cohort, and participants were not included in the invasive BP acquisition group.

Eligible participants were 18–60 years of age, had a body mass index (BMI) of 18–26 $kg/m^2$, had no contraindications to CT examination, and were able to cooperate with upper-limb CTA during cuff pressurization. Exclusion criteria included contraindications to imaging examination, pregnancy or suspected pregnancy, and any condition that, in the judgment of the investigators, made participation unsuitable.

Cohorts 2 and 3 were observational mechanistic validation cohorts designed to assess whether the prespecified physiological phenomena could be consistently observed, rather than to test equivalence or superiority between measurement methods. Consecutive eligible participants with evaluable recordings were included during the study period, and the reproducibility of the observed phenomena was assessed at the participant level.

For the study cohorts, demographic and anthropometric variables, including age, sex, height, weight, and body mass index (BMI), were recorded when available. In Cohort 1 (the cardiac catheterization cohort), additional information was collected on clinical history. These data are summarized in Supplementary Table 1.

###### S1.1.3 Ethics and informed consent

All study procedures were approved by the Ethics Committee of the First Affiliated Hospital of China Medical University, Shenyang, China (approval No. [2026] 2024-609-5). All procedures involving human participants were conducted in accordance with the ethical principles of the Declaration of Helsinki. Written informed consent was obtained from all participants before enrollment in the invasive cardiac-catheterization cohort and the healthy-volunteer CTA cohort. The noninvasive pressure-mapping study was approved under the same institutional protocol. No procedure performed solely for research altered the standard of clinical care delivered to patients in the catheterization cohort.

#### *Section S1.2 Integrated instrumentation and synchronized acquisition*

##### S1.2.1 Overview of the measurement systems

The integrated experimental platform consisted of the following subsystems: (1) a highly integrated multichannel electrophysiology recording system (HIERS™ multichannel physiological recorder; High Integrated EP Recording System; Shanghai Hongtong Industrial Co., Ltd., Shanghai, China), integrating electrophysiological recording, stimulation, $SpO_2$ monitoring, and invasive and noninvasive BP measurement, for physiological signal acquisition and vital sign monitoring in the catheterization laboratory; (2) a custom pressure-control system for cuff inflation, cuff deflation, and synchronized data transmission; (3) an invasive BP measurement system based on a 0.014-inch pressure guidewire (COMET™ II Pressure Guidewire, Boston Scientific, Marlborough, MA, USA; nominal diameter, 0.37 mm; calculated cross-sectional area (CSA), 0.108 mm$^2$) coupled to an FFR Link™ interface module (Boston Scientific), which received and transmitted BP signal from the pressure guidewire to the HIERS™ multichannel physiological recorder; (4) angiographic or guiding catheters were used as appropriate for the procedure, including RADIFOCUS™ Optitorque™ Angiographic Catheters (Terumo Corporation) and either Launcher™ Guide Catheters (Medtronic, Inc.) or March™ Guiding Catheters (Hunan APT Medical Inc.), with catheter curve and French size selected according to procedural requirements and target vessel anatomy; (5) a custom sound-acquisition system incorporating an electronic stethoscope chest piece adapted from an automatic auscultatory BP monitor; and (6) an intravascular ultrasound (IVUS) system consisting of a disposable IVUS diagnostic catheter (Model 02-6F60-01, 60 MHz, compatible with a 6F guide catheter) and an IVUS console (Model VH-60; Shenzhen Insight Lifetech Co., Ltd.).

##### S1.2.2 Synchronized acquisition architecture

Synchronous acquisition across multimodal devices was achieved through integrated signal receiving and signal generation modules within each system, with the HIERS™ multichannel physiological recorder, the digital subtraction angiography (DSA) system (Philips Allura Xper FD10, Neusoft NeuAngio 30C, or Siemens Artis Q.zen floor), the IVUS acquisition system, and the external trigger connected in parallel. Upon initiation of synchronous acquisition, the signal trigger outputs a pulse signal with an amplitude of 0.8 mV and a duration of 5 ms. Each device detects the rising edge of the pulse signal through its signal receiver and uses it as the synchronization timestamp. The signal trigger is a custom-designed trigger circuit. When IVUS is connected, the IVUS acquisition system itself serves as the signal-triggering device and automatically generates a pulse signal when acquisition begins. Interdevice connections are established using highly shielded coaxial cables, which effectively prevent false triggering caused by electrical interference.

##### S1.2.3 Pressure-control system

**Inflation module**. Cuff inflation and deflation were driven by a high-linearity, low-vibration piezoelectric air pump (Murata, MZB3004T04). The module provides a maximum static pressure of 60 kPa (19.5 Vdc) and a stable, low-pulsatility airflow with a no-load flow rate of 200 mL/min (18 Vdc). High-frequency piezoelectric actuation (~23 kHz) produces airflow with minimal pulsation and operates beyond the audible range, resulting in negligible acoustic noise. The driving displacement is on the order of tens of micrometers, yielding minimal transmitted mechanical vibration. These features were selected to avoid mechanically induced artifacts in oscillometric waveforms and to prevent interference with simultaneous acoustic signal acquisition. The module exhibits rapid dynamic response with short latency from activation to steady output and uses epoxy encapsulation and reinforced structural design to enhance resistance to shock and vibration during repeated operation.

**Pressure-sensing module**. Cuff pressure was continuously monitored using a piezoresistive pressure sensor (Freescale MPXV5050GP). The sensor provides temperature-compensated output over a wide operating range (−40°C to 125°C), with a maximum error of ±2.5% within 0°C to 85°C. Its sensing element is based on a silicon shear-stress strain gauge combined with metallized thin-film and bipolar processing, yielding good linearity and a sensitivity of approximately 90 mV/kPa. These characteristics enable stable real-time pressure feedback during dynamic cuff modulation.

**Control and regulation**. Closed-loop pressure control was implemented using a high-performance microcontroller (STM32 series; STMicroelectronics). Target pressure levels and inflation/deflation profiles were programmable, allowing high-response, high-precision regulation of cuff pressure and synchronized time alignment with invasive pressure recordings, imaging data, and acoustic signals.

##### S1.2.4 Cuff specification and scope of cuff-related analysis

Commercially available upper-arm oscillometric cuffs were used in this study, including the OMRON Medium Cuff HEM-CR24 Type B and the OMRON GS CUFF2 in size M. According to the manufacturer's instructions and publicly available product information, both cuffs are intended for upper-arm circumferences of 22–32 cm; the HEM-CR24 Type B cuff has approximate dimensions of 145 × 466 mm, whereas the GS CUFF2 M is the standard medium cuff used with the OMRON HBP-1120/HBP-1120U platform. In all measurements, the cuff was wrapped around the upper arm with the lower cuff edge positioned 1–2 cm above the antecubital fossa, and with the artery marker/tubing aligned along the medial aspect of the arm to match the course of the brachial artery. Cuff information is provided here to document the experimental hardware and measurement setup used during data acquisition, rather than to compare the performance or superiority of different cuff designs. Instead, the mechanistic focus of the present study was to identify oscillometric waveform feature changes that were shared across different cuffs, indicating common hemodynamic phenomena underlying cuff-based BP measurement.

##### S1.2.5 KS acquisition unit

KS were acquired using a compact, low-frequency, high-fidelity electronic auscultation module optimized for accurate reproduction of signals in the 20–200 Hz band, corresponding to the dominant spectral components of KS. To preserve low-frequency waveform integrity, no hardware high-pass filtering was applied in the acoustic front end. To ensure robust signal acquisition in the catheterization laboratory environment, very high-frequency components (approximately >100 kHz) were attenuated to reduce susceptibility to radiofrequency interference from surrounding medical equipment. The stethoscope chest piece was adapted from the built-in electronic stethoscope of an automatic auscultatory BP monitor and integrated into the custom acquisition system.

##### S1.2.6 Contact-pressure sensing unit

The axial distribution of cuff–tissue contact pressure was measured using a flexible external pressure-sensing pad (Mini pressure-sensing pad; JQ Industries, Weihai JQ Industrial Technology Co., Ltd., Weihai, China). The sensing layer comprised 1,024 sensing points arranged in a 32 × 32 matrix over a 200 × 200 mm active area, with a default pressure range of 0–40 kPa and a sampling frequency of 100 Hz. In the present study, this device was used exclusively for noninvasive mapping of cuff-transmitted surface pressure beneath the upper-arm cuff (Supplementary Fig. 2).

#### *Section S1.3 Core invasive and imaging measurements*

##### S1.3.1 Brachial arteriography

Brachial artery angiography was performed in 22 patients. These examinations were conducted in 2 principal settings during cuff-based BP measurement. In one subgroup, ipsilateral brachial angiography was performed alone via right radial arterial access. In the other subgroup, brachial angiography was performed in conjunction with simultaneous acquisition of mechanistic data using multiple pressure guidewires and/or IVUS, with the arterial access route selected according to the requirements of the combined procedure, including femoral access when necessary. Angiography was performed under standard sterile catheterization-laboratory conditions. The examined upper limb was positioned to permit stable imaging of the cuffed brachial segment while minimizing motion, direct external compression, and vasospastic stimuli. In all cases, the catheter tip was positioned at the proximal end of the cuff-covered segment rather than within the cuffed segment itself. Digital subtraction angiography was then performed by selective contrast injection from this proximal position, allowing visualization of brachial arterial morphology during cuff compression while preserving the intravascular working space required for pressure-guidewire and IVUS measurements. Because of technical limitations, brachial angiography and IVUS imaging were acquired in separate measurement runs (as shown in Fig. 2a of the main text).

##### S1.3.2 IVUS-based measurement of vascular CSA

IVUS-synchronized intravascular pressure measurements were performed in 28 patients using the IVUS system described above. The IVUS catheter used in this study operated at 60 MHz with a frame rate of 100 frames/s. Before each acquisition, the IVUS catheter was flushed and deaerated to establish a continuous acoustic-coupling fluid column, then connected to the console and pullback unit and advanced over the pressure guidewire through the guide catheter to the predefined vascular site. During each BP measurement run, the IVUS catheter was maintained at a fixed intravascular position, and continuous IVUS imaging was recorded throughout the cuff-measurement process to capture site-specific vascular cross-sectional changes over time. The pullback unit was connected during imaging acquisition, but automatic pullback was not used during the actual BP recording runs. After all BP measurements at one site had been completed, the IVUS catheter was repositioned to the next predefined arterial site and the same procedure was repeated.

For continuous CSA measurement, 30 sampling points were selected within each measurement cycle at approximately equal temporal intervals, with the additional requirement that both the maximal and minimal cross-sectional areas were included. For this analysis, one measurement cycle was defined as spanning the sequence from the onset of vascular expansion to maximal expansion, and then through the contraction phase to minimal CSA. At each sampling point, the luminal boundary was delineated along the inner intimal contour and the vascular CSA was calculated. Measurement consistency was evaluated using a standardized blinded protocol involving multiple delineators and independent supervisory review. At least 2 professionally trained and experienced clinicians independently analyzed the same data segment, and the final measurements were reviewed by a third and fourth specialist to ensure consistency and quality control. Throughout the measurement procedure, both the primary measurers and the reviewers were blinded to experimental information, including site identity and acquisition order.

###### S1.3.3 Reconstruction of whole-segment luminal volume from five IVUS measurement sites

To reconstruct the dynamic luminal volume of the cuff-covered arterial segment, IVUS-derived CSA waveforms were obtained at five predefined axial sites and combined into a whole-segment volume waveform. Because these five CSA recordings were acquired during five separate blood pressure measurements rather than simultaneously, temporal registration was performed before reconstruction. Heart rate and blood pressure remained stable across measurements, and cuff inflation was applied at a constant rate. Cuff pressure relative to invasive DBP and SBP was used as the common reference. For each recording, the number of cardiac cycles occurring while cuff pressure was between DBP and SBP was matched across measurements, and all beats from 5–6 cycles below DBP to 5–6 cycles above SBP were included for analysis. To account for beat-to-beat variation in cycle length, all included cardiac cycles were normalized in duration so that corresponding CSA waveforms from different measurements could be aligned on a common normalized time axis.

Axial delays caused by pulse-wave propagation were then corrected using electrocardiogram-derived pulse arrival time as the delay metric. The CSA waveform measured at Site ① was used as the temporal reference, and the beat-by-beat CSA waveforms from the remaining sites were shifted by their corresponding delays to align them with Site ①. Linear interpolation was used to fill the time gaps introduced by delay correction, preserving waveform continuity and uniform sampling across sites. After temporal normalization and delay correction, the CSA waveforms from the five sites were linearly interpolated along the vessel axis to reconstruct the continuous axial luminal area profile at each normalized time point. Whole-segment luminal volume was then calculated by integrating this axial CSA profile along the vessel axis.

###### S1.3.4 3D vascular reconstruction

Among the 28 patients who underwent IVUS examination, 4 patients additionally underwent cuff-inflation measurements with IVUS acquisition at 13 evenly distributed sub-cuff locations. These 13 measurement sites spanned the cuff-covered brachial arterial segment from proximal to distal. At each site, the pressure-guidewire sensor was positioned at the level of the corresponding IVUS imaging plane.

To reconstruct the continuous 3D configuration of the brachial artery beneath the cuff during BP measurement, the workflow comprised frame-wise IVUS contour extraction, frame-index recovery, multisource temporal alignment, and 3D volume generation.

For each measurement, the corresponding IVUS video stream was parsed frame by frame into RGB images. The subintimal contour on the IVUS images was manually delineated at 1-frame intervals. In parallel, a segmentation workflow guided by a vision foundation model was applied to the IVUS frames to identify the vessel region and generate candidate masks for contour regularization and quality control. The masks were binarized, abnormal or empty masks were excluded, and the outer boundary of the retained target region was extracted to obtain a structured frame-wise geometric representation of the vascular contour.

To recover temporal indices, numeric frame labels embedded in the IVUS video were extracted by optical character recognition (OCR) from a predefined region of interest after grayscale conversion, denoising, and binarization. The allowable character set was restricted to numeric characters, and the recognized sequence was further corrected using temporal-consistency constraints, thereby establishing a stable mapping between IVUS frame number and video-frame index.

The IVUS frame sequence, recovered frame indices, and contour results were then aligned by combining the OCR-derived mapping with externally recorded pressure and electrocardiographic timing information. Inter-site delay was determined from the relative timing of the simultaneously recorded pressure and electrocardiographic signals, after which each cardiac cycle was normalized and aligned across all 13 sites on a common time axis. This procedure enabled temporal correspondence among spatially separated IVUS measurements acquired during separate cuff-inflation runs under matched conditions.

After temporal alignment, the contours from the 13 spatially ordered sub-cuff locations were arranged sequentially along a common proximal-to-distal physical axis. Contour masks were normalized to a common spatial resolution and stacked along the axial direction to generate an initial 3D volume representation of the artery. To reduce discontinuity between adjacent slices, interpolation based on the signed distance function (SDF) was performed between neighboring contours. The interpolated volumes were then re-binarized to generate a high-resolution 3D dataset with continuous vascular geometry.

For dynamic reconstruction, the above procedure was repeated frame by frame throughout the aligned cardiac cycle, and adjacent contours were smoothly connected to form the instantaneous vascular geometry at each time point during cuff inflation. The reconstructed volumes were exported in standard volumetric format for visualization and analysis, and frame-by-frame rendering of the reconstructed geometries was used to generate a dynamic continuous video representing 3D brachial artery motion beneath the cuff.

##### S1.3.5 Invasive BP measurement and reference definition

**Pressure guidewire measurements.** Before use, the carrier tube was flushed with heparinized saline to remove air completely and to hydrate the hydrophilic segment of each pressure guidewire. Each pressure guidewire was connected to the FFR Link with the dedicated optical cable for automatic zeroing and was then advanced into the target vessel. For pressure equalization, the sensors of the pressure guidewires were positioned at the same vascular location just distal to the tip of the guide catheter, the guide catheter was flushed with heparinized saline until contrast was fully cleared, and the external pressure transducer was maintained at heart level. After waveform stabilization, each pressure guidewire was equalized against the same reference pressure signal, which in the present study was the catheter pressure signal provided by the fluid-filled catheter/transducer system. When more than one pressure guidewire was used in the same acquisition, inter-wire consistency was checked before recording by comparing the displayed pressures of all guidewires at the same vascular position against the same catheter reference signal. For single-guidewire recordings, the pressure guidewire was zeroed and equalized against the catheter reference signal before advancement to the target site. If concordance was not achieved, all pressure guidewires were withdrawn and the zeroing/equalization procedure was repeated until consistent pressure readings were obtained, after which BP recording was initiated.

**Catheter-based measurements.** Before invasive BP monitoring with a fluid-filled catheter system, the catheter–tubing–transducer assembly was prefilled with flush solution and fully deaerated to eliminate air bubbles and maintain a continuous fluid column. The pressure transducer was positioned at the level of the patient's right atrium and connected to the HIERS™ multichannel physiological recorder through a dedicated cable. For zero calibration, the catheter stopcock was opened to atmospheric pressure and the transducer was zeroed; the stopcock was then returned immediately to the monitoring pathway to initiate continuous invasive BP recording. The dynamic response characteristics of the catheter system were assessed in vivo by a fast-flush test. The measured natural frequency of the catheter system used in this study was 14.29 Hz, and the amplitude ratio of 0.184 corresponded to a damping coefficient of 0.474. These values were recorded to characterize the dynamic performance of the monitoring system [51].

**Reference BP definition.** Sub-cuff intra-brachial arterial pressure was used as the run-specific invasive reference pressure for each cuff BP measurement. For each run, the reference window was selected from a continuous, analyzable, artifact-free arterial pressure segment of approximately 30 s within 1 min before cuff inflation, whenever available. When feasible, the segment closest to the onset of cuff inflation was used. Window selection was based solely on signal quality and temporal proximity to cuff inflation, independent of the results of any non-invasive BP estimation method. Within each run-specific reference window, invasive SBP was defined as the mean of beat-wise systolic maxima, and invasive DBP as the mean of beat-wise diastolic minima. Beat-wise invasive mean arterial pressure (MAP) was calculated as the time integral of the pressure waveform over each cardiac cycle divided by cycle duration, and the final invasive MAP for that run was taken as the mean of these beat-wise values across the reference window. Thus, each cuff BP measurement run was paired with its own immediately preceding invasive reference BP values.

#### *Section S1.4 Non-invasive BP measurements*

##### S1.4.1 Auscultatory method

The sound-acquisition system was pre-adjusted and connected before measurement. After the BP cuff had been fixed on the patient's right upper arm, the point of strongest brachial artery pulsation distal to the cuff was identified by palpation and marked. The auscultation chest piece was placed at the marked location and fixed securely with an elastic band to ensure tight contact with the skin without adding direct compression to the brachial artery. KS signals during BP measurement were collected synchronously.

Based on previous studies of the KS frequency domain and the frequency-division analysis performed in this study, KS signals were divided into 3 frequency bands—low (20–50 Hz), medium (50–150 Hz), and high (150–900 Hz)—to characterize the frequency-domain features of different signal components [54–56]. If this frequency-division approach did not achieve adequate signal separation, the frequency bands were adjusted dynamically to maintain consistency of the three-band KS representation across participants.

To improve the accuracy of auscultatory BP interpretation, a combined visual–auditory strategy was used, with visual assessment as the primary basis and auditory assessment as supportive information. DBP interpretation relied mainly on visual identification of signal changes, with auditory input used for confirmation when necessary. In some subjects, the KS associated with DBP were continuous and did not show a distinct abrupt onset or offset because of brachial artery pulsation, but instead exhibited a marked timbre change at phase K5. Frequency-division processing facilitated separation of non-KS interference and enhanced identification of the relevant acoustic features.

SBP interpretation was also based primarily on visual assessment with auditory assistance. During most inflation recordings and some deflation recordings, the KS associated with SBP were extremely low in loudness, were easily overlapped with environmental noise, and were difficult to distinguish reliably by listening alone. However, because of the physiological mechanism of SBP formation, these signals displayed abrupt onset or disappearance. Frequency-division processing improved separation of weak signals from environmental noise and facilitated visual identification of the SBP transition.

For both DBP and SBP, 2 senior clinicians independently identified the relevant position on the basis of the frequency-divided KS signal, using visual assessment together with auditory review. For DBP, the target was the onset, disappearance, or timbre-change position. For SBP, the target was the position of sudden signal appearance or disappearance, while ensuring that the corresponding pressure was not lower than the maximum pressure that remained distinguishable by auditory assessment. If the 2 readings were inconsistent, or if the visually derived SBP value was lower than the auditory estimate, a third senior physician reviewed the recording and determined the final value. All assessors were blinded and were provided only with the KS segments and information on whether the BP measurement phase was inflation or deflation.

For each cuff-measurement run, MAP was estimated as DBP + (SBP − DBP)/3, using the auscultatory SBP and DBP obtained during the same run.

##### S1.4.2 Fixed-ratio amplitude coefficient method

The fixed-ratio amplitude coefficient method was used for oscillometric BP measurement. After extraction of the oscillometric waveform, the waveform envelope was constructed. The point with the maximum envelope amplitude was taken as the MAP. SBP and DBP were then estimated according to the following relations [19]:

$$K_d = \frac{A(\mathrm{DBP})}{A(\mathrm{m}ax)}, \tag{S1}$$

$$K_s = \frac{A(\mathrm{SBP})}{A(\mathrm{m}ax)}, \tag{S2}$$

where $A(max)$ is the oscillometric waveform amplitude at the envelope maximum, $A(\mathrm{DBP})$ is the oscillometric waveform amplitude corresponding to DBP, $A(\mathrm{SBP})$ is the oscillometric waveform amplitude corresponding to SBP, $K_d$ is the amplitude coefficient for DBP, and $K_s$ is the amplitude coefficient for SBP. Different research groups and oscillometric devices use different values for $K_d$ and $K_s$. In this study, $K_s = 0.593$ and $K_d = 0.717$ were used [19].

##### S1.4.3 Derivative-based oscillometric comparator

As an additional empirical comparator, a derivative-based oscillometric method was applied to the extracted oscillometric feature trajectory. BP landmarks were assigned according to maximal first-derivative changes in the corresponding trajectory. This analysis was used only as a supplementary local-feature comparator.

#### *Section S1.5 Morphology-Transition Oscillometry (MTO)*

##### S1.5.1 Conceptual overview

Morphology-Transition Oscillometry (MTO) was developed as a mechanistic framework for localizing DBP, MAP, and SBP from cuff oscillations by identifying pressure-dependent beat-to-beat changes in oscillogram morphology. Cuff oscillations arise from the superposition of pulsatile volume changes across arterial segments exposed to spatially heterogeneous external compression. As cuff pressure varies, the contribution of specific arterial segments to the oscillometric waveform changes at characteristic pressure levels, producing abrupt beat-to-beat morphological transitions.

In this study, SBP- and DBP-related MTO features were extracted using wavelet-domain analysis. Wavelet-domain analysis was performed with the Mexican-hat mother wavelet and dyadic scale sampling to localize transient changes in waveform curvature and time–scale structure. DBP was localized from valley-proximal beat-to-beat morphology transitions, whereas SBP was localized by dynamic tracking of a right-sided positive hotspot within the primary above-zero component. MAP was localized using the dual-notch crossover analysis described in Section S1.5.6.

For each measurement, the algorithm generated one unique candidate transition point for SBP and one for DBP based on predefined wavelet features. These candidate points were then verified under blinded conditions by at least two investigators familiar with the characteristic MTO features, without access to invasive BP reference values.

##### S1.5.2 Signal representation and wavelet transform

Let $p_{osc}(t)$ denote the oscillometric waveform after standard preprocessing and beat segmentation. Beats were indexed by $i = 1, \dots, K$.

For SBP-related analysis, a local baseline (zero line) was defined for each beat, and the primary above-baseline component was extracted:

$$x_i^A(t), \qquad t \in [0, T_i^A], \tag{S3}$$

where $T_i^A$ is the duration of the above-baseline segment.

The Mexican-hat (Ricker) mother wavelet was defined as:

$$\psi(t) = (1 - t^2)\exp\left(-\frac{t^2}{2}\right), \tag{S4}$$

For a beat segment $x_i(t)$, the continuous wavelet transform (CWT) was computed as:

$$W_i(s, \tau) = \frac{1}{\sqrt{s}} \int x_i(t)\, \psi\left(\frac{t-\tau}{s}\right) dt, \tag{S5}$$

where $s$ denotes scale and $\tau$ denotes time. Scales were restricted to a finite set $S = \{s_{min}, \dots, s_{max}\}$ such that the effective wavelet support remained commensurate with the time scale of the analysis window. The Mexican-hat wavelet used here is real-valued; therefore, the complex-conjugate symbol included in the general definition of the continuous wavelet transform is not required in Eq. S5. The mother-wavelet normalization constant is omitted because the present analysis compares relative coefficient changes within the same recording and does not compare absolute wavelet energy across different wavelet normalizations. The scale variable a controls temporal dilation of the wavelet, whereas $\tau$ specifies its temporal location within the analyzed beat segment. All scales were expressed in sampling-point units in the discrete implementation and were converted to equivalent periods using the sampling interval when physiological time scales were reported.

To make scale selection reproducible, scales were placed on a dyadic grid following Torrence and Compo, with $\delta j$ controlling the sampling density. In the present implementation, $\delta j = 0.125$ was used as the default dense grid for local morphology analysis [58].

$$s_j = s_0 2^{j\delta j}, \tag{S6}$$

For the Mexican-hat wavelet (DOG, $m = 2$), the equivalent Fourier period $\lambda(a)$ is related to scale a by the standard CWT conversion $\lambda(a) = 2\pi\sqrt{(2/5)}a \approx 3.97a$. Accordingly, admissible scales were determined from the physiologically defined ROI width rather than from an arbitrary fixed integer list [58,59].

$$\lambda(a) = 2\pi\sqrt{\frac{2}{5}}a \approx 3.97a. \tag{S7}$$

The lower bound was chosen so that the minimum equivalent period was approximately $2\Delta t$. Therefore, $a\_min \approx 2\Delta t/3.97$; with $fs = 1000$ Hz ($\Delta t = 1$ ms), the theoretical lower bound is about 0.50, and the discrete implementation therefore used $a\_min = 1$ [58].

$$a_{min} \approx \frac{2\Delta t}{3.97}. \tag{S8}$$

The upper bound can in principle be related to ROI width, where $W_{ROI}$ denotes the effective temporal width of the feature-extraction ROI. This ROI-matched interpretation remains useful mechanistically [59]. In the present SBP implementation described below, however, the discrete scale set was fixed to a ∈ [1,20] with $\delta j = 0.125$.

$$a_{max} = \frac{W_{ROI}}{3.97}. \tag{S9}$$

Coefficients were aggregated across scales into a one-dimensional curvature-saliency profile:

$$S_i(\tau) = \frac{1}{|S|}\sum_{s\in S} |W_i(s,\tau)|. \tag{S10}$$

$$E_i(\tau) = \frac{1}{|S|}\sum_{s\in S} W_i\,(s,\tau)^2, \tag{S11}$$

Equation S11 defines a complementary squared-coefficient descriptor. It is retained to document the complete feature formulation but was not used to replace the primary absolute-coefficient saliency profile in the main DBP and SBP decision rules. Unless otherwise specified, the results reported for MTO were obtained from the primary saliency definition rather than from the squared-energy alternative.

##### S1.5.3 Boundary-aware windowing principle

Window boundaries were used only to delimit analysis regions and were not themselves used as loci for feature extraction. Although the end of the above-baseline component (zero-line crossing) is a mathematical boundary, no decision statistics were extracted in its immediate vicinity. All decision statistics used for DBP and SBP localization were computed from interior regions of interest (ROIs) away from boundaries. Optional boundary stabilization, when used, was applied only to improve coefficient stability at truncation edges and did not alter the fact that feature extraction itself was restricted to the original interior ROI.

##### S1.5.4 DBP localization

Near DBP, transient equality between intra- and extravascular pressures produces pronounced mechanical transitions during the terminal downstroke immediately preceding the beat valley, yielding abrupt beat-to-beat changes in local curvature.

Let $t_{v,i}$ denote the valley time (minimum) of beat i. A fixed-length terminal window was defined as:

$$W_i^{DBP} = \left[t_{v,i} - \Delta t,\ t_{v,i}\right], \tag{S12}$$

where $\Delta t$ could be set as a constant (for example, 120 ms) or in adaptive form as:

$$\Delta t = 0.15T_i, \tag{S13}$$

In the primary analysis, $\Delta t$ was fixed at 120 ms to maintain an identical terminal-window duration across beats and measurements. Equation S13 states the corresponding heart-rate-scaled alternative, in which $T_i$ is the duration of beat i. The adaptive form was not combined with the fixed-window result and is retained only to describe the physiological scaling rule that may be used in future implementations.

Because $W_i^{DBP}$ is truncated at its left edge but physiologically anchored at the valley, optional boundary stabilization could be applied only at the truncation side by one-sided mirror extension, whereas no padding or exclusion was applied at the valley anchor. In all cases, feature extraction and decision statistics were interpreted only over the original interval.

A valley-proximal ROI was defined as the final $\delta t$ (typically 20–40 ms) of $W_i^{DBP}$. A beat-level score was defined as:

$$\mathrm{Score}_i^{DBP} = \max_{\tau \in ROI} S_i(\tau). \tag{S14}$$

In the present implementation, the default DBP ROI width was empirically set to 40 ms. Thus, $\mathrm{W\_DBP, ROI} = 40$ ms, which gives $\mathrm{a\_max, DBP} \approx 40/3.97 \approx 10.1$; the practical DBP scale range was therefore approximately $a \in [1,10]$.

$$W_{DBP,ROI} = 40\mathrm{ms}, \tag{S15}$$

$$a_{max,DBP} \approx \frac{40}{3.97} \approx 10.1. \tag{S16}$$

Beat-to-beat change in the wavelet-saliency score defined in Eq. S14 was computed as:

$$\Delta S_i^{DBP} = \mathrm{Score}_{i+1}^{DBP} - \mathrm{Score}_i^{DBP}. \tag{S17}$$

The DBP transition was localized by:

$$i_{DBP} = \operatorname*{argmax}_i \left|\Delta S_i^{DBP}\right|. \tag{S18}$$

The estimated DBP was obtained by mapping the transition index to cuff pressure, $DBP_{est} = g(i_{DBP})$. During cuff inflation, DBP was assigned to the cuff pressure at the onset of the later beat in the wavelet-defined transition pair. The beat-to-beat change in this score was subsequently computed for adjacent beats and plotted against beat index or cuff pressure. As cuff pressure approached DBP, the valley-proximal descending segment underwent its most prominent inter-beat morphological transition, producing the dominant turning point in the scatter plot of beat-to-beat change. DBP was therefore identified from this turning point, and the cuff pressure assigned to the corresponding beat within the maximally changing beat pair was taken as the predicted DBP. Thus, DBP was determined by the largest local morphological transition between adjacent beats, rather than by a fixed wavelet-energy threshold. Moderate variation in $\Delta t$ and ROI duration changed the magnitude of $\Delta S$ but did not materially shift the location of the maximal beat-to-beat transition; default values were therefore used for stability and simplicity.

##### S1.5.5 SBP localization

For SBP analysis, each oscillometric beat was first reduced to the primary above-zero component. For beat i, the local zero line was defined as described above, and the analysis window was the contiguous positive segment with the largest area above this zero line. This window was used as the only SBP wavelet-analysis region in the present implementation. No second fixed ROI, no late-primary subwindow, and no distal-pressure-defined window were used for SBP localization.

Let $x_i(\tau)$ denote the primary above-zero component of beat i, where $\tau$ is the local time index within that beat-specific segment. The Mexican-hat coefficient map was computed over the dyadic scale set $A$ defined in Section S1.5.2. Negative coefficients were not used for SBP hotspot tracking; instead, the positive coefficient map was defined as follows:

$$W_i^{+}(a,\tau) = \max(W_i(a,\tau), 0) \tag{S19}$$

Here, $W_i\ (a,\tau)$ is the Mexican-hat wavelet coefficient of beat i at scale a and local time $\tau$; $[x]_+$ denotes the positive-part operator, defined as $\max(x, 0)$; the analyzed scale a was restricted to the predefined dyadic scale set used for SBP hotspot tracking. Negative coefficients were set to zero only for the SBP hotspot-tracking step and were not removed from the preceding general CWT formulation.

A one-dimensional positive-hotspot curve was then obtained by averaging the positive coefficients over the scale axis:

$$H_i(\tau) = \frac{1}{|A|}\sum_{a\in A} W_i^{+}\ (a,\tau). \tag{S20}$$

The hotspot curve was lightly smoothed in the time direction before local-maximum detection. Candidate peaks were required to lie in the right-sided portion of the above-zero component, because the SBP-related morphology of interest corresponds to the late positive component of the primary oscillometric wave. In the first analyzed beat, the target hotspot was initialized as the rightmost valid positive hotspot after exclusion of the extreme right boundary. If no valid right-sided hotspot was detected, tracking was terminated rather than forced onto an unrelated component.

For each candidate hotspot m in beat i, the temporal support was defined from the nearest valleys of the hotspot curve on both sides of the candidate peak, with a small rightward extension to preserve the wave tail. Denoting the resulting temporal support by $T_{i,m}$ and the corresponding scale support by $A_{i,m}$, the local positive-coefficient patch was

$$B_{i,m} = \{W_i^{+}(a,\tau) : a \in A_{i,m},\ \tau \in T_{i,m}\}. \tag{S21}$$

The scale support was determined from the local scale profile within the candidate time interval and was expanded slightly to retain the full positive hotspot. The patch center was then calculated as a coefficient-weighted centroid in time and scale:

$$\tau_{i,m}^{c} = \frac{\sum_{\tau \in T_{i,m}} \tau \sum_{a \in A_{i,m}} W_i^{+}(a,\tau)}{\sum_{\tau \in T_{i,m}} \sum_{a \in A_{i,m}} W_i^{+}(a,\tau) + \varepsilon}, \tag{S22}$$

$$a_{i,m}^{c} = \frac{\sum_{a \in A_{i,m}} a \sum_{\tau \in T_{i,m}} W_i^{+}(a,\tau)}{\sum_{a \in A_{i,m}} \sum_{\tau \in T_{i,m}} W_i^{+}(a,\tau) + \varepsilon}. \tag{S23}$$

A candidate hotspot strength was defined as the mean positive coefficient within the patch multiplied by the square root of the patch area:

$$E_{i,m} = \frac{1}{\sqrt{N_{i,m}}} \sum_{r=1}^{N_{i,m}} w_{i,m,r}^{+} \tag{S24}$$

Here, the positive wavelet coefficients within candidate hotspot m of beat i are vectorized as $w_{i,m,r}^{+}$, $r = 1, \dots, N_{i,m}$, and $N_{i,m}$ is the total number of discrete time–scale samples in that hotspot. Each $w_{i,m,r}^{+}$ is taken from the positive coefficient map defined in Eq. S19. This definition gave weight to both the intensity and the spatial extent of the positive time-scale region, while avoiding an unrestricted dependence on window size.

After initialization, the target hotspot was tracked dynamically across beats. Candidate patches in beat i were first required to overlap the temporal support of the previously tracked hotspot by at least a minimum number of samples. This constraint prevented the algorithm from jumping to a different positive component after the SBP-related hotspot had weakened or disappeared. For each admissible candidate, the normalized temporal displacement, normalized scale displacement, patch correlation, and relative peak amplitude were defined as

$$d_{\tau,i,m} = \min\left(\frac{\left|\tau_{i,m}^{c} - \tau_{i-1}^{c}\right|}{G_i}, 1\right), \tag{S25}$$

$$d_{a,i,m} = \frac{\left|a_{i,m}^{c} - a_{i-1}^{c}\right|}{|A|}, \tag{S26}$$

$$r_{i,m} = \max\{\rho(\hat{\mathbf{b}}_{i-1}, \hat{\mathbf{b}}_{i,m}), 0\}, q_{i,m} = \frac{p_{i,m}}{\max_{\tau} H_i(\tau) + \varepsilon} \tag{S27}$$

Here, $\rho(\cdot,\cdot)$ is the Pearson correlation coefficient between the normalized vector representation of the hotspot tracked in beat i−1 and that of candidate m in beat i after both patches have been mapped to a common time–scale support. Negative correlations were truncated at zero because an anticorrelated patch was not considered a continuation of the same positive hotspot. The quantity $p_{i,m}$ is the peak value of candidate m on the positive-hotspot curve $H_i(\tau)$; $q_{i,m}$ is its peak value normalized by the largest positive-hotspot value in beat i; and $\varepsilon$ is a small positive constant used only to prevent division by zero. The tracking score was then computed as

$$M_{i,m} = w_\tau(1 - d_{\tau,i,m}) + w_a(1 - d_{a,i,m}) + w_r r_{i,m} + w_q q_{i,m}. \quad \text{(S28)}$$

The default weights were $w_\tau = 0.72$, $w_a = 0.12$, $w_r = 0.10$, and $w_q = 0.06$. Thus, temporal continuity was given the greatest weight, with smaller contributions from scale continuity, patch similarity, and peak amplitude. The admissible candidate with the largest $M_{i,m}$ was selected as the tracked hotspot. Tracking was terminated when no candidate had sufficient temporal overlap with the previous hotspot, when the best tracking score was below the acceptance threshold, or when the current candidate peak fell markedly relative to that of the preceding tracked beat. These termination rules were used to identify disappearance of the same right-sided morphology rather than to relabel a different residual component as the SBP target.

For each successfully tracked beat, the beat-level target-wave value was defined from the selected hotspot patch as

$$Q_i = E_{i,m_i} \max(M_{i,m_i}, 0.25), \quad \text{(S29)}$$

where $m_i$ denotes the selected candidate in beat i. For the first tracked beat, the matching factor was set to 1.0. Only consecutively tracked beats were retained for SBP decision making; once the first missing beat occurred, subsequent beats were excluded from the primary decision sequence. The resulting target-wave sequence was smoothed by a 3-beat moving average:

$$\tilde{Q}_i = \frac{Q_{i-1} + Q_i + Q_{i+1}}{3} \quad \text{(S30)}$$

Here, $Q_i$ is the tracked SBP-hotspot target value for beat i, and $\tilde{Q}_i$ is its centered three-beat moving average. At the first and last analyzed beats, for which a complete centered three-beat window was unavailable, the average was calculated from the available adjacent tracked beats without extrapolating values beyond the observed sequence.

SBP was identified as the beat immediately preceding entry into the final stable low-value plateau of the tracked SBP-related morphology. For each candidate beat i, the subsequent tracked values were collected in the forward set $Q_{i,L} = \tilde{Q}_{i+1}, \ldots, \tilde{Q}_{i+L}$, where L is the predefined number of forward beats used to assess whether the post-transition trajectory entered a stable low-value state.

$$D_i = \frac{\max(\tilde{Q}_i - \tilde{Q}_{i+1}, 0)}{SD_i + MAD_i + \varepsilon} \quad \text{(S31)}$$

Here, $SD_i$ is the standard deviation of the forward tracked values in $Q_{i,L}$; $MAD_i$ is the mean absolute difference between consecutive elements of that forward sequence; $[\cdot]_+$ is the positive-part operator; and $\varepsilon$ is a small positive stabilizing constant. A large $D_i$ therefore indicates a marked decline from beat i to beat i+1 followed by a comparatively stable forward trajectory. If all decision values were zero, the algorithm used the largest adjacent decrease in $\tilde{Q}_i$ as a fallback.

The SBP-associated beat was therefore determined by

$$k_{SBP} = \underset{i}{\operatorname{argmax}} D_i, \quad \text{(S32)}$$

and the estimated SBP was assigned as the cuff pressure corresponding to that beat:

$$\hat{P}_{SBP} = P_{cuff}(k_{SBP}). \quad \text{(S33)}$$

During cuff inflation, $P_{cuff}(k_{SBP})$ denotes the cuff pressure at the end of the identified beat, immediately before the tracked target-wave sequence enters its final stable low-value plateau.

Mechanistically, the tracked hotspot represents a late positive oscillometric component that weakens as distal pulsatile contribution becomes delayed, attenuated, and ultimately withdrawn under increasing cuff compression. The final SBP criterion therefore does not use a fixed amplitude threshold. Instead, it identifies the final beat before the tracked SBP-related morphology enters a stable low-value plateau. In practical terms, this implementation localizes SBP from the disappearance pattern of the same right-sided positive hotspot across consecutive beats, rather than from the absolute magnitude of a single beat or from a freely reidentified wavelet maximum.

##### S1.5.6 MAP localization by dual-notch crossover analysis

Conventional oscillometric algorithms estimate mean arterial pressure (MAP) from the maximum oscillation amplitude. This approach is based on the empirical assumption that, when cuff pressure approaches MAP, the mean transmural pressure of the sub-cuff brachial artery approaches zero and the arterial pressure–volume relation enters a relatively high-compliance range, thereby producing the largest arterial volume pulsation and the largest cuff-transmitted pressure oscillation. However, the maximum-amplitude beat can be displaced by transient blood pressure variation, beat-to-beat changes in stroke volume, or baseline drift during cuff measurement.

In morphology-transition oscillometry, MAP was therefore not assigned solely according to the maximum oscillation amplitude. Instead, MAP was determined from a dual-notch crossover on the descending limb of the single-beat oscillometric waveform. After cuff pressure exceeded DBP, a late-diastolic reflection-like notch gradually appeared on the terminal portion of the descending limb and became progressively more prominent. In parallel, the pre-existing dicrotic-like notch on the descending limb weakened as cuff pressure increased and disappeared once cuff pressure exceeded the pressure level corresponding to the dicrotic notch of the arterial pressure waveform. The MAP-related transition was defined as the crossover point at which the weakening dicrotic-like notch and the increasing late-diastolic reflection-like notch showed comparable prominence.

For quantitative localization, the descending limb of each single-beat oscillometric waveform was extracted and subjected to first-derivative analysis. For beat i, let $x_i^D(t)$ denote the portion of the single-beat oscillometric waveform extending from the principal positive peak to the subsequent trough. The first derivative of this descending-limb waveform was calculated as:

$$d_i(t) = \frac{dx_i^D(t)}{dt} \tag{S34}$$

Here, t is local time within beat i, $x_i^D(t)$ has units of cuff-pressure oscillation, and $d_i(t)$ has units of cuff-pressure oscillation per unit time. The derivative was evaluated after the same preprocessing used for the morphology analysis, without introducing an additional pressure-dependent normalization.

Two reproducible local derivative features were then searched within predefined early and late intervals of the descending limb. The early feature corresponded to the dicrotic-like notch, whereas the late feature corresponded to the late-diastolic reflection-like notch. Their prominences relative to the local derivative background were defined as:

$$J_{d,i} = \left|d_i(t_{d,i}) - \overline{d}_i(N_{d,i})\right| \tag{S35}$$

$$J_{\ell,i} = \left|d_i(t_{\ell,i}) - \overline{d}_i(N_{\ell,i})\right| \tag{S36}$$

Here, $t_{d,i}$ and $t_{\ell,i}$ are the derivative-feature locations of the dicrotic-like and late-diastolic notches in beat i, respectively. The sets $N_{d,i}$ and $N_{\ell,i}$ are the corresponding local background neighborhoods, and $\overline{d}_i(N)$ denotes the mean derivative over the specified neighborhood after excluding the feature extremum itself. The absolute differences $J_{d,i}$ and $J_{\ell,i}$ quantify feature prominence without assigning a common sign to derivative extrema whose orientation may vary with waveform polarity. The relative dominance of the late and early derivative features was represented by:

$$C_i = J_{\ell,i} - J_{d,i} \tag{S37}$$

A negative $C_i$ indicates dominance of the dicrotic-like feature, a positive $C_i$ indicates dominance of the late-diastolic feature, and a value near zero indicates comparable prominence of the two features. The MAP-related beat was defined as the analyzed beat at which the absolute prominence difference was smallest

$$i_{MAP} = \underset{i}{\operatorname{argmin}} |C_i| \tag{S38}$$

$$\hat{P}_{MAP} = \overline{P}_{cuff,i_{MAP}} \tag{S39}$$

Here, $J_{MAP}$ is the prespecified beat range in which the crossover was searched, $i_{MAP}$ is the selected beat index, and $\overline{P}_{cuff,i}$ is the mean cuff pressure during beat i. When $C_i$ changed sign between two consecutive beats, the beat with the smaller $|C_i|$ was selected rather than interpolating a pressure value not directly represented by an observed beat.

The dicrotic-like and late-diastolic reflection-like notches were identified as reproducible local derivative features on the descending limb. The cuff pressure corresponding to the beat at which these two derivative-defined notch components crossed over was assigned as the MTO-derived MAP. In recordings with stable beat-to-beat pressure, this crossover typically coincided with or lay close to the maximum-amplitude beat; in recordings with larger beat-to-beat pressure fluctuation, the dual-notch crossover was used preferentially to reduce dependence on amplitude alone.

##### S1.5.7 Time–scale localization

Wavelet analysis provides joint localization in time and scale subject to the Heisenberg uncertainty principle. By constraining DBP and SBP analyses to windows commensurate with the dominant mechanical time scales, the effective time–scale localization is confined to physiologically meaningful regions, such that extrema in $\Delta S$ predominantly reflect mechanical transitions rather than time–scale leakage.

#### *Section S1.6 Experimental protocols in the catheterization cohort*

Five complementary protocols were implemented in the catheterization cohort to address synchronized signal acquisition, site-specific vessel motion analysis, spatial mapping of sub-cuff arterial pressure, venous surrogate assessment of cuff-transmitted deep-tissue pressure, and BP accuracy assessment using a simplified invasive reference-pressure protocol.

To maintain consistent spatial terminology across catheterization protocols, sub-cuff measurement positions were described using two complementary spatial frameworks. First, a seven-site axial reference framework was used to define nominal positions across the cuff-covered brachial arterial segment. The seven reference sites were denoted as Site 0 to Site ⑥ from proximal to distal. Site 0 and Site ⑥ represented the proximal and distal edge-reference positions, respectively, whereas Site ① to Site ⑤ represented five predefined internal sub-cuff sites organized around the central cuff region. Site ③ represented the central cuff region, and the remaining internal sites represented paired proximal and distal reference positions within the cuff-covered segment. The five IVUS measurement sites corresponded to Site ① to Site ⑤. Site ⑤ represented the shared distal-internal reference site used across Protocols 1, 2, and 5. This seven-site framework served as a common coordinate system for protocol design and cross-protocol interpretation and does not imply that all seven sites were sampled in every participant or every protocol.

Second, selected arterial and venous pressure-mapping experiments used a local pressure-peak-zone configuration. In this configuration, pressure-guidewire sensors were concentrated within the central pressure-peak zone (PPZ) rather than distributed across the full cuff-covered segment. For arterial pressure-peak-zone recordings, the mapped region spanned 4 cm when five pressure guidewires were used and 5 cm when six pressure guidewires were used. The venous pressure-peak-zone configuration used the same central 4-cm cuff-relative region as the arterial five-guidewire pressure-peak-zone configuration. These local pressure-peak-zone recordings were used for high-resolution characterization of the dominant cuff-compressed region and should not be interpreted as equivalent to the global Site ① to Site ⑤ whole-cuff configuration.

##### S1.6.1 Protocol 1: Synchronized invasive arterial pressure, oscillometric waveform, and auscultatory signal acquisition during cuff BP measurement

Protocol 1 constituted the primary synchronized acquisition protocol of this study. It was designed to characterize intra-arterial pressure changes and oscillometric waveform behavior during cuff BP measurement by simultaneous acquisition of invasive arterial pressure, cuff pressure, cuff oscillometric waveforms, and auscultatory signals.

In Protocol 1, invasive arterial pressure was recorded from the shared distal-internal reference position, Site ⑤, in all analyzable recordings. This site corresponded to IVUS Site ⑤ in Protocol 2 and to the invasive reference position used in Protocol 5, and served as the principal intra-cuff arterial reference site for cross-protocol comparison. When vascular access, procedural conditions, and recording stability allowed, an additional pressure guidewire was positioned at Site 0, the proximal edge-reference position. This optional proximal channel was used to characterize proximal pressure remodeling and proximal-to-distal waveform separation during cuff pressurization, but it was not required for inclusion in the invasive-reference BP agreement analyses.

Two measurement modes were performed. For inflation-phase BP measurement, the cuff was inflated from baseline at a constant rate of 3 to 5 mmHg/s to a maximum pressure 30 mmHg above invasive SBP, after which the cuff was rapidly released to complete the run. For deflation-phase BP measurement, the cuff was first rapidly inflated to 30 mmHg above invasive SBP and was then deflated at a constant rate of 3 to 5 mmHg/s until cuff release. Invasive arterial pressure waveforms, cuff pressure signals, oscillometric waveforms, and auscultatory signals were recorded continuously and synchronously throughout each measurement run.

For waveform-based analysis, only the slow, approximately linear measurement phase was used: the slow inflation phase in inflation-mode runs and the slow deflation phase in deflation-mode runs. Rapid pressurization and rapid release phases were excluded from BP determination and morphology-transition analysis. Each inflation-mode measurement was repeated twice, and each deflation-mode measurement was also repeated twice. A 1-min rest period was provided after every measurement run to allow BP to return toward baseline before the next acquisition.

This protocol was used to analyze the relationship between oscillometric waveform features and sub-cuff intra-arterial pressure variation during cuff BP measurement, with particular attention to pressure-waveform changes near SBP and DBP. A total of 92 patients undergoing pressure-wire measurements were included in this protocol.

##### S1.6.2 Protocol 2: Site-specific IVUS and pressure measurements at representative sub-cuff locations

Protocol 2 was performed in participants drawn from Protocol 1 and followed the same cuff-inflation and cuff-deflation procedures as Protocol 1. Unlike Protocol 1, which used a fixed distal-internal pressure-guidewire position with an optional proximal reference, Protocol 2 interrogated representative internal sub-cuff positions across repeated measurement sessions. This protocol was designed to investigate the relationship between local vascular wall motion, IVUS-derived CSA, and intra-arterial pressure at defined axial positions beneath the cuff.

For the standard site-specific IVUS protocol, IVUS measurements were obtained at the five internal sites of the seven-site axial reference framework, corresponding to Sites ① to ⑤; the two edge-reference positions, Site 0 and Site ⑥, were not included in this standard five-site mapping protocol. These five internal positions were denoted as Site ① to Site ⑤ from proximal to distal. Site ③ represented the central cuff region, whereas Site ⑤ represented the shared distal-internal reference site used in Protocols 1 and 5. For the acoustic-mechanism analysis shown in Fig. 5a-c, an additional IVUS recording was obtained at the distal edge-reference position, Site ⑥, which was the IVUS location closest to the auscultation sensor.

At each IVUS site, the pressure-guidewire sensor was positioned at the same axial level as the IVUS imaging plane, allowing spatially matched pressure–CSA measurements. During each BP measurement run, the IVUS catheter was maintained at a fixed intravascular position, and continuous IVUS imaging was recorded throughout the cuff measurement process. After all measurements at one site had been completed, the IVUS catheter and pressure guidewire were repositioned to the next predefined site, and the same acquisition procedure was repeated.

IVUS measurements were not obtained at all five sites in every participant because of procedural constraints, including procedure duration, vascular access conditions, and catheter stability. However, all participants undergoing this protocol had IVUS acquisition at one or more sites at or distal to Site ③. Additional sites were examined in subsets of participants to provide more detailed site-resolved mechanistic information across the cuff-covered segment. This protocol provided spatially matched pressure–CSA data for analysis of local vessel-wall motion under cuff compression. A total of 28 patients were included in this protocol. In 4 participants, cuff-inflation measurements and IVUS data were obtained at 13 evenly distributed sub-cuff locations. After correction for wave propagation and temporal registration, the continuous 3D configuration of the sub-cuff brachial artery was reconstructed.

##### S1.6.3 Protocol 3: Multisite brachial arterial pressure measurements beneath the cuff

Protocol 3 was designed to characterize the spatial distribution of intra-arterial pressure beneath the cuff during BP measurement. In this protocol, multiple pressure guidewires were positioned at predefined axial locations along the brachial arterial segment beneath the cuff, enabling simultaneous acquisition of intra-arterial pressure at multiple sites during a single cuff measurement run. Depending on vascular anatomy, access feasibility, and procedural safety considerations, either five or six pressure guidewires were used. In total, this protocol was conducted in 8 patients.

Two multi-guidewire spatial-mapping configurations were used. The first configuration was designed for whole-cuff axial mapping. In this configuration, measurement positions were defined according to the seven-site axial reference framework. When five guidewires were used for whole-cuff axial mapping, the sensors were positioned at Site 0, Site ②, Site ③, Site ④, and Site ⑤. This five-guidewire whole-cuff configuration retained the proximal edge-reference position and sampled four internal sub-cuff sites, whereas Site ① was not sampled in this configuration. When six guidewires were used for whole-cuff axial mapping, the sensors were positioned at Site 0 to Site ⑤, thereby adding Site ① while still excluding the distal edge-reference position, Site ⑥.

The second configuration was designed to interrogate the PPZ beneath the cuff. In this configuration, pressure guidewires were concentrated around the PPZ rather than distributed across the full cuff-covered segment. For arterial pressure-peak-zone mapping, the measured region spanned 4 cm when five guidewires were used and 5 cm when six guidewires were used. This configuration was used to resolve local pressure-waveform changes within the dominant cuff-compressed region and was analyzed separately from the whole-cuff axial mapping configuration.

Signals from all pressure guidewires were recorded synchronously, allowing real-time assessment of pressure variation across the cuff-compressed arterial segment throughout cuff inflation and deflation. The whole-cuff configuration provided a direct representation of axial pressure heterogeneity across the cuff-loaded segment, whereas the PPZ configuration provided higher spatial resolution within the region of maximal external compression. In each patient, two cuff-inflation measurements and two cuff-deflation measurements were obtained at the predefined measurement positions. Thus, each patient contributed two inflation cycles and two deflation cycles for analysis at the corresponding locations.

In a subset of Protocol 3 experiments, additional simultaneous multisite measurements with synchronous recording from five or six pressure guidewires were performed across major arterial segments, including the ascending aorta, subclavian artery, axillary artery, and brachial artery, to examine the longitudinal continuity and propagation pattern of pressure transmission along the proximal-to-distal arterial pathway.

Among the patients included in Protocol 3, two patients who had both arterial and venous access available underwent an additional venous pressure protocol, as described in Protocol 4. Protocol 4 also included three additional venous-only patients who did not contribute invasive arterial pressure recordings.

##### S1.6.4 Protocol 4: Multisite brachial venous pressure measurements beneath the cuff

Protocol 4 was designed to use brachial venous pressure as a surrogate measure of cuff-transmitted deep-tissue pressure beneath the cuff. Brachial venous pressure measurements were performed in 5 patients. Two patients were drawn from the Protocol 3 cohort because both arterial and venous access were available during the catheterization procedure, whereas 3 additional patients underwent venous pressure measurements only. These 3 venous-only patients did not contribute invasive arterial pressure recordings and were not included in invasive-reference BP analyses.

Venous pressure mapping was performed using two configurations. In the whole-cuff internal-site configuration, venous pressure guidewires were positioned at Site ① to Site ⑤, corresponding to the five internal sub-cuff sites used for IVUS imaging. This configuration was used to characterize the axial distribution of cuff-transmitted deep-tissue pressure across the core cuff-covered segment.

In the pressure-peak-zone configuration, venous pressure guidewires were concentrated within the central 4-cm PPZ. This venous configuration used the same cuff-relative measurement region as the arterial five-guidewire pressure-peak-zone configuration in Protocol 3. It was used to resolve the local venous pressure distribution within the dominant cuff-compressed region and was analyzed separately from the global Site ① to Site ⑤ whole-cuff configuration.

Among the three venous-only patients, two underwent the standard five-guidewire venous pressure protocol. In the remaining venous-only patient, three venous pressure guidewires were used because of procedural and access constraints. For the whole-cuff internal-site configuration, one guidewire was maintained at the central site, Site ③, throughout the experiment, whereas the other two guidewires were sequentially repositioned to cover the remaining internal sites. This sequential configuration allowed venous pressure data to be obtained from all five internal sites. The simultaneously recorded Site ③ venous pressure served as the common within-run reference channel for pairing and comparing venous pressure values acquired across sequential site configurations.

The same cuff-inflation and cuff-deflation procedures were used for venous pressure measurements. Within each measurement run, all available venous pressure signals were recorded synchronously. In patients who also underwent arterial pressure mapping, arterial and venous measurements were performed sequentially rather than simultaneously. The venous pressure data were analyzed as surrogate measurements of cuff-transmitted deep-tissue pressure. The pressure-peak-zone venous measurements were spatially matched to the arterial five-guidewire pressure-peak-zone configuration, but no direct arterial–venous pressure pairing or arterial–venous mapping analysis was performed.

During cuff inflation and deflation, the pressures recorded by the venous pressure guidewires primarily reflected the external compressive load transmitted by the cuff to the deep tissues and the perivascular environment, rather than arterial hemodynamic pressure. Because the brachial vein is highly compliant and readily collapsible under external compression, the measured venous pressures were used as a surrogate measure of the compressive load applied to tissues and vessels beneath the cuff [52,53].

##### S1.6.5 Protocol 5: Repeated catheter-based invasive pressure measurements for BP accuracy assessment

Protocol 5 was designed as a simplified invasive pressure-recording protocol to facilitate efficient acquisition of reference-pressure data for BP accuracy assessment. In this protocol, invasive arterial pressure was obtained with a fluid-filled catheter system rather than with multiple pressure guidewires.

The invasive catheter was positioned at Site ⑤ within the seven-site axial reference framework, corresponding to IVUS Site ⑤ in Protocol 2 and to the distal-internal pressure-guidewire position used in Protocol 1, thereby maintaining consistency of the invasive reference position between Protocols 1 and 5.

Catheter preparation, zero calibration, dynamic-response assessment, and definition of invasive reference BP variables followed the procedures described in Section S1.3.5. Each subject then underwent three repeated cuff-deflation measurements. During each run, cuff pressure was recorded synchronously for extraction of the oscillometric waveform, and auscultatory signals were acquired simultaneously as an additional comparator. This protocol provided additional invasive-reference data for comparison of MTO, fixed-ratio amplitude coefficient analysis, and auscultatory BP measurement against invasive arterial pressure as the reference standard. A total of 75 patients were included in this protocol.

#### *Section S1.7 Experimental validation of the axial pressure distribution model*

To assess the validity of the theoretical model from complementary experimental perspectives, two validation approaches were used to evaluate (i) direct cuff–skin contact pressure and (ii) cuff-induced soft-tissue deformation and 3D cuff geometry at predefined cuff pressure levels.

##### S1.7.1 Surface pressure-mapping measurements

Surface pressure-mapping measurements were performed in 50 healthy participants using the flexible external pressure-sensing pad described above (Mini pressure-sensing pad; JQ Industries, Weihai JQ Industrial Technology Co., Ltd., Weihai, China). The sensor matrix contained 1,024 sensing points arranged in a 32 × 32 grid over a 200 × 200 mm active area and was sampled at 100 Hz. During each acquisition, the pad was positioned between the cuff and the upper-arm skin so that the cuff-covered segment was encompassed within the sensing area during controlled cuff inflation. Pressure data were exported as time-stamped framewise arrays and reconstructed into two-dimensional 32 × 32 pressure maps according to the physical sensor layout. For axial-profile analysis, the reconstructed maps were reduced by averaging across the column dimension to obtain row-wise temporal pressure profiles. No linear smoothing was applied along the column dimension. For visualization in the main text, a 3-point moving-average smoothing step was applied along the row dimension after column-wise averaging, whereas the unsmoothed profiles were retained for quality control and comparison. These measurements were used to characterize the spatiotemporal distribution of cuff-transmitted surface pressure and to compare the measured pressure field with the axial pressure-distribution pattern predicted by the model.

##### S1.7.2 Upper-limb CTA–based reconstruction under cuff compression

To characterize cuff–tissue–artery geometry under controlled external compression, upper-limb CTA was performed in 22 healthy volunteers at predefined cuff-pressure levels using retrospective electrocardiogram (ECG) gating to enable phase-resolved reconstruction throughout the cardiac cycle. Prior to scanning, brachial cuff BP was measured three times in each volunteer, and the average DBP and SBP values were used to define five target cuff-pressure conditions: DBP − 20 mmHg, DBP, a pressure level between DBP and SBP, SBP, and SBP + 20 mmHg. Across the cohort, 42 CTA acquisition runs were obtained, including 7 runs at DBP − 20 mmHg, 9 runs at DBP, 8 runs at a pressure level between DBP and SBP, 10 runs at SBP, and 8 runs at SBP + 20 mmHg. Twenty volunteers underwent two CTA acquisitions at different predefined cuff-pressure levels, and 2 volunteers underwent one CTA acquisition. Cuff pressure was maintained at the assigned target level during each scan. All volunteers were scanned using a photon-counting CT system (NeuViz P10, Neusoft Medical Systems). The transverse field of view was 250–300 mm, and the scan length was approximately 200 mm to ensure full coverage of the cuffed upper-arm segment. For photon-counting CT, the acquisition parameters were as follows: tube voltage,

120 kVp; rotation time, 0.27 s; pitch factor, 0.15; and collimation, 584 × 0.274 mm. Images were reconstructed at 33 phases of the R–R interval (3% increments) with a matrix size of 1,024 × 1,024, ClearInfinity level 7, reconstruction kernel H20, a slice thickness of 0.274 mm, and an increment of 0.137 mm. All image data were transferred to the NeuSpace workstation for post-processing. Segmentation of the BP cuff and brachial artery was performed using a semi-automated, AI-assisted workflow. All segmentation results were subsequently reviewed and manually refined to ensure accuracy. The finalized segmentation masks were then fused and reconstructed into 3D visualizations using volume-rendering techniques, enabling assessment of cuff configuration, soft-tissue deformation, and brachial artery geometry under controlled cuff compression.

#### *Section S1.8 Signal processing and analysis*

Cuff pressure signals were processed to extract the oscillometric waveform component used in subsequent waveform-based analyses. Specifically, a third-order Butterworth band-pass filter with a passband of 1–15 Hz was applied to the cuff pressure signal to isolate oscillometric waveforms [57]. The filtered oscillometric waveforms were then used for wavelet-based feature analysis, envelope extraction, and model-related analyses described below. Except for band-pass filtering of the cuff pressure signal for oscillometric waveform extraction, no additional filtering or smoothing was applied to the original invasive arterial pressure waveforms used for physiological interpretation. Signal processing, mathematical modeling, and data visualization were implemented in MATLAB (R2023a, MathWorks).

#### *Section S1.9 Statistical analysis*

The overall agreement, subgroup, repeated-measures, within-participant variability, and error-threshold analyses used 630 paired cuff measurements from 175 participants with invasive arterial pressure recordings. Analyses of arterial pressure changes and their relationships with oscillograms and KS used 124 synchronized measurements from 37 participants. Quantitative stage-specific IVUS-derived CSA analyses used 46 measurements from 25 participants. The three participants with venous pressure recordings only were included exclusively in the venous pressure substudy and were excluded from analyses requiring an invasive arterial reference. Continuous invasive arterial pressure was used as the reference standard. For each measurement, measurement error was calculated as the noninvasive estimate minus the corresponding invasive reference value. Agreement was evaluated separately for SBP, DBP, and MAP using bias, SD of the differences, 95% limits of agreement (LoA; bias ± 1.96 × SD), mean absolute error (MAE), root mean square error (RMSE), and Pearson correlation coefficients.

The primary Bland–Altman analyses were conducted at the measurement level using all paired measurements. To account for repeated measurements within participants, a repeated-measures agreement analysis was performed as a sensitivity analysis. For each method and BP parameter, measurement error was modeled using a linear mixed-effects model with a participant-specific random intercept. The model intercept represented the repeated-measures bias, and the between-participant and within-participant variance components were used to calculate repeated-measures LoA as $\text{bias} \pm 1.96 \times \sqrt{\sigma^2_{\text{between}} + \sigma^2_{\text{within}}}$. The 95% confidence intervals for the lower and upper LoA were estimated using 5,000 participant-level cluster bootstrap resamples, retaining all measurements from each sampled participant. A separate participant-level sensitivity analysis was performed by averaging all paired noninvasive and invasive measurements within each participant before calculating bias and LoA.

Subgroup analyses were conducted at the measurement level according to measurement phase (inflation or deflation), sex, age (<60, 60–69, or ≥70 years), BMI (<24, 24–27.9, or ≥28 kg/m²), and concurrent invasive SBP (<140, 140–159, or ≥160 mmHg). SBP categories were assigned separately for each measurement; consequently, the same participant could contribute measurements to more than one SBP category. Within each subgroup, bias, SD, 95% LoA, MAE, RMSE, and Pearson correlation coefficients were calculated. One-sample Wilcoxon signed-rank tests were used to assess whether the median measurement error differed from zero, and the resulting P values were adjusted within each measurement method using the Benjamini–Hochberg false discovery rate procedure.

Between-subgroup comparisons were based on measurement errors. Mann–Whitney U tests were used for subgroup variables with two levels, and Kruskal–Wallis tests were used for variables with three levels. For each measurement method, global P values were adjusted across the 15 comparisons defined by five subgroup variables and three BP parameters using the Benjamini–Hochberg procedure. When the global FDR-adjusted P value was <0.05 for a three-level subgroup variable, Dunn post hoc tests were performed, with pairwise P values adjusted using the Benjamini–Hochberg procedure.

Within-participant variability was evaluated using method-specific errors from repeated cuff measurements. For each participant, method, and BP parameter, the within-participant error variance was calculated across repeated measurements. A pooled within-participant error SD was then obtained by combining participant-specific variances weighted by their corresponding degrees of freedom. Differences in pooled within-participant error SD between MAA or KS and MTO were calculated by subtracting the MTO value from the corresponding MAA or KS value. Their 95% confidence intervals were estimated using 5,000 paired participant-level cluster bootstrap resamples, retaining all measurements and all methods from each sampled participant.

To further characterize clinically relevant errors, the absolute difference between each noninvasive estimate and its invasive reference was calculated for every measurement. For each method and BP parameter, the proportions of measurements with absolute errors greater than 10, 15, and 20 mmHg were reported descriptively. Statistical analyses were performed using R version 4.5.1 (R Foundation for Statistical Computing, Vienna, Austria), IBM SPSS Statistics version 26 (IBM Corp., Armonk, NY, USA), and MATLAB R2023a (MathWorks, Natick, MA, USA). All hypothesis tests were two-sided. Statistical significance was defined as P<0.05 or, where multiple-comparison correction was applied, an FDR-adjusted P<0.05.

### Supplementary Notes

#### *Section S2.1 Theoretical model of axial cuff contact pressure distribution*

In cuff-based BP measurement, inflation of the cuff transmits external pressure to the underlying vasculature through the intervening soft tissue [3,60]. Because the cuff has finite axial width and edge constraints, the transmitted normal contact pressure beneath the cuff need not be axially uniform [3,31,60]. The aim of the present model is not to predict the absolute contact pressure, but to derive a mechanically self-consistent expression for its relative axial distribution within the effective cuff width. The schematic diagram of the model is shown in Supplementary Fig. 3.

By definition, a normalized pressure-distribution function is introduced:

$$\alpha(z) = \frac{q(z)}{q(0)}, \tag{S40}$$

where $q(z)$ is the normal contact pressure at axial position $z$, and $z = 0$ denotes the cuff center.

##### S2.1.1 Geometric model and mechanical assumptions

The cuff–arm system is treated as axisymmetric. Let $W$ denote the effective axial width of the cuff, with axial coordinate $z$ centered at the origin. As a first-order idealization, the upper arm is represented by an equivalent cylinder of radius $R_a$ derived from the measured circumference $C$ [3,60]. Thus,

$$R_a = \frac{C}{2\pi}. \tag{S41}$$

To obtain an analytical solution, the following assumptions are adopted:

**1. Thin membrane hypothesis:** The cuff wall is modeled as a thin membrane, with bending stiffness neglected relative to membrane action [2,31,61].

**2. Quasi-static inflation:** Inflation is assumed sufficiently slow that inertial effects can be neglected and static equilibrium applies [2,61].

**3. Laplace-type normal equilibrium under contact:** For an axisymmetric membrane shell under a Laplace-type membrane equilibrium, the normal pressure resultant carried by membrane tension was denoted by $p_m(z)$ and expressed as [2,31,61]:

$$p_m(z) = N_\theta \kappa_\theta + N_z^{(m)} \kappa_z, \tag{S42}$$

Here, $N_\theta$ and $N_z^{(m)}$ are the circumferential and axial membrane-force resultants, respectively, with units of force per unit length; $\kappa_\theta$ and $\kappa_z$ are the corresponding principal curvatures, with units of inverse length; and $p_m(z)$ is the net normal pressure supported by membrane tension. The superscript $(m)$ identifies the axial membrane-force resultant and distinguishes it from the number of axial numerical sites used later in the reduced-order model.

**4. Continuous contact assumption:** Within the effective loaded width, gross separation between cuff and arm is neglected, and contact is assumed to remain continuous as a first-order approximation [3,60].

**5. Linear tissue support model:** The surrounding soft tissue is idealized as an equivalent local linear foundation of Winkler type, so that the normal reaction is proportional to the local indentation displacement [62,63]:

$$q(z) = k_t u(z), \tag{S43}$$

Here, $q(z)$ is the local normal contact reaction exerted by the tissue on the cuff membrane, $u(z)$ is the inward normal indentation of the membrane, and $k_t$ is the effective tissue-foundation stiffness with units of pressure per unit displacement. By Newtonian action–reaction, the magnitude of $q(z)$ is also the local contact pressure applied by the cuff to the tissue. In the revised notation, $q(z)$ is not used for the net membrane-tension pressure $p_m(z)$.

**6. Small-slope approximation:** The deformation field is assumed to satisfy $|u'(z)| \ll 1$ and $|u(z)|R_a \ll 1$, so that the curvature expressions may be linearized [2,61].

**7. Equivalent membrane force representation:** Complex structural details of the cuff, including weave, stitching, layer interaction, and friction, are homogenized into effective circumferential and axial membrane force resultants, $N_\theta$ and $N_z^{(m)}$. This homogenization is an author-defined simplification introduced to obtain a tractable closed-form model.

##### S2.1.2 Derivation of the governing equation

In the derivation below, $u(z)$ denotes the signed radial displacement of the cuff mid-surface relative to the undeformed cylindrical reference surface, with $u(z) > 0$ denoting inward indentation toward the arm centerline. The contact pressure $q(z)$ is taken as positive in compression and represents the compressive tissue reaction acting on the cuff membrane; by Newton's third law, it has the same magnitude as the pressure exerted by the cuff on the tissue. The principal curvatures $\kappa_\theta$ and $\kappa_z$ are defined with respect to the outward normal of the deformed cuff mid-surface, and the membrane-force resultants $N_\theta$ and $N_z^{(m)}$ are taken as positive in tension. All expressions below are written consistently under this convention.

The equivalent geometry of the cuff–tissue contact surface is defined as:

$$r(z) = R_a - u(z), \tag{S44}$$

where $u(z)$ denotes the local inward indentation of the membrane into the tissue under the above sign convention.

Under the small-slope approximation and the sign convention stated above, the principal curvatures of the axisymmetric surface are linearized as [2,61]:

$$\kappa_\theta \approx \frac{1}{R_a} + \frac{u(z)}{R_a^2}, \quad \kappa_z \approx -u''(z). \tag{S45}$$

The membrane normal-force balance is then written as [2,31,61]:

$$p - q(z) = p_m(z) = N_\theta \kappa_\theta + N_z^{(m)} \kappa_z \tag{S46}$$

Here, p is the spatially uniform net pressure driving the cuff membrane toward the arm before local tissue reaction is subtracted; $q(z)$ is the local tissue-contact reaction defined in Eq. S43; and $p_m(z)$ is the residual normal pressure supported by membrane tension. This separation prevents the contact pressure $q(z)$ from being used simultaneously for two different physical quantities.

Substituting the curvature relations in Eq. S45 together with $q(z) = k_t u(z)$ into Eq. S46 and rearranging gives the linear membrane–foundation equation:

$$N_z^{(m)} \frac{d^2 u}{dz^2} - K_{eff} u(z) + \left(p - \frac{N_\theta}{R_a}\right) = 0 \tag{S47}$$

Here, $K_{eff}$ is the combined linear restoring coefficient produced by the tissue foundation and the change in circumferential membrane curvature. Equation S47 assumes positive and approximately uniform effective membrane-force resultants, continuous cuff–tissue contact, quasi-static loading, and $|u|/R_a \ll 1$. It determines the axial displacement and relative contact-pressure shape within this linearized boundary-value problem; it does not resolve the full 3D stress field of the cuff, tissue, and arm. To simplify notation, define the equivalent normal stiffness:

$$K_{eff} = k_t + \frac{N_\theta}{R_a^2}. \tag{S48}$$

##### S2.1.3 Boundary conditions and analytical solution

Structural termination of the cuff and edge restraint motivate the following idealized boundary conditions:

$$u\left(\pm\frac{W}{2}\right) = 0\,, \quad u'(0) = 0\,. \tag{S49}$$

The first condition prescribes zero displacement at the effective edges, whereas the second follows from symmetry at the cuff center.

Provided that $N_z^{(m)} > 0$ and $K_{eff} > 0$ , so that λ is real and a physically relevant symmetric center-peaked solution exists, the analytical solution is then:

$$u(z) = u_p\left[1 - \frac{\cosh(\lambda z)}{\cosh\left(\frac{\lambda W}{2}\right)}\right], \tag{S50}$$

Where:

$$\lambda = \sqrt{\frac{K_{eff}}{N_z^{(m)}}}\,, \quad u_p = \frac{p - \frac{N_\theta}{R_a}}{K_{eff}}. \tag{S51}$$

The corresponding contact pressure distribution is $q(z) = ktu(z)$, with the same magnitude applying to the pressure exerted by the cuff on the tissue.

##### S2.1.4 Normalized axial pressure distribution

To suppress dependence on cuff pressure and uncertain material scales, the contact pressure is normalized by its central value:

$$\alpha(z) = \frac{q(z)}{q(0)}, \tag{S52}$$

Substituting the analytical solution and simplifying yields:

$$\alpha(z) = \frac{1 - \frac{\cosh(\lambda z)}{\cosh\left(\frac{\lambda W}{2}\right)}}{1 - \frac{1}{\cosh\left(\frac{\lambda W}{2}\right)}}. \tag{S53}$$

Introducing the dimensionless coordinate $\xi = 2z/W$, and the dimensionless structural parameter $\beta = \lambda W/2$, gives the compact form:

$$\alpha(\xi) = \frac{1 - \dfrac{\cosh(\beta\xi)}{\cosh(\beta)}}{1 - \dfrac{1}{\cosh(\beta)}}, \ \ \xi \in [-1,1]. \tag{S54}$$

###### S2.1.5 Physical significance of parameter β

The dimensionless parameter β integrates the relative effects of the following factors:

• The mechanical impedance of soft tissue against normal indentation.

• The cuff's axial resistance to local deformation.

• The geometric coupling of circumferential membrane forces on axial deformation.

• Constraints at the cuff edges limiting the bulge profile.

A larger β corresponds to a broader, flatter central high-pressure region with a steeper pressure decay near the cuff edges, whereas a smaller β corresponds to a more smoothly curved profile with pressure decreasing progressively from the center toward the edges. Thus, β controls the shape of the normalized pressure distribution rather than its absolute magnitude.

###### S2.1.6 Limitations

This theoretical model characterizes the relative axial distribution of contact pressure beneath a BP cuff rather than predicting its absolute magnitude. Its analytical form is obtained under several idealizations: the cuff is treated as a thin membrane with negligible bending stiffness; inflation is assumed to be quasi-static; the arm is approximated as an axisymmetric cylinder with continuous contact over the effective loaded width; soft tissue is represented by a local linear foundation; effective membrane-force resultants are taken as spatially uniform; and the governing geometry is linearized under small-slope conditions. Within this framework, the combined effects of tissue support, membrane mechanics, and cuff width are condensed into a single dimensionless shape parameter β which governs the normalized axial pressure profile. The model is therefore intended as a first-order mechanistic description of relative pressure distribution. The model also does not account for circumferential anatomical heterogeneity of the upper arm or for dynamic cuff–artery interactions during pulsation. It may not remain valid when soft tissues exhibit pronounced nonlinearity, when contact regions evolve substantially with indentation, or when deformation enters a large-strain regime, in which case higher-order mechanical models or direct experimental measurements are required.

#### *Section S2.2 Mechanistic model of brachial-artery behavior during cuff compression*

###### S2.2.1 Model overview

We developed a unified cuff–brachial artery interaction model to describe the continuous causal chain linking proximal arterial pressure input, local intraluminal pressure propagation, transmural-pressure redistribution, vascular opening and collapse, distal squeeze-emptying, refilling recovery, and cuff-oscillation output under axially nonuniform cuff compression. The model starts from the premise that sub-cuff external pressure is not axially uniform. Instead, it forms a relative pressure peak near the middle of the cuff and decreases toward the proximal and distal cuff edges. Under this external pressure field, local transmural pressure varies jointly with time and axial position, thereby determining luminal area, effective transmission, distal pressure-reservoir behavior, and the volume oscillations ultimately read out by the cuff air chamber.

The model is not a set of mutually independent empirical models for individual figures. It provides a single mechanistic framework. Different illustrative simulations may use different boundary inputs, cuff-pressure protocols, representative physiological states, or pressure-field remodeling parameters to show model predictions under different conditions; however, the principal chain—nonuniform external pressure, pressure propagation, transmural-pressure-dependent area response, negative-transmural-pressure collapse, functional closure, distal emptying/refilling, and volume-to-cuff-pressure readout—remains unchanged.

Throughout the model, proximal arterial pressure is treated as a boundary driving condition, and cuff pressure acts as an axially varying external load on the artery. The intraluminal pressure field and external pressure field together determine transmural pressure, which further determines local luminal area. Under severe compression, the model additionally introduces mechanism-inspired pressure-field remodeling terms to represent distal pressure accumulation, proximal mirror-like reflection, and local pressure waves near $DBP/SBP$ under propagation limitation and impedance mismatch. These terms are not post-processing steps applied to the final oscillogram. They enter the intraluminal pressure field and therefore continue to affect transmural pressure, area, total volume, and recovered cuff-pressure oscillations.

##### S2.2.2 Axial geometry and nonuniform cuff external-pressure field

Let $z$ denote the axial coordinate of the modeled arterial segment, with $z = 0$ at the axial cuff center. Let $L$ denote the total modeled length and $W_{cuff}$ the effective cuff width. The unified model discretizes the cuff-covered brachial artery segment into $N_z = 30$ axial sites, with inter-site spacing $\Delta z = L/(N_z - 1)$. Axial discretization determines not only spatial sampling but also cumulative segmental propagation delay and closure-related additional delay. Therefore, the reported unified model uses a fixed axial discretization so that numerical-discretization differences are not misinterpreted as physiological differences.

The cuff-induced local external pressure field is defined as

$$P_{ext}(z,t) = P_{cuff}(t)\alpha(z). \tag{S55}$$

Here, $P_{ext}(z,t)$ is the local cuff external pressure acting on the vascular wall at axial position $z$ and time $t$, in mmHg; $P_{cuff}(t)$ is the commanded or prescribed cuff pressure, in mmHg; and $\alpha(z)$ is a dimensionless normalized axial external-pressure distribution function ranging from 0 to 1.

Following a membrane-theory-inspired center-high/edge-low form, $\alpha(z)$ is written as

$$\alpha(z) = \begin{cases} \dfrac{1 - \cosh(\beta\xi)/\cosh(\beta)}{1 - 1/\cosh(\beta)}, & |z| \le W_{cuff}/2, \\ 0, & |z| > W_{cuff}/2, \end{cases} \tag{S56}$$

$$\xi = \frac{2z}{W_{cuff}}. \tag{S57}$$

Here, $\xi$ is the dimensionless normalized axial coordinate; $\xi = -1$ and $\xi = 1$ correspond to the proximal and distal cuff edges, respectively; and $\beta$ is a dimensionless axial shape parameter controlling how strongly external pressure is concentrated toward the cuff center. We used $\beta = 3.5$ as a representative value. Changing $\beta$ alters the sharpness of the axial external-pressure distribution, but within the tested range it does not alter the core mechanistic sequence by which nonuniform external pressure drives transmural-pressure redistribution, vascular collapse, distal emptying, and oscillogram formation [3,60,61]. Equations S56 and S57 are the numerical implementation of the normalized pressure shape derived in Section S2.1. They retain the analytical expression inside the cuff-covered interval and set the applied cuff pressure to zero outside that interval. Their purpose is to define the spatial external-pressure input used at every numerical site and time step, not to introduce a second independent pressure-distribution theory.

**Proximal boundary pressure input and cuff-pressure protocol.** The proximal intraluminal pressure $P_{in,prox}(t)$ is treated as a boundary condition rather than as an independent pressure waveform regenerated inside the model. Depending on the simulation purpose, this boundary input can be supplied by a standardized arterial-pressure template, a measured invasive arterial pressure waveform, or a constant-pressure control. The standardized template provides a physiologically plausible and controllable pulsatile input; measured input preserves subject-specific waveform morphology; and constant input is used only as a mechanistic control to isolate the effects of nonuniform external pressure and transmural-pressure changes on vascular collapse. For cardiac period $T$, the within-beat phase is defined as

$$\phi(t) = \frac{t}{T} + \phi_{shift} - \lfloor \frac{t}{T} + \phi_{shift} \rfloor \tag{S58}$$

Here, T=60/HR is the cardiac-cycle duration, HR is the prescribed heart rate in beats per minute, $\phi_{shift}$ is the dimensionless phase shift of the input waveform, and $\lfloor \cdot \rfloor$ denotes the floor function. Equation S58 extracts the fractional part of the phase argument and therefore ensures $0 \leq \phi(t) < 1$ without using the software-specific mod operator.

When the standardized template is used, the unnormalized pressure template is

$$\Psi(\phi) = M(\phi) + S(\phi) - N(\phi) + R(\phi) + U(\phi). \tag{S59}$$

Here, $\Psi(\phi)$ is the unnormalized arterial-pressure template; $M(\phi)$ is the main systolic peak; $S(\phi)$ is the shoulder or reflected-shoulder component; $N(\phi)$ is the notch term, entered with a negative sign because it represents a pressure-lowering component; $R(\phi)$ is the post-notch rebound component; and $U(\phi)$ is the late-diastolic runoff term. All components are dimensionless template functions.

The main peak is represented by a gamma-shaped waveform:

$$M(\phi) \propto \phi^{a_1} \exp(-b_1 \phi). \tag{S60}$$

Here, $a_1$ and $b_1$ control the upstroke steepness and decay speed of the main systolic peak, respectively. Both are dimensionless template-shape parameters. The proportionality symbol indicates that this component is subsequently normalized together with the full template, so no separate absolute amplitude is assigned.

The shoulder, notch, and post-notch rebound are represented by local Gaussian functions:

$$S(\phi) = a_{shoulder} \exp\left[-\frac{(\phi - 0.30)^2}{2(0.055)^2}\right], \tag{S61}$$

$$N(\phi) = a_{notch}\exp\left[-\frac{(\phi - 0.42)^2}{2(0.012)^2}\right], \tag{S62}$$

$$R(\phi) = a_{rebound}\exp\left[-\frac{(\phi - 0.465)^2}{2(0.018)^2}\right]. \tag{S63}$$

Here, $a_{shoulder}$, $a_{notch}$, and $a_{rebound}$ are the dimensionless amplitudes of the shoulder, notch, and post-notch rebound, respectively. The numerical centers and widths specify the phase locations and durations used in the reported implementation. These quantities define the prescribed boundary waveform only and are not estimated vascular-wall material parameters.

The late-diastolic runoff term is

$$U(\phi) = 0.22\exp[-4.2\max(\phi - 0.46,0)]. \tag{S64}$$

Here, $U(\phi)$ represents an exponential late-diastolic decay component; $\max(\phi - 0.46,0)$ makes this term primarily active after phase 0.46; and 0.22 and 4.2 are template amplitude and decay-rate parameters used only to generate the standardized boundary input.

After normalization and mild smoothing, the template is linearly mapped to a prescribed $SBP/DBP$ range to obtain the proximal boundary pressure:

$$P_{in,prox}(t) = P_{DBP} + (P_{SBP} - P_{DBP})\tilde{\Psi}[\phi(t)]. \tag{S65}$$

Here, $P_{in,prox}(t)$ is the proximal boundary intraluminal pressure, in mmHg; $P_{SBP}$ and $P_{DBP}$ are the input systolic and diastolic pressures, in mmHg; and $\tilde{\Psi}(\phi)$ is the normalized and smoothed pressure template, ranging from 0 to 1. This mapping ensures that the minimum of the template corresponds to $P_{DBP}$ and the maximum corresponds to $P_{SBP}$. In the reported implementation, the composite template was shifted to a nonnegative range, normalized to unity, transformed by an exponent of 0.92, and lightly smoothed with an 11-point Savitzky–Golay filter before being mapped to the prescribed SBP–DBP range. These operations standardize the proximal boundary waveform and do not add an independent cuff–artery mechanism.

$SBP$, $DBP$, and heart rate are input or protocol conditions, not fitted mechanistic parameters. Cuff pressure increases at approximately 3–5 mmHg/s according to the clinical measurement protocol and reaches approximately 30 mmHg above the input or reference $SBP$, thereby covering the full compression process from around $DBP$ to above $SBP$. The standardized template-shape parameters provide only a controllable boundary input; measured and constant inputs do not depend on this template.

##### S2.2.3 Distributed pressure propagation and transmural-pressure definition

Pressure propagation was represented by delayed site-to-site driving while the upstream segment remained functionally transmissive. When the upstream transmission state fell below the prescribed threshold, the downstream pressure state was no longer forced directly by the proximal waveform and instead relaxed toward an effective distal reservoir. This reduced-order rule was implemented as follows. Baseline propagation from the proximal to distal direction is determined by the baseline pulse-wave velocity $c_0$. The baseline propagation delay between adjacent axial sites is

$$\Delta\tau_0 = \frac{\Delta z}{c_0}. \tag{S66}$$

Here, $\Delta\tau_0$ is the baseline propagation delay between adjacent axial sites, in seconds; $\Delta z$ is the inter-site spacing, converted from mm to m during calculation; and $c_0$ is the baseline pulse-wave velocity, in m/s. The cumulative propagation delay arriving at axial site $i$ is denoted by $\tau_i$.

In addition to baseline propagation, the model allows propagation delay to increase as local closure deepens. Local occlusion severity is defined as

$$\Omega_i(t) = I_i(t)\big(1 - a_i(t)\big)^{\nu_A}\left(\frac{P_{-,i}(t)}{P_{tm,ref}}\right)^{\nu_P}. \tag{S67}$$

Here, $\Omega_i(t)$ is the dimensionless local occlusion severity at axial site $i$; $I_i(t)$ is an occlusion-condition indicator equal to 1 when $\alpha(z_i) > 0$ and $P_{tm,i} < 0$, and 0 otherwise; $a_i(t) = A_i(t)/A_{0,i}(t)$ is local relative area; $A_i(t)$ is local luminal area, in mm²; $A_{0,i}(t)$ is the corresponding zero-pressure reference area, in mm²; $P_{-,i}(t) = \max\big[-P_{tm,i}(t), 0\big]$ is the negative-transmural-pressure magnitude; $P_{tm,ref}$ is the reference pressure used to normalize negative transmural pressure, in mmHg; and $\nu_A$ and $\nu_P$ control the nonlinear contributions of area closure and negative transmural pressure to additional delay.

The segmental propagation delay is updated as

$$\Delta\tau_i = \Delta\tau_0\big(1 + g_{delay,occ}\Omega_i\big). \tag{S68}$$

Here, $\Delta\tau_i$ is the updated delay of the $i$th adjacent axial segment, and $g_{delay,occ}$ is a dimensionless closure-related delay gain. This additional delay represents progressive slowing of pressure transmission as the compressed region deepens. To avoid differences in axial discretization creating non-comparable cumulative delays, the unified model reported here uses 30 fixed axial sites. A representative baseline PWV of 6 m/s was used, with mechanistic exploration within a physiologically plausible range.

The local transmural pressure at each axial site is defined as

$$P_{tm}(z,t) = P(z,t) - P_{ext}(z,t). \tag{S69}$$

Here, $P_{tm}(z,t)$ is local transmural pressure, in mmHg; $P(z,t)$ is local intraluminal pressure, in mmHg; and $P_{ext}(z,t)$ is local external cuff pressure, in mmHg. $P_{tm} > 0$ indicates that intraluminal pressure exceeds external pressure and the vessel tends to remain open; $P_{tm} < 0$ indicates that external pressure exceeds intraluminal pressure and the vessel tends to compress or collapse.

When upstream transmission remains open, local intraluminal pressure is driven primarily by the delayed proximal input. When upstream transmission is functionally interrupted, local pressure decays toward an effective distal reservoir pressure $P_{res}^{eff}$ with time constant $\tau_{runoff}$:

$$P_i^{n+1} = \begin{cases} P_{in,prox}(t_n - \tau_i), & K_{i-1} > K_{crit}, \\ P_i^n + \dfrac{\Delta t}{\tau_{runoff}}\big(P_{res}^{eff} - P_i^n\big), & K_{i-1} \le K_{crit}. \end{cases} \tag{S70}$$

Here, $P_i^n$ and $P_i^{n+1}$ are the intraluminal pressures at axial site $i$ at the current and next time steps; $t_n$ is the $n$th time point; $\tau_i$ is the cumulative propagation delay to site $i$; $K_{i-1}$ is the effective conductance of the upstream adjacent site; $K_{crit}$ is the functional transmission-cutoff threshold; $\Delta t$ is the time step; $\tau_{runoff}$ is the time constant for local pressure to decay toward the effective distal reservoir pressure after interruption; and $P_{res}^{eff}$ is an effective distal reservoir pressure, not venous pressure. Equation S70 is an algorithmic reduced-order pressure-propagation rule. It switches between a delayed proximal input and relaxation toward an effective distal reservoir according to the upstream transmission state. It is not a finite-difference discretization of the complete one-dimensional momentum and continuity equations, and no local volumetric flow rate is inferred directly from this state update.

##### S2.2.4 Opening law under positive transmural pressure

To describe vascular behavior under positive transmural pressure, the model uses a Langewouters-type pressure-area law as the opening segment [64–66]. To avoid confusion with the closure-memory variable, the systolic/diastolic interpolation state is denoted by $q_{phase}$. The local normalized pressure is

$$p_n = \min\{1, \max\left[0, \frac{P - P_{DBP}}{P_{SBP} - P_{DBP}}\right]\} \quad \text{(S71)}$$

Here, $p_n$ is a dimensionless pressure-level coordinate bounded between 0 and 1; P is the local or prescribed arterial pressure used in the phase-dependent interpolation; and $P_{SBP}$ and $P_{DBP}$ are the reference systolic and diastolic pressures. The nested minimum and maximum functions are the mathematical form of the bounded normalization.

The upstroke gate is

$$g_\uparrow = \frac{1}{1 + \exp\left(-\dot{P}/s_{\dot{P}}\right)}. \quad \text{(S72)}$$

Here, $g_\uparrow$ is the pressure-upstroke weight, ranging from 0 to 1; $\dot{P}$ is the first derivative of pressure with respect to time, in mmHg/s; and $s_{\dot{P}}$ is the slope-scale parameter that controls smooth switching between upstroke and downstroke segments, in mmHg/s.

The phase state is

$$q_{phase} = g_\uparrow p_n^{\gamma_\uparrow} + (1 - g_\uparrow) p_n^{\gamma_\downarrow}. \quad \text{(S73)}$$

Here, $q_{phase}$ is the local phase-state variable used to smoothly interpolate between systolic and diastolic segments; $\gamma_\uparrow$ and $\gamma_\downarrow$ are the nonlinear exponents for the upstroke and downstroke segments.

A smootherstep function gives the systolic-segment weight:

$$w = 6q_{phase}^5 - 15q_{phase}^4 + 10q_{phase}^3. \quad \text{(S74)}$$

Here, $w$ is the systolic-segment weight ranging from 0 to 1. The smootherstep function provides smooth endpoint derivatives, avoiding abrupt transitions between systolic and diastolic area laws.

The zero-pressure reference area is

$$A_0 = (1-w)A_{0,dia} + wA_{0,sys}. \tag{S75}$$

Here, $A_0$ is the current phase-dependent zero-pressure reference area, in mm²; $A_{0,dia}$ and $A_{0,sys}$ are the diastolic- and systolic-segment zero-pressure reference areas, in mm².

Let $x = \max(P_{tm}, 0)$ denote the positive-transmural-pressure input. For each segment, the Langewouters-type area law is

$$A_b(x) = A_m\left[\frac{1}{2} + \frac{1}{\pi}\tan^{-1}\left(\frac{x-p_0}{p_1}\right)\right]. \tag{S76}$$

Here, $x$ is the positive-transmural-pressure input, in mmHg; $A_b(x)$ is the segment-specific equilibrium opening area, in mm²; $A_m$ is an area-scale parameter, in mm²; $p_0$ is the location parameter, in mmHg; and $p_1$ is the slope parameter, in mmHg. The positive-transmural-pressure opening area is then

$$A_{open} = (1-w)A_d(x) + wA_s(x). \tag{S77}$$

Here, $A_{open}$ is the opening-segment area under positive transmural pressure, and $A_d(x)$ and $A_s(x)$ are the diastolic- and systolic-segment Langewouters-type area functions. This component uses an opening segment based on measured large-artery pressure-area behavior as its direct mathematical source; its interpretation in brachial-artery collapse/opening behavior and modern tube-law parameterization is supported by related studies.

##### S2.2.5 Hybrid collapse law under negative transmural pressure

Under negative transmural pressure, the model uses a mechanistic hybrid collapse configuration. The core premise is that once the vessel enters a sufficiently negative-transmural-pressure range, the dominant behavior is no longer rapid repetitive reopening, but persistent collapse, residual slot-like patency, and further closure promoted by emptying. This component is intended to capture persistent collapse, slot-like residual patency, and emptying-assisted collapse under negative transmural pressure [65–67].

To avoid symbol conflicts, the closure-memory variable is denoted by $\chi_c$, and the residual-patency variable is denoted by $\lambda_p$. The negative-transmural-pressure magnitude is

$$P_{-} = \max(-P_{tm}, 0). \tag{S78}$$

Here, $P_{-}$ is the negative-transmural-pressure magnitude, in mmHg; when $P_{tm} \geq 0$, $P_{-} = 0$.

The contact-closure reference area and minimal residual leak area are

$$A_{contact} = f_{contact}A_0, \qquad A_{leak} = f_{leak}A_0. \tag{S79}$$

Here, $A_{contact}$ is the reference area for wall contact or near-contact; $A_{leak}$ is the minimal residual leak area retained numerically after functional closure; and $f_{contact}$ and $f_{leak}$ are dimensionless area fractions relative to $A_0$.

The geometric-collapse equilibrium area is

$$A_{geom}^{eq} = A_{contact} + \frac{A_0 - A_{contact}}{1 + (P_{-}/P_{buck})^{m_{buck}}}. \tag{S80}$$

Here, $A_{geom}^{eq}$ is the equilibrium area determined by the geometric collapse tendency under negative transmural pressure; $P_{buck}$ is the collapse reference pressure, in mmHg; and $m_{buck}$ is a dimensionless nonlinearity exponent.

The target value of the closure-memory variable is

$$\chi_{c,\infty} = 1 - \exp\left[-\left(P_{-}/P_{\chi}\right)^{n_{\chi}}\right]. \tag{S81}$$

Here, $\chi_{c,\infty}$ is the target value of the closure-memory variable $\chi_c$, ranging from 0 to 1; larger $\chi_c$ indicates a more persistent local closed state; $P_{\chi}$ is the reference negative transmural pressure for closure-memory formation, in mmHg; and $n_{\chi}$ is a nonlinear exponent.

The emptying-driven decay rate of the residual-patency variable $\lambda_p$ is

$$k_{drain} = k_{drain,0}[1 - \exp(-(P_{-}/P_{drain})^{n_{drain}})]\left(1 + k_{drain,\chi}\chi_c\right). \tag{S82}$$

Here, $k_{drain}$ is the emptying-driven decay rate of residual patency; $k_{drain,0}$ is the baseline drainage-drive coefficient; $P_{drain}$ is the drainage-drive reference pressure, in mmHg; $n_{drain}$ is a nonlinear exponent; and $k_{drain,\chi}$ represents the enhancement of drainage drive by closure memory.

The residual slot area is

$$A_{slot} = A_{leak} + \lambda_p(A_{contact} - A_{leak})(1 - \chi_c)^{\eta_{slot}}. \tag{S83}$$

Here, $A_{slot}$ is the residual slot-like channel area; $\lambda_p$ is the residual-patency variable, with $\lambda_p = 1$ representing a relatively patent residual channel and $\lambda_p = 0$ representing decay to the minimal leak area; and $\eta_{slot}$ controls the nonlinear decrease of slot area with closure memory.

The internal area state of the negative-pressure segment is

$$A_{neg} = (1 - \chi_c)A_{geom}^{eq} + \chi_c A_{slot}. \tag{S84}$$

Here, $A_{neg}$ is the internal area state of the negative-transmural-pressure segment; $A_{geom}^{eq}$ is the geometric-collapse reference area; $A_{slot}$ is the residual slot area; and $\chi_c$ weights the transition between geometric-collapse and residual-slot states.

The three internal states were updated explicitly at each time step. Their update rules were

$$A_{geom}^{n+1} = \begin{cases} A_{geom}^{n} - \Delta t \max\left(\dfrac{A_{geom}^{n} - A_{geom}^{eq,n}}{\tau_{geom,neg}}, 0\right), & P_{tm}^{n} < 0, \\ A_{geom}^{n} + \dfrac{\Delta t}{\tau_{geom,open}^{eff,n}}\left(A_0 - A_{geom}^{n}\right), & P_{tm}^{n} \geq 0, \end{cases} \tag{S85}$$

$$\chi_c^{n+1} = \begin{cases} \chi_c^{n} + \Delta t \max\left(\dfrac{\chi_{c,\infty}^{n} - \chi_c^{n}}{\tau_{\chi,neg}}, 0\right), & P_{tm}^{n} < 0, \\ \chi_c^{n} \exp\left(-\dfrac{\Delta t g_{open}^{n}}{\tau_{\chi,open}}\right), & P_{tm}^{n} \geq 0, \end{cases} \tag{S86}$$

$$\lambda_p^{n+1} = \begin{cases} \lambda_p^{n} \exp(-\Delta t k_{drain}^{n}), & P_{tm}^{n} < 0, \\ \lambda_p^{n} + \dfrac{\Delta t}{\tau_{p,refill}}\left(1 - \lambda_p^{n}\right), & P_{tm}^{n} \geq 0. \end{cases} \tag{S87}$$

Here, superscript $n$ denotes the current time step; $\Delta t$ is the numerical time step; $A_{geom}^{eq,n}$ is the instantaneous geometric-collapse target; $\tau_{geom,neg}$ is the collapse time constant under negative transmural pressure; and $\tau_{geom,open}^{eff,n}$ is the effective reopening time constant. The target $\chi_{c,\infty}^{n}$ and the time constants $\tau_{\chi,neg}$ and $\tau_{\chi,open}$ govern accumulation and release of closure memory. The opening drive $g_{open}^{n}$ was calculated as one minus the exponential of the negative 1.2 power of the normalized positive transmural pressure. In this definition, the positive part of transmural pressure is $\max(P_{tm}, 0)$, and $P_{open}$ is the positive-transmural-pressure scale for reopening. The quantity $k_{drain}^{n}$ is the state-dependent drainage rate of the residual-patency variable, and $\tau_{p,refill}$ is its refill time constant. After each update, the three states were restricted to their admissible ranges to prevent negative area or state values outside $[0,1]$.

The closure-mixing function is

$$G_{neg} = 1 - \exp[-(P_{-}/P_{close})^{n_{close}}]. \tag{S88}$$

Here, $G_{neg}$ is the negative-pressure closure-mixing function, ranging from 0 to 1; $P_{close}$ is the closure-mixing reference pressure, in mmHg; and $n_{close}$ is the closure-mixing nonlinear exponent.

The collapse-segment area is

$$A_{close} = \left(1 - G_{neg}\right)A_0 + G_{neg}A_{neg}. \tag{S89}$$

Here, $A_{close}$ is the negative-transmural-pressure collapse-segment area. When $G_{neg}$ approaches 0, the area approaches $A_0$; when $G_{neg}$ approaches 1, the area approaches $A_{neg}$.

A logistic function is used to smoothly switch between the opening and collapse segments:

$$s_w = \frac{1}{1 + \exp[-(P_{tm} - P_{sw})/w_{sw}]}. \tag{S90}$$

Here, $s_w$ is the smooth switching weight between the opening and collapse segments; $P_{sw}$ is the switching-center pressure, in mmHg; and $w_{sw}$ is the switching width, in mmHg. Larger $s_w$ gives greater contribution from the opening segment, whereas smaller $s_w$ gives greater contribution from the collapse segment.

The equilibrium area is

$$A_{eq} = s_w A_{open} + (1 - s_w)A_{close}. \tag{S91}$$

Here, $A_{eq}$ is the target equilibrium area determined jointly by transmural pressure and state variables; $A_{open}$ is the positive-transmural-pressure opening-segment area; and $A_{close}$ is the negative-transmural-pressure collapse-segment area.

Area dynamics are updated as

$$A_i^{n+1} = A_i^n + \frac{\Delta t}{\tau_{A,i}^n}\left(A_{eq,i}^n - A_i^n\right) + \frac{D_A \Delta t}{(\Delta z)^2}\left(A_{i+1}^n - 2A_i^n + A_{i-1}^n\right) \tag{S92}$$

Here, $A_i^n$ is the luminal area at axial site $i$ and time step $n$; $A_{eq,i}^n$ is the corresponding target area from the unified open/closed pressure–area relation; $\tau_{A,i}^n$ is the local relaxation time constant; $\Delta z$ is the axial site spacing; and $D_A$ is an axial numerical-regularization coefficient with units of length squared per unit time. The final term suppresses isolated site-to-site numerical discontinuities and is not interpreted as an independent biological diffusion process. Zero-normal-gradient boundary conditions were applied at the two axial ends when the second spatial difference was evaluated.

##### S2.2.6 Functional-closure criterion and effective closure boundary

Because the continuous collapse equations do not necessarily drive local area exactly to zero, the model distinguishes mathematical continuous area from physiologically interpretable functional closure [65–67]. Local effective conductance is defined as

$$K_i = \left(\frac{A_i}{A_{0,i}}\right)^{m_{cond}}. \tag{S93}$$

Here, $K_i$ is the dimensionless effective conductance at site $i$; $A_i/A_{0,i}$ is relative luminal area; and $m_{cond}$ is the area-to-conductance exponent used to map continuous area changes onto effective hydraulic transmission.

When $K_i \leq K_{crit}$, or equivalently when

$$\frac{A_i}{A_{0,i}} \leq \zeta_{close} = K_{crit}^{1/m_{cond}}, \tag{S94}$$

that location is interpreted as functionally closed. Here, $\zeta_{close}$ is the relative-area threshold for functional closure, and $K_{crit}$ is the conductance cutoff threshold. This thresholding step does not replace the continuous collapse law; it maps the output of the continuous equations to a physiologically interpretable effective closure boundary.

##### S2.2.7 Distal squeeze-emptying and distal refilling recovery

Distal emptying is based on an RC/Windkessel-like distal reservoir–compliance–outflow-resistance framework [12,15,68]. The initiation of emptying dominance, establishment of an emptying episode, and refilling/re-entry transitions are implemented as mechanistic state transitions within the propagation–collapse coupling framework.

When local collapse under negative transmural pressure transitions from rapid collapse dominance to emptying-dominated decay, the distal region is treated as entering a distal-emptying state. During this stage, some residual connectivity may persist between proximal and distal regions, but diastolic hydraulic connectivity is too weak to dominate distal pressure behavior. Distal squeeze-emptying therefore becomes the principal mechanism.

During emptying, distal pressure $P_d$ follows an RC/Windkessel-like form:

$$\frac{dP_d}{dt} = -\frac{P_d - P_{res}^{eff}}{\tau_{empty}}. \tag{S95}$$

$$\tau_{empty} = R_{out}^{eff} C_d^{eff}. \tag{S96}$$

Here, $P_d$ is distal pressure during emptying, in mmHg; $P_{res}^{eff}$ is the effective distal reservoir or attractor pressure within the current emptying episode, in mmHg; $\tau_{empty}$ is the distal emptying time constant, in seconds; $R_{out}^{eff}$ is effective distal outflow resistance; and $C_d^{eff}$ is effective distal compliance. Their product determines the pressure-decay time scale within the episode.

The effective distal reservoir pressure is

$$P_{res}^{eff} = P_{res,0}^{eff} + k_{res}\eta_{empty}. \tag{S97}$$

Here, $\eta_{empty}$ is the degree of distal emptying, ranging from 0 to 1; $P_{res,0}^{eff}$ is the baseline effective distal reservoir pressure, in mmHg; and $k_{res}$ is the coefficient by which emptying degree modulates the effective reservoir pressure, in mmHg.

Effective distal compliance is

$$C_d^{eff} = \max\left[C_{d,min}^{eff}, C_{d,0}^{eff}\left(\frac{A_d}{A_{0,d}}\right)^{n_C}\right] \tag{S98}$$

$$R_{out}^{eff} = \max\left[R_{out,min}^{eff}, \frac{R_{out,0}^{eff}\left(1 + k_{R,occ}\eta_{empty}^{n_R}\right)}{1 + k_{R,sq}S_{occ}}\right] \tag{S99}$$

Here, $A_d/A_{0,d}$ is the distal area ratio at the onset of an emptying episode; $C_{d,0}^{eff}$ and $C_{d,min}^{eff}$ are the baseline and lower-bound effective distal compliances; $n_C$ controls the dependence of effective compliance on distal area; $R_{out,0}^{eff}$ and $R_{out,min}^{eff}$ are the baseline and lower-bound effective outflow resistances; $\eta_{empty}$ is the normalized emptying state; $S_{occ}$ is the effective occlusion severity; and $k_{R,occ}$, $k_{R,sq}$, and $n_R$ are dimensionless shape parameters. The corresponding time constant is $\tau_{empty} = R_{out}^{eff}C_d^{eff}$. At the onset of each emptying episode, $P_{res}^{eff}$, $C_d^{eff}$, $R_{out}^{eff}$, and $\tau_{empty}$ were evaluated from the current distal area and compression state and were then held fixed until that episode ended. This episode-frozen implementation prevents rapid within-episode parameter fluctuations and represents a lumped effective distal reservoir rather than a directly measured distal vascular bed.

In the numerical implementation, the effective reservoir pressure, compliance, outflow resistance, and emptying time constant are determined at the start of each emptying episode from local distal area and compression state, and are then held fixed within that episode. Distal refilling recovery is a separate axial state transition: once local openness recovers, even a weakly reopened pathway can allow the larger systolic proximal-to-distal pressure gradient to drive blood back into the distal region. Emptying exit therefore does not represent simple stopping; it represents restoration of distal refilling capacity.

##### S2.2.8 Volume integration, recovered cuff pressure, and oscillogram extraction

The mapping from subcuff arterial volume to recovered cuff pressure follows the cuff air-chamber readout approach used in oscillometric modeling [12,15,68]. The equations use an ideal-gas approximation and a first-order chamber readout to convert distributed vascular volume changes into observable cuff-pressure oscillations.

The model pressure field $P_{model}(z,t)$ and area field $A_{model}(z,t)$ determine total subcuff arterial volume:

$$V_{total}(t) = \frac{1}{1000}\sum_{z \in cuff} A_{model}(z,t)\Delta z. \tag{S100}$$

Here, $V_{total}(t)$ is total arterial volume under the cuff, in mL; $A_{model}(z,t)$ is the model-layer local luminal area, in mm$^2$; $\Delta z$ is axial spacing, in mm; the summation over $z \in cuff$ indicates that only cuff-covered axial sites are integrated; and division by 1000 converts mm³ to mL because $1\ \mathrm{mL} = 1000\ \mathrm{mm}^3$.

The total volume is first smoothed by a moving average:

$$V_a(t_n) = \frac{1}{N_{W,n}} \sum_{j \in W_n} V_{total}\left(t_j\right) \tag{S101}$$

Here, $V_{total}(t_j)$ is the axially integrated arterial volume at sample j; $W_n$ is the set of available samples within the centered smoothing window around $t_n$; and $N_{W,n} = |W_n|$ is the number of samples in that window. At record boundaries, only available samples were used. The absolute air-chamber pressure corresponding to the recovered cuff pressure is

$$P_{abs}(t) = P_{cuff,rec}(t) + 760. \tag{S102}$$

Here, $P_{abs}(t)$ is chamber absolute pressure, in mmHg; $P_{cuff,rec}(t)$ is recovered cuff gauge pressure, in mmHg; and 760 mmHg approximates atmospheric pressure for conversion from gauge to absolute pressure.

The recovered cuff pressure satisfies

$$\frac{dP_{cuff,rec}}{dt} = \frac{dP_{cuff,cmd}}{dt} + \frac{P_{abs}(t)}{V_{0,cuff}} \dot{V}_a(t). \tag{S103}$$

Here, $dP_{cuff,rec}/dt$ is the rate of change of recovered cuff pressure; $dP_{cuff,cmd}/dt$ is the rate of change of commanded cuff pressure; $V_{0,cuff}$ is the equivalent cuff air volume, in mL; $\dot{V}_a$ is the rate of change of subcuff arterial volume, in mL/s; and $P_{abs}/V_{0,cuff}$ is the coupling coefficient from chamber-volume change to pressure change. This equation treats arterial volume change as an opposing perturbation to effective cuff-chamber volume.

The initial condition is

$$P_{cuff,rec}(t_0) = P_{cuff,cmd}(t_0). \tag{S104}$$

This initial condition aligns the recovered cuff-pressure trajectory with the commanded cuff pressure at the start of simulation. After recursive integration, a short-window smoothing with $N_P$ is applied to obtain the recovered cuff pressure.

**Oscillation extraction and detrending.** Filtering and detrending are used only to extract oscillatory components from the recovered cuff pressure in the same manner as the experimental cuff signal. They do not constitute an empirical criterion. In this model, the physical source of the oscillogram remains axial local area change and its coupling to cuff volume-pressure readout. The recovered cuff pressure is processed using the same oscillation extraction and detrending workflow used for experimental cuff signals, allowing simulated and measured oscillograms to be compared under the same readout definition. This step is an output-layer readout operation; it does not introduce a new physiological mechanism or any empirical blood-pressure rule.

##### S2.2.9 Mechanism-inspired pressure-field remodeling under severe compression

The principal model predicts that, when cuff compression limits local transmission and creates impedance mismatch between the distal compliant reservoir and proximal input, the intraluminal pressure field may develop distal pressure accumulation, proximal mirror-like reflection, and local pressure waves near $DBP/SBP$ [69–71]. These intrabeat fine-pressure structures are not fully resolved by nonuniform external pressure, baseline propagation, the pressure-area relation, and the collapse/emptying chain alone. Therefore, they are included as mechanism-inspired pressure-field remodeling terms in the intraluminal pressure field rather than as final oscillogram post-processing or graphical modifiers. Once added to $P(z,t)$, these terms continue to affect transmural pressure, vascular area, total volume, and recovered cuff-pressure oscillations.

**Distal pressure elevation with finite dynamic response.** Distal pressure elevation represents the model-predicted distal pressure-field enhancement under propagation limitation, local impedance mismatch, and distal pressure accumulation. Rather than applying instantaneous amplification, distal pressure enhancement is written as a dynamic state with finite rise and decay time constants to avoid nonphysiological pressure jumps between adjacent beats. This module acts distal to the refilling-recovery location and only when the local region is not in persistent emptying and has regained some refilling capacity.

An effective impedance ratio and pressure-enhancement coefficient are constructed from a stenosis/propagation-limitation severity state $\sigma$:

$$R_Z = 1 + k_Z \frac{\sigma^{n_Z}}{\sigma^{n_Z} + \sigma_Z^{n_Z}}, \qquad \Gamma = \frac{R_Z - 1}{R_Z + 1}. \tag{S105}$$

Here, $\sigma$ is a stenosis or propagation-limitation severity state ranging from 0 to 1; $R_Z$ is the dimensionless effective impedance ratio; $k_Z$ is the impedance-mismatch enhancement strength; $\sigma_Z$ is the severity scale producing half-maximal enhancement; $n_Z$ is a nonlinear exponent; and $\Gamma$ is the pressure-enhancement coefficient inferred from impedance mismatch, ranging from 0 to 1.

The positive pulsatile component relative to a local slow baseline is enhanced according to within-beat phase:

$$P_{pulse,mod} = P_{pulse,-} + [1 + \Gamma G_{beat}(\phi)] P_{pulse,+}. \tag{S106}$$

Here, $P_{pulse,mod}$ is the modified local pulsatile pressure component; $P_{pulse,+}$ is the positive pulsatile component relative to the local slow baseline; $P_{pulse,-}$ is the negative pulsatile component; $G_{beat}(\phi)$ is a within-beat or beat-level modulation function; and $\Gamma$ is the pressure-enhancement coefficient. The equation enhances only the positive pulsatile component to represent pressure accumulation and positive pressure release in the transmission-limited distal segment. Distal pressure elevation modifies the intraluminal pressure field that enters area and volume calculations; it is not post-processing of the final oscillogram.

**Proximal mirror-like pressure response.** The proximal mirror-like pressure response represents reflection-like components formed in the proximal pressure field when distal pressure accumulation occurs under propagation limitation and impedance mismatch. The module maps the distal enhanced pressure increment to the proximal pressure field according to an axial mirror relationship:

$$\Delta P_{prox,mirror}(z,t) = g_{mirror} \Delta P_{dist,boost}(z^*, t - \Delta\tau^*). \tag{S107}$$

Here, $\Delta P_{prox,mirror}(z,t)$ is the mirror-like pressure increment mapped to the proximal pressure field, in mmHg; $g_{mirror}$ is the mirror-reflection gain; $\Delta P_{dist,boost}(z^*, t - \Delta\tau^*)$ is the distal pressure-enhancement increment at the paired distal position after additional propagation delay; $z^*$ is the distal position paired to proximal position $z$ by axial mirroring; and $\Delta\tau^*$ is the additional delay of this mirror component. When the mirrored length exceeds the distal length that can be strictly paired, the remaining proximal reflection-like component continues to propagate with additional proximal delay. This term also enters the actual pressure field and is not used only for visualization.

**Local pressure waves associated with DBP and SBP.** As cuff pressure approaches $DBP$ and $SBP$, the model predicts local pressure packets, deepening notches, and double-peak-like transitions under propagation limitation and progressive proximal-distal decoupling. These fine morphologies are not assumed to arise solely from the basic opening/closure area law. Instead, they are represented as mechanism-inspired local pressure-wave terms under severe compression to describe additional pressure-field remodeling.

The local $DBP$-related pressure wave is written as

$$\Delta P_{DBP}(z,t) = A_{DBP} M_{DBP}(t) K_{DBP}[\phi(t), z]. \tag{S108}$$

Here, $\Delta P_{DBP}(z,t)$ is the $DBP$-related local pressure-wave increment, in mmHg; $A_{DBP}$ is its amplitude parameter, in mmHg; $M_{DBP}(t)$ is the maturity state of the $DBP$-related pressure wave, ranging from 0 to 1; $K_{DBP}[\phi(t), z]$ is a dimensionless kernel localized in both within-beat phase and axial position; and $\phi(t)$ is within-beat phase.

The $SBP$-related term consists of a post-systolic negative component and a subsequent positive component:

$$\Delta P_{SBP}(z,t) = -A_N M_{SBP}(t) K_N[\phi(t), z] + A_P M_{SBP}(t) K_P[\phi(t), z]. \tag{S109}$$

Here, $\Delta P_{SBP}(z,t)$ is the $SBP$-related local pressure-wave increment; $A_N$ and $A_P$ are the amplitude parameters of the post-systolic negative component and subsequent positive component, respectively, in mmHg; $M_{SBP}(t)$ is the maturity state of the $SBP$-related pressure wave; and $K_N[\phi(t), z]$ and $K_P[\phi(t), z]$ are the phase- and axis-localized kernels for the negative notch-like and positive second-peak-like components. These local pressure waves are written into the intraluminal pressure field and therefore continue to enter transmural pressure, vascular area, total volume, and recovered cuff-pressure oscillations. Their amplitudes and timing are constrained by physiologically plausible waveform behavior and are not unconstrained empirical degrees of freedom for blood-pressure estimation.

##### S2.2.10 Parameter summary and numerical implementation

To avoid conflating boundary conditions, mechanistic parameters, pressure-field remodeling parameters, and numerical implementation settings, we summarize parameters according to functional class. The main parameter table reports representative mechanistic parameters and their interpretation; a complete numerical-implementation table is provided separately to support reproducibility. Representative mechanistic parameters used to instantiate the unified cuff–brachial artery model are summarized in Supplementary Table 2, whereas numerical implementation settings required for reproducing the representative simulations are listed separately in Supplementary Table 3.

##### S2.2.11 Numerical implementation, model instantiation, and reproducibility

To balance mechanistic clarity and reproducibility, we use a two-level parameter-reporting strategy. The main table lists the representative parameters required for geometry, external pressure, boundary input, pressure propagation, pressure-area behavior, collapse, distal emptying, pressure-field remodeling, and cuff readout. The complete implementation table lists numerical settings that affect final pressure, area, volume, and oscillogram outputs.

The complete numerical-implementation parameters allow representative simulations to be reproduced, including the positions and widths of phase-localized kernels, smoothing windows, rise and decay constants of finite state variables, and numerical-stability constraints. These parameters serve numerical implementation and waveform presentation within the unified mechanism; they do not constitute additional physiological mechanisms outside the main model.

All representative simulations are generated from the same unified model. Different scenarios change only the boundary input, cuff-pressure protocol, representative physiological state, or pressure-field remodeling parameters to show predictions under different physiological conditions. They should not be interpreted as independent models. Constant input and no-remodeling conditions are mechanistic controls used to isolate the contributions of nonuniform external pressure, transmural pressure, and vascular collapse to the output waveform.

##### S2.2.12 Reconstruction of cuff oscillations from multisite IVUS-measured CSA

To test whether cuff oscillations can be generated directly from subcuff vascular motion rather than from an empirical oscillogram criterion, we used time-varying brachial-artery CSA sequences measured at multiple axial sites under the cuff as input. A continuous axial CSA distribution was reconstructed and inserted into the same volume-to-cuff-pressure readout relation used in the unified model [12,15,68].

Let measured CSA sequences $A_i(t)$ be obtained at axial positions $z_i$ $(i = 1, \dots, M)$. The continuous CSA distribution between adjacent measurement sites is approximated by linear interpolation:

$$A(z,t) = A_i(t) + \frac{A_{i+1}(t) - A_i(t)}{z_{i+1} - z_i}(z - z_i), \qquad z_i \le z \le z_{i+1}. \tag{S110}$$

Here, $A_i(t)$ and $A_{i+1}(t)$ are the measured CSAs at axial positions $z_i$ and $z_{i+1}$, in mm$^2$; $A(z,t)$ is the linearly interpolated continuous CSA between the two sites; $M$ is the number of measured axial sites; and $z$, $z_i$, and $z_{i+1}$ are in mm.

The continuous area distribution is then integrated along the cuff axis to obtain total subcuff arterial volume:

$$V_{CSA}(t) = \frac{1}{1000}\int_{-W_{cuff}/2}^{W_{cuff}/2} A\,(z,t)\,dz. \tag{S111}$$

Here, $V_{CSA}(t)$ is the subcuff arterial volume reconstructed from measured CSA; the integration range from $-W_{cuff}/2$ to $W_{cuff}/2$ denotes the cuff-covered segment; if $A$ is in mm$^2$ and $z$ is in mm, the integral is in mm$^3$ and is converted to mL by division by 1000. Substituting $V_{CSA}(t)$ into the same cuff volume-pressure coupling relation in S2.2.8 yields recovered cuff pressure and the extracted oscillogram. This analysis tests whether IVUS-measured local CSA changes can reproduce the main timing and morphology of measured cuff oscillations through volume-to-cuff-pressure readout.

##### S2.2.13 Local volume control experiments

Local control experiments were used to examine how specific axial volume components under the cuff contribute to $DBP$- and $SBP$-related oscillogram morphology. These analyses are causal perturbation tests applied to model volume components, not additional blood-pressure estimation criteria. In each control, the original cuff-pressure trajectory and unperturbed volume components are retained; the modified total volume is passed again through the same cuff volume-pressure readout and oscillation-extraction workflow.

For any axial region $\Omega$, the local volume component is defined as

$$V_{\Omega}(t) = \frac{1}{1000}\int_{\Omega} A\,(z,t)\,dz. \tag{S112}$$

Here, $\Omega$ is an arbitrarily specified axial region; $V_{\Omega}(t)$ is the local volume component in that region; and $A(z,t)$ is the local area distribution in the corresponding region.

**DBP-related local volume replacement.** In the $DBP$-related control, the replacement region was adaptively determined from the membrane-theory external-pressure distribution and a fast-collapse threshold under the reference cuff pressure in the target window. This defines the $DBP$-dominant axial region $\Omega_{DBP}$ and reflects the requirement that the $DBP$-dominant region should follow the model-predicted PPZ rather than an arbitrary fixed fraction of cuff width.

Let $P_{c,ref}$ be the mean cuff pressure in the $DBP$-control target window. To avoid sign ambiguity for negative transmural pressure, the negative-transmural-pressure magnitude required for substantial rapid collapse is defined as a positive value $P_{col}^{*}$. The external-pressure distribution threshold is

$$\alpha_{th} = \frac{DBP + P_{col}^{*}}{P_{c,ref}}. \tag{S113}$$

Here, $\alpha_{th}$ is the external-pressure distribution threshold for identifying the $DBP$-dominant axial region; $P_{c,ref}$ is the mean cuff pressure in the $DBP$-control target window; $P_{col}^{*}$ is the positive negative-transmural-pressure magnitude needed for rapid collapse to become prominent; and $DBP$ is reference diastolic pressure. If $\alpha_{th}$ falls outside 0–1, it is bounded according to the definition of the normalized external-pressure distribution.

The $DBP$ control region is therefore

$$\Omega_{DBP} = \{z\colon \alpha(z) \geq \alpha_{th}\}. \tag{S114}$$

Here, $\Omega_{DBP}$ is the $DBP$-dominant axial region satisfying the external-pressure threshold, typically corresponding to the cuff PPZ. The corresponding local volume is

$$V_{DBP}(t) = \frac{1}{1000}\int_{\Omega_{DBP}} A\,(z,t)\,dz. \tag{S115}$$

Here, $V_{DBP}(t)$ is the local volume component of the $DBP$-dominant region. Within a user-defined target time window, $V_{DBP}(t)$ is replaced by a donor segment from another time window. If donor and target windows differ in length, the donor segment is first linearly resampled to the target length; a 50-ms crossfade is then applied at both endpoints to preserve continuity. The modified total volume is

$$V_{ctrl}(t) = V_{total}(t) - V_{DBP}(t) + \tilde{V}_{DBP}(t). \tag{S116}$$

Here, $V_{ctrl}(t)$ is the controlled total volume after replacement; $V_{total}(t)$ is the original model total volume; $V_{DBP}(t)$ is the original $DBP$-dominant regional volume; and $\tilde{V}_{DBP}(t)$ is the replacement $DBP$ local-volume segment obtained after donor-window resampling and crossfade. Finally, $V_{ctrl}(t)$ is passed again through the same recovered cuff-pressure calculation, band-pass filtering, and lower-envelope detrending workflow.

**SBP-related distal volume-locking control.** In the $SBP$-related control, late distal volume re-expansion is suppressed to test its contribution to $SBP$-related morphology. If multiple emptying-entry events are present, the locking time $t_{hold}$ is chosen as the penultimate emptying-entry event; if only one usable event is present, that event is used.

Let $V_{dist}(t)$ be the distal volume component generated by the model. From $t_{hold}$ onward, the controlled distal volume is defined as the cumulative minimum:

$$\tilde{V}_{dist}(t) = \min_{\tau \in [t_{hold}, t]} V_{dist}(\tau), \qquad t \geq t_{hold}. \tag{S117}$$

Here, $\tilde{V}_{dist}(t)$ is the controlled distal volume after the locking time $t_{hold}$; $V_{dist}(t)$ is the original distal volume; and $t_{hold}$ is determined by the distal emptying-entry event. This definition prevents distal volume from re-expanding after $t_{hold}$; it can only remain unchanged or decrease. For $t < t_{hold}$, $\tilde{V}_{dist}(t) = V_{dist}(t)$.

The corresponding controlled total volume is

$$V_{ctrl}(t) = V_{prox}(t) + \tilde{V}_{dist}(t). \tag{S118}$$

Here, $V_{prox}(t)$ is the proximal volume component that is not locked; $\tilde{V}_{dist}(t)$ is the locked distal volume component; and $V_{ctrl}(t)$ is the recomputed total volume for the $SBP$ distal-locking control. This operation preserves proximal volume contribution while suppressing distal re-expansion and late distal pulsatile recovery. The recovered cuff pressure and oscillogram are then recalculated from $V_{ctrl}(t)$. If $SBP$-related morphology changes, this indicates that the late oscillogram morphology depends on distal emptying, delayed distal recovery, and proximal-distal volume mismatch rather than only on the global oscillogram envelope.

$DBP$ and $SBP$ controls can be performed individually or in combination. All control segments generate only modified oscillograms and do not change the original unified-model simulation results.

##### S2.2.14 Model-derived DBP- and SBP-related oscillogram morphologies

In the unified model, the emergence, enhancement, and attenuation of cuff oscillations correspond to continuous vascular-state transitions rather than empirical envelope features [14,19]. At low cuff pressure, most vascular cross-sections remain open and local volume pulsation is small; therefore, oscillation amplitude is weak. As cuff pressure enters the vicinity of $DBP$, the cuff PPZ first enters a more prominent cyclic compression–reopening state. Diastolic and systolic CSA diverge, total volume pulsation increases, and the oscillogram enters a rapid-rising phase. Thus, $DBP$-related information can be interpreted as the vascular-mechanical transition at which cyclic compression–reopening becomes established in the central dominant region, rather than as an isolated empirical amplitude threshold.

As cuff pressure further approaches $SBP$, the closure region expands, closure duration increases, proximal-distal connectivity decreases, and distal squeeze-emptying strengthens. Distal pressure and distal volume pulsation become increasingly delayed and attenuated relative to proximal pulsation. Thus, $SBP$-related information is expressed mainly through oscillogram morphological changes associated with restricted pulsatile transmission, distal emptying, delayed distal recovery, and proximal-distal volume mismatch, rather than through a fixed-amplitude ratio.

##### S2.2.15 Summary of the supplementary model framework

Together, the unified model and local volume control analyses link axially nonuniform cuff external pressure to the spatiotemporal evolution of subcuff arterial CSA, local volume components, and recovered cuff oscillations. $DBP$-related morphology is associated with the establishment of cyclic compression in the PPZ. $SBP$-related morphology is associated with restricted distal pulsatile transmission, delayed distal refilling, and proximal-distal volume mismatch under high cuff pressure. These analyses support the interpretation that cuff oscillogram morphology arises from spatially localized vascular dynamics and their coupling to cuff volume-pressure readout, rather than from the global oscillogram envelope itself [14,19].

#### *Section S2.3 Pressure-waveform changes at different recording sites*

##### S2.3.1 Axial intra-arterial waveform changes at 6 sub-cuff sites

During cuff inflation, representative simultaneous invasive recordings from 6 axial sub-cuff sites were used to illustrate the site-dependent remodeling of intra-arterial pressure waveforms beneath the cuff. These representative recordings were selected from experiments in which all 6 predefined sub-cuff sites were successfully sampled, thereby providing broad axial coverage from the proximal edge-reference position through the five internal sub-cuff positions.

At the proximal cuff-edge site (Site 0), the intra-arterial pressure waveform remained largely similar to its baseline configuration. However, once the local cuff pressure exceeded the diastolic level, the notch pattern on the descending limb began to change. Specifically, while the original dicrotic notch remained identifiable, an additional reflection-like notch gradually appeared and became increasingly prominent near the waveform trough. This change persisted until cuff deflation.

More pronounced waveform changes were observed at the 2 adjacent sites located distally along the axial direction from the proximal cuff edge (Site ① and ②). When the local cuff pressure at these sites exceeded diastolic pressure, the diastolic portion of the waveform progressively deformed, and the original trough disappeared, being replaced by a pressure contour that rose with increasing cuff pressure. When the intrinsic arterial pressure at a given phase transiently exceeded the local cuff pressure, the waveform briefly returned toward a near-baseline morphology. In contrast, when the intrinsic arterial pressure fell below the local cuff pressure, the waveform changed progressively with cuff pressure. Once the local cuff pressure exceeded systolic pressure, the pulsatile waveform disappeared and was replaced by a nonpulsatile pressure contour that continued to rise with cuff pressure.

The measurement sites located within the central PPZ of the cuff and at its distal side (Site ③ to ⑤) showed waveform changes that were broadly similar in pattern but more synchronized in timing. At these sites, morphological alteration typically occurred within the same cardiac cycle after cuff pressure exceeded the diastolic level. As cuff pressure approached systolic pressure, weak residual pressure oscillations were observed. These residual oscillations persisted for approximately 5 cardiac cycles and then disappeared within the same cardiac cycle after cuff pressure exceeded systolic pressure. Their disappearance consistently preceded that observed at the proximal site. With further cuff pressurization, the residual pressure contour declined gradually.

During cuff deflation, these changes showed a broadly mirror-image reversal of the inflation process. At the proximal cuff-edge site (Site 0), the pressure waveform remained closest to baseline throughout, with only notch changes present while cuff pressure remained above the diastolic level; these notch changes disappeared as cuff pressure decreased further, and the waveform returned fully to baseline morphology. At the other 2 sites located within the proximal half of the cuff (Site ① and ②), the pressure contour decreased progressively with decreasing cuff pressure, and pressure oscillations reappeared earlier than at the other sites. In contrast, the sites within and distal to the central PPZ (Site ③ to ⑤) exhibited delayed recovery and showed nearly simultaneous reappearance of residual pressure oscillations.

Supplementary Figs. 4 and 5 illustrate this process using inflation and deflation recordings obtained from 2 patients.

###### S2.3.2 Waveform changes from the central under-cuff site to the ascending aorta

Supplementary Fig. 6 shows the pressure-waveform changes recorded at sequential sites in the ascending aorta, the origin of the brachiocephalic trunk, the right subclavian artery, the right axillary artery, the proximal sub-cuff brachial site (Site 0), and the central under-cuff site (Site ③). When cuff pressure exceeded the diastolic level, the effect of cuff compression on the proximal pressure waveform propagated retrogradely from the brachial artery toward more proximal segments, reaching the right subclavian artery, where the waveform exhibited an overall upward shift. No significant change was observed at the origin of the brachiocephalic trunk.

#### *Section S2.4 Additional oscillometric waveform features and arterial pressure remodeling*

###### BP changes and oscillogram morphology during cuff BP measurement

Unless otherwise specified, analyses of intra-arterial pressure changes and corresponding oscillogram features during cuff BP measurement were based on 124 synchronized measurements from 37 participants. Quantitative stage-specific CSA analyses were based on 46 measurements from 25 of the 28 participants who underwent IVUS and were used to characterize the patterns of arterial motion throughout cuff BP measurement.

**Phase I: Cuff pressure less than or equal to DBP (Cuff pressure ≤ DBP).** During Phase I (Supplementary Fig. 10), cuff pressure did not exceed DBP. At the proximal site, intra-arterial SBP showed only a slight increase from baseline, whereas DBP remained essentially unchanged. A similar pattern was observed in the mid-distal artery, where SBP increased only mildly and DBP showed little change. Overall, before cuff pressure exceeded DBP, intra-arterial pressure remained largely stable at both proximal and mid-distal sites, with only minor systolic-pressure changes and no appreciable alteration in diastolic pressure. IVUS showed that the maximal systolic CSA of the brachial artery at each measurement site remained close to baseline (20.72 ± 5.78 vs. 21.80 ± 5.82 mm$^2$), whereas the minimal diastolic CSA showed a slight decrease relative to the preceding state (17.26 ± 4.79 vs. 21.1 ± 5.56 mm$^2$; decline rate: 0.54 ± 0.42 mm$^2$/s), with only limited CSA variation during the systolic-diastolic cycle (n = 25 participants; 46 measurements; of the 28 participants who underwent IVUS examination, 25 participants with 46 analyzable measurements were included in the quantitative stage-specific CSA analysis).

**Phase II: From DBP to disappearance of the mid-distal pressure pulse (DBP < Cuff pressure ≤ SBP).** Phase II began when cuff pressure exceeded the DBP level (Supplementary Fig. 10). IVUS showed that the brachial artery within the PPZ became markedly compressed from the onset of diastole, with the minimal diastolic area decreasing rapidly (0.54 ± 0.42 vs. 1.65 ± 1.07 $mm^2/s$), whereas the maximal systolic area remained relatively stable in the early stage (decline rate: 0.24 ± 0.22 $mm^2/s$) (Supplementary Fig. 10, blue dashed arrow in Phase II). This marked the initial pressure-induced collapse of the artery. At the same time, the negative component of the oscillometric waveform synchronized with diastole increased abruptly (Supplementary Fig. 10, orange dashed arrow in Phase II). Within the same cardiac cycle, the diastolic trough of the oscillometric waveform showed a sudden steep downward deflection, corresponding to the first pressure-induced collapse of the sub-cuff brachial artery and providing a reliable oscillometric landmark for DBP localization.

As the brachial artery within the PPZ became compressed and progressively occluded during diastole, diastolic intra-arterial pressure at this site and in the distal brachial artery gradually increased (Supplementary Fig. 10, green dashed arrow in Phase II). Synchronously, IVUS showed that the rate of change in brachial artery diastolic CSA slowed markedly (decline rate: 1.65 ± 1.07 vs. 0.33 ± 0.36 $mm^2/s$), with the minimum diastolic CSA eventually decreasing to 0 (Supplementary Fig. 10, red dashed arrow in Phase II), and the notch phase of the area-time curve shortened and began to disappear (Supplementary Fig. 10, green arrow in Phase II).

A diastolic reflection-like notch appeared synchronously in the proximal pressure waveform and the corresponding oscillometric waveform (Supplementary Fig. 10, red arrow in Phase II). Synchronous brachial artery angiography indicated that this feature did not represent a classical pressure-wave reflection. Rather, as distal resistance progressively increased, antegrade diastolic flow decelerated and transiently accumulated proximally, producing a secondary forward-propagating expansion that appeared as an upward deflection in the diastolic waveform tail (Supplementary Video 9). By mid-Phase II, the original dicrotic notch in the pressure waveform within the PPZ and in the oscillometric waveform had begun to weaken and disappear in some participants, while the reflection-like notch emerged; during this transition, the timing of the two features could partially overlap (Supplementary Fig. 10, blue arrow in Phase II). This morphological transition occurred as cuff pressure approached the MAP level. Wavelet-based localization of the DBP-related morphology transition is described in section S1.5.4 and illustrated in Fig. 4b-e of the main text.

As cuff pressure increased further, the oscillometric wave reached its maximum amplitude. Proximal SBP increased from 145.34 ± 22.43 mmHg to 157.68 ± 26.18 mmHg, proximal DBP increased from 76.15 ± 12.05 mmHg to 79.80 ± 12.44 mmHg, and proximal pulse pressure increased from 69.19 ± 17.97 mmHg to 77.88 ± 21.09 mmHg. Mid-distal SBP increased from 150.57 ± 21.21 mmHg to 164.72 ± 25.55 mmHg, mid-distal DBP increased from 78.38 ± 11.87 mmHg to 114.83 ± 28.80 mmHg, whereas mid-distal pulse pressure decreased from 72.19 ± 17.17 mmHg to 49.88 ± 21.28 mmHg. IVUS showed that brachial artery CSA began to decline markedly during systole (decline rate: 0.85 ± 0.90 $mm^2/s$; Supplementary Fig. 10, blue dashed arrow in Phase II). At this point, cuff pressure approached and progressively exceeded the dicrotic-notch level, and the dicrotic notch in the pressure waveform within the PPZ and in the oscillometric wave gradually disappeared (Supplementary Fig. 10, blue arrow in Phase II, Supplementary Video 10).

The oscillometric wave and its corresponding cuff pressure at the time point defined by the intersection between disappearance of the dicrotic notch and appearance of the diastolic reflection-like component showed the highest consistency with MAP calculated by the invasive pressure integral method (105.39 ± 14.29 vs. 105.76 ± 14.69 mmHg, P = 0.281, n = 175 participants; 630 measurements) (Supplementary Fig. 11). Furthermore, the corresponding cuff pressure of the oscillometric wave at its maximum amplitude is slightly higher than the invasively measured MAP (108.13± 18.25 vs. 105.76 ± 14.69 mmHg, P < 0.001, n = 175 participants; 630 measurements). Subsequently, a double-peaked morphology began to appear in the proximal and mid-segment pressure waveforms (Supplementary Fig. 10, red box in Phase II), originating from secondary vascular expansion during systole (Supplementary Video 10).

During phase II, proximal SBP and DBP increased modestly, accompanied by a modest increase in proximal pulse pressure. In the mid-distal segment, SBP increased slightly in parallel with the proximal segment, whereas DBP continued to rise with increasing cuff pressure, leading to progressive narrowing of pulse pressure.

As cuff pressure continued to increase and approached SBP, the pressure waveform in the mid-distal segment progressively lost the morphology of the original BP waveform, and residual low-amplitude pulsations, referred to as pressure "small waves," began to appear (Supplementary Fig. 10, blue box in Phase II). Relative to the proximal pressure waveform, these small waves were markedly delayed and attenuated. IVUS showed that the open CSA and pulsation amplitude of the mid-distal brachial artery were substantially reduced (decline rate: 2.83 ± 2.08 $mm^2/s$) (Supplementary Fig. 10, red dashed arrow in Phase II), indicating that the vessel could no longer maintain an effective flow-transmission channel. As blood traversed this severely compressed, intermittently occluded, and narrowed arterial segment, transmission to the distal artery became progressively delayed, increasing the time required for the pressure pulsation to reach the distal segment. This stage typically lasted approximately five cardiac cycles (4.81 ± 2.58). Meanwhile, proximal SBP began to decline and gradually approached the pre-measurement level (mean proximal SBP: 144.97 ± 23.42 vs. 145.34 ± 22.43 mmHg). During this sustained transition, a series of visually recognizable oscillogram morphology changes emerged, as detailed below.

**Visual oscillogram features during small-wave appearance.** When cuff pressure was slightly below SBP, the delayed distal pulsatile component described above progressively separated from the main oscillogram peak, shifted onto its descending limb, and decreased in amplitude. When the artery remained patent, the relatively small proximal–distal delay allowed the corresponding volume pulsations to overlap temporally and jointly form the main oscillogram peak. With marked arterial compression and narrowing, the increasing delay progressively separated the proximal and distal volume contributions in time, causing the main peak to resolve into two temporally distinct components. As distal blood flow decreased, the delayed distal component contributed progressively less to the total oscillogram. This temporal separation and attenuation produced a sequence of visually recognizable oscillogram morphology changes, mainly including peak-side shift in 64 of 124 measurements, amplitude transition in 60 of 124 measurements, and transient double-peak morphology in 26 of the 64 measurements with peak-side shift. Complete oscillograms are shown in Supplementary Figs. 12-15.

**Phase III: Complete occlusion of the mid-distal arterial segment (Cuff pressure > SBP).** When cuff pressure increased just above the SBP level, the mid-distal brachial artery became completely occluded during both systole and diastole, thereby entering Phase III (Supplementary Fig. 10). IVUS and synchronous pressure measurements showed complete disappearance of mid-distal vascular pulsation and the distal pressure waveform, with blood no longer able to pass through the occluded segment. Notching or a double-peaked morphology appeared in the proximal pressure waveform, accompanied by synchronous changes in the oscillometric waveform. These features arose from a reflected component generated when systolic flow was obstructed by the occluded sub-cuff brachial artery (Supplementary Video 9).

After cuff pressure exceeded SBP, proximal arterial volume pulsation became the principal source of the oscillometric waveform. Because cuff-induced external pressure was axially nonuniform, the pressure applied to the artery near the proximal cuff edge remained below systolic intra-arterial pressure, allowing systolic blood flow to continue entering the proximal sub-cuff artery. As blood propagated distally toward the PPZ, external pressure progressively increased, preventing flow from traversing the PPZ and allowing it to reach only to or near this region. Nevertheless, this proximal systolic inflow produced sufficient arterial volume pulsation to generate cuff oscillations. Thus, after complete PPZ occlusion, the remaining oscillometric waveform was generated by proximal arterial volume pulsation rather than distal flow transmission.

**Visual oscillogram features during small-wave disappearance.** At SBP, disappearance of the distal pulsatile component indicated loss of the small-wave-related contribution to the oscillogram. On the oscillogram, this was reflected by disappearance of the feature component on the descending limb of the main peak. Morphologically, the oscillogram mainly showed abrupt whole-peak or local narrowing of the main peak in 84 of 124 measurements and marked reduction in overall amplitude in 89 of 124 measurements. These patterns were not mutually exclusive. The different visual patterns may reflect interindividual differences in vascular properties, body habitus, and arterial pressure waveform morphology. In some cases, the transition was also accompanied by disappearance of a subtle notch on the descending limb. After this transition, the oscillatory signal was dominated mainly by the remaining proximal pulsatile component. Representative oscillograms are shown in Supplementary Figs. 16-19.

**Distinguishing distal small-wave and reflection-related components.** Beyond the patterns described in the main text, we also observed that, in some measurements, a notch-related component remained detectable to the right of the main coefficient peak after termination of the distal small-wave tracking chain. Supplementary Fig. 21 shows a representative example. The peak-to-notch delay of the tracked distal small-wave component followed a continuous beat-to-beat trajectory and tended to increase progressively with increasing cuff pressure before MTO termination. In contrast, the subsequently detected reflection-related component generally remained at a relatively stable delay from the main coefficient peak across beats and typically exhibited lower local wavelet coefficient energy than the distal small-wave component. This reflection-related component also remained detectable after MTO termination, forming a temporal trajectory distinct from that of the distal small-wave component. The vertical dashed line indicates MTO termination. Thus, the distal small-wave component was characterized by a pressure-dependent increase in temporal delay and eventual disappearance together with loss of distal arterial pulsation, whereas a post-termination component that persisted at a relatively stable delay and lower wavelet energy was classified as a reflection-related proximal response rather than evidence of continued distal transmission. Persistent post-termination components were therefore not considered a continuation of the distal small-wave tracking chain.

Together with the Mexican-hat wavelet results in Fig. 4g,h of the main text, these visual oscillogram features converge on the same key event: loss of the distal arterial contribution to the oscillometric waveform.

#### *Section S2.5 Hemodynamic basis and quantitative analysis of KS boundaries*

At the IVUS site closest to the acoustic sensor, the directly measured descriptor of vascular opening was the rate of change in luminal CSA. It was defined as

$$\dot{A}_j(t) = \frac{dA_j(t)}{dt} \tag{S119}$$

Here, $A_j(t)$ is the luminal CSA at IVUS site $j$ and time $t$, and $\dot{A}_j(t)$ is the CSA expansion rate in mm²/s. Positive values indicate luminal expansion, whereas negative values indicate contraction.

$$\dot{A}_j(t_n) \approx \frac{A_j(t_{n+1}) - A_j(t_{n-1})}{2\Delta t} \tag{S120}$$

For framewise IVUS measurements, the derivative was evaluated by the centered finite difference in Eq. S120, where $\Delta t$ is the time interval between consecutive analyzed frames. Forward or backward differences were used only at the first or last available frame of a measurement segment.

$$\dot{A}_{j,k}^{\max} = \max_{t} \dot{A}_j(t) \tag{S121}$$

Here, $\mathcal{B}_k$ is the time interval of beat $k$ and $\dot{A}_{j,k}^{max}$ is the maximum positive CSA expansion rate during that beat. This beat-wise value was used when comparing vascular opening with the presence or absence of a KS. No universal expansion-rate threshold was imposed, because the observed sound-generation threshold varied substantially among participants and measurement states.

$$v_{r,j}(t) = \frac{1}{2\sqrt{\pi A_j(t)}} \frac{dA_j(t)}{dt} \tag{S122}$$

If a length-per-time descriptor was required, an equivalent circular lumen with $A_j(t) = \pi r_j^2(t)$ was assumed, which gives the radial expansion velocity $v_{r,j}(t)$ in Eq. S122. This conversion is an equivalent geometric representation and does not imply that the compressed lumen remained circular.

In the arterial-pressure observation subset (37 participants; 124 synchronized measurements), we statistically analyzed the relationship between K5 and the cardiac beat corresponding to invasive DBP. We also evaluated the relationship between K1 and the SBP-related characteristic hemodynamic event, defined as the final distal pressure small wave during cuff inflation and the first distal pressure small wave during cuff deflation. K5 did not stably correspond to invasive DBP: it occurred before DBP in 47 measurements, after DBP in 40 measurements, and within the same cardiac beat as DBP in the remaining 37 measurements, indicating variable alignment between the K5 acoustic boundary and invasive DBP. Compared with premeasurement invasive DBP in the same measurements, K5-based DBP showed a small nonsignificant bias (Bias = −0.57 ± 6.00 mmHg, P = 0.652). For SBP, the K1-related acoustic boundary did not consistently coincide with the corresponding distal pressure-small-wave boundary. Distal pressure small waves remained detectable without corresponding KS in 72 of 124 measurements when cuff pressure was slightly below SBP, whereas KS coexisted with distal pressure small waves in 52 measurements. Corresponding arterial pulsation was assessed in the recordings with synchronized IVUS. Compared with premeasurement invasive SBP in the same measurements, K1-based SBP showed a systematic negative bias (Bias = −5.28 ± 8.95 mmHg, P < 0.001). In recordings with synchronized IVUS and KS data (n = 25; N = 46), the estimated KS-generation thresholds differed between K5 and K1 and showed marked interindividual variability (K5, 117.78 ± 75.03 mm²/s; K1, 107.56 ± 108.55 mm²/s), limiting the feasibility of using a fixed KS threshold to infer DBP or SBP. These findings indicate that K5 and K1 are influenced by individualized KS-generation thresholds and therefore do not accurately localize DBP and SBP. These KS results were obtained in the 37-participant arterial-pressure observation subset; the overall comparison of BP measurement methods used 630 paired measurements from 175 participants, as reported in Section S2.7.

#### *Section S2.6 Mechanistic amplitude-ratio framework based on the unified cuff-artery model*

##### S2.6.1 Purpose and scope

This section does not re-derive the unified cuff-brachial artery model described above. Instead, it analyzes the cuff oscillogram generated by that model and defines the maximum-amplitude beat, the diastolic- and systolic-pressure-related beats, and their amplitude ratios relative to the maximum oscillation amplitude. The purpose is to explain the mechanistic basis of these amplitude ratios and why fixed amplitude ratios used in the traditional maximum-amplitude method lack mechanistic universality.

Traditional maximum-amplitude algorithms first identify the maximum of the cuff-oscillation envelope and usually assume that this point corresponds to mean arterial pressure. Fixed or empirically modified amplitude ratios before and after the maximum are then used to estimate DBP and SBP, effectively treating these ratios as empirical population constants. In the unified model, by contrast, the cuff oscillogram is generated by axial integration of the luminal-area field over the cuff-covered arterial segment, followed by cuff air-chamber volume-to-pressure readout. Therefore, the maximum-amplitude beat and the DBP- and SBP-related beats are outputs of the coupled cuff-artery system rather than independently imposed empirical envelope points. Their corresponding amplitude ratios can be expressed analytically within the model, but the resulting expressions depend on multiple vascular, cuff, tissue, and hemodynamic parameters that are not directly available during routine cuff measurement. Consequently, these ratios cannot in practice be reduced to universally calculable fixed coefficients, which explains why conventional maximum-amplitude algorithms rely on empirically determined amplitude ratios.

Only the mathematical definitions and explanatory relations required for the amplitude-ratio framework are introduced here. The vascular mechanics, cuff external pressure, transmural pressure, area response, functional closure, distal emptying/refilling, and pressure-field remodeling retain the meanings established in the unified model. Symbols used only for earlier model-specific event definitions are not carried forward unless redefined in the present section.

##### S2.6.2 Beatwise oscillation amplitude and general amplitude ratio

Let $\mathrm{P_{cuff,rec}(t)}$ denote the cuff pressure recovered from the unified model. The model cuff oscillogram is obtained by applying the same oscillation-extraction procedure used for the experimental cuff signal:

$$\mathrm{P_{osc}(t)} = \mathcal{F}\{\mathrm{P_{cuff,rec}(t)}\}. \tag{S123}$$

Here, $P_{osc}(t)$ is the extracted model cuff oscillogram, in mmHg; $P_{cuff,rec}(t)$ is the recovered cuff pressure derived from model-predicted volume changes, in mmHg; and $\mathcal{F}\{\cdot\}$ is the oscillation-extraction operator, including filtering, detrending, and the output-level processing used to match the experimental signal definition. This step defines the observable oscillatory component and introduces no new vascular mechanism.

Let $\mathcal{B}_k = [t_{k,0}, t_{k,1}]$ denote the time window of beat $k$. The beatwise oscillation amplitude is defined as

$$A_k = \max_{t\in\mathcal{B}_k} P_{osc}(t) - \min_{t\in\mathcal{B}_k} P_{osc}(t). \tag{S124}$$

Here, $A_k$ is the peak-to-trough oscillation amplitude of beat $k$, in mmHg; $\mathcal{B}_k$ is the corresponding beat window; and $t_{k,0}$ and $t_{k,1}$ are the onset and end times of that beat. The peak-to-trough definition is used to remain comparable with the envelope amplitude in conventional oscillometry. If a specific morphology analysis is required, the amplitude operator may be replaced by a predefined main-peak amplitude or positive-peak amplitude, but the same definition must then be used throughout the analysis.

The mean cuff pressure associated with beat $k$ is

$$\bar{P}_{cuff,k} = \frac{1}{t_{k,1}-t_{k,0}} \int_{t_{k,0}}^{t_{k,1}} P_{cuff,cmd}(t)\, dt. \tag{S125}$$

Here, $\bar{P}_{cuff,k}$ is the mean commanded cuff pressure during beat $k$, in mmHg; $P_{cuff,cmd}(t)$ is the commanded or protocol-defined cuff pressure, in mmHg; and $t_{k,1} - t_{k,0}$ is the duration of beat k, in seconds. When cuff pressure varies only slightly within one beat, the midpoint cuff pressure may be used as an approximation.

For any cuff pressure $P_c$, the mechanistic amplitude ratio is defined as

$$K(P_c) = \frac{A(P_c)}{A_{max}}. \tag{S126}$$

Here, $K(P_c)$ is the dimensionless mechanistic amplitude ratio at cuff pressure $P_c$; $A(P_c)$ is the beatwise oscillation amplitude corresponding to that cuff pressure, in mmHg; and $A_{max}$ is the maximum beatwise oscillation amplitude over the entire measurement, in mmHg. Equation S126 defines a ratio relative to the maximum amplitude; it does not imply that $P_c$ corresponds to any fixed empirical blood-pressure point.

##### S2.6.3 Maximum-amplitude beat and its relation to mean arterial pressure

The maximum-amplitude beat is defined by

$$k_{max} = \arg\max_{k} A_k. \tag{S127}$$

Here, $k_{max}$ is the index of the beat with the largest oscillation amplitude, and $A_k$ is the beatwise oscillation amplitude.

The maximum beatwise oscillation amplitude is

$$A_{max} = A_{k_{max}}. \tag{S128}$$

Here, $A_{max}$ is the maximum beatwise oscillation amplitude over the full measurement, in mmHg, and $A_{k_{max}}$ is the amplitude of the maximum-amplitude beat.

The cuff pressure associated with the maximum-amplitude beat is

$$P_{max} = \bar{P}_{cuff,k_{max}}. \tag{S129}$$

Here, $P_{max}$ is the mean cuff pressure of the maximum-amplitude beat, in mmHg, and $\bar{P}_{cuff,k_{max}}$ is the mean cuff pressure of beat $k_{max}$.

In the conventional maximum-amplitude method, $P_{max}$ is often assumed to approximate mean arterial pressure. In the present model, however, $P_{max}$ represents the cuff pressure at which the total subcuff volume pulsation, after cuff readout, reaches its maximum. The total volume of the cuff-covered arterial segment is

$$V_{total}(t) = \int_{\Omega_{cuff}} A\,(z,t)\,dz. \tag{S130}$$

Here, $V_{total}(t)$ is the total arterial volume under the cuff, in $mm^3$ or after conversion in mL; $A(z,t)$ is the local luminal area at axial position $z$ and time $t$, in $mm^2$; $\Omega_{cuff}$ is the cuff-covered axial region; and $dz$ is the axial integration element, in mm.

In the discrete model, Eq. S130 becomes

$$V_{total}(t) \approx \sum_{j=1}^{N_z} A_j\,(t)\Delta z. \tag{S131}$$

Here, $A_j(t)$ is the local luminal area at axial site $j$, in $mm^2$; $N_z$ is the number of axial sites; $\Delta z$ is the axial spacing, in mm; and $j$ is the axial-site index.

The beatwise total-volume pulsation is

$$\Delta V_k = \max_{t\in\mathcal{B}_k} V_{total}(t) - \min_{t\in\mathcal{B}_k} V_{total}(t). \tag{S132}$$

Here, $\Delta V_k$ is the total volume pulsation of beat $k$ over the cuff-covered segment, and $\mathcal{B}_k$ is the beat window.

The maximum-amplitude beat can therefore be expressed as

$$k_{max} = \arg\max_{k} \mathcal{R}(\Delta V_k). \tag{S133}$$

Here, $\mathcal{R}(\cdot)$ is the compound readout operator that maps total volume pulsation to cuff-pressure oscillation amplitude, including cuff air-chamber volume-to-pressure coupling, recovered cuff pressure, oscillation extraction, and beatwise amplitude calculation. Equation S133 shows that the maximum-amplitude point is the maximum of the axially integrated volume pulsation after cuff readout, not necessarily the pressure at which a local arterial segment has maximal compliance.

The beatwise total-volume pulsation reflects the coupled effects of open-segment compliance, cyclic collapse and reopening, distal emptying and refilling, and propagation asynchrony. In the numerical model, these mechanisms are not calculated as four unique and mutually exclusive additive signals. Their contributions are inferred from the corresponding state variables, axial-region outputs, and controlled segment-replacement analyses. The following small-oscillation formulation is therefore used only to show how coupled local responses enter the axially integrated cuff signal, rather than to assert an exact additive decomposition of total volume pulsation.

This mechanistic distinction also provides the rationale for the morphology-based MAP criterion used in MTO. Rather than assigning MAP solely to the maximum-amplitude beat, MTO identifies MAP from the crossover between a weakening dicrotic-like notch and an increasing late-diastolic reflection-like notch on the descending limb of the oscillometric waveform. Thus, the MAP estimate is treated as a morphology-transition point in the integrated volume-derived oscillation, rather than as a purely amplitude-defined maximum.

##### S2.6.4 Axial integral expression under a small-oscillation approximation

To obtain an interpretable local representation, the complete time-domain model was linearized only for small pulsatile perturbations around a specified cuff-pressure state. This approximation does not replace the nonlinear simulation. It relates the complex amplitude of local pressure, area, and volume pulsations and shows how these local responses contribute to the integrated cuff oscillation.

$$\widetilde{A}(z, P_c) = C_A^{eff}(z, P_c)\, T_p(z, P_c)\, H_A(z, P_c, \omega)\, e^{-i\omega\tau(z,P_c)}\, \widetilde{P}_{in}. \quad \text{(S134)}$$

Here, $\widetilde{A}(z, P_c)$ is the fundamental complex amplitude of local area pulsation at axial position $z$ and cuff pressure $P_c$, in $mm^2$; $C_A^{eff}(z, P_c)$ is the local effective area compliance, in $mm^2/mmHg$; $T_p(z, P_c)$ is the effective pulsatile transmission factor; $H_A(z, P_c, \omega)$ is the local dynamic area-response function; $\omega$ is the fundamental angular frequency of the cardiac cycle, in rad/s; $\tau(z, P_c)$ is the effective propagation delay to position $z$, in seconds; $e^{-i\omega\tau(z,P_c)}$ is the phase factor induced by propagation delay; $\widetilde{P}_{in}$ is the fundamental complex amplitude of the proximal input pressure, in mmHg; and $i$ is the imaginary unit.

The corresponding complex amplitude of total volume pulsation is

$$\widetilde{V}(P_c) = \int_{\Omega_{cuff}} \widetilde{A}\,(z, P_c)\, dz. \quad \text{(S135)}$$

Here, $\widetilde{V}(P_c)$ is the complex amplitude of total volume pulsation at cuff pressure $P_c$, in $mm^3$ or after conversion in mL; $\widetilde{A}(z, P_c)$ is the local area-pulsation complex amplitude; and $\Omega_{cuff}$ is the cuff-covered axial region.

Under this approximation, the beatwise cuff oscillation amplitude can be written as

$$A(P_c) \approx G_{cuff}(P_c)\, \left|\widetilde{V}(P_c)\right|. \quad \text{(S136)}$$

Here, $A(P_c)$ is the beatwise oscillation amplitude at cuff pressure $P_c$, in mmHg; $G_{cuff}(P_c)$ is the cuff air-chamber volume-to-pressure readout gain, with units interpretable as mmHg/mL; $\widetilde{V}(P_c)$ is the complex amplitude of total volume pulsation; and $|\cdot|$ denotes the complex modulus. Equations S134–S136 are small-oscillation analytical approximations for interpretation. All numerical amplitudes and boundary-related ratios reported from the model were calculated from the full nonlinear time-domain simulation rather than by substituting a universal parameter set into these linearized expressions.

These equations show that the maximum oscillation amplitude depends on local effective compliance, pulsatile transmission, dynamic area response, propagation delay, axial integration, and cuff readout. Thus, even if a local arterial segment has high compliance near MAP, the global cuff oscillation maximum can shift because of diastolic closure-length expansion, distal emptying/refilling, and proximal-distal desynchronization.

##### S2.6.5 Mechanistic DBP amplitude ratio

In this framework, DBP is not assigned by a fixed envelope ratio before the maximum amplitude. Instead, it is linked to the beat in which cyclic compression and reopening begin to emerge in the central high-pressure region after cuff pressure enters the DBP range. For a given subject- and state-specific parameter set p, let $A(P_c; p)$ denote the beatwise oscillation-amplitude envelope generated by the full model as a function of cuff pressure $P_c$. An interior maximum-amplitude pressure $P_{max}$ satisfies

$$A'(P_{max}; p) = 0, \quad A''(P_{max}; p) < 0. \tag{S137}$$

Here, $p$ collects the cuff-loading, vascular pressure–area, collapse, transmission, distal reservoir, delay, and readout parameters used in the model. Equation S137 defines the maximum of the model-generated amplitude envelope; it does not impose that $P_{max}$ must equal invasive MAP.

$$K_b(p) = \frac{A(P_b; p)}{A(P_{max}; p)}, b \in \{DBP, SBP\}. \tag{S138}$$

Here, $P_b$ is the DBP- or SBP-related pressure defined by the operational beat criterion, and $K_b(\boldsymbol{p})$ is the corresponding model-induced amplitude ratio relative to the maximum-amplitude beat. Equations S134–S138 establish the sequence by which local pressure–area response, pulsatile transmission, phase delay, axial integration, and cuff readout determine an individualized ratio. They do not yield a universal closed-form numerical coefficient, because the parameter set p varies across participants, cuff conditions, and measurement states.

As an operational definition comparable with the input DBP, the DBP-related beat is

$$\mathrm{k_{DBP}} = \min\{\mathrm{k}: \bar{\mathrm{P}}_{\mathrm{cuff,k}} > \mathrm{P_{DBP}}\}. \tag{S139}$$

Here, $\mathrm{k_{DBP}}$ is the index of the DBP-related beat; $\bar{\mathrm{P}}_{\mathrm{cuff,k}}$ is the mean cuff pressure of beat k, in mmHg; $\mathrm{P_{DBP}}$ is the input or reference diastolic pressure, in mmHg; and $\min\{\cdot\}$ denotes the earliest beat satisfying the condition. This definition corresponds to the first beat after cuff pressure exceeds DBP. If a morphology-based criterion is used, the beat can be further required to show the DBP-related central compression-reopening pattern.

The oscillation amplitude of the DBP-related beat is

$$\mathrm{A_{DBP}} = \mathrm{A_{k_{DBP}}}. \tag{S140}$$

Here, $\mathrm{A_{DBP}}$ is the oscillation amplitude of the DBP-related beat, in mmHg, and $\mathrm{A_{k_{DBP}}}$ is the beatwise amplitude of beat $\mathrm{k_{DBP}}$.

The mechanistic DBP amplitude ratio is defined as

$$\mathrm{K_{DBP}} = \frac{\mathrm{A_{DBP}}}{\mathrm{A_{max}}}. \tag{S141}$$

Here, $\mathrm{K_{DBP}}$ is the dimensionless ratio between the DBP-related beat amplitude and the maximum beatwise amplitude; $\mathrm{A_{DBP}}$ is the DBP-related beat amplitude; and $\mathrm{A_{max}}$ is the maximum beatwise oscillation amplitude. This ratio is not a prescribed empirical constant. Negative-transmural-pressure collapse and open-closed transition dynamics explain why this beat gains amplitude, but the final $\mathrm{K_{DBP}}$ is computed from the total cuff oscillation amplitude of that beat, not from any isolated local collapse term.

##### S2.6.6 Mechanistic SBP amplitude ratio

In this framework, SBP is not assigned by a fixed envelope ratio after the maximum amplitude. Instead, it is linked to the final beat before cuff pressure exceeds SBP that still preserves effective pulsatile transmission and total volume pulsation. The operational definition is

$$k_{SBP} = \max\{k: \bar{P}_{cuff,k} < P_{SBP}\}. \tag{S142}$$

Here, $k_{SBP}$ is the index of the SBP-related beat; $\bar{P}_{cuff,k}$ is the mean cuff pressure of beat $k$, in mmHg; $P_{SBP}$ is the input or reference systolic pressure, in mmHg; and $\max\{\cdot\}$ denotes the last beat satisfying the condition. This definition corresponds to the last beat before cuff pressure exceeds SBP. If a morphology-based criterion is used, the beat can be further required to retain pre-SBP effective distal transmission or total volume pulsation.

The oscillation amplitude of the SBP-related beat is

$$A_{SBP} = A_{k_{SBP}}. \tag{S143}$$

Here, $A_{SBP}$ is the oscillation amplitude of the SBP-related beat, in mmHg, and $A_{k_{SBP}}$ is the beatwise amplitude of beat $k_{SBP}$.

The mechanistic SBP amplitude ratio is defined as

$$K_{SBP} = \frac{A_{SBP}}{A_{max}}. \tag{S144}$$

Here, $K_{SBP}$ is the dimensionless ratio between the SBP-related beat amplitude and the maximum beatwise amplitude; $A_{SBP}$ is the SBP-related beat amplitude; and $A_{max}$ is the maximum beatwise amplitude. This ratio reflects the combined influence of the functional closure boundary, distal emptying, refilling recovery, proximal-distal propagation delay, local pressure waves, and proximal-distal volume mismatch near high cuff pressure.

These operational definitions correspond to the inflation protocol used in the present analysis. For deflation protocols, the inequality direction and beat-ordering criterion should be adjusted accordingly.

##### S2.6.7 Reduced mechanistic grouping of amplitude-ratio determinants

The preceding sections provide the operational definitions of beatwise oscillation amplitude, the maximum-amplitude beat, the DBP/SBP-related beats, and the mechanistic amplitude ratios. Therefore, the amplitude ratio is not formulated here as an empirical expression determined by a small number of independent physiological constants. Instead, it is treated as an output of the full time-domain unified cuff-artery model. The lower-level variables in the full model include cuff external-pressure distribution, open-segment pressure-area behavior, negative-transmural-pressure collapse, effective transmission loss, distal emptying/refilling, propagation delay, pressure-field remodeling, and cuff air-chamber readout. Directly inserting all lower-level variables into $K_{DBP}$ and $K_{SBP}$ would produce a high-dimensional expression with limited interpretability.

To avoid treating aggregated descriptors as independent and directly measurable scalar physiological parameters, lower-level variables with related mechanistic effects are grouped into six mechanistic domains: cuff loading geometry, open-state arterial compliance, negative-pressure collapse behavior, residual hydraulic conductance and functional disconnection, distal emptying-refilling dynamics, and propagation asynchrony with pressure-field remodeling. These domains are denoted as: $G_1$, $G_2$, $G_3$, $G_4$, $G_5$, $G_6$.

Here, $G_i$ denotes the ith mechanistic domain rather than a single scalar parameter. Each $G_i$ corresponds to a subset of lower-level parameters and state variables. After grouping the lower-level parameters by mechanistic domain, the full lower-level parameter vector can be written as

$$p = \left(p_{G_1}, p_{G_2}, p_{G_3}, p_{G_4}, p_{G_5}, p_{G_6}\right). \tag{S145}$$

Here, p is the full lower-level parameter vector, and $p_{G_i}$ is the parameter subvector belonging to the ith mechanistic domain. This grouping does not alter the preceding definitions of the amplitude ratios; $K_{DBP}$ and $K_{SBP}$ are still computed from the unified time-domain model, axial area integration, cuff air-chamber readout, and beatwise amplitude calculation.

Under this grouping, the DBP and SBP amplitude ratios can be written as model-induced functions of the six lower-level parameter groups:

$$K_{DBP} = F_{DBP}\left(p_{G_1}, p_{G_2}, p_{G_3}, p_{G_4}, p_{G_5}, p_{G_6}\right), \tag{S146}$$

$$K_{SBP} = F_{SBP}\left(p_{G_1}, p_{G_2}, p_{G_3}, p_{G_4}, p_{G_5}, p_{G_6}\right). \tag{S147}$$

Here, $F_{DBP}$ and $F_{SBP}$ denote the DBP and SBP amplitude-ratio functions induced by the unified cuff-artery model. Equations S146 and S147 are not closed-form empirical equations, nor do they assume that $K_{DBP}$ or $K_{SBP}$ is linear. They indicate only that the mechanistic amplitude ratios depend on coupled lower-level mechanistic domains rather than on fixed amplitude ratios.

##### S2.6.8 Six mechanistic domains and their lower-level determinants

The six mechanistic domains are summarized in Supplementary Table 4 and further defined below. Each domain groups lower-level model quantities with related effects on axial volume pulsation and cuff-pressure readout; none should be interpreted as a single directly measured physiological constant.

$G_1$: cuff loading geometry. This domain describes how cuff geometry and arm-cuff coupling shape the axial distribution of external pressure applied to the artery. A compact descriptor may be written as

$$G_1 = \mathcal{C}_\alpha\{\alpha(z)\}, \tag{S148}$$

where $\alpha(z)$ is the normalized axial external-pressure distribution, and $\mathcal{C}_\alpha\{\cdot\}$ denotes a summary functional that captures the concentration, width, and axial location of the pressure-peak region. $G_1$ is influenced by cuff width, bladder structure, wrapping tightness, soft-tissue thickness, fat-to-muscle ratio, soft-tissue compressibility, and the position of the brachial artery relative to the cuff center.

$G_2$: open-state arterial compliance. This domain represents the sensitivity of luminal area to transmural pressure when the vessel remains on the open segment. It can be described by the local slope of the open-segment pressure-area relation:

$$G_2 = \left.\frac{\partial A_{open}}{\partial P_{tm}}\right|_{P_{tm}>0}. \tag{S149}$$

Here, $A_{open}$ is the open-segment luminal area, and $P_{tm}$ is transmural pressure. $G_2$ is affected by baseline arterial diameter, arterial wall elasticity, arterial stiffness, age, vascular tone, pulse pressure, and hypertensive vascular remodeling. A larger $G_2$ increases the ability of pressure pulsation to generate volume pulsation before marked compression develops.

$G_3$: negative-pressure collapse behavior. This domain summarizes the vessel response once external pressure exceeds intraluminal pressure sufficiently to drive negative-transmural-pressure collapse. An effective collapse-susceptibility descriptor may be written as

$$P_{col}^{*} = \inf\left\{P_{neg}: \frac{A_{neg}(P_{neg})}{A_0} \leq \eta_{col}\right\}, \tag{S150}$$

where $P_{neg} = \max(-P_{tm}, 0)$ is the magnitude of negative transmural pressure, $A_{neg}$ is the negative-pressure segment area, $A_0$ is the zero-pressure reference area, and $\eta_{col}$ is a predefined relative-area level used to indicate appreciable collapse. The nonlinearity of collapse progression can be summarized as

$$N_{col} = -\frac{\partial \log(A_{neg}/A_0)}{\partial \log(P_{neg}/P_{ref})}. \tag{S151}$$

Here, $P_{ref}$ is a reference negative transmural pressure. Together, $P_{col}^{*}$, $N_{col}$, closure-memory dynamics, and collapse/reopening time constants constitute $G_3$. This domain is influenced by wall thickness, perivascular soft-tissue support, tissue stiffness, nonlinear wall elasticity, residual slot-like patency, and collapse geometry.

$G_4$: residual hydraulic conductance and functional disconnection. This domain describes the conversion from a small but nonzero residual lumen to effective loss of pressure transmission. The effective conductance may be written as

$$K_{cond}(z,t) = \left(\frac{A(z,t)}{A_0(z)}\right)^{m_{cond}}, \tag{S152}$$

where $K_{cond}$ is the local effective conductance, $A(z,t)$ is the local luminal area, $A_0(z)$ is the corresponding reference area, and $m_{cond}$ is the area-conductance exponent. Functional disconnection is represented when

$$K_{cond}(z,t) \leq K_{crit}, \text{or equivalently} \frac{A(z,t)}{A_0(z)} \leq K_{crit}^{1/m_{cond}}. \tag{S153}$$

Here, $K_{crit}$ is the effective conductance cutoff. This domain is influenced by residual channel area, blood viscosity, distal resistance, collapse morphology, proximal-distal pressure gradient, and residual conducting-channel geometry.

$G_5$: distal emptying-refilling dynamics. This domain represents the effective distal reservoir behavior once proximal-distal communication becomes restricted. For example, one compact descriptor of distal emptying-refilling dynamics can be written as

$$G_5 = \left(\frac{T}{\tau_{empty}}, \eta_{empty}^{\max}, \frac{t_{refill}}{T}\right) \tag{S154}$$

Here, $T/\tau_{empty}$ compares the cardiac-cycle duration with the effective distal emptying time constant; $\eta_{empty}^{\max}$ is the maximum normalized emptying state reached during the relevant beat or episode; and $t_{refill}/T$ is the refill-recovery duration normalized by the cardiac period. Thus, $G_5$ is a three-component descriptor of distal emptying and refilling rather than a single scalar physiological constant. This form avoids algebraic cancellation that would occur if $T/\tau_{empty}$ were multiplied again by normalized resistance and compliance while $\tau_{empty} = R_{out}^{eff} C_{eff}^{d}$.

$G_6$ : propagation asynchrony and pressure-field remodeling. This domain combines compression-related propagation delay and pressure-field reshaping under severe cuff compression. The propagation-asynchrony component can be summarized as

$$G_{6a}(P_{cuff}) = \frac{1}{N_z T} \sum_{j=1}^{N_z} \left|\tau_j(P_{cuff}) - \tau_{j,0}\right|, \tag{S155}$$

where $N_z$ is the number of axial sites, T is the cardiac period, $\tau_j(P_{cuff})$ is the cumulative propagation delay to axial site j at cuff pressure $P_{cuff}$, and $\tau_{j,0}$ is the corresponding baseline delay without marked compression. The pressure-field remodeling component can be summarized as

$$G_{6b} = w_d \Gamma_{dist} + w_r \Gamma_{refl} + w_l \frac{|A_{DBP,wave}| + |A_{SBP,notch}| + |A_{SBP,peak}|}{PP}. \tag{S156}$$

Here, $\Gamma_{dist}$ is the distal pressure-enhancement coefficient, $\Gamma_{refl}$ is the proximal mirror-like reflection gain, $A_{DBP,wave}$ is the DBP-related local pressure-wave amplitude, $A_{SBP,notch}$ and $A_{SBP,peak}$ are the SBP-related notch-like and second-peak amplitudes, PP is pulse pressure, and $w_d$, $w_r$, and $w_\ell$ are normalized weights. $G_6$ is therefore governed by pulse-wave velocity, compression-related additional delay, occluded length, impedance mismatch, distal pressure accumulation, reflected waves, input waveform morphology, and proximal-distal coupling.

##### S2.6.9 Relation to conventional fixed amplitude-ratio methods

Conventional fixed-ratio methods can be written as

$$K_b^{fixed} = c_b, \qquad b \in \{DBP, SBP\}, \tag{S157}$$

where $K_b^{fixed}$ is the fixed amplitude ratio used for blood-pressure point b, and $c_b$ is an empirical constant. This formulation implicitly assumes that the amplitude ratio associated with DBP or SBP is approximately universal across subjects.

In the present mechanistic framework, a conventional fixed ratio can be interpreted as a population-average projection of individualized model-induced functions under a specific population, cuff condition, and measurement protocol:

$$K_b^{fixed} \approx \mathbb{E}_{\mathcal{P}}\left[F_b\left(p_{G_1}, p_{G_2}, p_{G_3}, p_{G_4}, p_{G_5}, p_{G_6}\right)\right], \qquad b \in \{DBP, SBP\}. \tag{S158}$$

Here, $F_b$ is the amplitude-ratio function for blood-pressure point b induced by the unified cuff-artery model, and $\mathbb{E}_{\mathcal{P}}[\cdot]$ denotes expectation over a defined target population and measurement condition. Equation S158 indicates that conventional fixed ratios are not universal physiological constants, but average projections of coupled mechanistic domains in a restricted sample. Thus, when cuff loading, vascular compliance, collapse behavior, residual hydraulic conductance, distal emptying/refilling, propagation asynchrony, or pressure-field remodeling differs across individuals, the appropriate DBP and SBP amplitude ratios should also vary.

##### S2.6.10 Summary

This section reformulates the mechanistic DBP and SBP amplitude ratios as individualized outputs of the unified cuff-artery model rather than fixed empirical ratios. The maximum-amplitude beat corresponds to the maximum cuff-readout of axially integrated subcuff volume pulsation and need not correspond to mean arterial pressure. The DBP and SBP amplitude ratios,

$K_{DBP}$ and $K_{SBP}$, are determined by six coupled mechanistic domains: cuff loading geometry, open-state arterial compliance, negative-pressure collapse behavior, residual hydraulic conductance and functional disconnection, distal emptying-refilling dynamics, and propagation asynchrony with pressure-field remodeling. Conventional fixed amplitude ratios should therefore be regarded as empirical population-level projections under specific cuff-loading and vascular mechanical conditions, not as universal mechanistic constants. This framework provides a mechanistic basis for interindividual variability in oscillometric amplitude ratios and a theoretical interface for model-constrained individualized blood-pressure estimation.

#### *Section S2.7 Clinical performance analyses*

We performed supplementary clinical performance analyses to test whether the primary agreement pattern was preserved under additional empirical comparators, clinically interpretable error thresholds, prespecified subgroup strata, and repeated-measure designs. Unless otherwise stated, analyses used all 630 cuff-measurement runs from 175 participants with invasive arterial reference pressure.

##### S2.7.1 Additional derivative-based comparator analysis

As an additional empirical comparator, we evaluated a derivative-based oscillometric method using the same invasive-reference dataset. BP values derived from this method were compared with invasive BP, KS, and MTO using the agreement metrics described in Section S1.9. The derivative-based method showed wider limits of agreement than MTO across SBP, DBP, and MAP, most notably for SBP, for which the bias was −9.28 mmHg, the SD was 22.75 mmHg, the 95% limits of agreement were −53.87 to 35.31 mmHg, and the RMSE was 24.55 mmHg (Supplementary Table 5). This analysis was included as a supplementary local-feature comparator and was not part of the primary MTO framework.

##### S2.7.2 Error-threshold analysis

To further characterize clinically relevant measurement error, we summarized the proportions of measurements with absolute BP differences exceeding predefined thresholds. Absolute error was defined as the absolute value of the non-invasive estimate minus the corresponding invasive reference pressure. The analysis was performed for thresholds greater than 10, 15, and 20 mmHg (Supplementary Table 6).

MTO had fewer large errors than MAA and KS for all BP parameters. For SBP, absolute error exceeded 10 mmHg in 11.75% of MTO measurements, compared with 46.51% for MAA and 40.00% for KS. Absolute SBP error exceeded 15 mmHg in 2.86% of MTO measurements, compared with 24.92% for MAA and 22.38% for KS, and exceeded 20 mmHg in 0.32% of MTO measurements, compared with 11.11% for MAA and 12.22% for KS. For DBP, the corresponding proportions for errors greater than 10, 15, and 20 mmHg were 10.95%, 1.27%, and 0.16% for MTO; 29.37%, 13.02%, and 4.60% for MAA; and 16.83%, 4.76%, and 0.48% for KS. For MAP, absolute errors greater than 10, 15, and 20 mmHg occurred in 8.89%, 1.59%, and 0.48% of MTO measurements, respectively, compared with 33.02%, 16.03%, and 7.14% for MAA, and 32.06%, 14.29%, and 3.49% for KS. Thus, MTO reduced not only mean error and dispersion but also the frequency of clinically relevant outlying errors.

Cumulative error distributions further characterized the differences in measurement performance among the three methods (Supplementary Fig. 27). Across SBP, DBP, and MAP, the

MTO curves generally rose more steeply and were shifted to the left relative to those of MAA and auscultation, indicating an overall shift toward smaller absolute errors. The separation was already evident at clinically relevant thresholds of 10 and 15 mmHg and persisted across the error range. These distributions therefore show that the improved performance of MTO was not confined to differences in mean bias or summary error metrics, but reflected a broader reduction in absolute measurement error across individual measurements.

##### S2.7.3 Stratified subgroup performance

We assessed stratified agreement performance across predefined measurement and participant strata, including measurement phase, sex, age, BMI and baseline invasive SBP (Supplementary Table 7). These analyses were designed to evaluate consistency rather than to derive subgroup-specific correction rules. The interpretation therefore focused on within-stratum agreement metrics, including bias, SD, 95% limits of agreement (LoA), MAE, RMSE, and correlation.

Across 39 subgroup-parameter cells, defined by 13 strata and three BP parameters, MTO had the lowest SD, narrowest LoA, and lowest RMSE in all 39 cells. MTO also had the lowest MAE in 38 of 39 cells; in the remaining cell, DBP in participants aged 70 years or older, the MAE was nearly identical for MTO and KS (5.30 versus 5.25 mmHg), while MTO retained lower SD, RMSE, and LoA width. This pattern indicates that the main agreement advantage of MTO was not driven by a single measurement condition or participant subgroup.

For inflation runs (N = 227), MTO showed an SBP bias of 0.59 mmHg, an SD of 5.59 mmHg, and 95% LoA of −10.38 to 11.55 mmHg. For deflation runs (N = 403), the corresponding values were 0.28 mmHg, 6.83 mmHg, and −13.11 to 13.66 mmHg, respectively. The same direction of effect was observed for DBP and MAP, for which MTO maintained lower dispersion than MAA and KS within both measurement phases.

Across sex, age, BMI, and baseline invasive SBP strata, MTO continued to show near-zero SBP bias and lower dispersion than the empirical comparators. In the concurrent invasive SBP ≥160 mmHg stratum, the MTO SBP SD was 7.11 mmHg and the 95% LoA were −14.90 to 12.98 mmHg; even in this high-pressure stratum, the LoA remained narrower than those of MAA and KS. Thus, the subgroup analyses support the robustness of morphology-based BP localization rather than suggesting a subgroup-restricted effect.

##### S2.7.4 Between-subgroup tests

Between-subgroup tests were performed on signed measurement errors (noninvasive estimate minus invasive reference) to assess differences across predefined subgroup variables (Supplementary Table 8). These tests were interpreted as supportive heterogeneity analyses, not as evidence for selecting different methods in different subgroups.

For MTO, SBP measurement error did not differ significantly by measurement phase, sex, or age group, but differed across BMI and concurrent invasive-SBP groups. DBP measurement error differed by measurement phase, age group, and BMI group, whereas MAP measurement error differed by sex, BMI group, and concurrent invasive-SBP group. This heterogeneity is expected because measurement-error distributions can change with BP range, vascular state, and measurement conditions. However, these subgroup effects did not reverse the within-stratum ranking: MTO retained lower error dispersion than MAA and KS across the predefined strata.

##### S2.7.5 Repeated-measures agreement sensitivity analysis

Because several cuff-measurement runs were obtained from the same participant, repeated-measures Bland-Altman analyses were performed to evaluate whether the agreement results were affected by within-participant clustering (Supplementary Table 9). The difference between each non-invasive estimate and the invasive reference pressure was modeled with participant as a random intercept. Confidence intervals were estimated by participant-level cluster bootstrap resampling with 5000 replicates.

After accounting for repeated measurements, MTO continued to show minimal bias and narrower LoA than MAA and KS. For SBP, repeated-measures bias was 0.40 mmHg for MTO, compared with −6.79 mmHg for MAA and −6.87 mmHg for KS; the corresponding LoA were −12.35 to 13.16 mmHg for MTO, −27.87 to 14.29 mmHg for MAA, and −27.16 to 13.42 mmHg for KS. For DBP, the MTO bias was −0.12 mmHg with LoA of −12.57 to 12.33 mmHg, compared with 4.51 mmHg and −13.49 to 22.51 mmHg for MAA, and −1.33 mmHg and −15.83 to 13.18 mmHg for KS. For MAP, MTO showed a bias of −0.56 mmHg and LoA of −12.16 to 11.03 mmHg, compared with 2.21 mmHg and −20.60 to 25.02 mmHg for MAA, and −6.94 mmHg and −21.91 to 8.04 mmHg for KS.

Participant-level sensitivity analyses gave consistent estimates. Patient-averaged MTO biases were 0.41 mmHg for SBP, −0.16 mmHg for DBP, and −0.60 mmHg for MAP, with LoA of −10.64 to 11.46 mmHg, −10.99 to 10.68 mmHg, and −10.18 to 8.98 mmHg, respectively. These results indicate that the observed agreement pattern was not driven by unequal numbers of repeated measurements per participant.

##### S2.7.6 Within-participant repeatability of method-specific BP errors

To directly assess repeatability under repeated cuff measurements, we quantified within-participant variability of method-specific BP errors. For each run, error was defined as the non-invasive BP estimate minus the invasive BP reference value. Within-participant error variances were calculated for each participant, method, and BP parameter, then combined across participants to obtain the pooled within-participant error SD. The repeatability coefficient was calculated as $1.96\sqrt{2}SD_w$, where $SD_w$ denotes the pooled within-participant error SD. Differences versus MTO were estimated using participant-level bootstrap resampling with 5000 replicates (Supplementary Table 10).

MTO showed the lowest pooled within-participant error SD and the lowest repeatability coefficient for SBP, DBP, and MAP. For SBP, the pooled within-participant error SD was 3.93 mmHg for MTO, compared with 4.99 mmHg for MAA and 6.18 mmHg for KS. For DBP, the pooled within-participant error SD was 3.78 mmHg for MTO, 6.60 mmHg for MAA, and 4.55 mmHg for KS. For MAP, the corresponding values were 3.96 mmHg for MTO, 8.09 mmHg for MAA, and 4.62 mmHg for KS. All comparator-minus-MTO differences in pooled within-participant SD were positive and statistically significant. These findings show that the smaller error of MTO was accompanied by lower within-participant fluctuation of method-specific BP error across repeated runs.

### Supplementary Tables

##### Supplementary Table 1. Baseline characteristics of the study cohorts

| **Variable** | **Cohort 1 Cardiac catheterization cohort** | **Cohort 2 Pressure-mapping cohort** | **Cohort 3 CTA imaging cohort** |
|---|---|---|---|
| **A. Demographic and anthropometric characteristics** | | | |
| Participants, no. | 178 | 50 | 22 |
| Age, years | 60.79 ± 9.91 | 44.22 ± 18.25 | 35.27 ± 4.62 |
| Male sex, no. (%) | 125 (70.22) | 28 (56.00) | 11 (50.00) |
| Height, cm | 167.70 ± 7.78 | 169.98 ± 8.30 | 172.23 ± 7.30 |
| Weight, kg | 70.48 ± 13.04 | 70.02 ± 13.67 | 68.00 ± 9.25 |
| Body mass index, kg/m² | 24.95 ± 3.49 | 24.13 ± 3.79 | 22.85 ± 1.90 |
| B. Clinical history characteristics of Cohort 1 | | | |
| History of smoking, no. (%) | 77 (43.26) | — | — |
| History of alcohol use, no. (%) | 52 (29.21) | — | — |
| History of cardiac disease, no. (%) | 144 (80.90) | — | — |
| History of diabetes mellitus, no. (%) | 63 (35.39) | — | — |
| History of hypertension, no. (%) | 98 (55.06) | — | — |
| Regular use of antihypertensive medication, no. (%) | 104 (58.43) | — | — |
| History of cerebrovascular disease, no. (%) | 21 (11.80) | — | — |
| History of kidney disease, no. (%) | 9 (5.06) | — | — |
| Relevant family history, no. (%) | 15 (8.43) | — | — |

Note: Values are mean ± SD or number (%). Percentages were calculated using the number of participants in the corresponding cohort as the denominator; for Cohort 1, percentages were calculated using 178 as the denominator unless otherwise specified. Cohort 1 included 175 patients with invasive arterial pressure recordings who were used for invasive-reference BP, agreement, error-distribution, repeated-measures, repeatability, and mechanistic arterial-pressure analyses, and 3 additional patients who contributed venous pressure measurements only and were not included in invasive arterial-pressure reference analyses. Clinical history variables were recorded for Cohort 1 only. Cohort 2 and Cohort 3 were separate healthy-participant cohorts and did not overlap with the catheterization-laboratory cohort. An em dash indicates that the variable was not recorded for that cohort or was not applicable to that cohort-specific analysis; it does not indicate a value of zero. CTA, computed tomographic angiography; no., number; SD, standard deviation.

**Supplementary Table 2. Representative mechanistic parameters for instantiation of the unified cuff–brachial artery interaction model in the reported numerical simulations**

| Parameter group | Parameter | Unit | Representative value | Interpretation / role |
|---|---|---|---|---|
| Geometry and cuff loading | Modeled brachial-artery length $L$ | mm | 120 | Principal compressed brachial-artery segment represented under the cuff. |
| Geometry and cuff loading | Effective cuff width $W_{cuff}$ | mm | 120 | Effective axial width of upper-arm cuff loading. |
| Geometry and cuff loading | Number of axial sites $N_z$ | dimensionless | 30 | Fixed axial discretization for the reported simulations. |
| Geometry and cuff loading | Axial external-pressure shape parameter $\beta$ | dimensionless | 3.5 | Controls the center-high/edge-low external-pressure distribution. |
| Boundary input and protocol | Proximal input pressure $P_{in,prox}$ | mmHg | standardized/measured/constant | Boundary driving pressure; input form does not alter the cuff-artery mechanism. |
| Boundary input and protocol | Input SBP/DBP | mmHg | input-defined | Input systolic and diastolic pressures; not fitted model parameters. |
| Boundary input and protocol | Cuff inflation rate | mmHg/s | 3-5 | Inflation rate used in the representative measurement protocol. |
| Boundary input and protocol | Maximum cuff pressure | mmHg | SBP + 30 | Upper compression level used to cover the range from DBP to above SBP. |
| Pressure propagation | Baseline PWV $c_0$ | m/s | 6 | Representative baseline pulse-wave velocity. |
| Pressure propagation | Closure-related additional delay gain $g_{delay,occ}$ | dimensionless | 4.0 | Scales transmission slowing under severe compression. |
| Pressure-area relation | Positive-transmural-pressure opening-law parameters $A_m$, $P_0$, and $P_1$ | mm² or mmHg | $A_m$ = 22.55/21.61; $P_0$ = 10/6; $P_1$ = 10/20 | Systolic/diastolic opening-segment parameters for the positive-transmural-pressure area law. |
| Collapse and closure | Residual slot-area fraction $f_{leak}$ | dimensionless (A/A0) | 0.003 | Residual slot-like channel retained numerically after near-complete collapse. |
| Collapse and closure | Negative-transmural-pressure collapse reference $P_{buck}$ | mmHg | 2.6 | Reference pressure controlling negative-pressure-driven collapse strength. |

| Parameter group | Parameter | Unit | Representative value | Interpretation / role |
|---|---|---|---|---|
| Collapse and closure | Functional-closure threshold $\zeta_{close}$ | dimensionless (A/A0) | 0.02 | Relative-area criterion for mapping continuous area to effective closure. |
| Distal emptying/refilling | Effective distal reservoir pressure $P_{res,eff}$ | mmHg | 60 | Effective distal pressure attractor after functional interruption; not venous pressure. |
| Distal emptying/refilling | RC/Windkessel emptying time scale $\tau_{empty}$ | s | state-dependent | Event-specific emptying time scale determined by distal compliance and outflow resistance. |
| Distal emptying/refilling | Reconnection time scale | s | 0.030 | Baseline time scale for distal pressure to follow proximal drive outside emptying episodes. |
| Pressure-field remodeling | Distal pressure-enhancement strength $k_Z$ | dimensionless | 3.0 | Scales distal pressure accumulation under transmission limitation and impedance mismatch. |
| Pressure-field remodeling | Distal-enhancement dynamic follower | s | 0.03/0.080 | Rise/decay time constants for distal pressure elevation. |
| Pressure-field remodeling | Proximal mirror-like reflection gain $g_{mirror}$ | dimensionless | 1.0 | Maps distal pressure accumulation to proximal reflection-like pressure components. |
| Pressure-field remodeling | DBP/SBP local pressure-wave amplitudes $A_{DBP}$, $A_N/A_P$ | mmHg | $A_{DBP}$ = 9.2; $A_N/A_P$ = 2.0/2.0 | Amplitudes for DBP local waves and SBP-related notch/second-peak pressure remodeling. |
| Volume readout | Equivalent cuff air volume $V_{0,cuff}$ | mL | 250 | Air-chamber volume used to convert vascular-volume change into recovered cuff pressure. |
| Volume readout | Sampling frequency | Hz | 1000 | Matched to the experimental signal-acquisition frequency. |
| Volume readout | Oscillation extraction band | Hz | 1-15 | Band used to extract model cuff oscillations consistently with experimental processing. |

Note: Values are representative preset values used to instantiate the reported numerical simulations; they are not subject-specific fitted parameters. Except for explicit input variables, parameters were kept fixed across simulations. Input variables include $P_{in,prox}$, SBP/DBP, and the cuff-pressure protocol.

Slash convention: for $A_m$, $P_0$, and $P_1$, "/" separates systolic/diastolic segment values. In nonsegment rows, "/" separates the paired quantities specified in that row, such as rise/decay time constants or $A_N/A_P$ notch/second-peak amplitudes. $\zeta_{close}$ = 0.02 is an approximate reference criterion for effective functional closure. τempty is state-dependent and is determined by effective distal vascular compliance and outflow resistance at the start of each distal-emptying episode, rather than by a fixed preset value.

Input definition: $P_{in,prox}$ supports 3 forms: a standardized arterial-pressure template, a measured invasive arterial-pressure waveform, and a constant-pressure control. These alternative boundary inputs do not alter the core cuff-artery interaction mechanism.

**Supplementary Table 3. Numerical implementation settings for reproducibility of the representative simulations of the unified cuff–brachial artery model**

| Parameter group | Parameter | Unit | Representative value | Interpretation / role |
|---|---|---|---|---|
| Boundary input template | Phase shift, shoulder, notch, and rebound parameters | dimensionless | 0.90; 0.18; 0.10; 0.07 | Template settings used only to generate the standardized proximal boundary pressure waveform. |
| External pressure/propagation | Additional-delay smoothing window and cap | samples/multiplier | 3; 8.0 | Axial smoothing window and per-segment cap for additional propagation delay. |
| Collapse state | $\chi_c/\lambda_p$ auxiliary exponents and weights | Dimensionless; mmHg; s; $s^{-1}$ | $P_\chi$ = 5.0; $n_\chi$ = 1.6; $\tau_\chi$ = 0.030/0.060; $k_{drain,0}$ = 6.0 | Numerical implementation of closure memory, residual patency, and emptying drive. |
| Segment transition | Positive/negative transmural-pressure switching width | mmHg | $P_{sw}$ = 0.20; $w_{sw}$ = 0.90 | Smooth transition between opening and collapse segments. |
| Distal enhancement | Severity smoothing and distal-enhancement parameters | s or dimensionless | $\tau_\sigma$ = 0.03; $\sigma_Z$ = 0.18; $k_Z$ = 3.0 | Slow severity state and local baseline for distal pressure enhancement. |
| Distal enhancement | Dynamic follower rise/decay constants | s | 0.03/0.080 | Finite response time constants for distal pressure elevation. |
| Proximal mirror | Axial mirror pairing and additional propagation delay | dimensionless | $g_{mirror}$ = 1.0; delay scale = 1.0 | Mirror-like mapping of distal pressure increments to the proximal pressure field. |
| DBP/SBP local waves | Phase-localization kernel locations and widths | phase | $\phi_{DBP}$ = 0.935, $\sigma_{DBP}$ = 0.075; $\phi_{SBP,N}$ = 0.258, $\sigma_{SBP,N}$ = 0.024; $\phi_{SBP,P}$ = 0.385, $\sigma_{SBP,P}$ = 0.032 | Intrabeat localization of DBP- and SBP-related local pressure waves. |
| Volume readout | Short-window smoothing of volume and recovered cuff pressure | samples | 5/3 | Smoothing windows applied to total volume and recovered cuff pressure, respectively. |
| Output processing | Filter order | order | 3 | Filter order used for cuff-oscillation extraction. |

Note: These settings specify the numerical implementation used to reproduce the representative simulations. They do not constitute additional physiological mechanisms and were not used as free fitting parameters for blood-pressure estimation.

The 4 boundary-input template values are, in order, the phase-shift parameter and the shoulder, notch, and rebound template-amplitude parameters. Standardized-template settings are used only when generating a simulated proximal input pressure waveform; measured invasive input and constant-pressure control inputs do not use this template. Slash convention: "/" denotes paired quantities specified in the row, including $\tau_\chi$ rise/decay time constants, distal-enhancement follower rise/decay constants, and volume/recovered-cuff-pressure smoothing windows.

**Supplementary Table 4. Six mechanistic domains underlying the mechanistic amplitude-ratio framework**

| Domain | Name | Main mechanistic meaning |
|---|---|---|
| $G_1$ | Cuff loading geometry | Cuff width, loading geometry, axial concentration of external pressure, width of the central pressure-peak region, and the position of the brachial artery relative to the cuff center. |
| $G_2$ | Open-state arterial compliance | Luminal-area response to pulsatile intraluminal pressure under positive transmural pressure, reflecting the overall influence of the open-segment pressure-area law. |
| $G_3$ | Negative-pressure collapse behavior | Susceptibility to marked area reduction under negative transmural pressure, collapse nonlinearity, closure memory, and collapse/reopening dynamics. |
| $G_4$ | Residual hydraulic conductance and functional disconnection | Residual slot-like patency, area-conductance mapping, effective conductance cutoff, and the functional disconnection boundary. |
| $G_5$ | Distal emptying-refilling dynamics | Distal reservoir pressure, distal compliance, outflow resistance, emptying time scale, refilling capacity, and reconnection dynamics. |
| $G_6$ | Propagation asynchrony and pressure-field remodeling | Compression-related additional delay, proximal-distal desynchronization, distal pressure enhancement, proximal mirror-like reflection, and local pressure waves. |

Note: The six domains group lower-level model parameters and state variables according to their shared effects on axial volume pulsation and cuff-pressure readout. Each domain denotes a mechanistic grouping rather than a single directly measured scalar physiological parameter.

**Supplementary Table 5. Overall agreement of the derivative method, auscultatory method, and MTO against invasive blood pressure**

| Parameter | Method | Bias (95%LoA) (mmHg) | SD (mmHg) | MAE (mmHg) | RMSE (mmHg) | Correlation | P value |
|---|---|---|---|---|---|---|---|
| SBP | Derivative method | -9.28 (-53.87, 35.31) | 22.75 | 18.52 | 24.55 | 0.49 | <0.001 |
| | KS | -6.58 (-26.78, 13.61) | 10.30 | 9.67 | 12.22 | 0.89 | <0.001 |
| | MTO | 0.39 (-12.17, 12.95) | 6.41 | 4.79 | 6.41 | 0.96 | 0.100 |
| DBP | Derivative method | 4.65 (-21.59, 30.89) | 13.39 | 10.36 | 14.16 | 0.48 | <0.001 |
| | KS | -1.15 (-15.35, 13.04) | 7.24 | 5.63 | 7.33 | 0.84 | <0.001 |
| | MTO | 0.14 (-12.11, 12.39) | 6.25 | 5.05 | 6.25 | 0.87 | 0.388 |
| MAP | Derivative method | 3.50 (-27.15, 34.15) | 15.64 | 11.46 | 16.01 | 0.52 | <0.001 |
| | KS | -6.77 (-21.32, 7.79) | 7.42 | 8.08 | 10.04 | 0.87 | <0.001 |
| | MTO | -0.37 (-11.97, 11.24) | 5.92 | 4.59 | 5.93 | 0.92 | 0.281 |

Note: Values are shown for all 630 cuff-measurement runs. Bias was calculated as noninvasive estimate minus invasive reference pressure. LoA denotes limits of agreement, calculated as bias ± 1.96 × SD. Correlation denotes Pearson correlation coefficient. KS denotes Korotkoff sound auscultatory method; MTO denotes Morphology-Transition Oscillometry; SBP, systolic blood pressure; DBP, diastolic blood pressure; MAP, mean arterial pressure; MAE, mean absolute error; RMSE, root mean square error.

**Supplementary Table 6. Error-threshold analysis of the maximum-amplitude algorithm, auscultatory method, and MTO against invasive blood pressure**

| Parameter | Method | Absolute difference >10 mmHg (%) | Absolute difference >15 mmHg (%) | Absolute difference >20 mmHg (%) |
|---|---|---|---|---|
| SBP | MAA | 46.51 | 24.92 | 11.11 |
| | KS | 40.00 | 22.38 | 12.22 |
| | MTO | 11.75 | 2.86 | 0.32 |
| DBP | MAA | 29.37 | 13.02 | 4.60 |
| | KS | 16.83 | 4.76 | 0.48 |
| | MTO | 10.95 | 1.27 | 0.16 |
| MAP | MAA | 33.02 | 16.03 | 7.14 |
| | KS | 32.06 | 14.29 | 3.49 |
| | MTO | 8.89 | 1.59 | 0.48 |

Note: Values are shown as percentages of all cuff-measurement runs. Absolute difference was calculated as the absolute value of the noninvasive estimate minus the corresponding invasive reference pressure. MAA denotes maximum-amplitude algorithm; KS denotes Korotkoff sound auscultatory method; MTO denotes Morphology-Transition Oscillometry; SBP, systolic blood pressure; DBP, diastolic blood pressure; MAP, mean arterial pressure.

**Supplementary Table 7. Subgroup-specific performance of noninvasive blood pressure measurement methods (Bland-Altman and correlation analyses)**

| Subgroup | Parameter | Method | Bias (mmHg) | 95% LoA (mmHg) | SD (mmHg) | MAE (mmHg) | RMSE (mmHg) | Correlation | Adjusted P value |
|---|---|---|---|---|---|---|---|---|---|
| Inflation (No. of measurements=227) | SBP | MAA | -7.39 | -25.64 to 10.86 | 9.31 | 9.85 | 11.87 | 0.912 | <0.001 |
| | | KS | -5.27 | -24.56 to 14.01 | 9.84 | 8.78 | 11.14 | 0.907 | <0.001 |
| | | MTO | 0.59 | -10.38 to 11.55 | 5.59 | 4.18 | 5.61 | 0.970 | 0.552 |
| | DBP | MAA | 8.42 | -9.24 to 26.07 | 9.01 | 9.99 | 12.31 | 0.808 | <0.001 |
| | | KS | 0.11 | -13.19 to 13.41 | 6.78 | 5.26 | 6.77 | 0.855 | 0.605 |
| | | MTO | 2.03 | -8.02 to 12.07 | 5.13 | 4.52 | 5.50 | 0.915 | <0.001 |
| | MAP | MAA | 4.13 | -15.84 to 24.10 | 10.19 | 7.79 | 10.97 | 0.829 | <0.001 |
| | | KS | -4.95 | -18.49 to 8.58 | 6.91 | 6.84 | 8.49 | 0.890 | <0.001 |
| | | MTO | 0.00 | -11.27 to 11.26 | 5.75 | 4.50 | 5.73 | 0.926 | 0.729 |
| Deflation (No. of measurements=403) | SBP | MAA | -6.46 | -28.46 to 15.55 | 11.23 | 10.48 | 12.94 | 0.861 | <0.001 |
| | | KS | -7.32 | -27.90 to 13.25 | 10.50 | 10.17 | 12.79 | 0.883 | <0.001 |
| | | MTO | 0.28 | -13.11 to 13.66 | 6.83 | 5.13 | 6.83 | 0.949 | 0.297 |
| | DBP | MAA | 2.84 | -14.00 to 19.69 | 8.59 | 6.81 | 9.04 | 0.772 | <0.001 |
| | | KS | -1.87 | -16.38 to 12.65 | 7.40 | 5.84 | 7.63 | 0.830 | <0.001 |
| | | MTO | -0.92 | -13.81 to 11.96 | 6.57 | 5.35 | 6.63 | 0.851 | 0.021 |
| | MAP | MAA | 1.39 | -22.45 to 25.22 | 12.16 | 9.35 | 12.22 | 0.746 | 0.165 |
| | | KS | -7.79 | -22.52 to 6.95 | 7.52 | 8.78 | 10.82 | 0.862 | <0.001 |
| | | MTO | -0.57 | -12.36 to 11.22 | 6.01 | 4.65 | 6.03 | 0.912 | 0.179 |
| Female (No. of measurements=177) | SBP | MAA | -6.84 | -28.71 to 15.02 | 11.16 | 10.83 | 13.06 | 0.883 | <0.001 |
| | | KS | -8.31 | -29.87 to 13.26 | 11.00 | 11.15 | 13.76 | 0.888 | <0.001 |
| | | MTO | -0.17 | -14.18 to 13.84 | 7.15 | 5.37 | 7.13 | 0.948 | 0.998 |
| | DBP | MAA | 4.04 | -13.56 to 21.65 | 8.98 | 7.87 | 9.83 | 0.643 | <0.001 |
| | | KS | -0.95 | -15.45 to 13.54 | 7.40 | 5.82 | 7.44 | 0.754 | 0.134 |
| | | MTO | 0.31 | -12.89 to 13.52 | 6.74 | 5.53 | 6.72 | 0.791 | 0.648 |
| | MAP | MAA | 3.37 | -23.87 to 30.61 | 13.90 | 10.70 | 14.26 | 0.655 | 0.005 |
| | | KS | -8.19 | -23.10 to 6.72 | 7.61 | 8.89 | 11.16 | 0.843 | <0.001 |
| | | MTO | -2.12 | -13.66 to 9.42 | 5.89 | 4.60 | 6.24 | 0.903 | <0.001 |
| Male (No. of measurements=453) | SBP | MAA | -6.77 | -27.07 to 13.53 | 10.36 | 10.03 | 12.37 | 0.881 | <0.001 |
| | | KS | -5.91 | -25.41 to 13.59 | 9.95 | 9.09 | 11.56 | 0.896 | <0.001 |
| | | MTO | 0.60 | -11.33 to 12.54 | 6.09 | 4.56 | 6.11 | 0.960 | 0.134 |

| | | | | | | | | | |
|---|---|---|---|---|---|---|---|---|---|
| | DBP | MAA | 5.17 | -12.85 to 23.18 | 9.19 | 7.99 | 10.54 | 0.794 | <0.001 |
| | | KS | -1.23 | -15.32 to 12.86 | 7.19 | 5.56 | 7.29 | 0.855 | <0.001 |
| | | MTO | 0.07 | -11.80 to 11.94 | 6.06 | 4.86 | 6.05 | 0.887 | 0.684 |
| | MAP | MAA | 1.99 | -18.58 to 22.55 | 10.49 | 8.03 | 10.67 | 0.818 | <0.001 |
| | | KS | -6.21 | -20.49 to 8.07 | 7.29 | 7.76 | 9.57 | 0.879 | <0.001 |
| | | MTO | 0.32 | -11.05 to 11.68 | 5.80 | 4.59 | 5.80 | 0.925 | 0.283 |
| Age < 60 (No. of measurements=234) | SBP | MAA | -6.15 | -27.05 to 14.75 | 10.66 | 9.83 | 12.29 | 0.876 | <0.001 |
| | | KS | -5.81 | -25.83 to 14.22 | 10.22 | 9.22 | 11.73 | 0.897 | <0.001 |
| | | MTO | 0.39 | -11.74 to 12.53 | 6.19 | 4.73 | 6.19 | 0.959 | 0.484 |
| | DBP | MAA | 3.54 | -17.80 to 24.88 | 10.89 | 8.11 | 11.43 | 0.749 | <0.001 |
| | | KS | -1.33 | -17.02 to 14.36 | 8.01 | 6.15 | 8.10 | 0.813 | 0.013 |
| | | MTO | -1.02 | -13.74 to 11.70 | 6.49 | 5.21 | 6.55 | 0.869 | 0.038 |
| | MAP | MAA | -0.05 | -20.34 to 20.25 | 10.36 | 7.56 | 10.33 | 0.812 | 0.990 |
| | | KS | -6.70 | -22.17 to 8.78 | 7.89 | 8.34 | 10.34 | 0.851 | <0.001 |
| | | MTO | 0.29 | -10.86 to 11.45 | 5.69 | 4.35 | 5.69 | 0.926 | 0.484 |
| 60 ≤ Age < 70 (No. of measurements=295) | SBP | MAA | -7.03 | -28.23 to 14.17 | 10.82 | 10.67 | 12.88 | 0.870 | <0.001 |
| | | KS | -7.36 | -27.90 to 13.18 | 10.48 | 10.28 | 12.79 | 0.884 | <0.001 |
| | | MTO | 0.43 | -12.65 to 13.52 | 6.68 | 4.96 | 6.68 | 0.948 | 0.533 |
| | DBP | MAA | 5.04 | -10.28 to 20.36 | 7.82 | 7.51 | 9.29 | 0.766 | <0.001 |
| | | KS | -1.76 | -14.69 to 11.18 | 6.60 | 5.36 | 6.82 | 0.823 | <0.001 |
| | | MTO | 0.15 | -11.79 to 12.09 | 6.09 | 4.84 | 6.08 | 0.841 | 0.790 |
| | MAP | MAA | 3.47 | -21.36 to 28.30 | 12.67 | 9.91 | 13.12 | 0.739 | <0.001 |
| | | KS | -7.46 | -20.81 to 5.89 | 6.81 | 8.28 | 10.10 | 0.873 | <0.001 |
| | | MTO | -0.96 | -12.47 to 10.54 | 5.87 | 4.67 | 5.94 | 0.901 | 0.038 |
| Age ≥ 70 (No. of measurements=101) | SBP | MAA | -7.59 | -26.52 to 11.33 | 9.66 | 10.02 | 12.25 | 0.930 | <0.001 |
| | | KS | -6.11 | -25.53 to 13.31 | 9.91 | 8.91 | 11.60 | 0.925 | <0.001 |
| | | MTO | 0.23 | -11.82 to 12.29 | 6.15 | 4.42 | 6.12 | 0.971 | 0.314 |
| | DBP | MAA | 7.34 | -7.75 to 22.43 | 7.70 | 8.89 | 10.61 | 0.782 | <0.001 |
| | | KS | 1.02 | -12.37 to 14.41 | 6.83 | 5.25 | 6.87 | 0.814 | 0.038 |
| | | MTO | 2.78 | -7.64 to 13.20 | 5.32 | 5.30 | 5.98 | 0.880 | <0.001 |
| | MAP | MAA | 4.78 | -14.35 to 23.91 | 9.76 | 8.34 | 10.82 | 0.839 | <0.001 |
| | | KS | -4.89 | -20.09 to 10.31 | 7.76 | 6.89 | 9.14 | 0.890 | <0.001 |
| | | MTO | -0.16 | -12.82 to 12.49 | 6.46 | 4.93 | 6.43 | 0.927 | 0.902 |
| BMI < 24 (No. of measurements=248) | SBP | MAA | -8.34 | -26.06 to 9.37 | 9.04 | 10.12 | 12.29 | 0.929 | <0.001 |
| | | KS | -7.53 | -27.69 to 12.63 | 10.29 | 10.13 | 12.73 | 0.911 | <0.001 |
| | | MTO | -0.66 | -12.79 to 11.47 | 6.19 | 4.51 | 6.21 | 0.968 | 0.179 |

| | | | | | | | | | |
|---|---|---|---|---|---|---|---|---|---|
| | DBP | MAA | 4.86 | -14.73 to 24.45 | 9.99 | 8.29 | 11.09 | 0.739 | <0.001 |
| | | KS | -1.95 | -13.94 to 10.03 | 6.11 | 5.01 | 6.41 | 0.866 | <0.001 |
| | | MTO | 0.16 | -11.31 to 11.63 | 5.85 | 4.82 | 5.84 | 0.882 | 0.618 |
| | MAP | MAA | 1.45 | -19.97 to 22.87 | 10.93 | 8.07 | 11.00 | 0.824 | 0.074 |
| | | KS | -7.70 | -21.25 to 5.85 | 6.91 | 8.60 | 10.34 | 0.902 | <0.001 |
| | | MTO | -0.99 | -11.61 to 9.63 | 5.42 | 4.24 | 5.50 | 0.941 | 0.039 |
| 24 ≤ BMI < 28 (No. of measurements=283) | SBP | MAA | -4.79 | -27.32 to 17.74 | 11.50 | 10.02 | 12.43 | 0.841 | <0.001 |
| | | KS | -5.54 | -25.90 to 14.81 | 10.38 | 9.33 | 11.76 | 0.879 | <0.001 |
| | | MTO | 1.21 | -11.40 to 13.82 | 6.43 | 4.90 | 6.53 | 0.949 | 0.007 |
| | DBP | MAA | 5.63 | -10.94 to 22.21 | 8.46 | 8.06 | 10.15 | 0.801 | <0.001 |
| | | KS | 0.00 | -15.07 to 15.06 | 7.69 | 5.98 | 7.67 | 0.820 | 0.890 |
| | | MTO | 1.01 | -11.35 to 13.37 | 6.31 | 5.21 | 6.38 | 0.867 | 0.038 |
| | MAP | MAA | 3.66 | -20.35 to 27.67 | 12.25 | 9.62 | 12.76 | 0.726 | <0.001 |
| | | KS | -5.80 | -20.55 to 8.95 | 7.52 | 7.54 | 9.49 | 0.844 | <0.001 |
| | | MTO | 0.60 | -11.60 to 12.80 | 6.23 | 4.87 | 6.24 | 0.897 | 0.141 |
| BMI ≥ 28 (No. of measurements=99) | SBP | MAA | -8.65 | -29.30 to 12.00 | 10.54 | 11.25 | 13.59 | 0.843 | <0.001 |
| | | KS | -7.19 | -26.65 to 12.28 | 9.93 | 9.47 | 12.22 | 0.866 | <0.001 |
| | | MTO | 0.66 | -12.23 to 13.54 | 6.57 | 5.19 | 6.57 | 0.946 | 0.434 |
| | DBP | MAA | 2.61 | -14.00 to 19.21 | 8.47 | 6.80 | 8.82 | 0.754 | 0.004 |
| | | KS | -2.43 | -18.29 to 13.43 | 8.09 | 6.22 | 8.41 | 0.788 | 0.014 |
| | | MTO | -2.41 | -14.98 to 10.16 | 6.41 | 5.19 | 6.82 | 0.851 | 0.007 |
| | MAP | MAA | 1.02 | -20.05 to 22.09 | 10.75 | 8.20 | 10.74 | 0.744 | 0.571 |
| | | KS | -7.18 | -23.02 to 8.66 | 8.08 | 8.30 | 10.78 | 0.829 | <0.001 |
| | | MTO | -1.58 | -13.07 to 9.92 | 5.86 | 4.69 | 6.04 | 0.915 | 0.058 |
| SBP < 140 (No. of measurements=249) | SBP | MAA | -3.66 | -22.73 to 15.41 | 9.73 | 8.42 | 10.38 | 0.632 | <0.001 |
| | | KS | -5.43 | -23.16 to 12.30 | 9.05 | 8.39 | 10.54 | 0.642 | <0.001 |
| | | MTO | 1.46 | -9.91 to 12.83 | 5.80 | 4.54 | 5.97 | 0.854 | 0.007 |
| | DBP | MAA | 3.67 | -11.75 to 19.08 | 7.86 | 6.63 | 8.66 | 0.704 | <0.001 |
| | | KS | -0.28 | -13.31 to 12.75 | 6.65 | 5.11 | 6.64 | 0.807 | 0.249 |
| | | MTO | -0.08 | -11.39 to 11.22 | 5.77 | 4.69 | 5.76 | 0.818 | 0.884 |
| | MAP | MAA | 1.18 | -16.34 to 18.70 | 8.94 | 6.62 | 9.00 | 0.624 | 0.236 |
| | | KS | -5.32 | -18.10 to 7.47 | 6.52 | 6.93 | 8.41 | 0.763 | <0.001 |
| | | MTO | 1.20 | -8.94 to 11.35 | 5.18 | 4.16 | 5.30 | 0.849 | <0.001 |
| 140 ≤ SBP < 160 (No. of measurements=180) | SBP | MAA | -7.70 | -27.35 to 11.95 | 10.03 | 10.35 | 12.62 | 0.354 | <0.001 |
| | | KS | -5.76 | -25.77 to 14.25 | 10.21 | 9.03 | 11.70 | 0.374 | <0.001 |
| | | MTO | 0.41 | -11.59 to 12.41 | 6.12 | 4.53 | 6.12 | 0.571 | 0.434 |

| | | | | | | | | | |
|---|---|---|---|---|---|---|---|---|---|
| | DBP | MAA | 4.52 | -11.85 to 20.89 | 8.35 | 7.47 | 9.48 | 0.703 | <0.001 |
| | | KS | -1.55 | -16.03 to 12.93 | 7.39 | 5.65 | 7.53 | 0.816 | 0.005 |
| | | MTO | -0.15 | -11.61 to 11.32 | 5.85 | 4.76 | 5.84 | 0.846 | 0.790 |
| | MAP | MAA | 2.00 | -19.17 to 23.17 | 10.80 | 8.85 | 10.95 | 0.477 | 0.024 |
| | | KS | -6.49 | -21.38 to 8.40 | 7.60 | 7.94 | 9.98 | 0.687 | <0.001 |
| | | MTO | -0.31 | -11.99 to 11.37 | 5.96 | 4.66 | 5.95 | 0.815 | 0.692 |
| SBP ≥ 160 (No. of measurements=201) | SBP | MAA | -9.86 | -31.52 to 11.79 | 11.05 | 12.44 | 14.79 | 0.657 | <0.001 |
| | | KS | -8.75 | -31.27 to 13.78 | 11.49 | 11.83 | 14.42 | 0.671 | <0.001 |
| | | MTO | -0.96 | -14.90 to 12.98 | 7.11 | 5.32 | 7.16 | 0.808 | 0.227 |
| | DBP | MAA | 6.61 | -14.73 to 27.96 | 10.89 | 10.03 | 12.72 | 0.687 | <0.001 |
| | | KS | -1.88 | -17.03 to 13.27 | 7.73 | 6.27 | 7.94 | 0.791 | 0.002 |
| | | MTO | 0.67 | -13.26 to 14.61 | 7.11 | 5.76 | 7.12 | 0.815 | 0.147 |
| | MAP | MAA | 4.20 | -24.29 to 32.69 | 14.54 | 11.41 | 15.09 | 0.527 | <0.001 |
| | | KS | -8.80 | -24.25 to 6.64 | 7.88 | 9.63 | 11.80 | 0.752 | <0.001 |
| | | MTO | -2.37 | -14.48 to 9.74 | 6.18 | 5.07 | 6.60 | 0.856 | <0.001 |

Note: Values are based on measurement-level analyses. The 95% limits of agreement (LoA) were calculated as bias ± 1.96 × SD. MAE denotes mean absolute error, and RMSE denotes root mean square error. Correlation coefficients are Pearson correlation coefficients between noninvasive and invasive measurements. Adjusted P values were derived from two-sided one-sample Wilcoxon signed-rank tests assessing whether the subgroup-specific bias differed from zero and were corrected within each measurement method using the Benjamini–Hochberg false discovery rate procedure. Subgroups based on SBP were assigned according to the concurrent invasive SBP for each measurement. P values <0.001 are reported as <0.001. MAA, maximum-amplitude algorithm; KS, Korotkoff sound method; MTO, Morphology-Transition Oscillometry; SBP, systolic blood pressure; DBP, diastolic blood pressure; MAP, mean arterial pressure; SD, standard deviation.

**Supplementary Table 8. Between-subgroup comparisons of measurement error across predefined subgroup variables**

| Subgroup Variable | Parameter | Method | Test | Adjusted P value | Post hoc results |
|---|---|---|---|---|---|
| Inflation/Deflation | SBP | MAA | Mann-Whitney U | 0.394 | |
| | | KS | | 0.041 | |
| | | MTO | | 0.894 | |
| | DBP | MAA | | <0.001 | |
| | | KS | | <0.001 | |
| | | MTO | | <0.001 | |
| | MAP | MAA | | <0.001 | |
| | | KS | | <0.001 | |
| | | MTO | | 0.216 | |
| Gender | SBP | MAA | | 0.898 | |
| | | KS | | 0.012 | |
| | | MTO | | 0.397 | |
| | DBP | MAA | | 0.215 | |
| | | KS | | 0.457 | |
| | | MTO | | 0.823 | |
| | MAP | MAA | | 0.515 | |
| | | KS | | 0.017 | |
| | | MTO | | <0.001 | |
| Age group | SBP | MAA | Kruskal-Wallis | 0.687 | Main test not significant |
| | | KS | | 0.206 | Main test not significant |
| | | MTO | | 0.894 | Main test not significant |
| | DBP | MAA | | <0.001 | Age < 60 vs 60 ≤ Age < 70; Age < 60 vs Age ≥ 70; 60 ≤ Age < 70 vs Age ≥ 70 |
| | | KS | | <0.001 | Age < 60 vs Age ≥ 70; 60 ≤ Age < 70 vs Age ≥ 70 |
| | | MTO | | <0.001 | Age < 60 vs 60 ≤ Age < 70; Age < 60 vs Age ≥ 70; 60 ≤ Age < 70 vs Age ≥ 70 |
| | MAP | MAA | | <0.001 | Age < 60 vs 60 ≤ Age < 70; Age < 60 vs Age ≥ 70 |
| | | KS | | 0.006 | Age < 60 vs Age ≥ 70; 60 ≤ Age < 70 vs Age ≥ 70 |
| | | MTO | | 0.068 | Main test not significant |
| BMI group | SBP | MAA | | <0.001 | BMI < 24 vs 24 ≤ BMI < 28; 24 ≤ BMI < 28 vs BMI ≥ 28 |
| | | KS | | 0.080 | Main test not significant |
| | | MTO | | 0.004 | BMI < 24 vs 24 ≤ BMI < 28 |
| | DBP | MAA | | 0.015 | BMI < 24 vs BMI ≥ 28; 24 ≤ BMI < 28 vs BMI ≥ 28 |
| | | KS | | 0.008 | BMI < 24 vs 24 ≤ BMI < 28; 24 ≤ BMI < 28 vs BMI ≥ 28 |
| | | MTO | | <0.001 | BMI < 24 vs BMI ≥ 28; 24 ≤ BMI < 28 vs BMI ≥ 28 |
| | MAP | MAA | | 0.037 | BMI < 24 vs 24 ≤ BMI < 28; 24 ≤ BMI < 28 vs BMI ≥ 28 |
| | | KS | | 0.012 | BMI < 24 vs 24 ≤ BMI < 28 |

| | | | | |
|---|---|---|---|---|
| | | MTO | 0.003 | BMI < 24 vs 24 ≤ BMI < 28; 24 ≤ BMI < 28 vs BMI ≥ 28 |
| SBP group | SBP | MAA | <0.001 | SBP < 140 vs 140 ≤ SBP < 160; SBP < 140 vs SBP ≥ 160; 140 ≤ SBP < 160 vs SBP ≥ 160 |
| | | KS | 0.003 | SBP < 140 vs SBP ≥ 160; 140 ≤ SBP < 160 vs SBP ≥ 160 |
| | | MTO | 0.007 | SBP < 140 vs SBP ≥ 160 |
| | DBP | MAA | <0.001 | SBP < 140 vs SBP ≥ 160; 140 ≤ SBP < 160 vs SBP ≥ 160 |
| | | KS | 0.163 | Main test not significant |
| | | MTO | 0.216 | Main test not significant |
| | MAP | MAA | 0.008 | SBP < 140 vs SBP ≥ 160 |
| | | KS | <0.001 | SBP < 140 vs SBP ≥ 160; 140 ≤ SBP < 160 vs SBP ≥ 160 |
| | | MTO | <0.001 | SBP < 140 vs 140 ≤ SBP < 160; SBP < 140 vs SBP ≥ 160; 140 ≤ SBP < 160 vs SBP ≥ 160 |

Note: Between-subgroup comparisons were conducted at the measurement level using measurement error. Mann–Whitney U tests were used for subgroup variables with two levels, and Kruskal–Wallis tests were used for variables with three levels. Overall P values were adjusted within each measurement method for 15 comparisons, comprising five subgroup variables and three blood pressure parameters, using the Benjamini–Hochberg false discovery rate procedure. When the overall adjusted P value was <0.05 for a three-level subgroup variable, Dunn's test was used for post hoc pairwise comparisons, with P values adjusted using the Benjamini–Hochberg procedure; only statistically significant pairs are listed. SBP categories were assigned separately for each measurement according to the concurrent invasive SBP. MAA, maximum-amplitude algorithm; KS, Korotkoff sound method; MTO, Morphology-Transition Oscillometry; SBP, systolic blood pressure; DBP, diastolic blood pressure; MAP, mean arterial pressure.

**Supplementary Table 9. Repeated-measures agreement and sensitivity analyses accounting for multiple cuff-measurements**

| Parameter | Method | Repeated-measures BA with cluster bootstrap | | | Participant-level mean | |
|---|---|---|---|---|---|---|
| | | Bias (mmHg) | Lower 95% LoA [95% CI] (mmHg) | Upper 95% LoA [95% CI] (mmHg) | Bias (mmHg) | 95% LoA (mmHg) |
| **SBP** | MAA | -6.79 | -27.87 [-29.93, -25.59] | 14.29 [11.66, 17.01] | -6.78 | -26.24 to 12.68 |
| | KS | -6.87 | -27.16 [-29.10, -25.04] | 13.42 [11.54, 15.18] | -6.91 | -24.49 to 10.67 |
| | MTO | 0.40 | -12.35 [-14.05, -10.68] | 13.16 [11.75, 14.53] | 0.41 | -10.64 to 11.46 |
| **DBP** | MAA | 4.51 | -13.49 [-16.36, -10.78] | 22.51 [19.94, 25.32] | 4.41 | -10.04 to 18.87 |
| | KS | -1.33 | -15.83 [-17.44, -14.16] | 13.18 [11.50, 14.74] | -1.37 | -13.91 to 11.18 |
| | MTO | -0.12 | -12.57 [-14.01, -11.13] | 12.33 [11.01, 13.58] | -0.16 | -10.99 to 10.68 |
| **MAP** | MAA | 2.21 | -20.60 [-23.25, -18.05] | 25.02 [21.93, 28.00] | 2.17 | -16.45 to 20.79 |
| | KS | -6.94 | -21.91 [-23.85, -20.00] | 8.04 [6.50, 9.52] | -7.04 | -19.90 to 5.82 |
| | MTO | -0.56 | -12.16 [-13.70, -10.70] | 11.03 [9.88, 12.13] | -0.60 | -10.18 to 8.98 |

Note: The analysis dataset comprised 630 paired measurements from 175 participants. In the repeated-measures Bland–Altman analysis, between-participant and within-participant variance components were estimated using a random-intercept mixed-effects model. The 95% confidence intervals (95% CI) for the lower and upper limits of agreement (LoA) were obtained from 5,000 participant-level cluster bootstrap resamples. In the participant-level mean analysis, all available paired measurements were averaged within each participant before the bias and limits of agreement were calculated. LoA denotes limits of agreement; MTO, Morphology-Transition Oscillometry; MAA, maximum-amplitude algorithm; KS, Korotkoff sound; SBP, systolic blood pressure; DBP, diastolic blood pressure; MAP, mean arterial pressure.

**Supplementary Table 10. Within-participant variability of blood pressure measurement errors**

| Parameter | Method | Pooled within-participant error SD (mmHg) | Difference vs. MTO in pooled SD [95% CI] (mmHg) |
|---|---|---|---|
| SBP | MAA | 4.99 | 1.06 [0.67 to 1.46] |
| | KS | 6.18 | 2.25 [1.65 to 2.85] |
| | MTO | 3.93 | Reference |
| DBP | MAA | 6.60 | 2.82 [0.87 to 5.12] |
| | KS | 4.55 | 0.77 [0.33 to 1.25] |
| | MTO | 3.78 | Reference |
| MAP | MAA | 8.09 | 4.13 [2.73 to 5.71] |
| | KS | 4.62 | 0.65 [0.29 to 1.03] |
| | MTO | 3.96 | Reference |

Note. Within-participant variability was calculated from method-specific BP errors, defined as the non-invasive BP estimate minus the invasive BP reference value, across repeated cuff-measurements within each participant. The pooled within-participant error SD was calculated by combining participant-level within-participant variances weighted by degrees of freedom. Differences versus MTO were calculated as comparator minus MTO; therefore, positive values indicate greater within-participant error variability than MTO. MAA, maximum amplitude algorithm; KS, Korotkoff sounds method; MTO, Morphology-Transition Oscillometry; SBP, systolic blood pressure; DBP, diastolic blood pressure; MAP, mean arterial pressure; CI, confidence interval; SD, standard deviation.

### Supplementary Figures

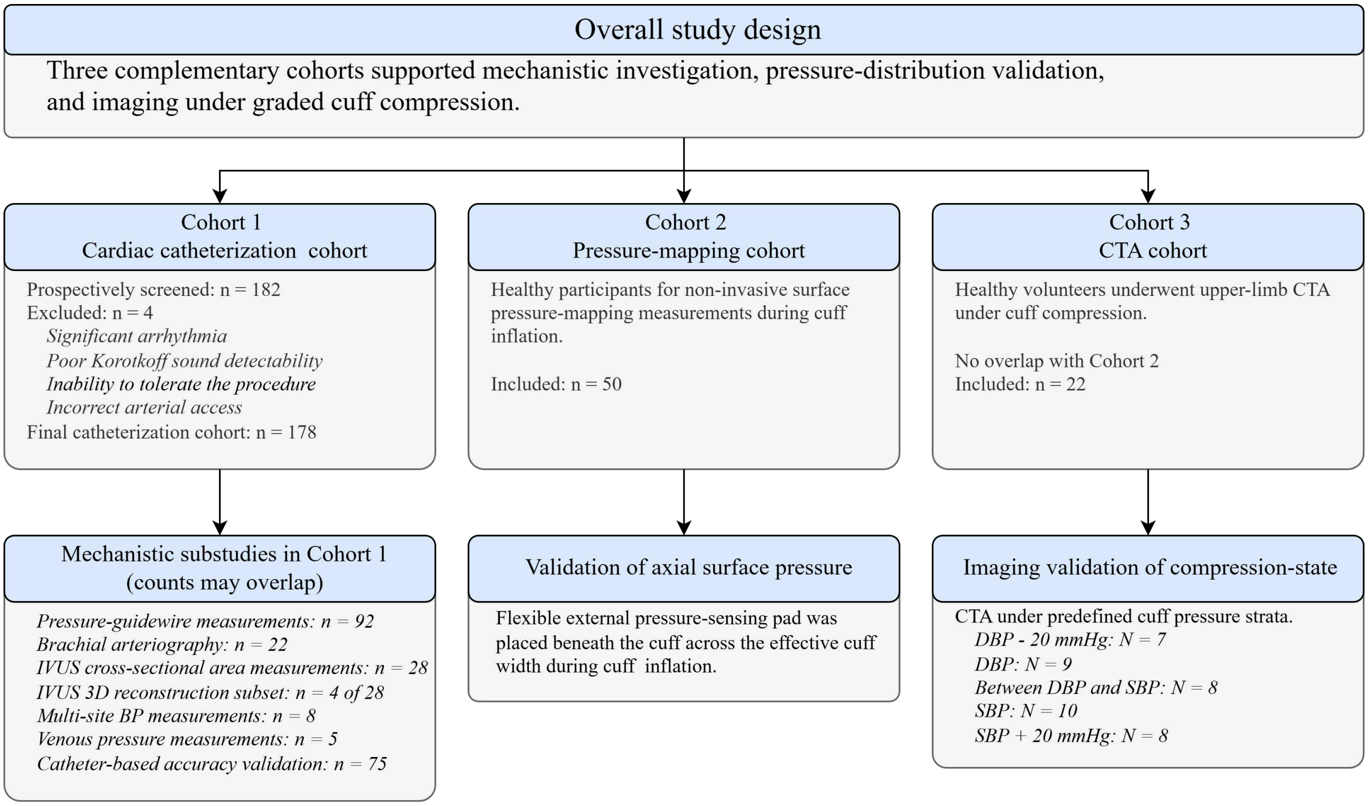

**Supplementary Fig. 1. Overall study design and cohort structure.**
Three complementary cohorts were included for invasive mechanistic measurements, validation of cuff-induced surface pressure distribution, and upper-limb CTA under graded cuff compression. Counts for Cohort 1 substudies are not mutually exclusive and should not be summed to obtain the number of unique participants. Lowercase n denotes the number of participants, whereas uppercase N denotes the number of measurements. Abbreviations: BP, blood pressure; CTA, computed tomographic angiography; DBP, diastolic blood pressure; IVUS, intravascular ultrasound; MAP, mean arterial pressure; SBP, systolic blood pressure.

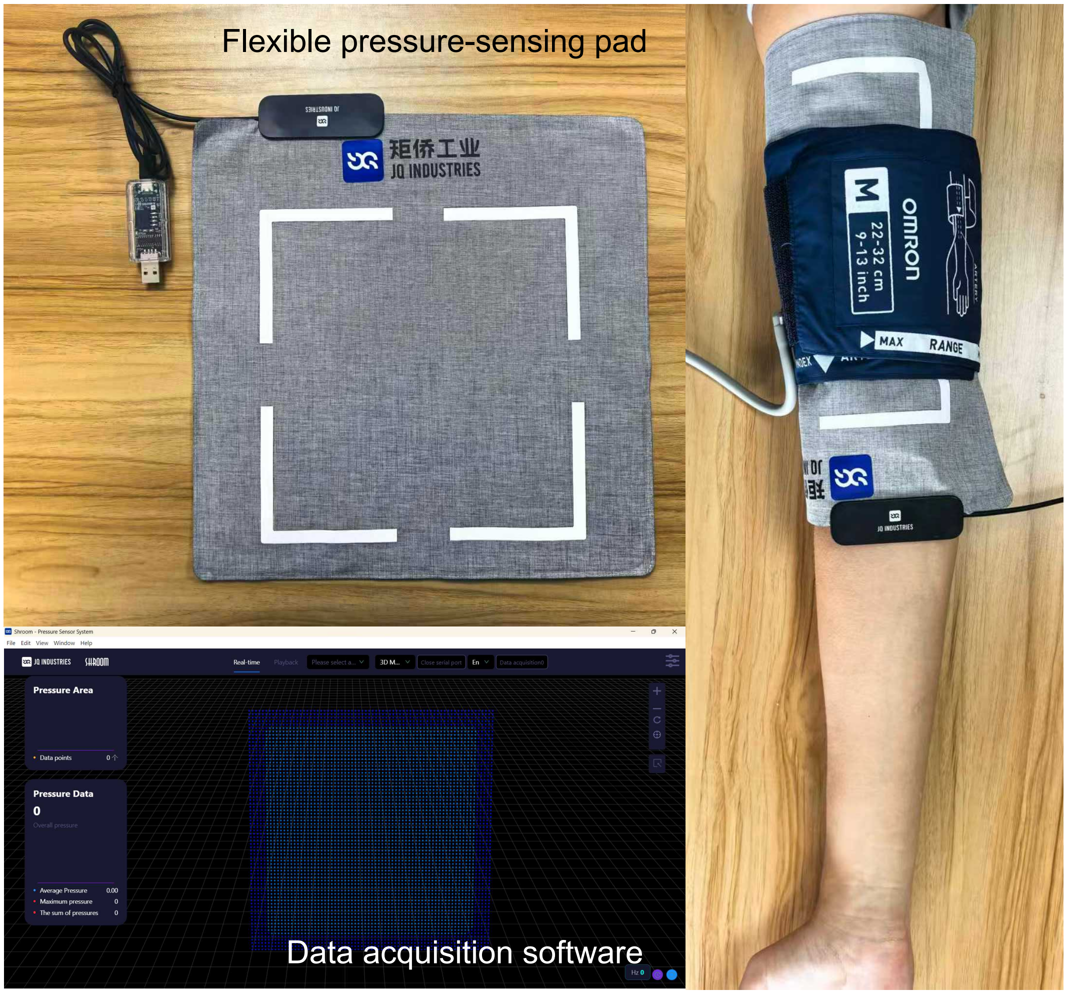

**Supplementary Fig. 2. External pressure-mapping system used for measurement of cuff-transmitted surface pressure.**

The contact-pressure sensing unit consisted of a flexible pressure-sensing pad (Mini pressure-sensing pad; JQ Industries, Weihai JQ Industrial Technology Co., Ltd., Weihai, China) with 1,024 sensing points arranged in a 32 × 32 matrix over a 200 × 200 mm active area. In the noninvasive pressure-mapping cohort, the pad was positioned between the upper-arm cuff and the skin during controlled cuff inflation to record spatiotemporal surface-pressure fields at 100 Hz. Framewise pressure maps were reconstructed from the exported sensor arrays, and row-wise temporal pressure profiles were obtained by averaging across the column dimension; the 3-point row-smoothed profile was used for visualization in the main text.

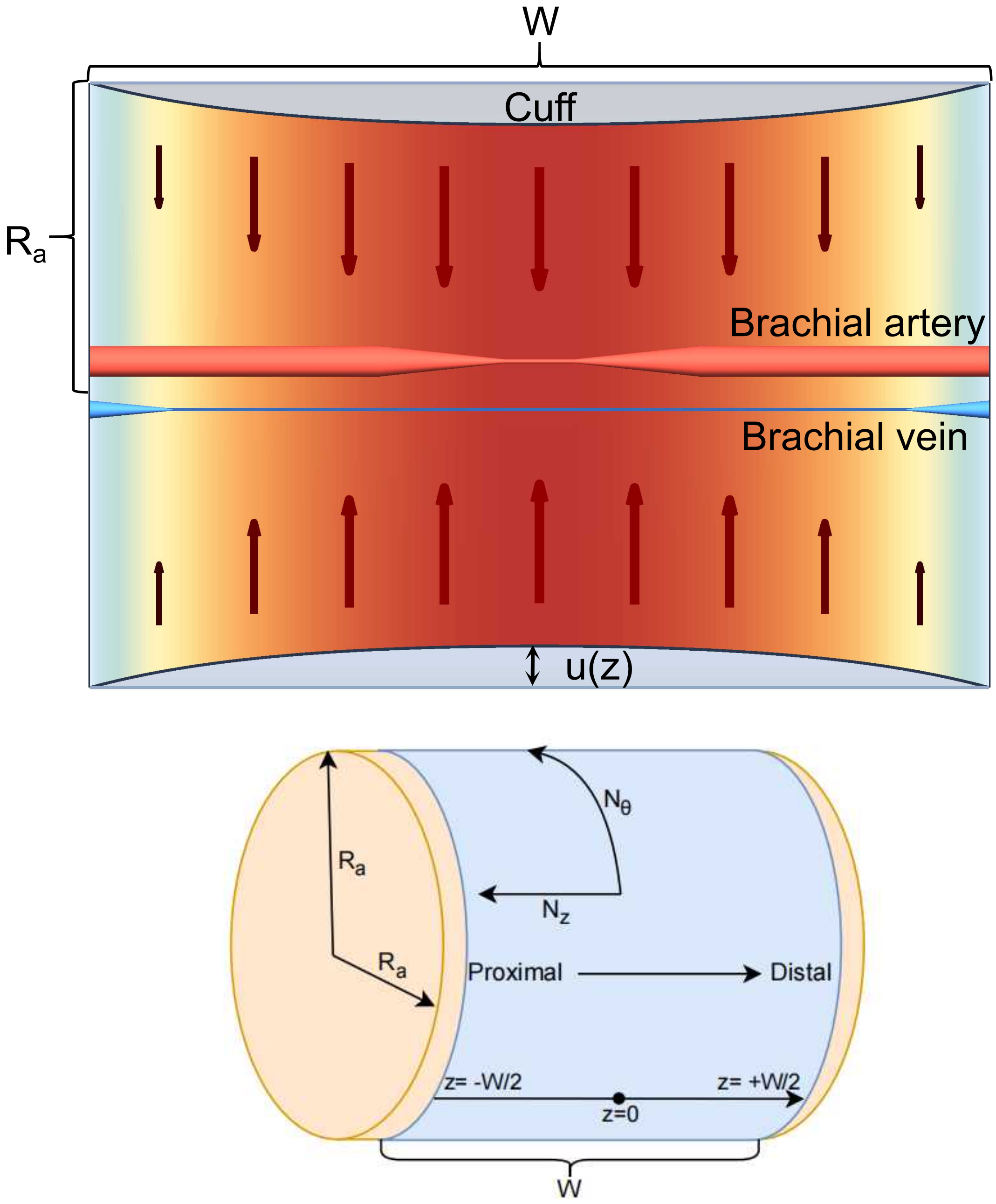

**Supplementary Fig. 3. Idealized computational schematic of axial pressure distribution beneath the cuff and compression states of the artery and vein.**

The schematic summarizes the conceptual framework used to derive the normalized axial contact-pressure distribution beneath the cuff. The upper panel shows the cuff-loaded region as a thin-membrane system transmitting external pressure to soft tissue and to the underlying brachial vessels, producing a center-enhanced axial loading pattern. The lower panel defines the principal geometric and mechanical variables used in the derivation, including the effective cuff width $W$, equivalent upper-arm radius $R_a$, local indentation displacement $u(z)$, and the circumferential and axial membrane-force resultants $N_\theta$ and $N_z$. The artery and vein are shown qualitatively to illustrate how stronger central loading and weaker edge loading are expected to generate different local compression states along the cuff axis. This nonuniform loading pattern forms the external-pressure input for the subsequent cuff–brachial artery coupling model.

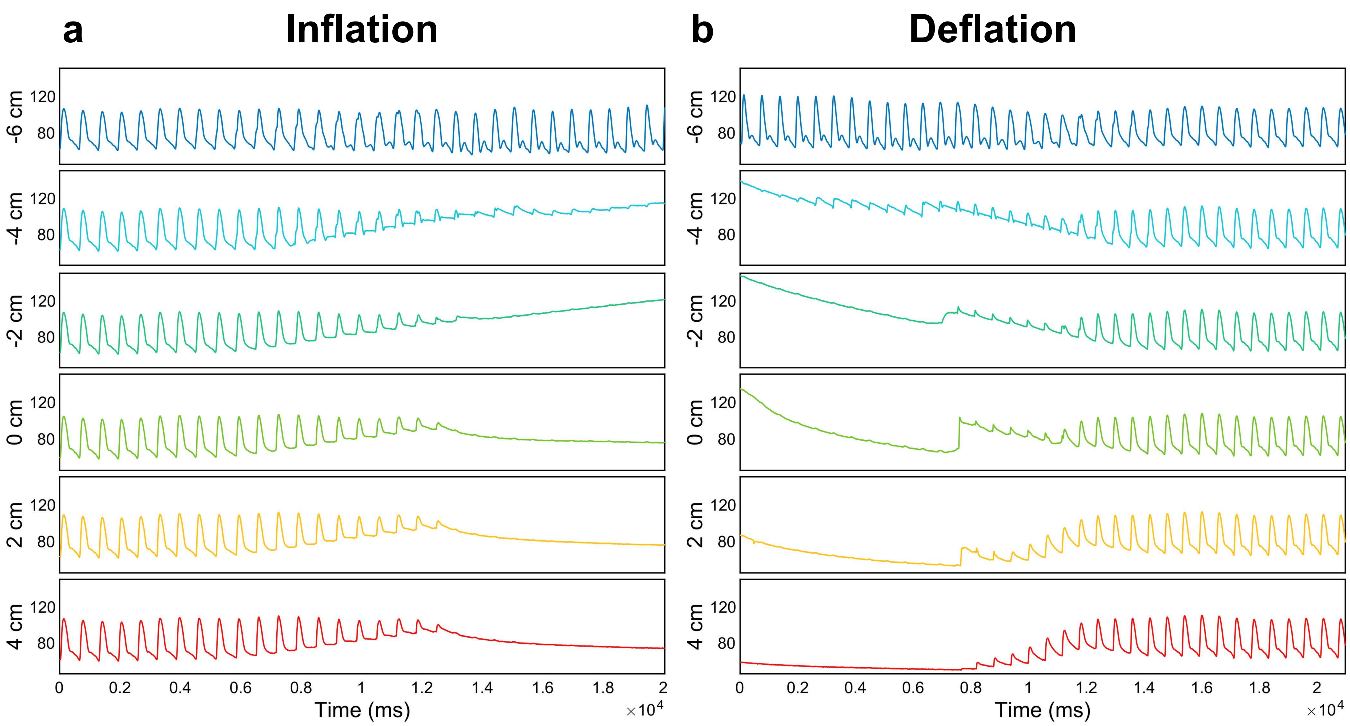

**Supplementary Fig. 4. Complete intra-arterial pressure recordings from predefined sub-cuff sites spanning the cuff-covered segment (participant 1).**

**a–b**, Intra-arterial pressure waveforms recorded simultaneously from predefined measurement sites distributed across the full cuff-covered segment. Cuff-inflation and deflation measurements are shown, providing complete axial pressure profiles during cuff-based BP measurement in this participant. Vertical axis unit: mmHg.

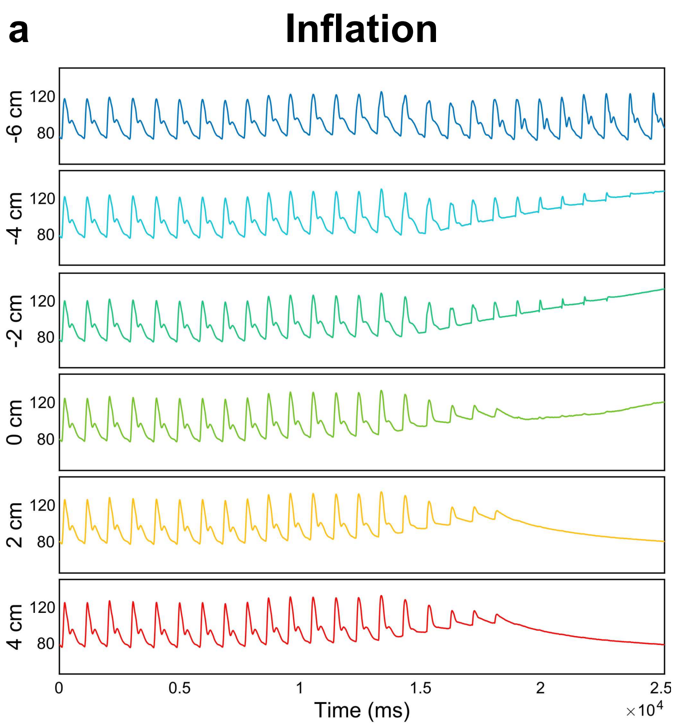

**Supplementary Fig. 5. Complete intra-arterial pressure recordings from predefined sub-cuff sites spanning the cuff-covered segment (participant 2).**

**a–b**, Intra-arterial pressure waveforms recorded simultaneously from predefined measurement sites distributed across the full cuff-covered segment. Cuff-inflation and deflation measurements are shown, providing complete axial pressure profiles during cuff-based BP measurement in this participant. Vertical axis unit: mmHg.

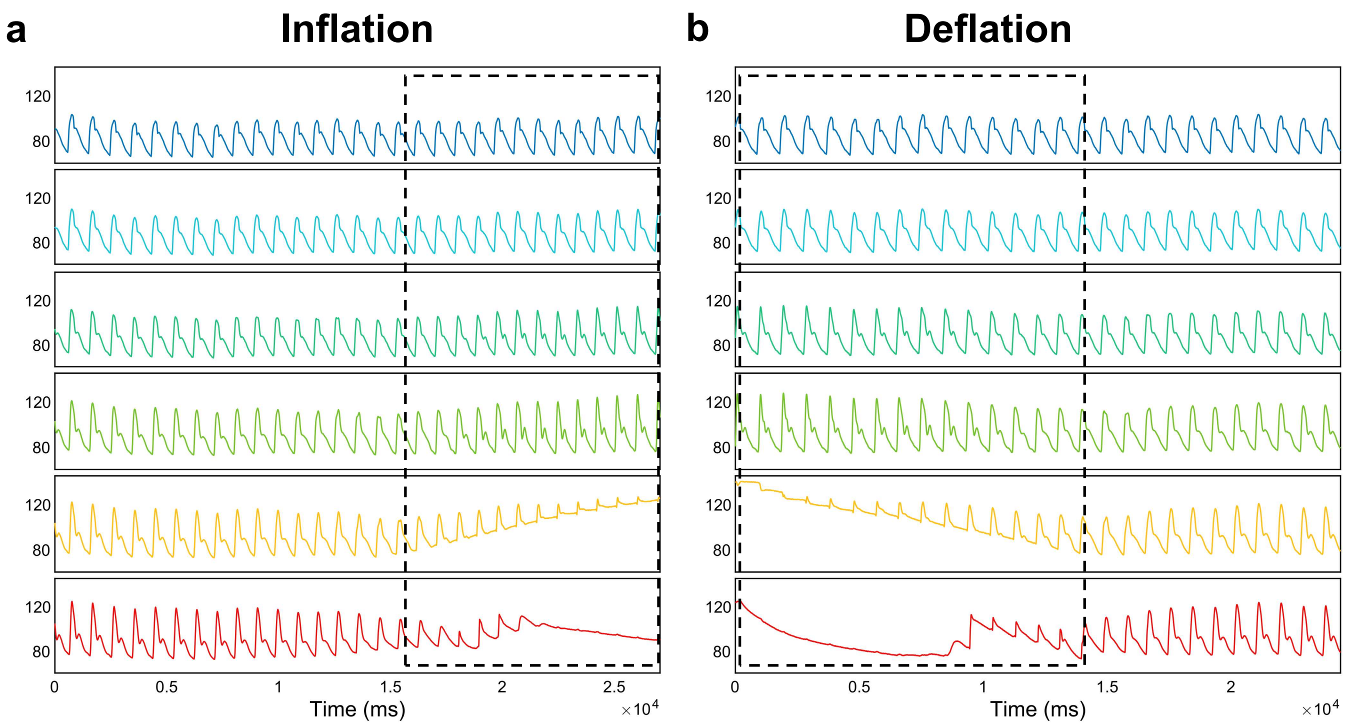

**Supplementary Fig. 6. Segmental intra-arterial pressure recordings from the ascending aorta to the sub-cuff brachial artery.**

**a–b**, Measurement sites were positioned sequentially at the ascending aorta, the brachiocephalic origin, the right subclavian artery, the right axillary artery, a proximal sub-cuff brachial site, and a central sub-cuff brachial site (from top to bottom). The cuff inflation and deflation measurements are shown, illustrating segmental pressure changes from the central arteries to the cuff-compressed brachial segment. Vertical axis unit: mmHg.

**a**

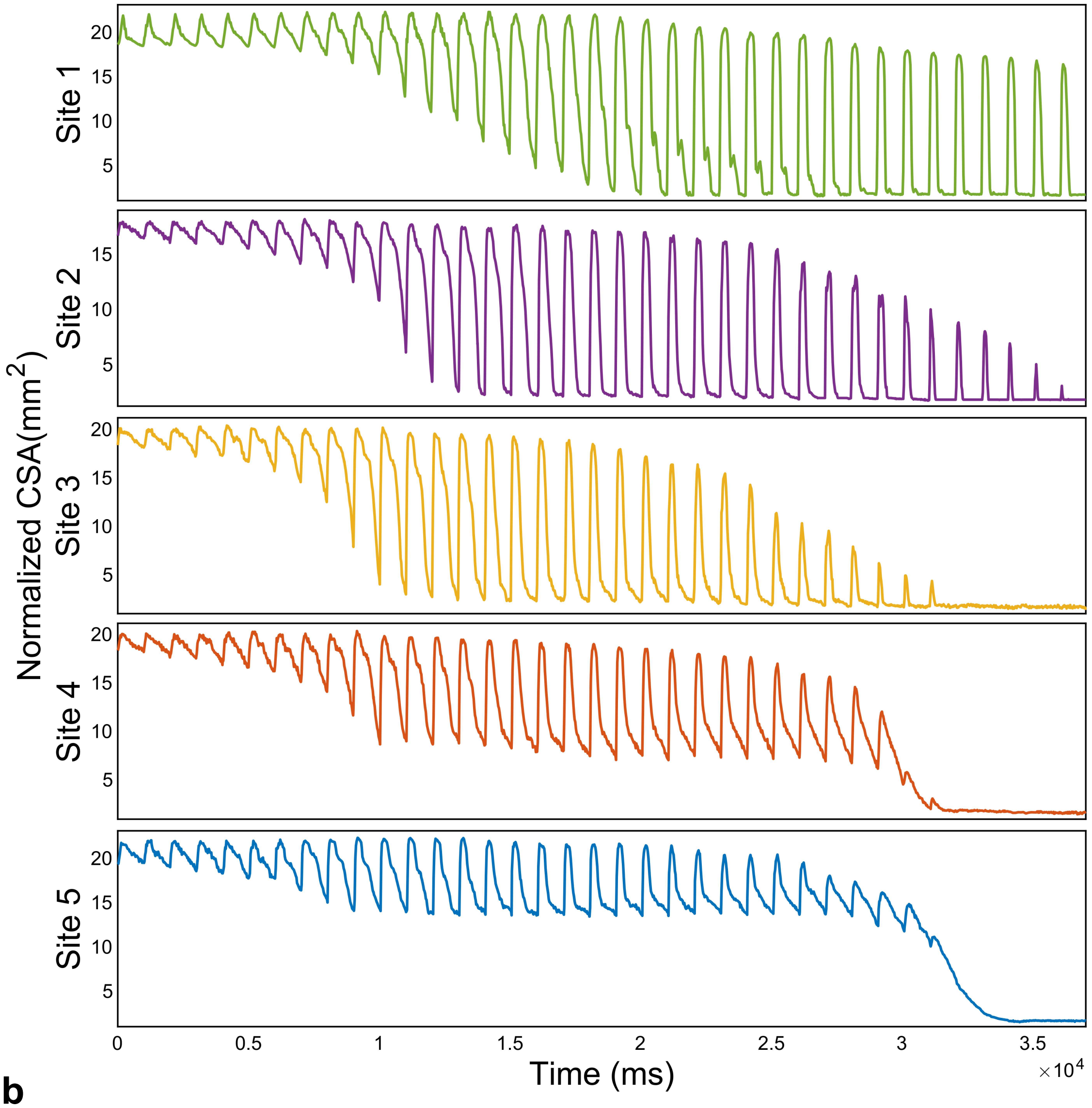

**b**

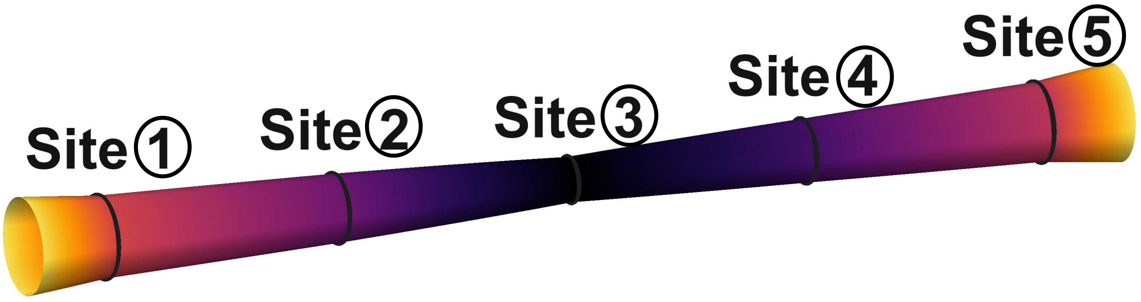

**Supplementary Fig. 7. Time-normalized CSA waveforms and volume reconstruction using 5-site linear interpolation.**

**a**, Time-normalized CSA waveforms from five IVUS measurement sites, with each cardiac cycle normalized to the same duration. The top panel shows CSA time series for Sites ① through ⑤.

**b**, Volume reconstruction based on linear interpolation of the CSA data from the five sites, used to estimate the volume change of the cuff-covered arterial segment.

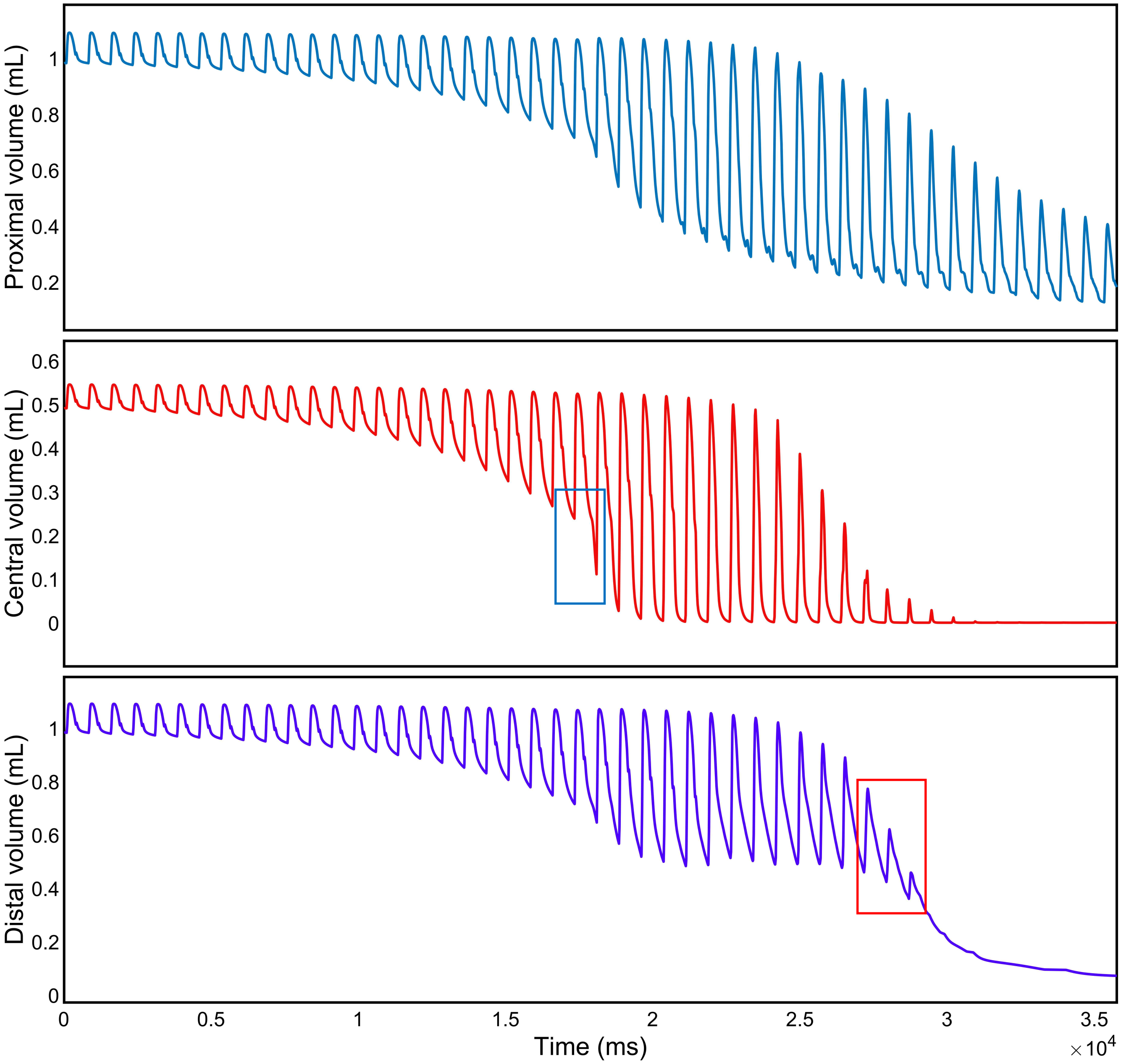

**Supplementary Fig. 8. Model-predicted volume changes in the proximal segment, central PPZ, and distal segment of the sub-cuff brachial artery.**

The top, middle, and bottom panels show the model-predicted volume changes over time in the proximal segment, central PPZ, and distal segment of the brachial artery beneath the cuff, respectively. The three traces exhibit distinct temporal patterns, indicating marked axial heterogeneity in the volumetric response of the artery under cuff compression. The blue box in the middle panel marks the abrupt decrease in volume in the central PPZ when cuff pressure reaches DBP, indicating rapid vessel collapse in this region. The red box in the bottom panel marks the disappearance of volume pulsations in the distal segment when cuff pressure reaches SBP, indicating the loss of distal arterial pulsation.

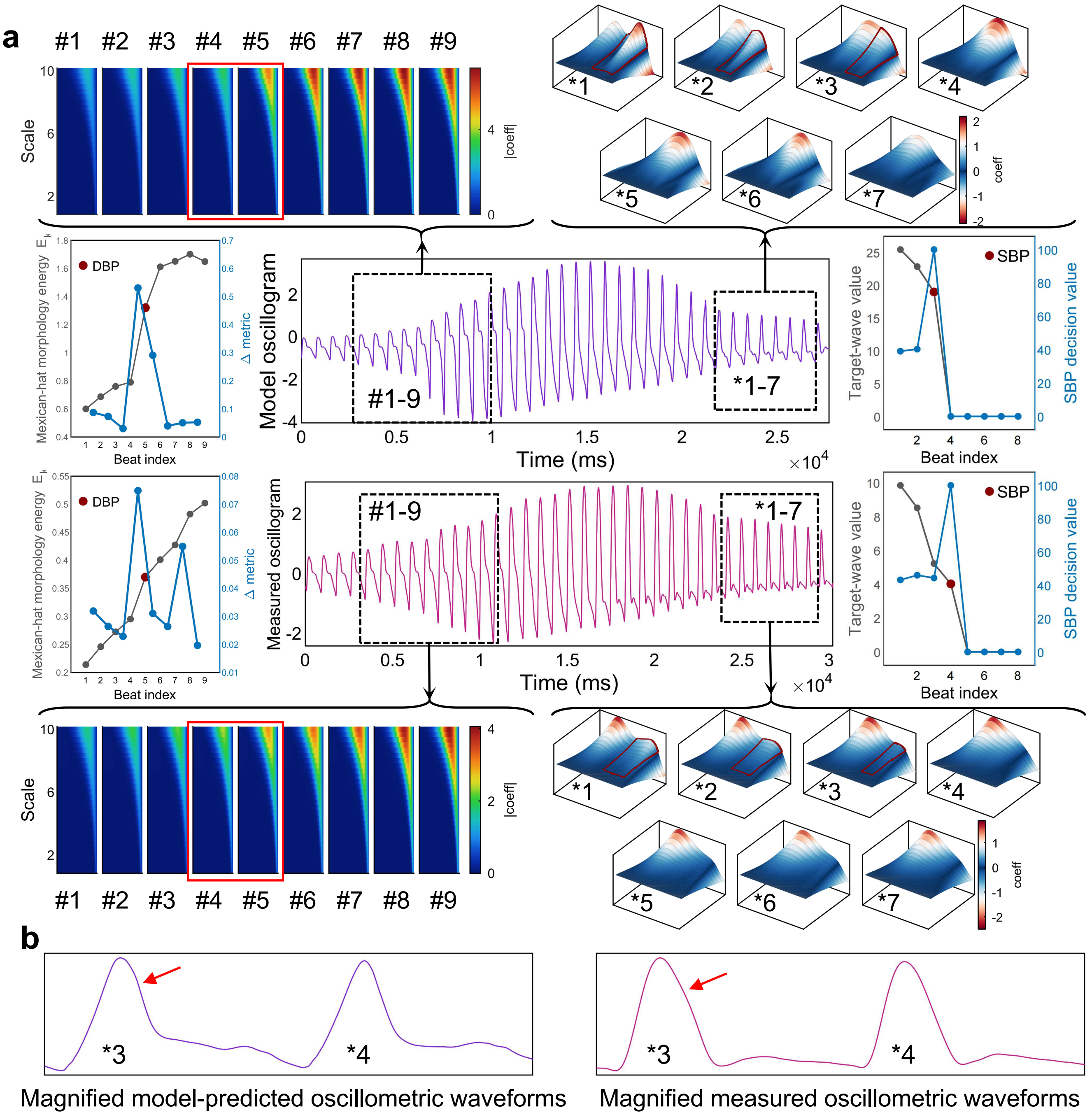

**Supplementary Fig. 9. Identification of SBP- and DBP-related oscillogram features in the model-predicted and measured oscillograms from the same subject in Fig. 3.**

**a**, The upper row shows the model-predicted oscillogram generated using the same subject-specific BP input, and the lower row shows the corresponding measured oscillogram. Mexican-hat wavelet analysis was applied to both signals to identify the characteristic feature-transition points associated with DBP and SBP. Insets and dashed boxes highlight representative local waveform segments used for feature extraction. The left summary panels show the beat-wise DBP-related morphology metric, and the right summary panels show the SBP-related target-wave metric; red markers indicate the identified DBP and SBP points, respectively. The corresponding wavelet coefficient maps and three-dimensional coefficient surfaces illustrate the evolution of local waveform morphology around the selected beats.

**b**, The left and right panels show locally magnified model-predicted and measured oscillometric waveforms, respectively, representing two adjacent cardiac cycles as cuff pressure crossed SBP. Red arrows indicate the distal small-wave-related component.

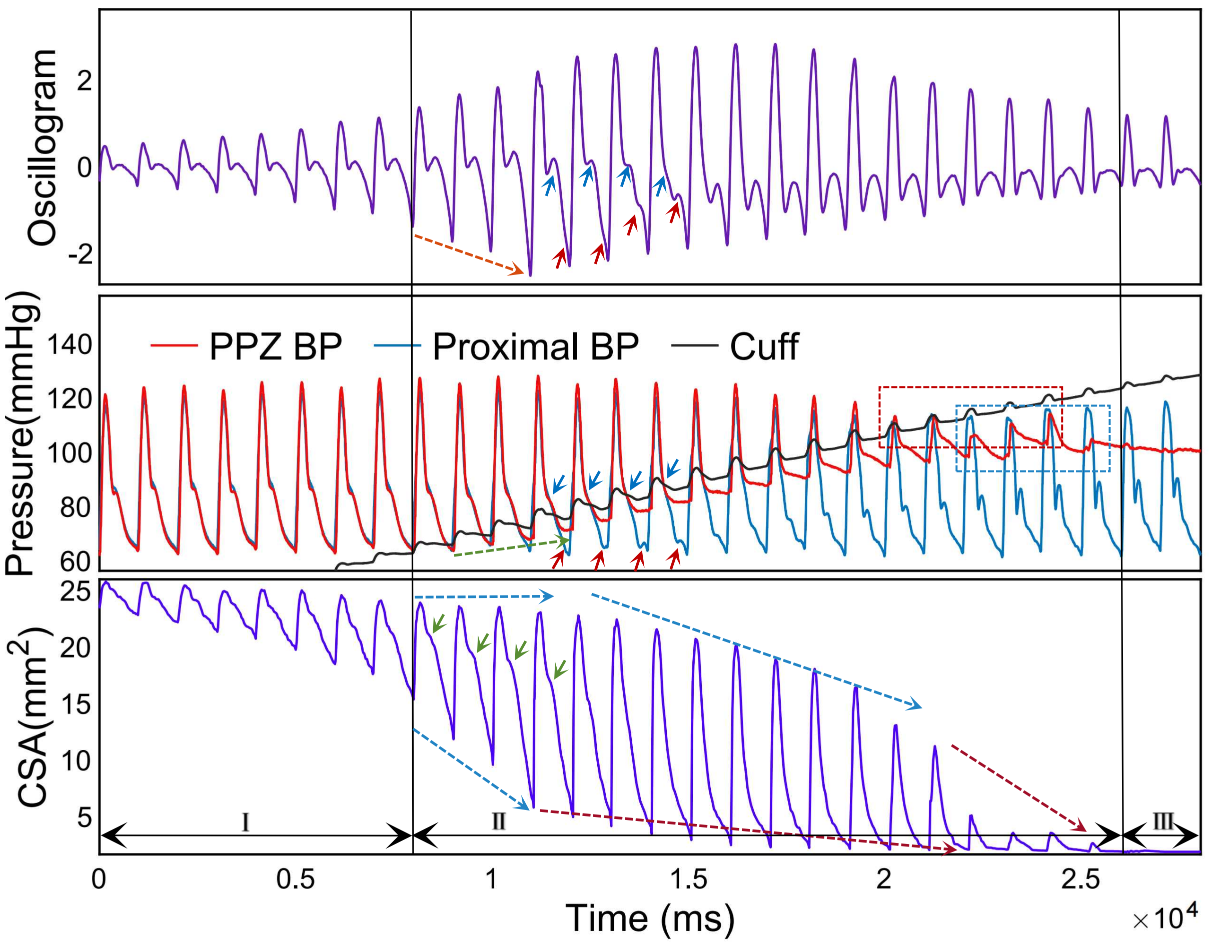

**Supplementary Fig. 10. Additional oscillogram features revealed by co-registered oscillogram, intra-arterial pressure, and IVUS recordings during cuff inflation.**

Co-registered recordings obtained during cuff inflation. The top panel shows the oscillogram extracted from cuff pressure. The middle panel shows cuff pressure together with intra-arterial pressures recorded at Site 0 and Site ③. The bottom panel shows IVUS-derived cross-sectional area measured at Site ③. Roman numerals denote sequential waveform phases. Colored arrows and boxes annotate phase-specific transitions, including emergence of reflection-like components, attenuation and disappearance of the dicrotic notch, formation of proximal double peaks, appearance of residual distal pressure small waves, and last IVUS pulsatility in phase II.

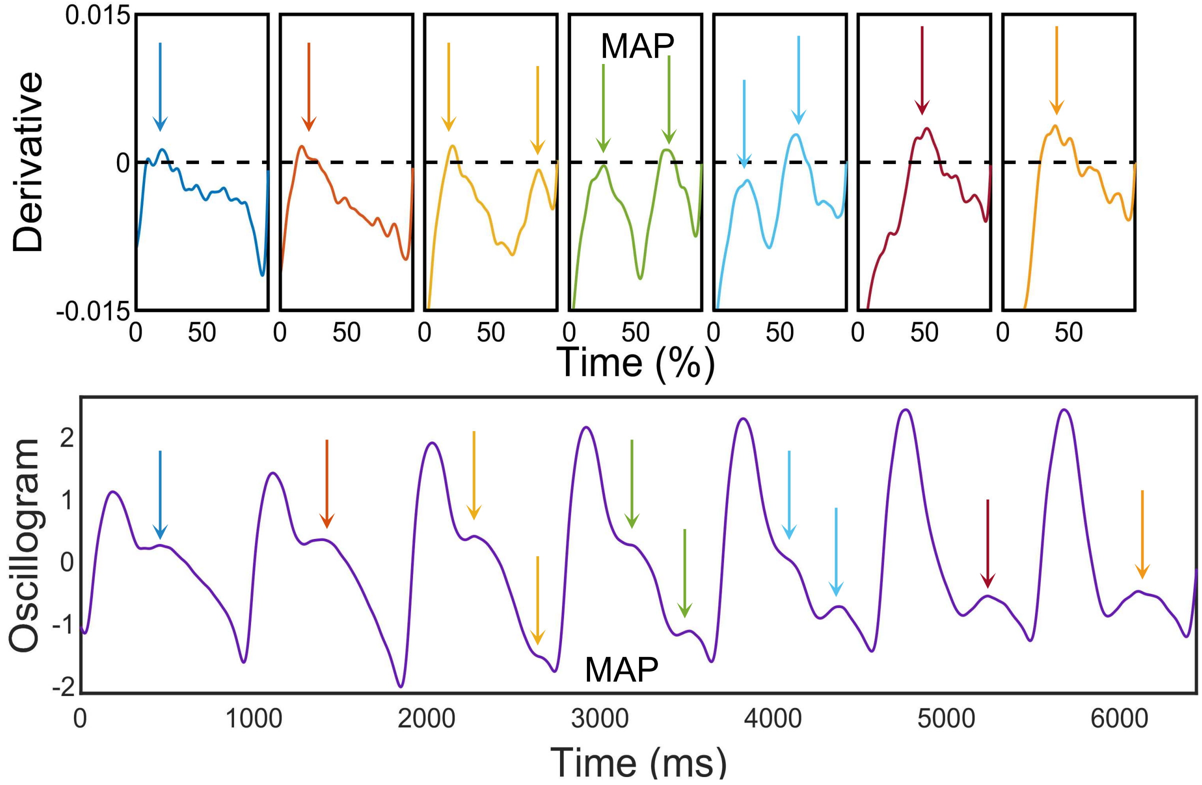

**Supplementary Fig. 11. Notch recognition and MAP positioning.**
First-derivative–based localization of the dicrotic-like notch and reflection-like notch, used for MAP identification in the oscillogram.

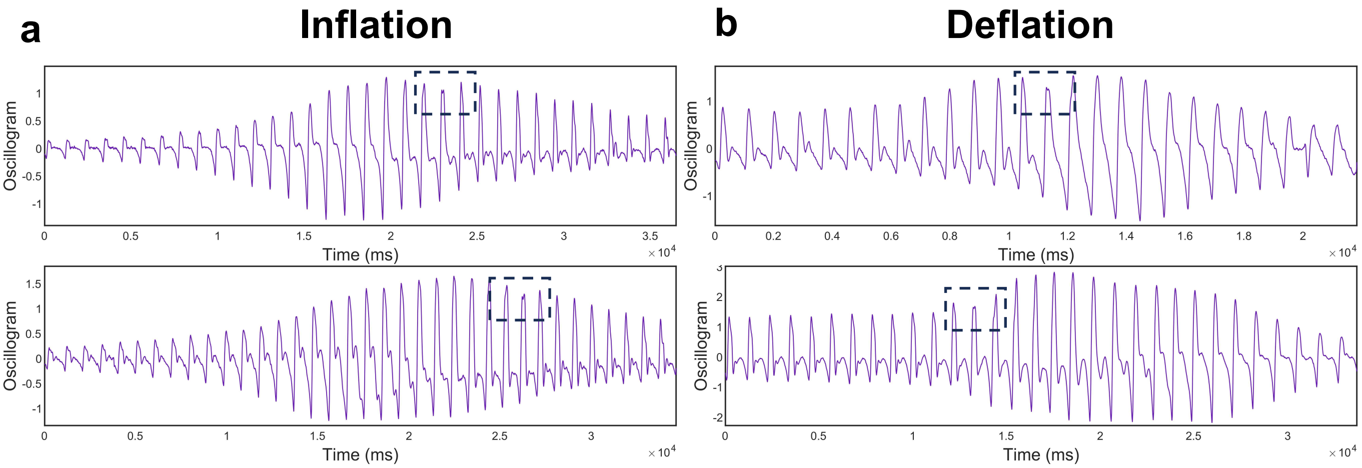

**Supplementary Fig. 12. Additional examples of oscillogram features associated with residual distal pressure small waves in different subjects (double-peak transitions).**

**a–b**, Oscillograms from different participants illustrating double-peak transitions associated with residual distal pressure small waves. The left and right columns show inflation and deflation recordings, respectively.

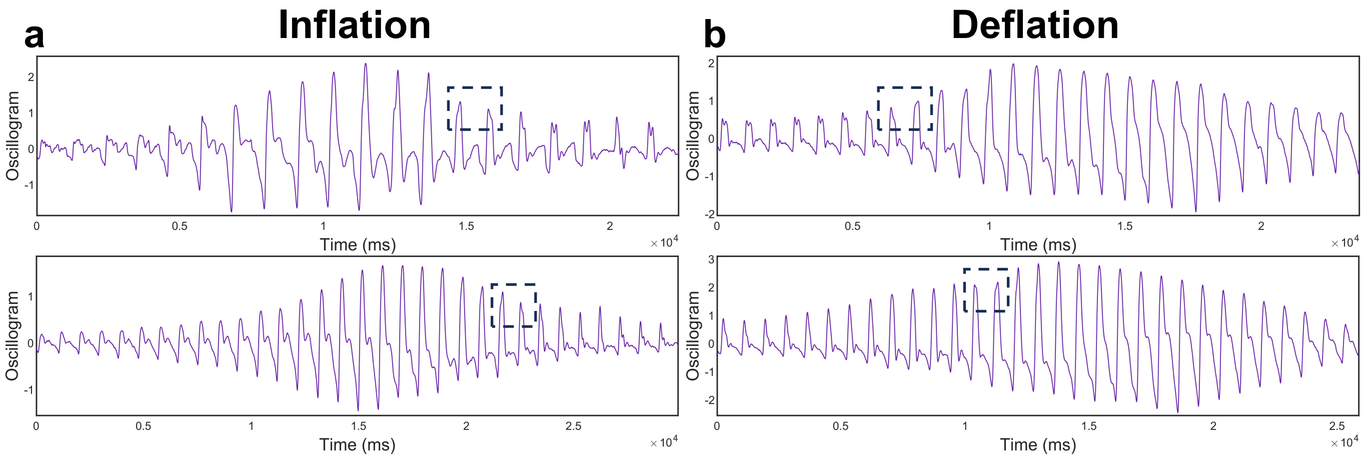

**Supplementary Fig. 13. Additional examples of oscillogram features associated with residual distal pressure small waves in different subjects (peak-side shift).**

**a–b**, Oscillograms from different participants illustrating peak-side shift associated with residual distal pressure small waves. The left and right columns show inflation and deflation recordings, respectively.

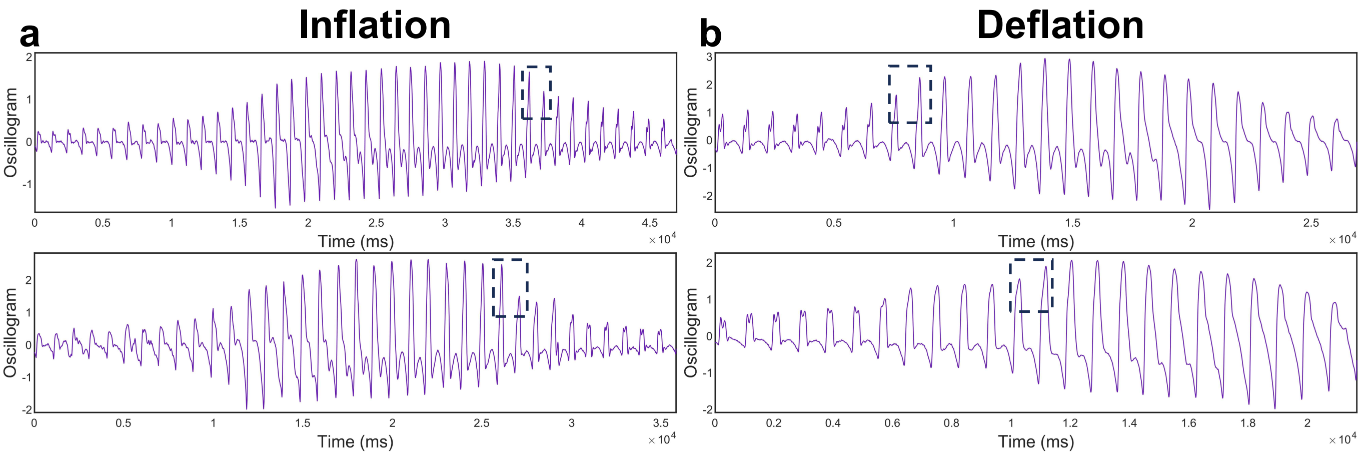

**Supplementary Fig. 14. Additional examples of oscillogram features associated with residual distal pressure small waves in different subjects (amplitude transitions).**

**a–b**, Oscillograms from different participants illustrating amplitude transitions associated with residual distal pressure small waves. The left and right columns show inflation and deflation recordings, respectively.

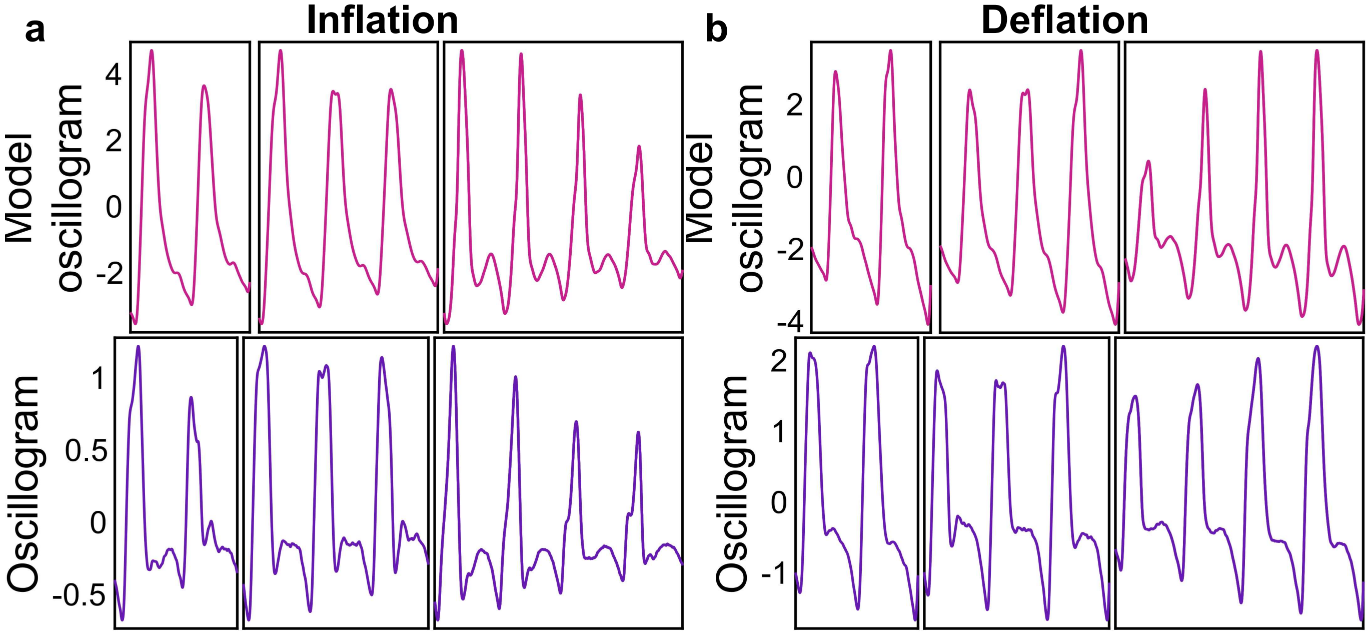

**Supplementary Fig. 15. Model prediction and measured observations of oscillogram features associated with the emergence of small waves.**

**a**–**b**, Local magnified oscillogram segments showing model-predicted and measured features associated with the emergence of small waves during cuff inflation and deflation, respectively. In each panel, the upper row shows model-predicted oscillograms, and the lower row shows measured oscillograms. From left to right, the representative feature patterns are peak-side shift, double-peak transition, and amplitude transition.

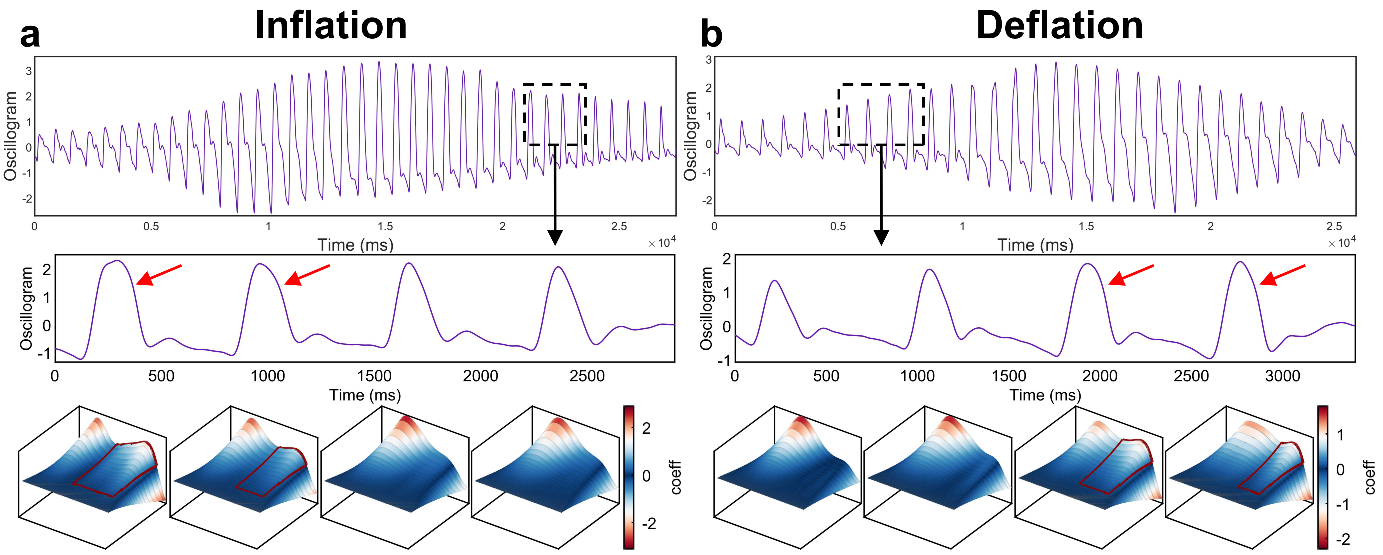

**Supplementary Fig. 16. Additional examples of SBP–related oscillogram feature changes in different subjects (whole-peak narrowing/broadening of the main peak).**

**a–b**, Representative oscillograms from different subjects illustrating whole-peak narrowing/broadening associated with SBP identification. The top panels show complete oscillograms; the middle panels show magnified four consecutive single-cycle oscillometric waveforms around the transition, with red arrows marking the small-wave components; and the bottom panels show the corresponding Mexican-hat wavelet scalograms. The left and right columns represent cuff inflation and deflation, respectively.

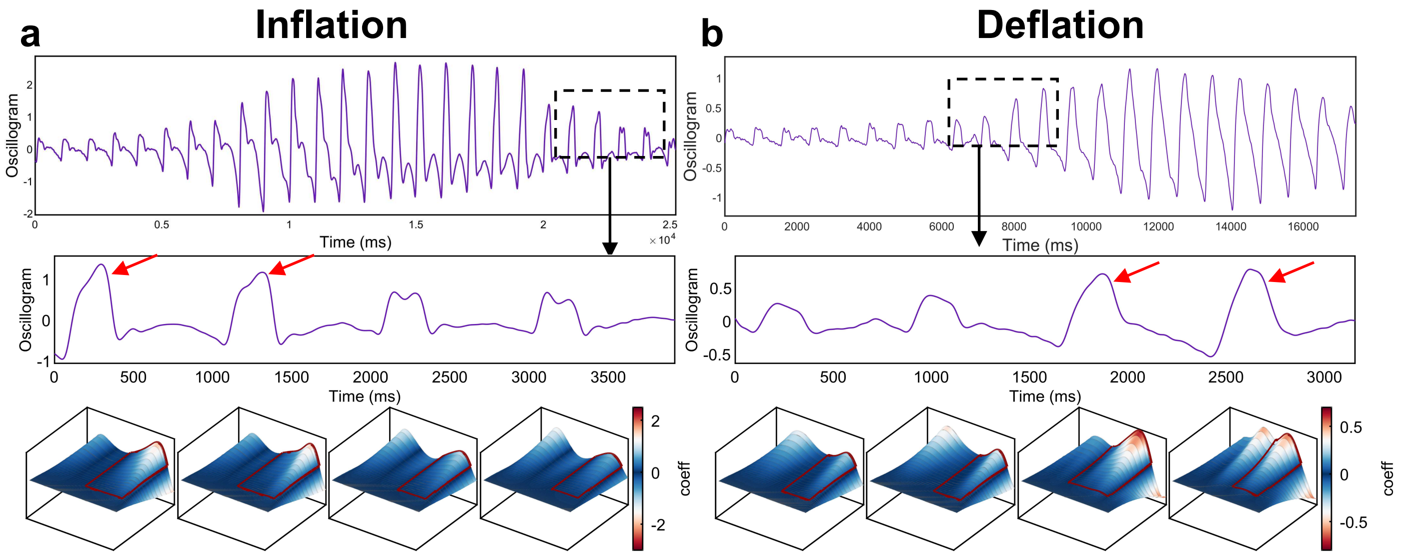

**Supplementary Fig. 17. Additional examples of SBP–related oscillogram feature changes in different subjects (amplitude changes).**

**a–b**, Representative oscillograms from different subjects illustrating amplitude changes associated with SBP identification. The top panels show complete oscillograms; the middle panels show magnified four consecutive single-cycle oscillometric waveforms around small-wave disappearance/appearance, with red arrows marking the small-wave components; and the bottom panels show the corresponding Mexican-hat wavelet scalograms. The left and right columns represent cuff inflation and deflation, respectively.

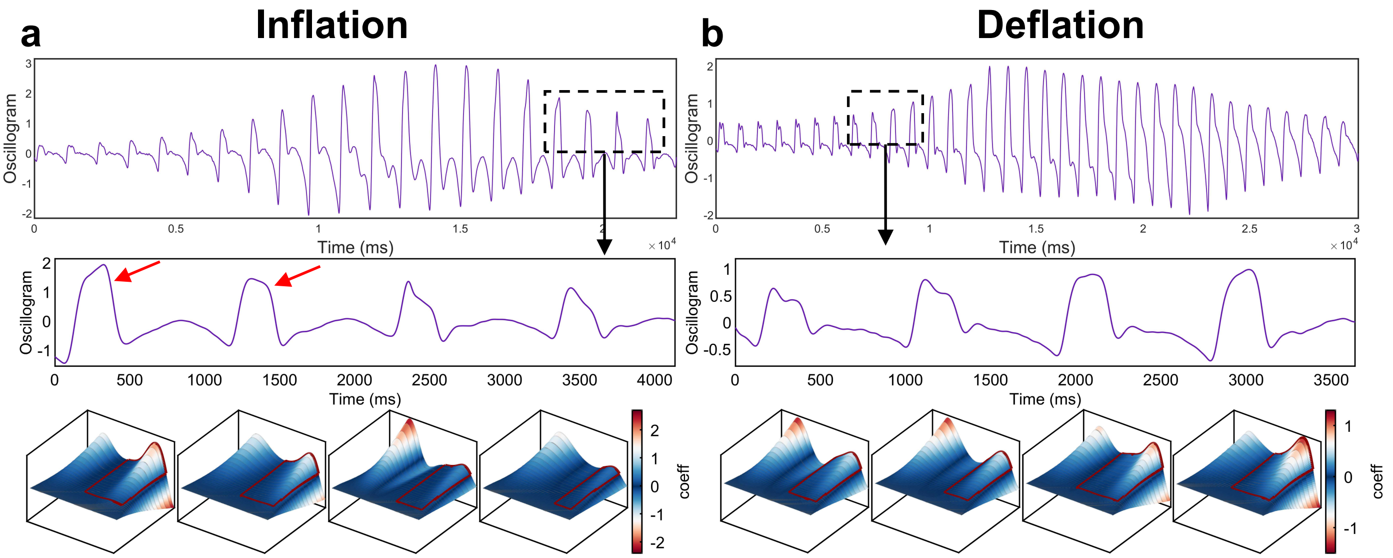

**Supplementary Fig. 18. Additional examples of SBP–related oscillogram feature changes in different subjects (appearance/disappearance of the descending-limb notch).**

**a–b**, Representative oscillograms from different subjects illustrating descending-limb notch appearance/disappearance associated with SBP identification. The top panels show complete oscillograms; the middle panels show magnified four consecutive single-cycle oscillometric waveforms around the transition, with red arrows marking the small-wave components; and the bottom panels show the corresponding Mexican-hat wavelet scalograms. The left and right columns represent cuff inflation and deflation, respectively.

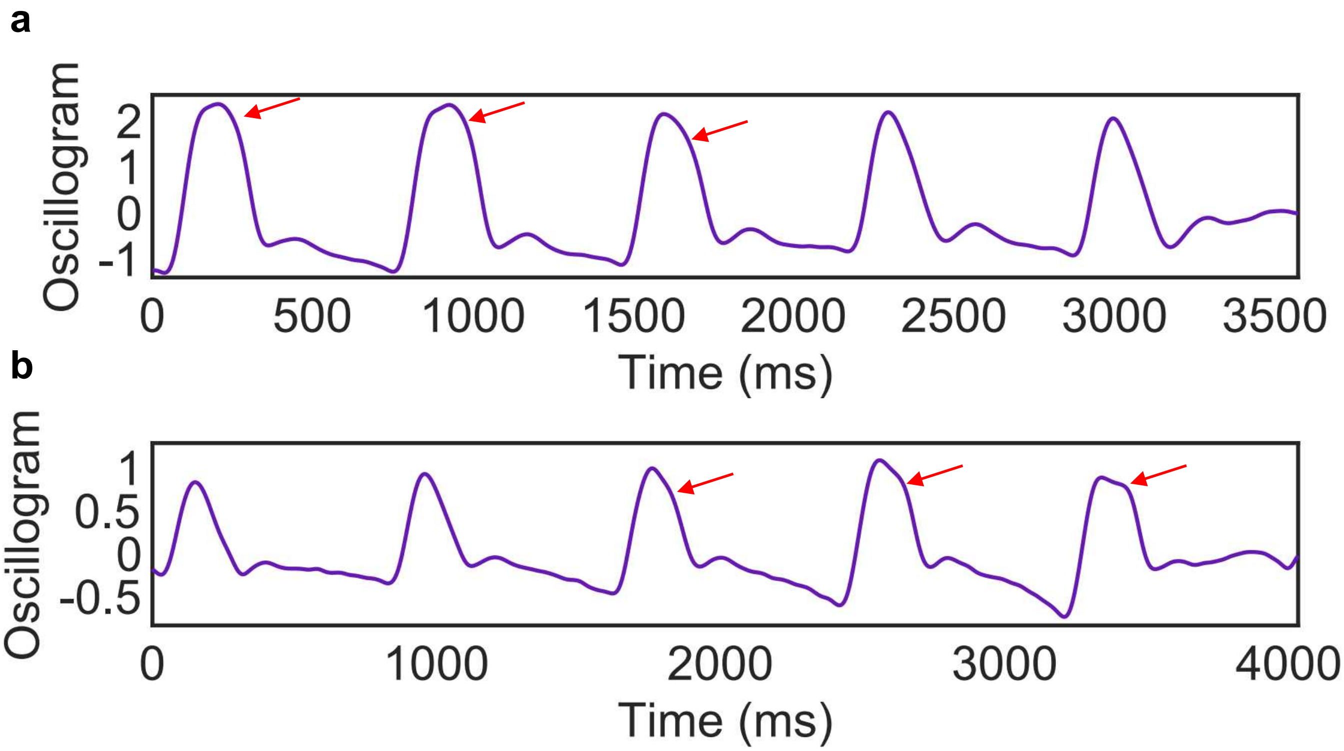

**Supplementary Fig. 19. Measured observations of oscillogram features associated with small-wave disappearance and reappearance.**

**a**–**b**, Locally magnified oscillogram segments during cuff inflation and deflation, respectively, showing the features associated with small-wave disappearance and reappearance, respectively. Red arrows indicate the small-wave-related component on the descending limb.

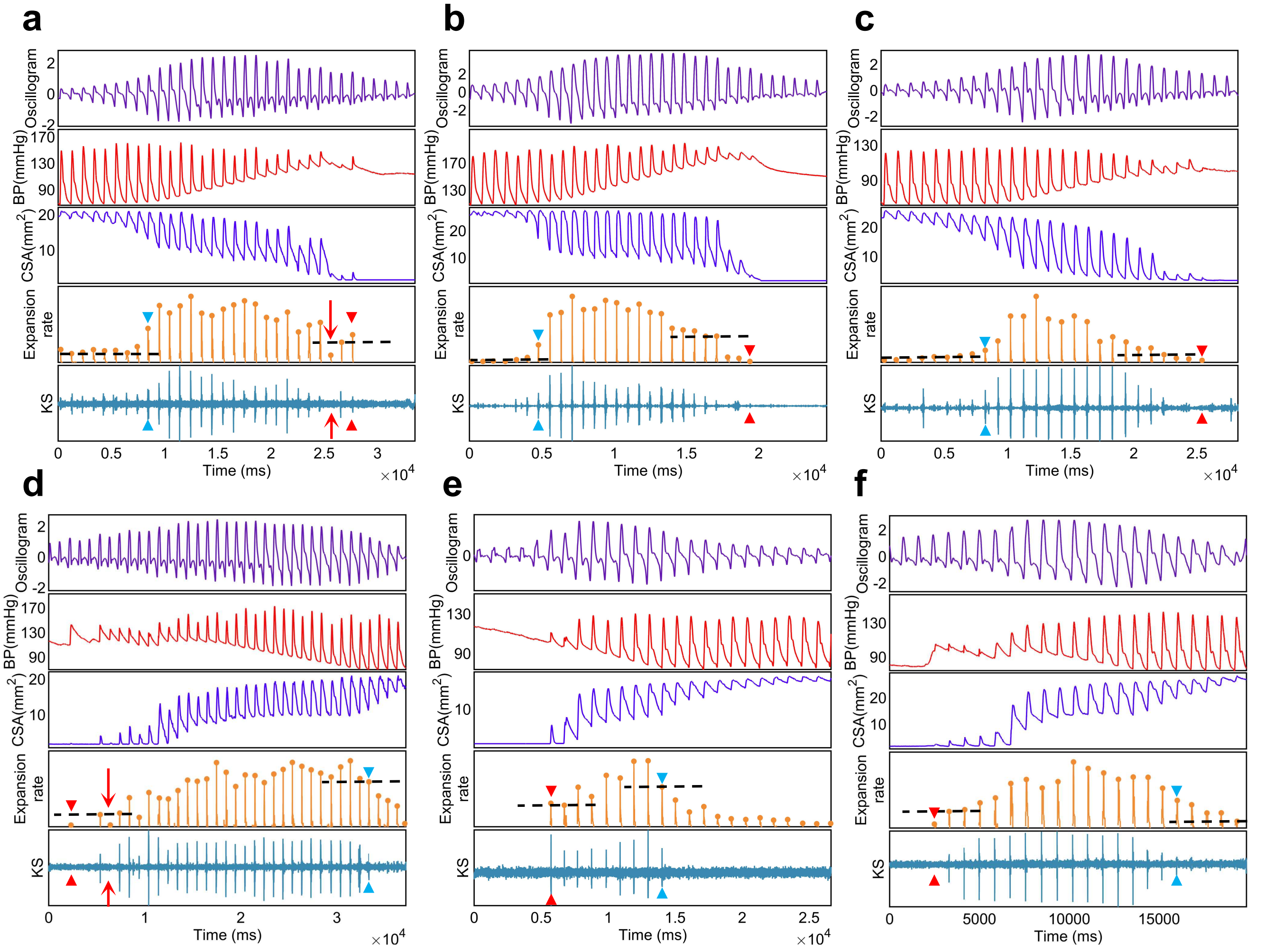

**Supplementary Fig. 20. Expansion-rate-based analysis of Korotkoff sound signals.**
Additional synchronized representative recordings of the oscillogram, Site ④ pressure, Site ④ cross-sectional area (CSA), Site ④ CSA expansion rate, and Korotkoff sound signals during cuff-based BP measurement are shown from top to bottom. Panels a to c show representative recordings during cuff inflation, and panels d to f show representative recordings during cuff deflation. Black dashed lines indicate the case-specific KS detection thresholds. Red arrows indicate auscultatory gaps. Blue and red triangles indicate invasive DBP and SBP, respectively.

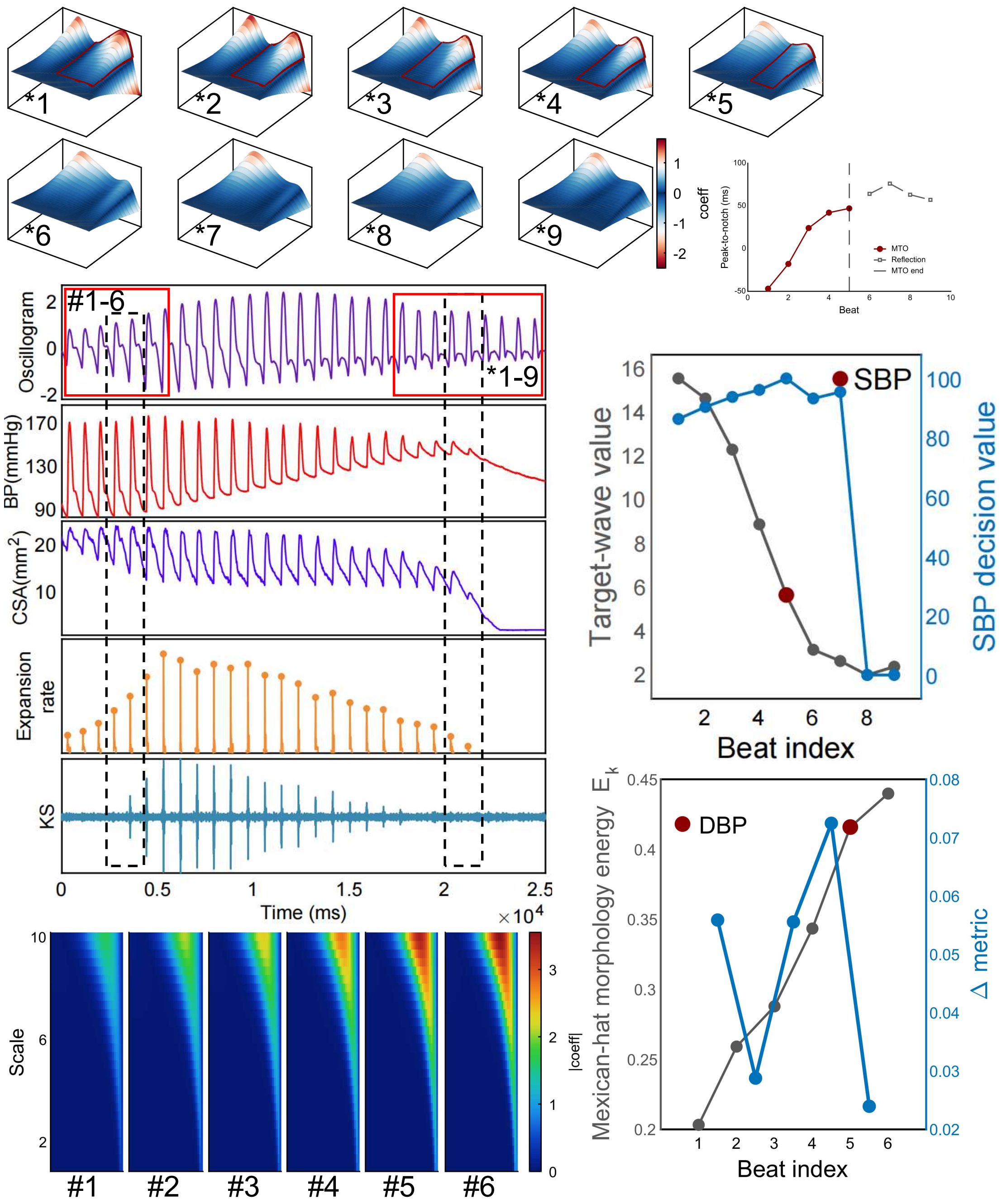

**Supplementary Fig. 21. Accuracy comparison between MTO and auscultation.**

The same measurement from the same patient as shown in Fig. 5d is used. The two red solid boxes indicate the waveform ranges used for DBP and SBP analysis, respectively. The black dashed boxes indicate the two adjacent cardiac cycles during which cuff pressure equaled DBP and SBP. Wavelet analysis results are shown in the heat maps and scatter plots. In this measurement, auscultation provided an accurate DBP estimate but an inaccurate SBP estimate. The top-right panel shows the peak-to-notch delay of the small-wave and reflection-related components, with the dashed line marking MTO termination.

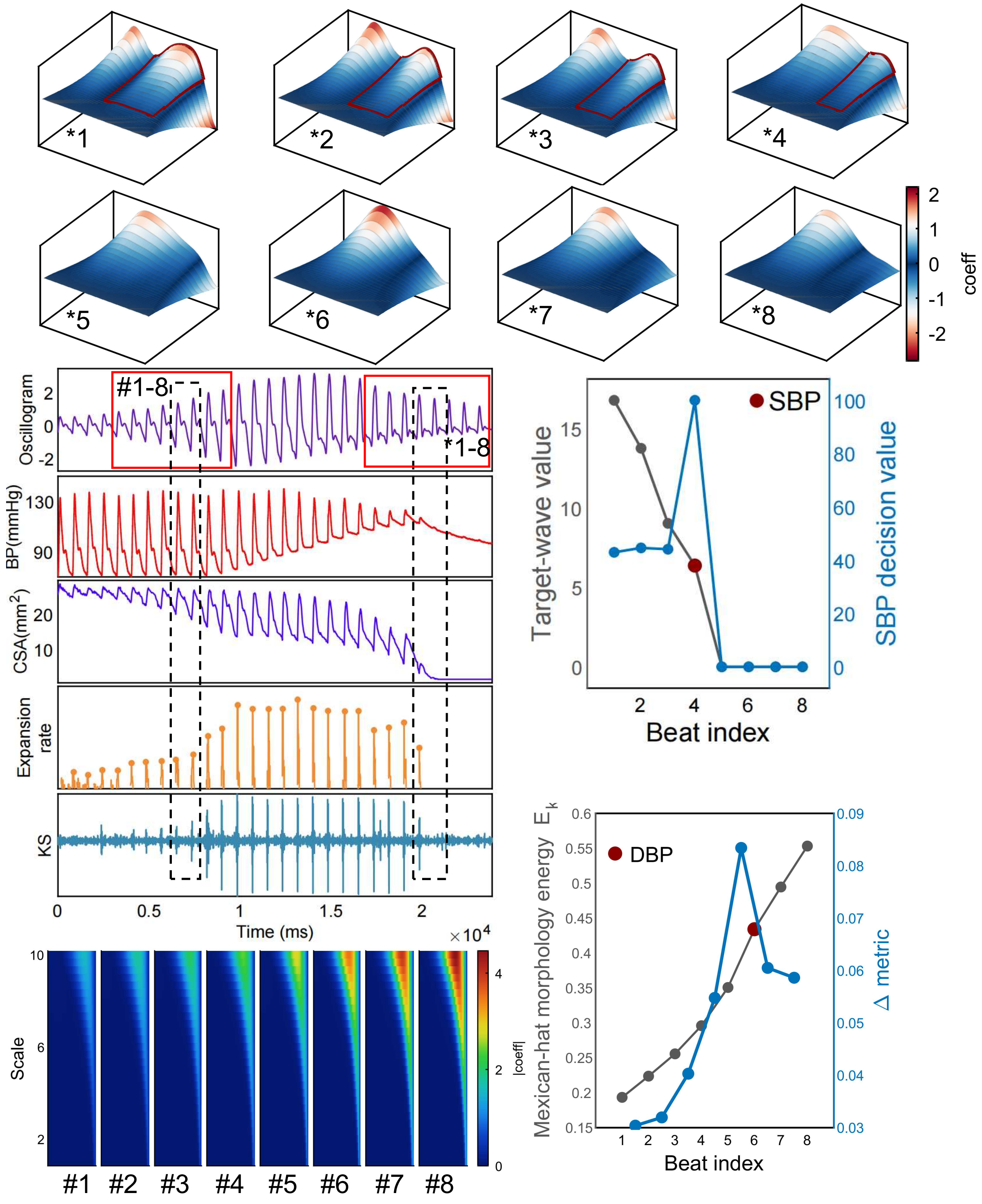

**Supplementary Fig. 22. Accuracy comparison between MTO and auscultation.**
The same measurement from the same patient as shown in Fig. 5e is used. The two red solid boxes indicate the waveform ranges used for DBP and SBP analysis, respectively. The black dashed boxes indicate the two adjacent cardiac cycles during which cuff pressure equaled DBP and SBP. Wavelet analysis results are shown in the heat maps and scatter plots. In this measurement, auscultation provided an accurate SBP estimate but an inaccurate DBP estimate.

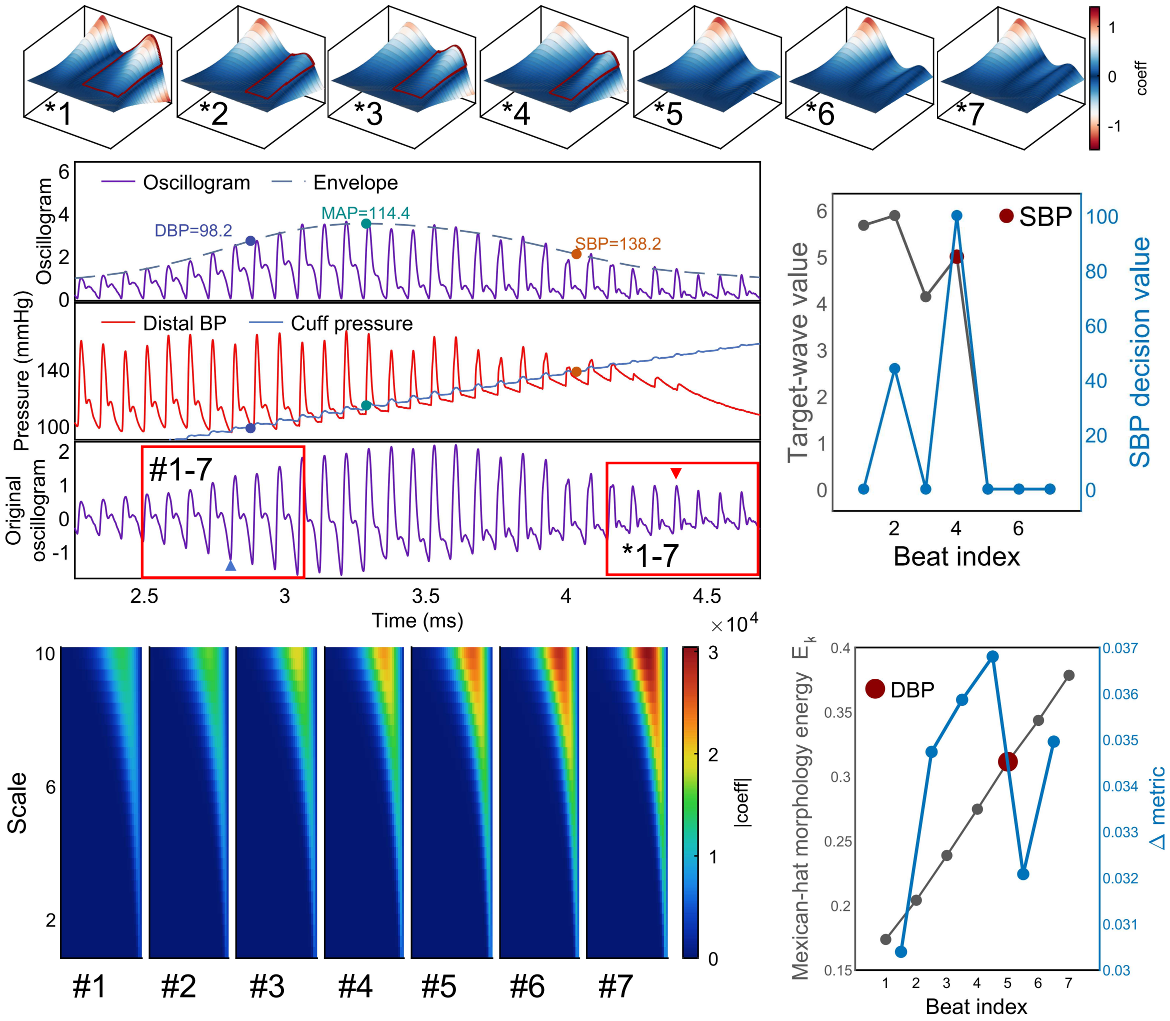

**Supplementary Fig. 23. Accuracy comparison between MTO and MAA.**

The same measurement from the same patient as shown in Fig. 5f is used. The MAA method was applied to the detrended oscillogram, whereas MTO analyzed the original oscillogram. The two red solid boxes indicate the waveform ranges used for DBP and SBP analysis, respectively. Wavelet analysis results are shown in the heat maps and scatter plots. In this measurement, MAA underestimated SBP.

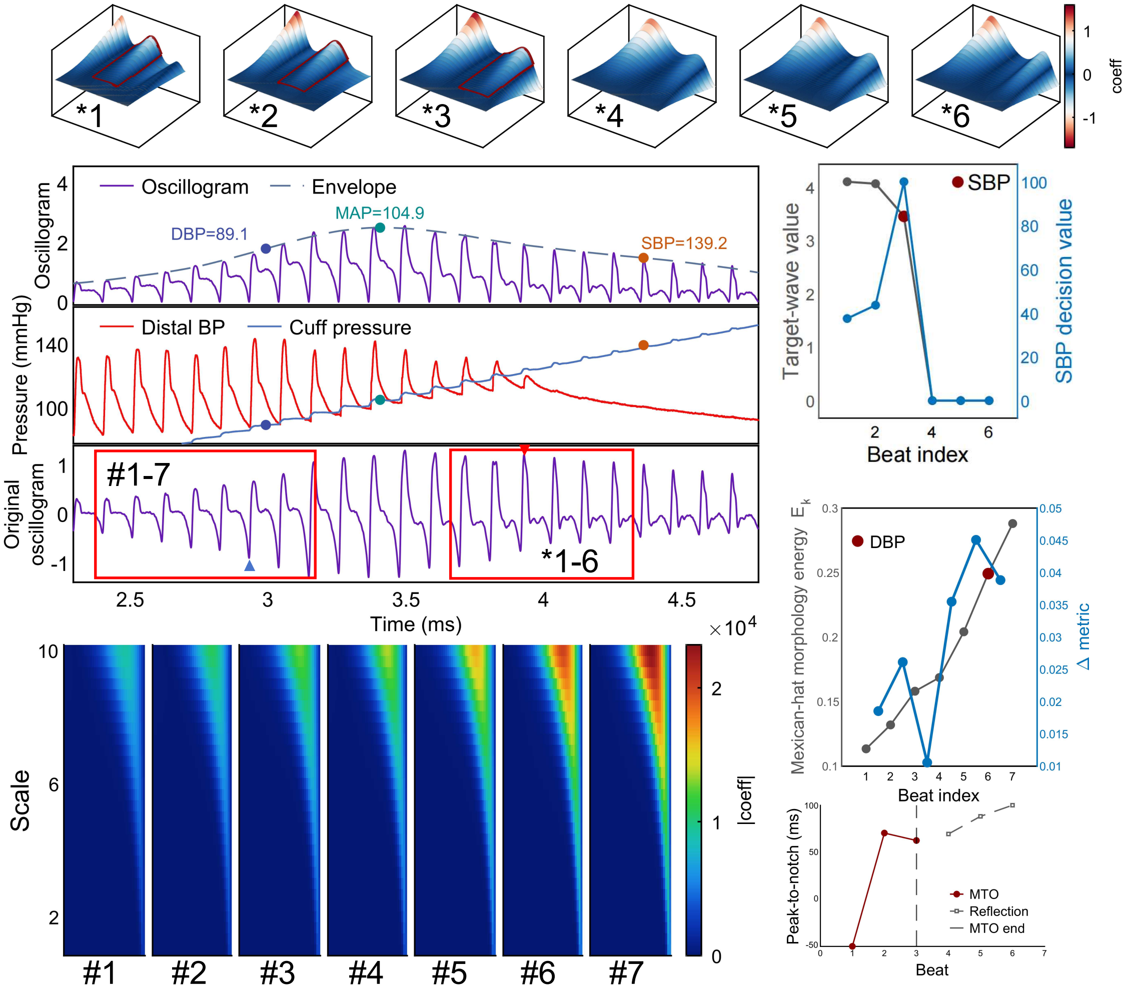

**Supplementary Fig. 24. Accuracy comparison between MTO and MAA.**

This is a representative example in which the amplitude-coefficient method overestimated SBP. The MAA method was applied to the detrended oscillogram, whereas MTO analyzed the original oscillogram. The two red solid boxes indicate the waveform ranges used for DBP and SBP analysis, respectively. Wavelet analysis results are shown in the heat maps and scatter plots. The bottom-right panel shows the peak-to-notch delay of the small-wave and reflection-related components, with the dashed line marking MTO termination.

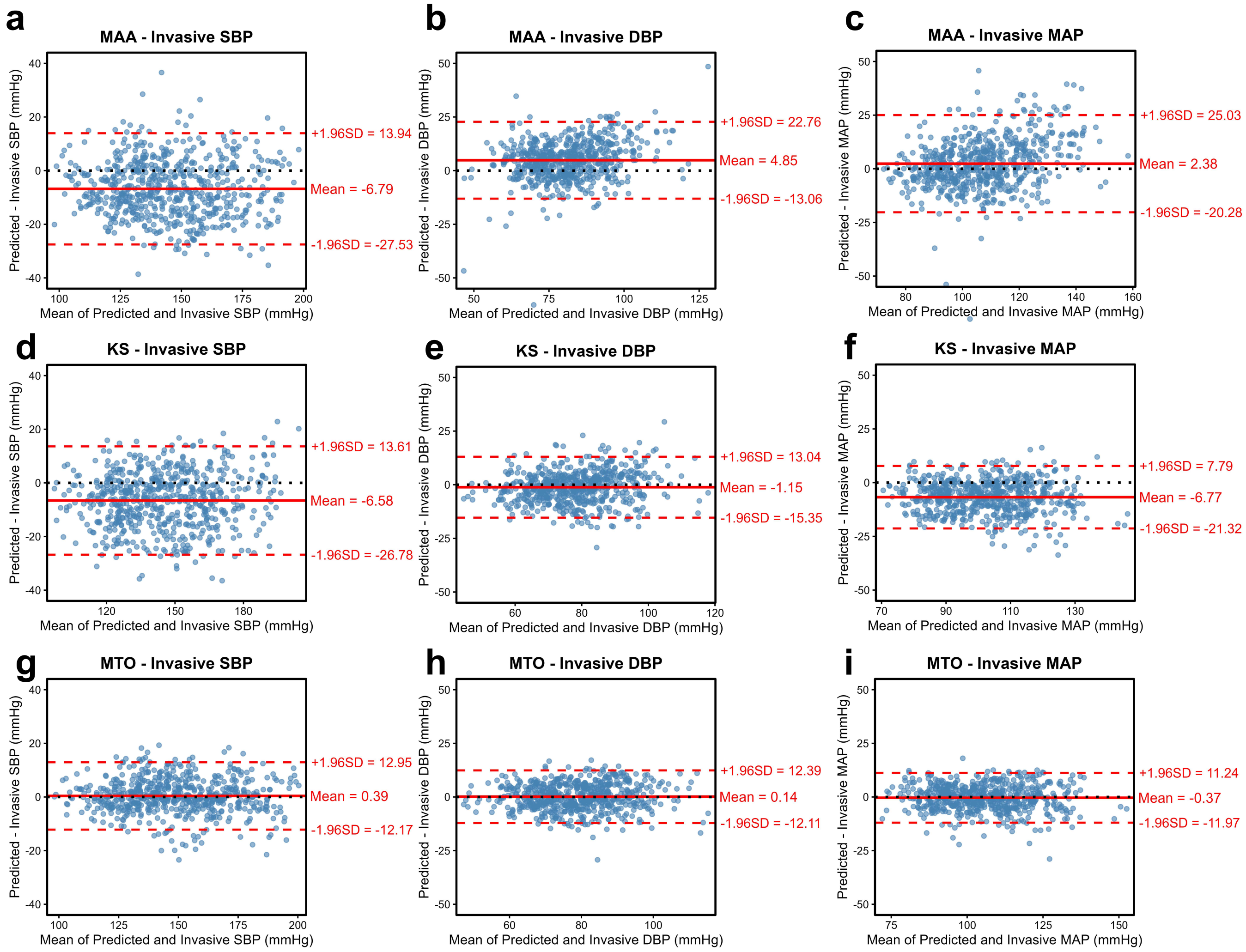

**Supplementary Fig. 25. Bland–Altman analyses of noninvasive BP methods relative to invasive BP.**

Bland–Altman analyses comparing the maximum-amplitude algorithm (MAA), Korotkoff sounds (KS), and the proposed MTO with invasive SBP, DBP, and MAP. The solid red line indicates the mean bias, and the dashed red lines indicate the ±1.96 SD limits of agreement.

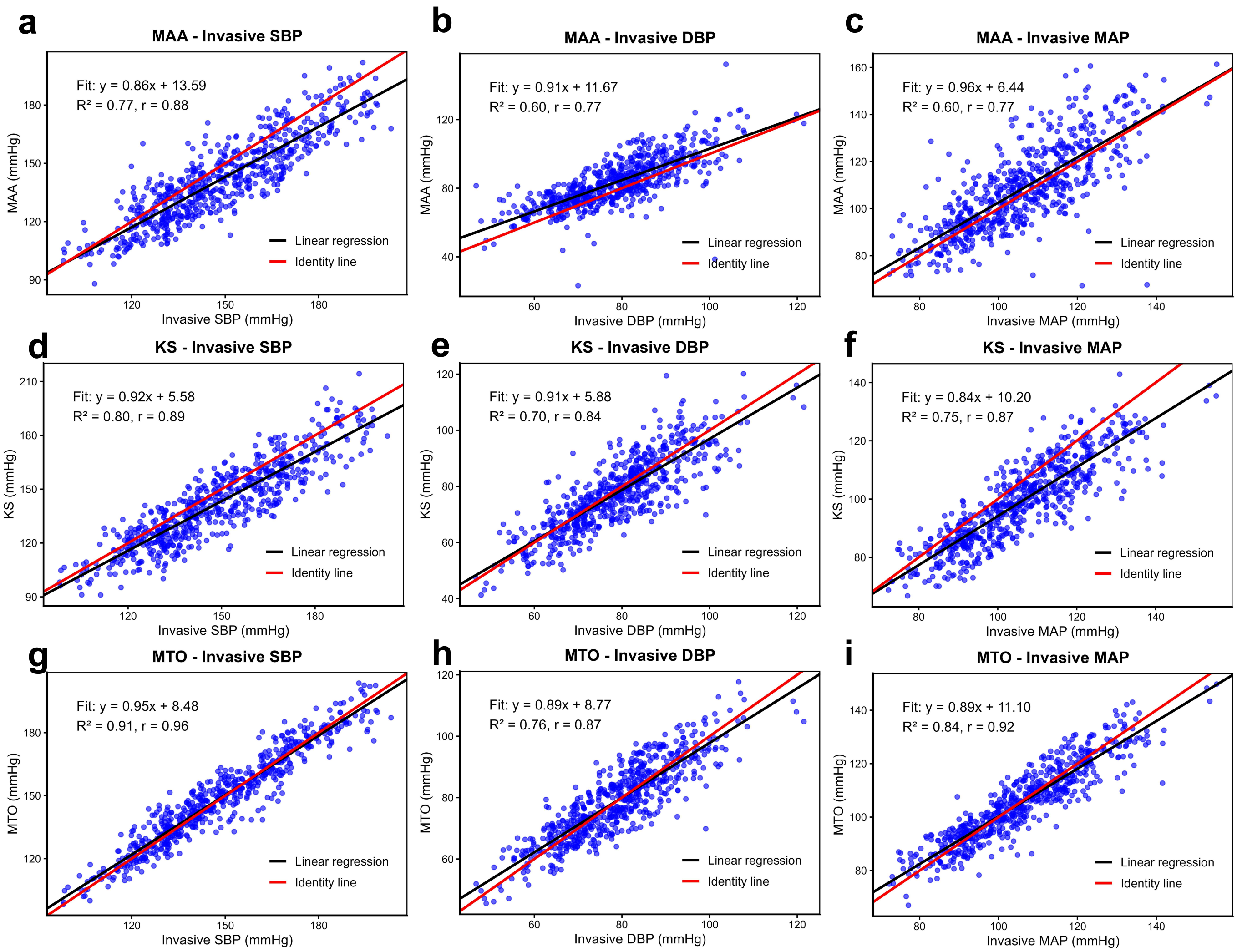

**Supplementary Fig. 26. Correlation analyses of noninvasive BP methods relative to invasive BP.** Linear correlation analyses comparing MAA, KS, and MTO with the corresponding invasive SBP, DBP, and MAP values. Black lines indicate fitted linear regressions, and Pearson correlation coefficients are shown in each panel.

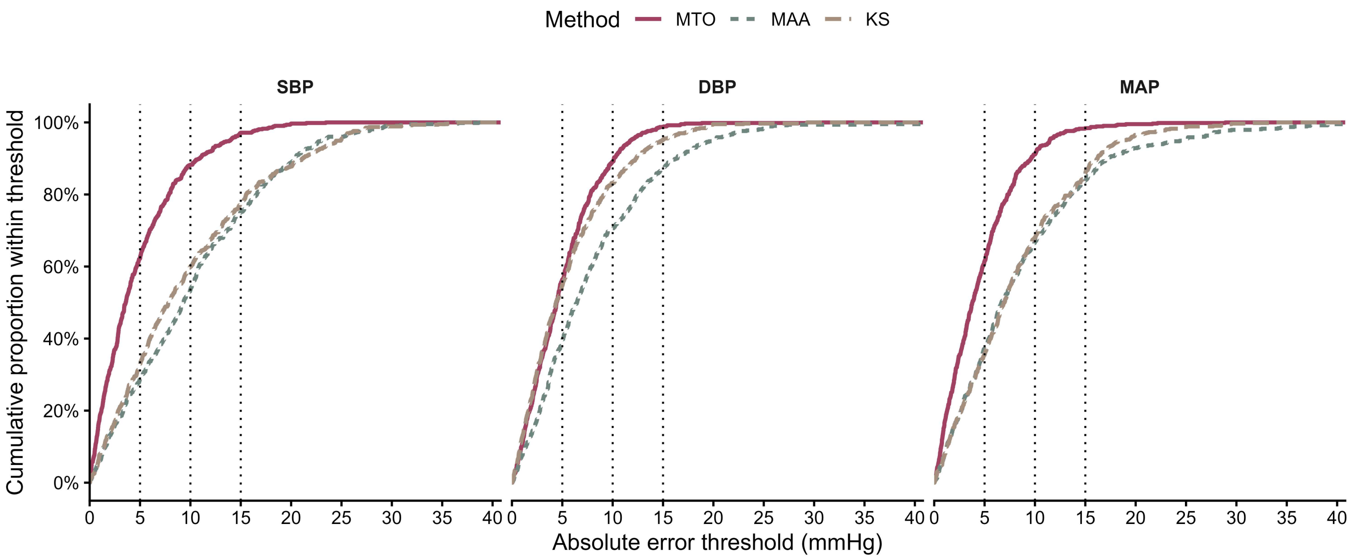

**Supplementary Fig. 27. Cumulative distributions of absolute errors for MTO, MAA, and auscultation relative to invasive blood pressure.**

Cumulative proportions of measurements with absolute errors within increasing thresholds are shown for SBP, DBP and MAP. Vertical dotted lines indicate absolute-error thresholds of 5, 10, and 15 mmHg. A steeper, left-shifted curve indicates that a greater proportion of measurements fall within a given error threshold.
